# Multi-Agent Dynamic Refinement Outperforms Static RAG in Clinical Reasoning for Complex Nephrology Cases

**DOI:** 10.64898/2026.07.15.26358121

**Authors:** Yuichiro Yano, Hiroaki Kakizaki, Hajime Nagasu, Seiji Kishi, Takeo Koshida, Yoshihito Nihei, Akira Hirano, Yuka Sugawara, Takahiro Imaizumi, Yuki Osakabe, Yusuke Sakaguchi, Masaomi Nangaku, Hirotake Mori, Toshio Naito, Mizuki Ohashi, Shoichi Maruyama, Isao Matsui, Yoshitaka Isaka, Hirokazu Okada, Yusuke Suzuki, Naoki Kashihara

## Abstract

**Background:** Large language models (LLMs) struggle with dynamic, longitudinal clinical reasoning. We developed a Multi-Stage Iterative Clinical Reasoning Agent framework to address this gap and systematically decouple the clinical efficacy of static retrieval-augmented generation (RAG) from dynamic self-refinement.

**Methods:** Ten complex longitudinal nephrology cases, rigorously selected via a modified Delphi consensus technique, were blindly evaluated by four board-certified nephrologists and a multi-model AI panel. We compared three architectures across nine cognitive steps: (Model A) a baseline frontier LLM, (Model B) an LLM augmented with static guideline-based RAG, and (Model C) our proposed multi-agent framework featuring RAG integrated with iterative self-critique and refinement.

**Results:** In human evaluations (20-point scale), Model C (mean 17.2, SD 1.2) significantly outperformed both Model A (16.1, 1.3) and Model B (16.2, 1.2) (*P* < 0.001). Implementing static RAG (Model B) yielded no significant improvement over the baseline. Automated AI evaluations (15-point scale) corroborated these findings: Model C (14.7, 0.6) outscored Model A (14.2, 0.9, *P* < 0.001) and Model B (14.3, 0.9, *P* = 0.01). While monolithic models exhibited severe score degradations in planning-heavy tasks such as dynamic differential diagnoses, the multi-agent framework effectively intercepted error cascades, achieving significantly higher diagnostic accuracy (mean 17.6, *P* = 0.019) and therapeutic management scores (17.3, *P* = 0.002).

**Conclusions:** Static knowledge retrieval alone fails to enhance frontier LLM performance in longitudinal medical reasoning. Distributing clinical workflows into a multi-agent dynamic refinement pipeline significantly improves reasoning completeness, intercepts error cascades, and safely resolves planning bottlenecks in complex patient care.

## Introduction

The integration of Large Language Models (LLMs) into healthcare marks a transformative shift from basic information retrieval to sophisticated clinical reasoning systems. [1,2] Advanced prompting techniques, such as Chain-of-Thought (CoT), have enabled LLMs to externalize intermediate reasoning steps, thereby improving diagnostic accuracy and transparency. [3,4] Furthermore, recent breakthroughs utilizing Reinforcement Learning (RL), exemplified by models like DeepSeek-R1 and Med-R1, have cultivated emergent reasoning capabilities by allowing models to learn through exploration. [5–7] However, the application of these frontier models in high-stakes medical environments remains constrained by critical limitations. Recent rigorous evaluations, such as the HealthBench framework, reveal that despite achieving high average scores, state-of-the-art models exhibit concerning fragility and unreliability in worst-case, critical scenarios. [8,9]

Clinical decision-making is rarely a static, single-encounter event; rather, it demands longitudinal reasoning across dynamic patient states. Physicians continuously update differential diagnoses and therapeutic strategies as new clinical data—such as laboratory test results and treatment responses—unfold over time. While recent multi-agent collaborative frameworks (e.g., MDTeamGPT) have successfully simulated multi-disciplinary team consultations, they primarily focus on cross-sectional case analysis. [10] Adapting LLMs to process temporal sequences of complex medical data and dynamically self-correct their reasoning remains a critical, unsolved frontier in medical AI.

To address this gap, we developed a Multi-Stage Iterative Clinical Reasoning Agent framework designed to process longitudinal medical data. Specifically, the system sequentially evaluates chronological clinical inputs—initial patient data, subsequent test results, and treatment information—to mirror real-world diagnostic workflows. While modern advanced LLMs are often hypothesized to have already internalized vast amounts of medical knowledge during their pre-training phases, the actual efficacy and necessity of incorporating external knowledge bases in complex reasoning tasks remain a subject of debate. To systematically ascertain whether Retrieval-Augmented Generation (RAG) is truly effective compared to a model’s intrinsic knowledge, and to decouple the effects of knowledge retrieval from dynamic reasoning, we designed a comparative study. We validated our framework in the domain of nephrology by conducting a rigorous blind evaluation of longitudinal case reports, assessed by four independent board-certified nephrologists. We compared the performance of three architectural models: (1) a baseline LLM relying solely on intrinsic knowledge without external augmentation, (2) an LLM augmented with static RAG based on clinical practice guidelines, and (3) our proposed RAG framework integrated with a multi-agent system (Initial Assessor, Clinical Evaluator, and Refinement agent) for dynamic refinement.

## Methods

### Methodology for Case Selection

An expert panel comprising three nephrologists (SK, TK, and YN) established predefined selection criteria emphasizing diagnostic complexity, educational value, and comprehensive coverage of the nephrology field. To ensure the selected cases reflected contemporary clinical practice, we conducted a literature search of CEN Case Reports via PubMed for relevant articles published between January 2024 and October 2025. This initial search yielded 53 candidate case reports.

We subsequently employed a modified Delphi technique to rigorously evaluate and select the optimal cases. Following an initial screening by two facilitators (HN and AH), the expert panel independently rated the clinical suitability of 28 shortlisted candidate cases across two consecutive Delphi rounds. Based on the aggregated quantitative ratings and qualitative peer feedback from these rounds, the panel convened for a final consensus meeting to review the 20 highest-scoring cases. Ultimately, 10 cases were selected to ensure a balanced, high-yield, and comprehensive representation of the nephrology spectrum.

A comprehensive description of the predefined selection criteria, the complete literature search strategy, and the detailed step-by-step procedures of the Delphi process are provided in the Supplementary Material.

### Proposed Architecture and Evaluation Framework

We developed a multi-stage, iterative multi-agent architecture built on the Dify platform, utilizing gpt-5.4-2026-03-05 as the base model. The system mirrors the clinical workflow through three cascaded steps: Initial Clinical Assessment, Diagnostic Refinement, and Therapeutic Evaluation (Figure 1). To overcome the limitations of standalone models, each step integrates retrieval-augmented generation (RAG) using a physician-curated knowledge base, followed by an automated self-reflection and refinement loop. Within this iterative process, an initial reasoning agent generates a response, a clinical evaluator agent critiques it for logical consistency and safety, and a refinement agent finalizes the output before propagating it to the subsequent clinical step.

**Figure 1.**
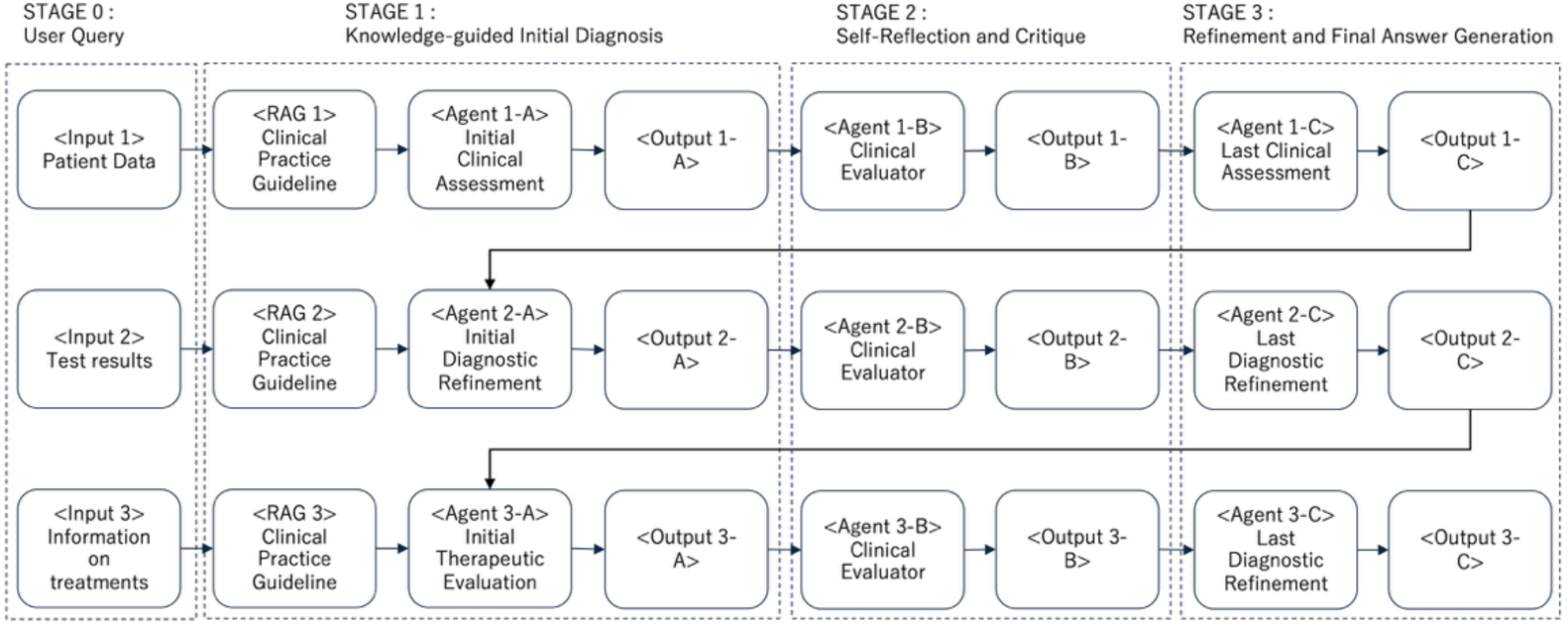
Comparison Conditions for Component Evaluation. The proposed method has a structure in which three steps, aligned with the clinical workflow, are connected vertically in a cascade. Each step corresponds to an independent clinical subtask: Step 1 performs the Initial Clinical Assessment, Step 2 the Diagnostic Refinement, and Step 3 the Therapeutic Evaluation.

To isolate the specific contributions of knowledge augmentation and self-critique, we evaluated three experimental conditions: a baseline model (LLM only) (Figure S1), a knowledge-grounded model (LLM + RAG) (Figure S2), and the complete proposed architecture (LLM + RAG + Self-Refinement) (Figure 1). The clinical reasoning quality of each condition was systematically decomposed into nine distinct cognitive tasks (Table 1). [11] Performance was rigorously assessed through two parallel approaches: a blinded, randomized evaluation by expert physicians using a 5-point Likert scale, and a multi-model LLM-as-a-judge framework (utilizing gpt-5.4-2026-03-05, Gemini 3.1 Pro Preview, and Claude Opus 4.7) carefully designed to mitigate individual model biases.

**Table 1.** The nine cognitive steps comprising clinical reasoning and the reasoning tasks evaluated.

| Clinical step | Cognitive step | Reasoning task evaluated |
| --- | --- | --- |
| **Step 1: Initial Clinical Assessment** | 1. Medical summarization | Summarize the patient as a medical problem |
|  | 2. Differential diagnosis | Present three differential diagnoses (the most likely / the “must-not-miss”) |
|  | 3. Additional data acquisition | Propose additional history-taking and physical examination to test the hypotheses |
|  | 4. Diagnostic planning | Formulate an investigation plan with its purpose and expected results |
| **Step 2: Diagnostic Refinement** | 5. Interpretation of results | Interpret the test results in the context of the patient’s condition |
|  | 6. Diagnostic updating | Update the diagnostic reasoning and present the most likely working diagnosis |
|  | 7. Treatment planning | Propose a treatment plan with its rationale and alternative options |
| **Step 3: Therapeutic Evaluation** | 8. Monitoring and risk management | Propose strategies to monitor treatment efficacy, specific risks, and countermeasures |
|  | 9. Contingency planning | Propose actions for cases that fail to improve or that deteriorate suddenly |

Comprehensive methodological details—including exact API versions, system prompts, the RAG configuration process, the nine-step cognitive decomposition matrix, and the specific scoring rubrics—are provided in the Supplementary Material.

### Statistical Analysis

The generated outputs were blindly assessed using a predefined rubric by four independent board-certified nephrologists (maximum 20 points; 5 per evaluator) and, concurrently, by three distinct AI models to provide an objective counterpart (maximum 15 points; 5 per model). Total scores were aggregated by summing the itemized scores across all clinical cases and reasoning steps.

Continuous variables are presented as means with standard deviations (SD). Global performance differences across the three architectural paradigms were evaluated using the Kruskal-Wallis test. Post-hoc pairwise comparisons were performed using the Wilcoxon rank-sum test, with p-values adjusted via the Bonferroni correction. To identify vulnerabilities within specific clinical reasoning steps, we analyzed score distributions per question and employed heatmap visualizations coupled with step-specific Kruskal-Wallis tests.

A two-sided p-value < 0.05 was considered statistically significant. All analyses were performed using R software (version 4.5.3; R Foundation for Statistical Computing, Vienna, Austria).

### Ethics Statement

Ethical review and Institutional Review Board (IRB) approval were waived for this study as it falls outside the scope of human subjects research. The study involved a secondary analysis of publicly available, fully anonymized case reports derived from published literature. This complies with our institutional policies and national ethical guidelines, which explicitly exempt research utilizing only widely available, anonymized existing data with established academic value. The study did not involve any experiments on human participants or animals, and no identifiable personal health information was accessed.

## Results

### Overall Clinical Reasoning Performance and the Efficacy of Knowledge Augmentation

We first evaluated the overall performance of each model. In the human evaluation (Figure 2, out of a maximum score of 20 points), the baseline LLM (Model A) achieved a mean score of 16.1 (SD = 1.3). Augmenting the system with static RAG (Model B) resulted in a comparable mean score of 16.2 (SD = 1.2; ns vs. Model A). Conversely, our proposed multi-agent framework (Model C) achieved a mean score of 17.2 (SD = 1.2), achieving significantly higher scores compared to both Model A and Model B (*P* < 0.001).

**Figure 2.**
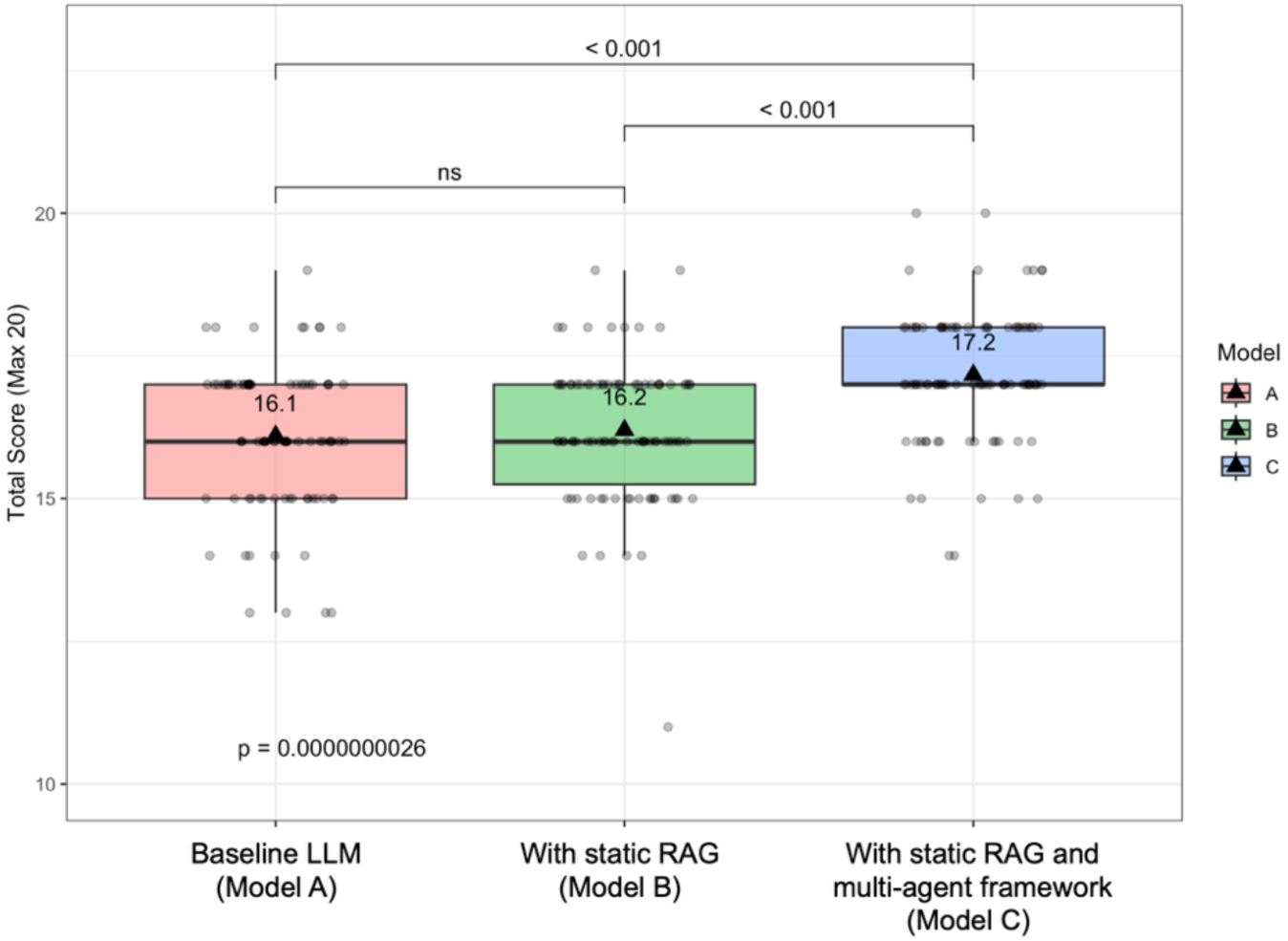
Comparison of overall clinical reasoning performance among architectural models by human evaluations. Box plots overlaid with jittered individual data points represent the distribution of total scores across the three architectural models, by the blinded evaluation by board-certified nephrologists (maximum score = 20). Black triangles and text labels indicate the mean score for each model. Global p-values were derived from the Kruskal-Wallis test, and post-hoc pairwise comparisons were performed using the Wilcoxon rank-sum test with Bonferroni correction.

The AI evaluation (Figure S3, out of a maximum score of 15 points) yielded remarkably similar trends. The baseline Model A and the static RAG Model B achieved mean scores of 14.2 (SD = 0.9) and 14.3 (SD = 0.9), respectively, with no statistically significant difference between them (ns). However, Model C achieved the highest mean score of 14.7 (SD = 0.6), showing statistically significant differences when compared to both Model A (*P* < 0.001) and Model B (*P* = 0.01).

### Identification of Inherent Challenges in Specific Clinical Reasoning Steps

We analyzed the pooled scores across all models to identify specific clinical steps where AI-generated outputs inherently demonstrated vulnerabilities. In the human evaluation (Figure S4, evaluated on the 20-point scale), evaluators recorded the lowest mean score in Q8 (Treatment evaluation; Mean: 16.3), while also assigning relatively lower scores to Q2 (Differential Diagnoses), Q3 (Physical Examinations), Q4 (Diagnostic plan), and Q6 (Diagnostic reassessment) (all Mean: 16.4), compared to baseline data summarization (Q1; Mean: 16.7) or test interpretations (Q5; Mean: 16.7).

The AI evaluation (Figure S5, evaluated on the 15-point scale) clearly delineated similar challenging tasks, with the lowest mean scores consistently observed in Q2 (Differential Diagnoses; Mean: 13.7), Q4 (Diagnostic plan; Mean: 13.7), and Q7 (Treatment planning; Mean: 13.7).

### Temporal Dynamics of Iterative Refinement in Longitudinal Clinical Workflows

In the human evaluation (Figure 3), during the Initial Assessment stage, Model C recorded the highest score in Q3 (Mean: 17.7, *P* = 0.016). This trend was supported by the AI evaluation (Figure S6), which also indicated a strong trend toward higher scores for Model C in Q3 (Mean: 14.6, *P* = 0.079). Moving to the Diagnostic Refinement stage, human evaluators found that Model C demonstrated the highest diagnostic accuracy score in Q5 (Mean: 17.6, *P* = 0.019).

**Figure 3.**
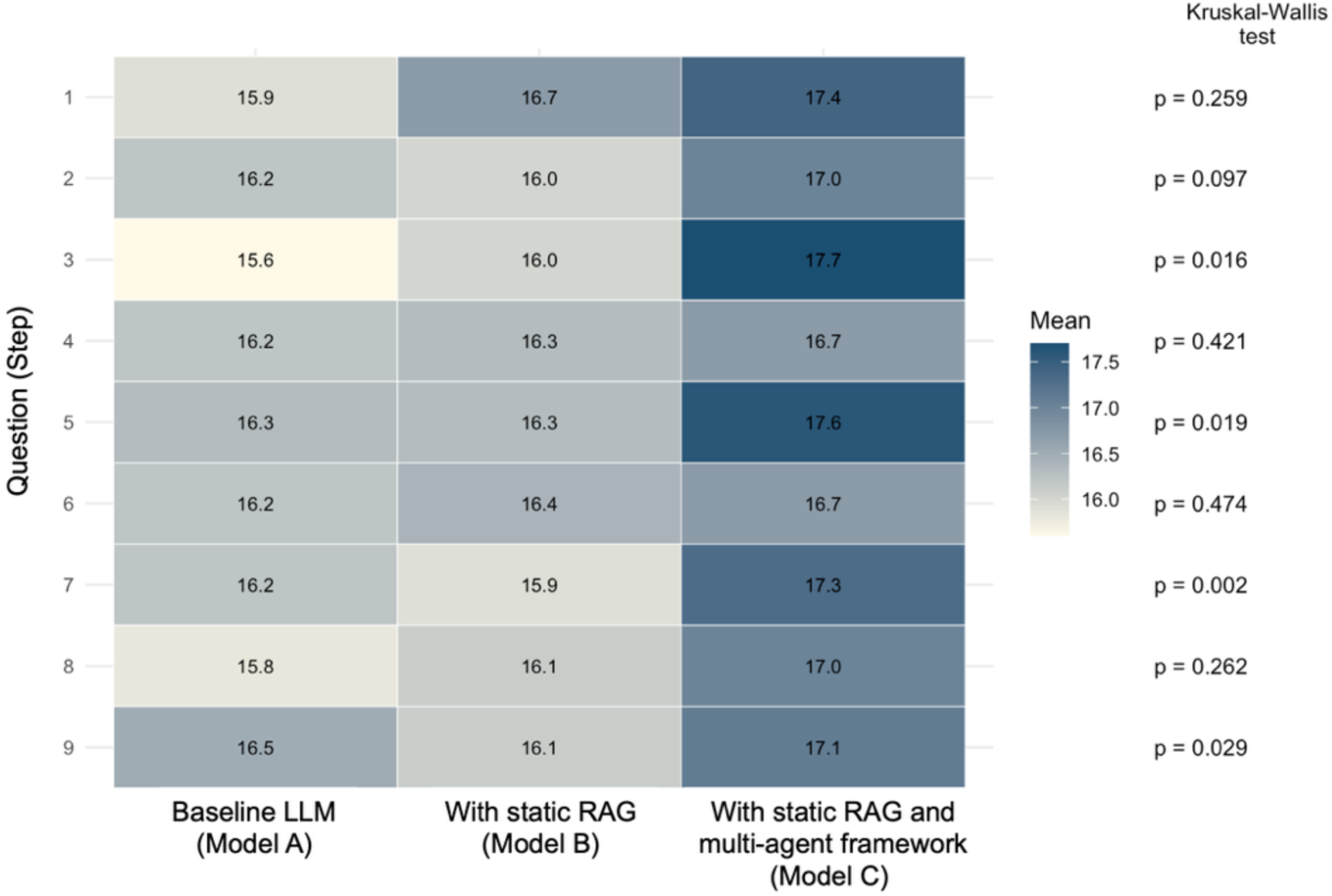
Heatmap of average scores by question step and paradigm by human evaluations. The matrix visualizes the mean score for each clinical reasoning step across the three architectural models for the human expert evaluation, with darker colors representing superior performance. The right-hand panels display the corresponding p-values from question-specific Kruskal-Wallis tests, indicating whether performance differences between models were statistically significant at that specific step.

Concurrently, the AI evaluation highlighted a highly significant difference in Q6, where Model C achieved a notably high mean score, reaching the maximum ceiling of the 15-point scale (Mean: 15.0, *P* = 0.001), showcasing the effectiveness of the multi-agent critique in synthesizing complex laboratory findings. The most pronounced divergence between models was observed in the Therapeutic Management stage. Based on human assessment, Model C achieved the highest scores in Q7 (Mean: 17.3, *P* = 0.002) and Q9 (Mean: 17.1, *P* = 0.029). The AI evaluation echoed this later-stage divergence, with Model C recording the highest score in Q8 (Mean: 14.9, *P* = 0.002).

## Discussion

We demonstrate that augmenting a frontier LLM with static RAG (Model B) yielded no statistically significant improvement over the unaugmented baseline (Model A) in either human or AI automated evaluations. This suggests that modern frontier architectures have already internalized the core corpus of medical literature within their parametric knowledge base. [12] The primary bottleneck in medical AI is rarely static fact retrieval, but rather the dynamic contextual synthesis of time-varying clinical metrics (e.g., electrolyte fluctuations, acid-base equilibrium, fluid dynamics). In uncertain domains like nephrology, static RAG may deplete the model’s limited context window with redundant guideline texts, wasting attention resources that should otherwise track subtle temporal variances. This aligns with recent longitudinal EHR evaluations (e.g., EHR-RAG), which demonstrate that flat, vector-search RAG often induces information dropping and incomplete evidence coverage. [13]

Our step-by-step analysis uncovers a cognitive topology across monolithic language models, characterized by severe score degradations at planning-heavy junctures: Q2 (Differential Diagnoses), Q4 (Diagnostic Plan), and Q7/Q8 (Therapeutic Management). This aligns with findings by Rao et al., [14] which demonstrated that while state-of-the-art models achieve near-perfect accuracy (>90%) on static final diagnoses once all data is provided, their failure rates exceed 80% when generating evolving differential diagnoses. The models might exhibit premature closure bias, providing an early single diagnostic possibility based on sparse initial inputs and ignoring competing alternatives. This deficit stems not from a lack of medical knowledge, but from a failure in reasoning completeness under uncertainty. MedR-Bench revealed a performance gap [15]: while models achieve 81.34% accuracy in static knowledge retrieval, their success rate plummets to just 22.34% when tasked with managing dynamic, patient-specific clinical workflows. While LLMs excel at passive informational summarization (Q1, Q5), they encounter severe cognitive constraints in forward-planning tasks that require weighting hidden variables like subsequent test results or treatment responses.

Our proposed Multi-Stage Iterative framework (Model C) resolves these cognitive gaps through architectural scaffolding. Grounded in dual-process theory, monolithic LLMs operate predominantly under "System 1" (intuitive, rapid statistical associations).

Emulating "System 2" (analytical, logical self-verification) within AI requires a structural framework that deliberately distributes cognitive load and segregates reasoning phases. [16,17] Standalone LLMs are statistical engines that lack internal hypothesis testing, preventing them from actively questioning their own assumptions. [12] The Clinical Evaluator acts as an external metacognitive auditor, forcing the system to cross-examine early diagnostic claims against incoming lab results, effectively breaking the single-turn pattern-matching dependency. In our system, Agent B prevents a cascade of cognitive errors by challenging Agent A’s (the Initial Assessor) preliminary assumptions and cross-examining them against incoming, longitudinal lab results. This architecture aligns with ClinicalAgents [18], which use Monte Carlo Tree Search (MCTS) and dual-memory configurations for hypothesis falsification, and TeamMedAgents [19], which leverage Salas’ teamwork theory to achieve mutual monitoring and dynamic consensus. Our framework extends this concept into a specialized temporal dimension, utilizing its distributed architecture to dynamically modulate response depth and context-awareness across shifting patient timelines.

Our automated AI evaluation echoed the human evaluation trends, suggesting both Model C’s overwhelming statistical superiority and the shared structural vulnerabilities across Q2, Q4, and Q7. This indicates that model-based clinical evaluation could achieve tight alignment with human expert consensus when guided by objective, granular grading rubrics. This dual validation confirms that scalable, automated meta-evaluation frameworks may be utilized to track future architectural iterations, mitigating the astronomical costs and time latencies traditionally mandatory for expert manual chart reviews. [20]

Several limitations must be acknowledged. First, clinical case reports represent inherently curated, clean, and chronologically organized narratives compiled for educational purposes. Real-world hospital systems operate on fragmented and messy electronic health records (EHRs) where vital parameters are frequently missing or ambiguous. Second, clinical medicine remains heavily dependent on non-text signals. Our current multi-agent system does not process visual diagnostics (e.g., ultrasound imaging, histopathology slides) or auditory signals (e.g., a patient’s acute level of distress), which are vital for holistic bedside triage. Third, potential data leakage exists because our dataset comprises case reports from subscription-based journals; although these are not open-access, the LLMs might still have internalized them during pre-training. Fourth, multi-agent framework (Model C) invokes the LLM three times per step (generation, critique, refinement), whereas Models A and B use single-pass queries. This leaves the contribution of specialized multi-agent role-playing versus an expanded inference-time compute budget unseparated.

In summary, our research provides evidence that expert-level clinical reasoning may not be accomplished through the mere scaling of static knowledge retrieval layers. By organizing clinical workflows into a distributed, multi-agent dynamic refinement pipeline, our Multi-Stage framework intercepts error cascades, resolves the planning bottleneck, and achieves a statistically significant breakthrough in reasoning completeness. As commercial healthcare models rapidly transition toward clinical deployment, adopting robust multi-agent orchestration will be paramount to bridge the gap between marketing claims and the unforgiving, safe reality of bedside patient care.

## Data Availability Statement

The datasets generated or analyzed during this study are available from the corresponding author on reasonable request.

## Author contribution

YY conceptualized the study and developed the methodology. HN, SK, TK, and YN curated the data. HK and MO conducted the formal analysis. AH, MN, HM, TN, SM, IM, YI, HO, YS, and NK supervised the study. TN acquired funding. YY drafted the manuscript, and all authors reviewed, edited, and approved the final version.

## Declaration of Generative AI Use

The authors used Gemini 3.5 to refine the English composition. All AI-generated content was critically reviewed by the authors. The experimental evaluation of the three LLMs (gpt-5.4-2026-03-05, Gemini 3.1 Pro Preview, and Claude Opus 4.7) carefully designed to mitigate individual model biases.) was conducted independently of these writing-assistant tools.

