## Supplemental Method, Figure, Tables for "Multi-Agent Dynamic Refinement Outperforms Static RAG in Clinical Reasoning for Complex Nephrology Cases"

Running headline: Multi-Agent Nephrology Clinical Reasoning

Yuichiro Yano;[1,2] Hiroaki Kakizaki:[3] Hajime Nagasu;[4] Seiji Kishi;[5] Takeo Koshida;[5] Yoshihito Nihei;[6] Akira Hirano;[4] Yuka Sugawara;[7] Takahiro Imaizumi:[8] Yuki Osakab;[5] Yusuke Sakaguchi;[9] Masaomi Nangaku;[7] Hirotake Mori; [1] Toshio Naito; [1] Mizuki Ohashi;[1] Shoichi Maruyama;[10] Isao Matsui;[9] Yoshitaka Isaka;[9] Hirokazu Okada;[11] Yusuke Suzuki;[6]and Naoki Kashihara[4]

1. Department of General Medicine, Juntendo University Faculty of Medicine, Tokyo, Japan
2. Artificial Intelligence Incubation Farm, Juntendo University Faculty of Medicine, Tokyo, Japan
3. PeopleMedia, Inc, Tokyo, Japan
4. Department of Nephrology and Hypertension, Kawasaki Medical School, Kurashiki, Okayama, Japan
5. Department of Endocrinology, Metabolism and Nephrology, Kochi Medical School, Kochi University, Kohasu, Oko-cho, Nankoku, Kochi, 783-8505, Japan.
6. Department of Nephrology, Juntendo University Faculty of Medicine, Tokyo, Japan
7. Division of Nephrology and Endocrinology, The University of Tokyo Graduate School of Medicine, Tokyo, Japan
8. Department of Advanced Medicine, Nagoya University Hospital, Nagoya, Japan.
9. Department of Nephrology, The University of Osaka Graduate School of Medicine, Suita, Japan.
10. Department of Nephrology, Nagoya University Graduate School of Medicine, Aichi, Japan
11. Department of Nephrology, Faculty of Medicine, Saitama Medical University, Saitama, Japan.

**Corresponding author:**
Yuichiro Yano, MD, PhD, FAHA

Professor, Department of General Medicine
Director, AI Incubation Farm
Juntendo University Faculty of Medicine
ZIP: 113-8421 2-1-1, Hongo, Bunkyo-Ku, Tokyo, JAPAN


**Supplementary methods**

**Ethics Statement:**

Ethical review and Institutional Review Board (IRB) approval were waived for this study as it falls outside the scope of human subjects research. The study involved a secondary analysis of publicly available, fully anonymized case reports derived from published literature. This complies with our institutional policies and national ethical guidelines, which explicitly exempt research utilizing only widely available, anonymized existing data with established academic value. The study did not involve any experiments on human participants or animals, and no identifiable personal health information was accessed.

***Methodology for Case Selection to Evaluate Large Language Models Using the Delphi Technique***

**1. Preparatory Phase: Establishing Selection Criteria**

An expert panel, comprising three nephrologists (SK, TK, and YN), conducted a preliminary discussion to establish consensus on the essential criteria for case selection in the context of clinical reasoning in nephrology. The agreed-upon criteria were as follows:

i. **Comprehensive Coverage of Nephrology:** The cases should collectively cover a wide spectrum of the field of nephrology.
ii. **Avoidance of Simple Cases:** Cases with a limited or straightforward differential diagnosis were to be avoided.
iii. **Complexity of Diagnosis and Pathophysiology:** Priority was given to cases involving complex diagnostic processes or intricate pathophysiology.
iv. **Balance between Complexity and Rarity:** While prioritizing complexity (criterion iii), extremely rare diseases infrequently encountered in clinical practice were to be avoided where possible.
v. **Potential for High Educational Value:** Cases should present opportunities for significant learning, such as highlighting potential diagnostic errors or critical decision points.
vi. **Complexity in Post-Diagnostic Management:** The cases should involve complex considerations regarding post-diagnostic management and the course of treatment.

### 2. Literature Search and Candidate Case Compilation

To ensure that the selected cases reflected contemporary clinical practice following the publication of the CKD Clinical Practice Guidelines 2023 (published in June 2023), we searched for recent case reports in CEN Case Reports, a peer-reviewed journal dedicated to nephrology case reports. We searched PubMed for case reports published between January 2024 and October 2025 using the following query: ("CEN Case Rep"[Journal]) AND ("Case Reports"[Publication Type]) AND ("2024/01"[Date - Publication] : "2025/10"[Date - Publication]) NOT ("Infant"[Mesh] OR "Child"[Mesh] OR "Adolescent"[Mesh] OR "Neoplasms"[Mesh] OR "Genetic Diseases, Inborn"[Mesh] OR "Genetic Predisposition to Disease"[Mesh] OR "COVID-19"[Mesh] OR "SARS-CoV-2"[Mesh] OR "red yeast rice"[tiab] OR "Monascus"[tiab] OR "Renal Dialysis"[Mesh] OR "Peritoneal Dialysis"[Mesh] OR "Kidney Transplantation"[Mesh]). The final search yielded 53 candidate case reports.

### 3. The Delphi Process for Case Evaluation

**Round 1:**

1. Two non-voting facilitators (HN and AH) first reviewed the titles of all 53 candidate cases and excluded those judged inappropriate for the present study, narrowing the pool to 28 cases. They then distributed this list of 28 candidate cases to the expert panel, together with an evaluation form. Each case was accompanied by a summary and, where possible, a link to the full-text article. When a sufficient number of cases was available, open-access articles were prioritized to avoid copyright issues.
2. Each panelist (SK, TK, and YN) independently and anonymously rated the “suitability” of each candidate case on a 5-point Likert scale (1 = Not at all suitable, 2 = Not very suitable, 3 = Neutral/undecided, 4 = Suitable, and 5 = Highly suitable). The specific evaluation criteria for each item are detailed in Supplementary Table 1.
3. For each case, panelists were required to provide a written rationale for their rating, outlining the key criteria they considered and any concerns they had.
4. The completed evaluation forms were returned to the facilitators.

**Analysis and Feedback (Post-Round 1):**

1. The facilitators collected the responses and calculated the mean and median suitability ratings for each case.
2. The facilitators compiled and synthesized the panelists’ written comments for each case.

3. A feedback report summarizing the quantitative scores and qualitative comments from the panelists for each case was prepared.

**Round 2:**

1. The facilitators distributed this feedback report to the panel along with the same list of candidate cases (or a list narrowed based on scores).
2. The panelists reviewed the aggregated results and peer feedback, then reassessed each case. Panelists who chose to change their rating submitted revised scores and rationales; otherwise, their Round 1 ratings were carried forward to form the second-round evaluation.

**Consensus and Subsequent Rounds:**

1. The facilitators analyzed the results of Round 2, summarizing the suitability of each case by its mean and median rating. The cases were then ranked according to these ratings to inform the subsequent selection.
2. A third round, to be conducted in the same manner, was planned in case further refinement of the rankings was required. The rankings were essentially unchanged between Round 1 and Round 2; accordingly, no third round was conducted.

**4. Final Case Selection Meeting**

1. Based on the final results of the Delphi process, including the final mean and median scores and the summarized comments, the facilitators prepared a shortlist of the 20 highest-scoring cases.
2. The facilitators then convened a final consensus meeting (held in person or online) with the expert panel. During this meeting, the shortlist and the complete, anonymized results of the Delphi rounds were presented.
3. In this final stage, the panel discussed the shortlisted cases not only on their individual merits but also in the context of the entire set, ensuring that the final selection of 10 cases met criterion i: comprehensive coverage of nephrology.

**5. Final Analysis and Reporting**

The final set of 10 selected cases and the detailed rationale for their selection, including the results from the Delphi process and the final consensus meeting, were documented for reporting.

**Proposed Architecture**

The proposed method has a structure in which three steps, aligned with the clinical workflow, are connected vertically in a cascade (Figure S1, S2, 1). Each step corresponds to an independent clinical subtask: Step 1 performs the Initial Clinical Assessment, Step 2 the Diagnostic Refinement, and Step 3 the Therapeutic Evaluation.

The design of this architecture is grounded in the following three lines of prior evidence. First, it has been reported that having multiple LLM agents collaborate in a division of roles yields medical reasoning performance superior to that of a single model [1,2]. Second, augmenting an LLM with external knowledge—such as clinical practice guidelines—via retrieval-augmented generation (RAG) has been shown to improve the factuality, completeness, and safety of its outputs [3]. Third, approaches in which an LLM critiques and iteratively revises its own output (e.g., Self-Refine and Reflexion) have been reported to improve output quality across diverse tasks without requiring additional training [4,5]. Indeed, agent-based systems that combine retrieval-based grounding in clinical guidelines and drug formularies with structured reasoning have recently been shown to approach the level of specialist physicians in the domain of treatment and management as well [6]. The present study is distinguished by integrating these insights into each subtask of clinical reasoning and implementing knowledge augmentation, self-critique, and refinement as a single iterative flow.

Within each step, an iterative process comprising the following three stages was carried out. In the first stage (Knowledge-guided Initial Diagnosis), after augmenting the model with clinical guideline–derived knowledge through RAG from the knowledge base described below, the initial reasoning agent (Agent X-A) generates the initial output for that step (Output X-A). In the second stage (Self-Reflection and Critique), the clinical evaluation agent (Agent X-B, Clinical Evaluator) critiques the initial output and identifies issues from the perspectives of logical consistency, clinical validity, and safety (Output X-B). In the third stage (Refinement and Final Answer Generation), the refinement agent (Agent X-C) revises the output in light of the critique and generates the final output for that step (Output X-C).

The connection between steps was designed so that the refined final output of each step is propagated to the subsequent step. Specifically, Output 1-C is provided as the input to Agent 2-A in Step 2, and Output 2-C as the input to Agent 3-A in Step 3, so that the results of self-critique and refinement in the upstream steps are cumulatively reflected in the downstream diagnostic and therapeutic reasoning.

**Decomposition of Clinical Reasoning into Nine Cognitive Steps**

To systematically evaluate the clinical reasoning of the proposed architecture, we decomposed the clinical reasoning across the three steps into nine cognitive steps, following our previous study [7] (see below Table). The final output of each step (Agents X-A through X-C) is evaluated as a set of specific reasoning tasks corresponding to these cognitive steps. Specifically, Step 1 (Initial Clinical Assessment) corresponds to cognitive steps 1–4, Step 2 (Diagnostic Refinement) to cognitive steps 5–7, and Step 3 (Therapeutic Evaluation) to cognitive steps 8–9.

| Clinical step | Cognitive step | Reasoning task evaluated |
|---|---|---|
| **Step 1: Initial Clinical Assessment** | 1. Medical summarization | Summarize the patient as a medical problem |
| | 2. Differential diagnosis | Present three differential diagnoses (the most likely / the "must-not-miss") |
| | 3. Additional data acquisition | Propose additional history-taking and physical examination to test the hypotheses |
| | 4. Diagnostic planning | Formulate an investigation plan with its purpose and expected results |
| **Step 2: Diagnostic Refinement** | 5. Interpretation of results | Interpret the test results in the context of the patient's condition |
| | 6. Diagnostic updating | Update the diagnostic reasoning and present the most likely working diagnosis |
| | 7. Treatment planning | Propose a treatment plan with its rationale and alternative options |
| **Step 3: Therapeutic Evaluation** | 8. Monitoring and risk management | Propose strategies to monitor treatment efficacy, specific risks, and countermeasures |
| | 9. Contingency planning | Propose actions for cases that fail to improve or that deteriorate suddenly |

**Table. The nine cognitive steps comprising clinical reasoning and the reasoning tasks evaluated (based on our previous study [7])**

**Models and RAG Configuration**

For all agents (Agents X-A, X-B, and X-C), the large language model gpt-5.4-2026-03-05 was used as the base model. The role of each agent was defined by a system prompt [prompts to be provided in the Appendix].

To ensure reproducibility, the version of each API-based model used in this study was fixed (snapshot-pinned). The snapshot dates of the models used as the base model and as the LLM-as-a-judge evaluator models were as follows: gpt-5.4-2026-03-05 as of March 5, 2026; Gemini 3.1 Pro Preview as of February 19, 2026; and Claude Opus 4.7 as of April 16, 2026.

As the reference knowledge base for RAG, we utilized a highly structured, multidimensional clinical knowledge base specifically engineered by expert physicians to align with standard clinical reasoning workflows. This knowledge base was derived from the *Evidence-based Clinical Practice Guidelines for CKD 2023* (published June 2023 by the Japanese Society of Nephrology). To transform flat guideline texts into an agent-interpretable cognitive format, physicians manually compiled and annotated each clinical recommendation across ten specialized programmatic fields:

1. Anatomical/Clinical Domain Classification: Taxonomy fields defining the clinical axis, including *Chapter* (e.g., Hypertension/CVD), *Subgroup* (e.g., Cardio-Renal Syndrome), *Item* (e.g., Goals of Anti-hypertensive Management), and *Target Disease* (e.g., Chronic Kidney Disease).

2. Clinical Intent Categorization: Mapping of textual evidence to fundamental diagnostic and therapeutic actions, labeling sections into actionable classes such as *Risk Factors*, *Comorbidities/Inter-disease Relations*, *Treatment/Management*, and *Basic Information*.

3. Context-Question-Answer Triplets: For each guideline recommendation, a verified clinical *Context* was extracted, which was then paired with an expert-formulated clinician *Question* (e.g., investigating precise blood pressure targets based on proteinuria status) and a definitive, comprehensive *Answer* synthesized directly from the guideline's consensus statements.

4. Evidence Grounding: Rigid association of each knowledge chunk with up to three primary peer-reviewed source literatures (e.g., large-scale clinical trials and epidemiological studies from *The Lancet* or *NEJM*) to ensure absolute traceability.

Through this methodology, the static guidelines were decomposed into precise, fact-contained clinical query-response vectors. For information retrieval during the inferencetime execution, text-embedding-3-large was deployed to vectorize these structured nodes into high-dimensional embeddings. At each multi-stage clinical step, the matching relevant knowledge chunks were dynamically retrieved via cosine similarity and appended to the context window of the initial reasoning agent. In the field of clinical medicine, it has already been reported that such clinician-structured RAG frameworks significantly mitigate hallucination cascades and improve both the factuality and safety of LLM outputs under severe uncertainty [3].

**Implementation**

All applications used in this study—the proposed architecture, control conditions 1 and 2, and the processing flows for RAG and each agent—were built on Dify, an LLM application development platform. Each agent (Agents X-A, X-B, and X-C), the inter-step cascade, and the RAG retrieval process were implemented using Dify's workflow functionality. The RAG knowledge base was constructed using Dify's built-in RAG (knowledge) feature, and text-embedding-3-large was used for the embeddings. All frontier models and the embedding model used in this study were accessed from Dify via each provider's API, and the version of each model was fixed. This unified the implementation of preprocessing, prompts, and data flow across conditions, thereby ensuring reproducibility.

**Comparison Conditions**

To isolate the contribution of each component of the proposed architecture, we compared the following three conditions. In all conditions, the cascade structure of the three steps—clinical assessment → diagnosis → treatment—was commonly retained, and each condition propagated the final output of each step within that condition to the subsequent step.

Condition 1 (LLM only) is a control representing base-model performance, in which each step was composed solely of the initial reasoning agent (Agent X-A) without using RAG. The inter-step connection was made via Output X-A.

Condition 2 (LLM + RAG) is a condition in which RAG (retrieval augmentation using text-embedding-3-large) was added to each step, with each step composed solely of Agent X-A; comparison with Condition 1 isolates the contribution of RAG. The inter-step connection was made via Output X-A.

Condition 3 (Proposed architecture) is the complete version, in which the full processing of Stages 1–3 (Agent X-A → X-B → X-C) is performed at each step; comparison with Condition 2 evaluates the contribution of the self-critique and refinement mechanism. The inter-step connection was made via the refined Output X-C.

### Evaluation Method

#### Human Evaluation by Expert Physicians

The outputs of each condition were independently evaluated by 4 expert physicians in nephrology. The evaluators were blinded to the condition assignment, and the presentation order of the conditions was randomized for each case. Following our previous study [7], clinical reasoning was decomposed into nine cognitive steps (Table 1), and the reasoning quality at each cognitive step was scored on a 5-point Likert scale (1 = inadequate to 5 = excellent) that took accuracy, clinical validity, and safety into account [scoring rubric provided in the Appendix]. Inter-rater agreement among the multiple evaluators was assessed using the intraclass correlation coefficient, ICC(2,k). Blinded, randomized evaluation by expert physicians has also been adopted in prior studies evaluating the clinical reasoning and management capabilities of LLMs [6,7]

#### LLM-as-a-Judge

To complement the human evaluation and extend the scoring, a multi-model LLM-as-a-judge evaluation was used in parallel. Although LLM-as-a-judge using a strong LLM as the evaluator has been reported to show high agreement with human evaluation, systematic biases such as position bias and verbosity bias have also been pointed out [8]. To reduce scoring bias attributable to any single model, three large language models with different foundations—namely gpt-5.4-2026-03-05, Gemini 3.1 Pro Preview, and claude-opus-4-7—were used as evaluators. Each evaluator model was provided with the reference standard for the case and the scoring rubric, and independently scored the outputs of the three stages (Stages 1–3) for each condition, using the same criteria and scale as the human evaluation (nine cognitive steps; 5-point Likert scale) [evaluation prompts provided in the Appendix].

It should be noted that one of the evaluator models, gpt-5.4-2026-03-05, is identical to the base model used to generate the outputs, raising a concern of self-preference bias. Therefore, as a sensitivity analysis, we confirmed that the main conclusions did not change when using the scores from only the two models excluding this model

**Figure S1. A structure of the baseline LLM model (LLM only: Model A)**

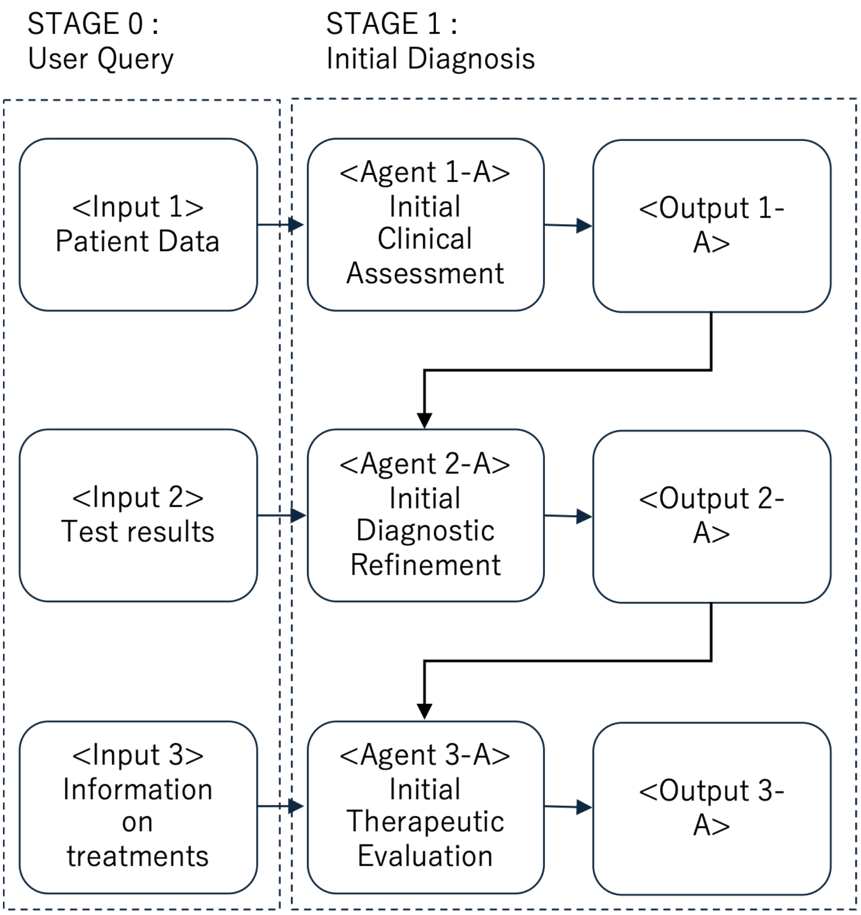

**Figure S2. A structure of the knowledge-grounded model (LLM + RAG: Model B)**

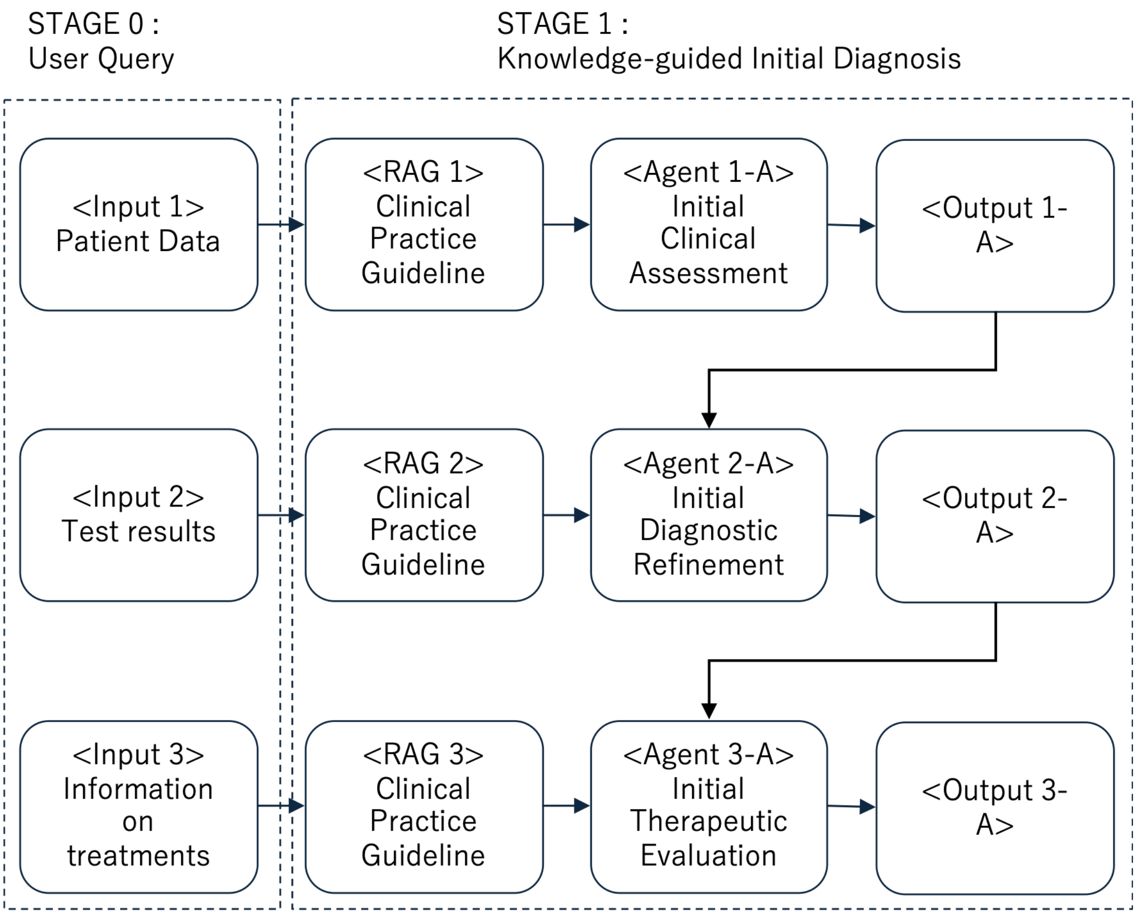

**Figure S3. Comparison of overall clinical reasoning performance among architectural models by AI evaluations.**

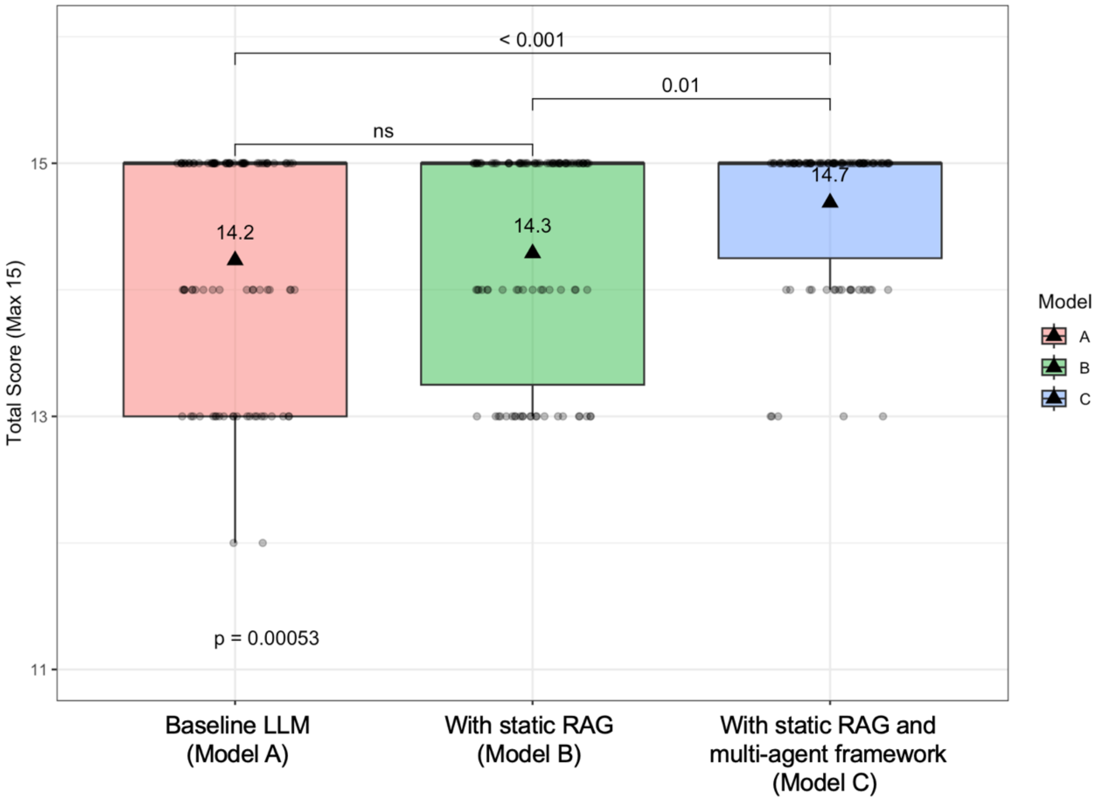

Box plots overlaid with jittered individual data points represent the distribution of total scores across the three architectural models, by the three different AI evaluation (maximum score = 15). Black triangles and text labels indicate the mean score for each model. Global p-values were derived from the Kruskal-Wallis test, and post-hoc pairwise comparisons were performed using the Wilcoxon rank-sum test with Bonferroni correction.

**Figure S4. Performance distribution per individual question step.**

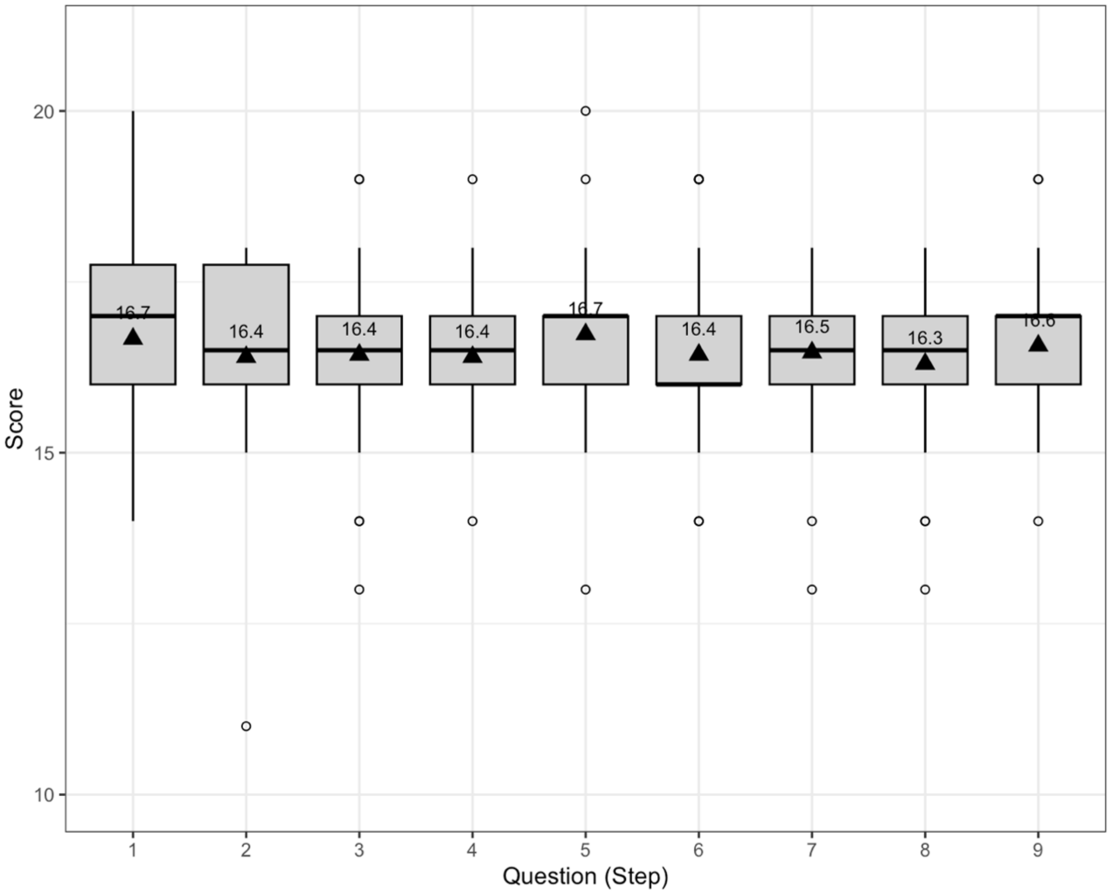

Box plots display the distribution of scores across specific questions (steps) regardless of the model evaluated, for the human expert evaluation. Triangles and text labels denote mean values, reflecting the inherent difficulty or vulnerability of each clinical reasoning step.

**Figure S5. Performance distribution per individual question step by AI evaluations.**

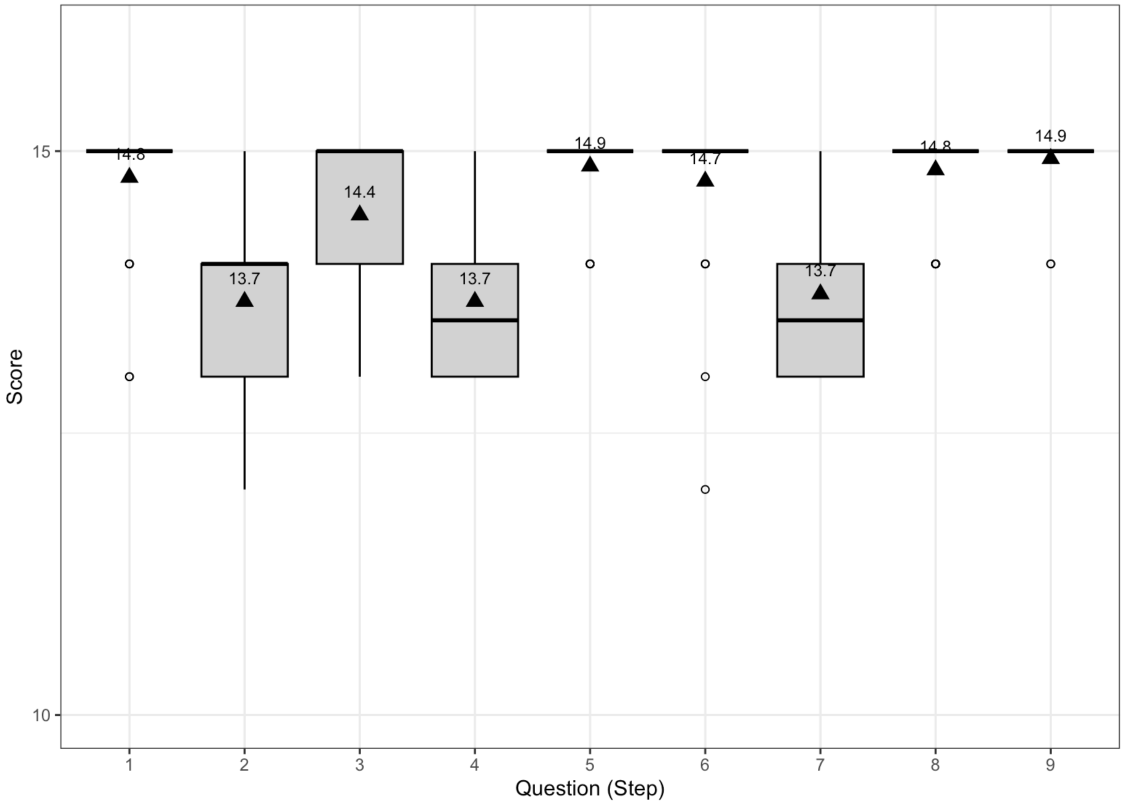

Box plots display the distribution of scores across specific questions (steps) regardless of the model evaluated, for the AI evaluation. Triangles and text labels denote mean values, reflecting the inherent difficulty or vulnerability of each clinical reasoning step.

**Figure S6. Heatmap of average scores by question step and paradigm by AI evaluations.**

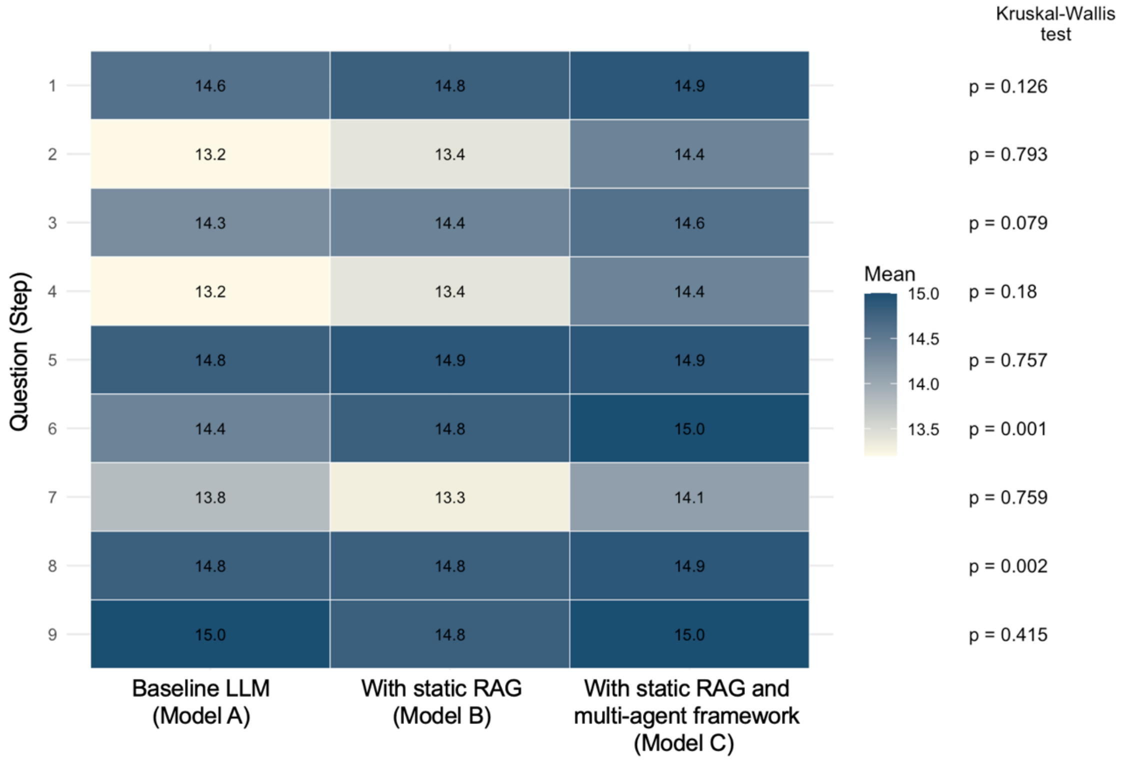

The matrix visualizes the mean score for each clinical reasoning step across the three architectural models for the AI evaluation, with darker colors representing superior performance. The right-hand panels display the corresponding p-values from question-specific Kruskal-Wallis tests, indicating whether performance differences between models were statistically significant at that specific step.

**Supplementary Table 1. Prompts and Evaluation Criteria for the Phased Assessment of Clinical Reasoning**

| Question | Prompt to the LLM | Evaluation Criteria |
|---|---|---|
| **Phase I: Before the Consultation** | | |
| Q1 | How would you summarize this patient as a medical problem? Please explain, focusing on the key points. | Accurately and concisely summarizes the patient's problem, incorporating age, gender, timeline, and key symptom characteristics. |
| Q2 | Hearing this chief complaint, what differential diagnoses do you consider? Please list three. What is the most likely? What must not be missed? Please also explain your reasoning. | Provides a broad and prioritized list of differential diagnoses, considering frequency and severity, and explains the rationale. |
| Q3 | To verify the hypotheses you've made, what additional questions would you ask or what physical exams would you perform? What do you expect to clarify with that history-taking or examination? | Proposes focused history or physical exam steps with clear purpose tied to confirming or ruling out differential diagnoses. |
| Q4 | To narrow down the diagnosis, what tests do you plan to order? Please tell me the purpose for ordering those tests and the expected results. | Suggests a logical test plan aligned with diagnostic refinement, considering cost-effectiveness and invasiveness. |

| Phase II: After Test Results | | |
|---|---|---|
| Q5 | How do you interpret these test results? Please explain the points of particular interest and what they mean in the context of this patient's condition. | Interprets results in clinical context, explaining their medical significance rather than just stating abnormalities. |
| Q6 | Based on all the information, how has your diagnostic thinking changed? What is the most likely diagnosis (Working | Revises and reprioritizes differential diagnoses appropriately in light of new information, and explains reasoning. |

| | | |
|---|---|---|
| | Diagnosis) at this point? | |
| Q7 | What kind of treatment plan would you create for this patient? Please also explain the rationale for your chosen treatment and other possible options. | Proposes a treatment plan considering both evidence-based medicine (EBM) and patient-specific factors, including justification and alternatives. |

| **Phase III: During Treatment Course** | | |
|---|---|---|
| Q8 | How will you monitor the effectiveness and safety of the treatment? What are the specific risks to watch out for, and what are your concrete countermeasures for them? | Identifies indicators for treatment response and potential side effects or complications, and proposes a monitoring and response plan. |
| Q9 | If the patient's condition does not improve as expected or suddenly changes, how would you think and act? | Describes a process for reassessment of diagnosis or treatment plan, including rapid response and flexible decision-making. |
| The evaluation was conducted using a step-by-step prompting approach to simulate a realistic, temporally structured reasoning process. The clinical reasoning process for each case was divided into three phases as shown in the table. Initially, only the 'Phase I' section of the case was provided to each model to answer the corresponding questions (Q1-Q4); information from Phase II and III was withheld. Upon completion of Phase I, the 'Phase II' section was presented, and the relevant questions (Q5-Q7) were asked, still without disclosing Phase III information. Finally, the 'Phase III' section was provided for the models to answer the last set of questions (Q8-Q9). The outputs from each phase were then evaluated and scored by four nephrology experts on a 5-point scale: 5 = perfect and highly informative, 4 = good and mostly correct, 3 = acceptable, 2 = incomplete or partially incorrect, and 1 = incorrect or entirely unhelpful. The scores for each model were aggregated to evaluate the differences in total scores across the models. | | |

**Supplementary Table 2. Responses from the Three Architectural Models to Clinical Reasoning Questions Across Nine Cases**

| | |
|---|---|
| **Case 1: A case of PLA2R-positive membranous nephropathy with subsequent development of IgG4-related disease** [9] | |
| **Phase I – Q1: Summarizing medical problems** | |
| Baseline LLM (Model A) | # 1. Problem representation<br>A **60-year-old man** with **new-onset edema and frothy urine for about 1 month**, found to have **proteinuria on routine health screening**, with **no known prior medical history** and **no previous abnormal urinalysis**, presenting with a picture most suggestive of **recently developed glomerular disease causing significant proteinuria, possibly nephrotic syndrome**.<br><br>**Key points**<br>- **Age/sex**: 60-year-old man<br>- **Time course**: subacute, about 1 month<br>- **Main features**: pitting leg edema, frothy urine, newly detected proteinuria<br>- **Clinical implication**: these symptoms strongly suggest **heavy proteinuria**, likely of **glomerular origin**<br>- **Syndromic framing**: concern for **nephrotic syndrome** (proteinuria, edema; hypoalbuminemia/hyperlipidemia not yet confirmed) |
| With Static RAG (Model B) | 1. **Problem representation**<br><br>A **60-year-old man** with **about 1 month** of **pitting lower-extremity edema** and **frothy urine**, newly found to have **proteinuria** on routine health screening, with **no known relevant past history**.<br>This presentation strongly suggests a **glomerular cause of significant proteinuria, possibly nephrotic syndrome**, given the edema and frothy urine. The key clinical issue is to determine whether this is **primary glomerular disease** or **secondary kidney involvement** from a systemic disorder. |
| With Static RAG and Mult-agent framework (Model C) | # 1. Problem representation<br>A **60-year-old man** with **subacute (~1 month) bilateral pitting leg edema and frothy urine**, found for the **first time** to have **proteinuria on routine health screening**, has a presentation most concerning for **suspected nephrotic syndrome or nephrotic-range proteinuric glomerular disease**, which still requires confirmation with **protein quantification and serum albumin**.<br><br>**Key features**<br>- **Age/sex:** 60-year-old man<br>- **Time course:** subacute, about 1 month<br>- **Main findings:** edema, frothy urine, newly detected proteinuria<br>- **Syndromic implication:** likely significant **glomerular protein loss**<br>- **Important caveat:** nephrotic syndrome is **suspected, not yet confirmed**, until heavy proteinuria, hypoalbuminemia, and supportive labs are documented |
| **Phase I – Q2: Differential diagnoses and rationale** | |
| Baseline LLM (Model A) | ## 2. Differential diagnosis<br>### Top 3 differential diagnoses<br>1. **Membranous nephropathy**<br>2. **Minimal change disease** |

| | |
|---|---|
| | 3. **Diabetic kidney disease or other secondary glomerular disease**<br>(more broadly, secondary nephrotic syndrome such as amyloidosis, lupus, infection-related, malignancy-associated)<br><br>### Most likely diagnosis<br>**Membranous nephropathy**<br><br>**Reasoning**<br>- In a **60-year-old adult** with **new nephrotic-range proteinuria symptoms**, membranous nephropathy is a common cause of adult nephrotic syndrome.<br>- The presentation is **subacute** and dominated by **edema/frothy urine**, without information suggesting nephritic syndrome (e.g., gross hematuria, rapidly progressive course).<br>- Older age also raises the possibility of **secondary membranous nephropathy**, including malignancy-associated disease.<br><br>### Must not be missed<br>**Secondary nephrotic syndrome due to malignancy or systemic disease**, and **rapidly progressive glomerulonephritis if nephritic features are present**<br><br>**Reasoning**<br>- In older adults, newly developed glomerular proteinuria can be associated with **solid tumors, infections, autoimmune disease, or amyloidosis**.<br>- Although the current story favors nephrotic syndrome, missing a **nephritic/RPGN process** would be dangerous because it can cause rapid irreversible kidney failure.<br>- Severe nephrotic syndrome also carries risk of **venous thromboembolism**, especially in membranous nephropathy. |
| With Static RAG (Model B) | 2. **Differential diagnosis**<br><br>**Top 3 differential diagnoses**<br>1. **Membranous nephropathy**<br>- Most likely in a 60-year-old adult with new-onset significant proteinuria/possible nephrotic syndrome.<br>- Common cause of nephrotic syndrome in older adults.<br><br>2. **Minimal change disease or focal segmental glomerulosclerosis (FSGS)**<br>- Both can present with edema and heavy proteinuria.<br>- FSGS is especially important in adults; minimal change disease is less common at this age but still possible.<br><br>3. **Diabetic kidney disease / secondary glomerular disease (including amyloidosis, lupus, infection-related, malignancy-associated disease)**<br>- Secondary causes must be considered in any adult with new nephrotic-range proteinuria, especially at age 60.<br>- Amyloidosis or malignancy-associated membranous nephropathy are important possibilities in older adults.<br><br>**Most likely:** **Membranous nephropathy**<br>- Age and presentation fit well with adult-onset nephrotic syndrome.<br><br>**Must not be missed:** **Secondary nephrotic syndrome due to malignancy or systemic disease**, and **rapidly progressive glomerulonephritis if hematuria/renal dysfunction is present** |

| | |
|---|---|
| | - Because management and prognosis change substantially.<br>- In older adults, membranous nephropathy can be associated with solid malignancy.<br>- If renal function is declining or active urine sediment is present, urgent evaluation is needed. |
| With Static RAG and Mult-agent framework (Model C) | # 2. Prioritized differential diagnosis<br><br>### Top differential diagnoses<br>1. **Membranous nephropathy**<br>2. **FSGS or minimal change disease**<br>3. **Amyloidosis or secondary nephrotic syndrome**<br> - including **malignancy-associated membranous nephropathy** and infection-related causes such as **HBV, HCV, HIV**<br><br>### Other important considerations<br>- **Diabetic kidney disease**<br> - possible but less strongly supported at present because no diabetes history is given<br>- **Lupus nephritis or other immune-complex glomerulopathy**<br> - less likely without systemic features, but worth screening if indicated<br>- **IgA nephropathy or other glomerulonephritides**<br> - less typical if edema and isolated heavy proteinuria predominate<br><br>### Most likely diagnosis<br>**Membranous nephropathy**<br><br>**Why**<br>- It is a common cause of **nephrotic syndrome in older adults**<br>- The presentation of **new edema + frothy urine + proteinuria** is very compatible with a **nephrotic-pattern glomerular disease**<br>- In this age group, both **primary membranous nephropathy** and **secondary membranous disease** should be considered<br><br>### Must not miss<br>**Secondary nephrotic syndrome due to malignancy, monoclonal disease, or chronic infection**<br><br>**Examples**<br>- **Malignancy-associated membranous nephropathy**<br>- **Amyloidosis / monoclonal gammopathy**<br>- **HBV-, HCV-, or HIV-associated glomerular disease**<br><br>**Why**<br>- In older adults, newly recognized nephrotic syndrome may be the first sign of a serious underlying systemic disorder<br>- Missing these would delay disease-specific treatment |
| **Phase I – Q3: Necessary physical examinations and reasons** | |
| Baseline LLM (Model A) | # 3. Additional history and physical examination<br>### Additional questions<br>1. **How severe is the edema? Any weight gain, facial edema, abdominal distension, dyspnea?**<br> - To assess degree of volume retention and possible nephrotic syndrome complications. |

| | |
|---|---|
| | 2. **Any hematuria, dark urine, oliguria, hypertension symptoms?**<br>- To look for nephritic features suggesting other glomerulonephritides.<br><br>3. **Any history of diabetes, hypertension, autoimmune symptoms, chronic infection, hepatitis risk, or malignancy symptoms (weight loss, anorexia, smoking history)?**<br>- To evaluate secondary causes.<br><br>4. **Medication history**: NSAIDs, herbal drugs, recent antibiotics, immune checkpoint inhibitors<br>- Drug-induced nephrotic syndrome is possible, especially minimal change disease or membranous nephropathy.<br><br>5. **Thrombotic symptoms**: unilateral leg pain/swelling, chest pain, dyspnea<br>- Nephrotic syndrome increases thromboembolic risk<br><br>### Physical examination<br>1. **Blood pressure, weight, volume status**<br>- To quantify severity and assess renal/vascular involvement.<br><br>2. **Distribution of edema**: legs, sacrum, periorbital area, ascites, pleural effusion signs<br>- To assess nephrotic syndrome severity.<br><br>3. **Cardiopulmonary exam**<br>- To distinguish renal edema from heart failure and evaluate fluid overload.<br><br>4. **Skin/joint exam**<br>- Rash, purpura, arthritis may suggest systemic autoimmune disease or vasculitis.<br><br>5. **Abdominal/lymph node exam**<br>- To screen for malignancy or chronic systemic disease. |
| With Static RAG (Model B) | 3. **Additional history and physical examination**<br><br>**History**<br>- **Urine symptoms:** amount of urine, gross hematuria, duration/progression of frothy urine<br>- To assess severity and whether nephritic features coexist.<br>- **Edema history:** facial edema, weight gain, ascites, dyspnea<br>- To evaluate volume status and severity of hypoalbuminemia/fluid retention.<br>- **Systemic symptoms:** fever, rash, arthralgia, fatigue, neuropathy<br>- To screen for lupus, vasculitis, amyloidosis, infection-related disease.<br>- **Diabetes/hypertension history**<br>- To assess common secondary kidney diseases.<br>- **Drug exposure:** NSAIDs, herbal medicines, antibiotics<br>- Important causes of minimal change disease or secondary membranous nephropathy.<br>- **Infection risk/history:** hepatitis B/C, HIV<br>- Secondary causes of glomerular disease.<br>- **Malignancy symptoms:** weight loss, anorexia, cough, gastrointestinal symptoms<br>- Important given age and possible membranous nephropathy.<br>- **Thrombotic symptoms:** leg pain, chest pain, dyspnea<br>- Nephrotic syndrome increases thrombosis risk. |

| | |
|---|---|
|  | **Physical examination**<br>- **Blood pressure**<br> - Helps characterize glomerular disease severity and chronicity.<br>- **Assessment of edema** (legs, sacrum, eyelids), **jugular venous pressure**, **lung exam**, **ascites**<br> - To assess fluid overload.<br>- **Skin/joint exam** for rash, purpura, synovitis<br> - To look for systemic autoimmune disease/vasculitis.<br>- **Cardiopulmonary and abdominal exam**<br> - To assess heart failure, liver disease, ascites, masses.<br>- **General exam for malignancy clues** such as lymphadenopathy/cachexia<br> - To identify secondary causes. |
| With Static RAG and Mult-agent framework (Model C) | # 3. Additional history and physical examination<br><br>### Focused history<br>1. **Edema severity and progression**<br> - Weight gain, facial swelling, scrotal edema, abdominal distension, dyspnea<br> - Helps assess severity of suspected nephrotic syndrome and fluid overload<br><br>2. **Urinary symptoms**<br> - Gross hematuria, decreased urine output, nocturia<br> - Helps distinguish nephrotic from nephritic or mixed presentations<br><br>3. **History of hypertension or prior kidney disease**<br> - Prior elevated blood pressure, prior abnormal creatinine, earlier urinary abnormalities<br> - Helps identify chronic kidney disease or long-standing renal disease<br><br>4. **Systemic symptoms**<br> - Rash, arthralgia, oral ulcers, photosensitivity, neuropathy, fevers<br> - Screens for autoimmune disease, vasculitis, amyloidosis, or systemic illness<br><br>5. **Diabetes history**<br> - Known diabetes, polyuria, polydipsia, visual complications<br> - Evaluates diabetic kidney disease, though currently not strongly supported<br><br>6. **Infection and immunologic triggers**<br> - Hepatitis risk, HIV risk, recent infections, recent vaccinations<br> - Relevant to secondary glomerular diseases and occasional immune triggers<br><br>7. **Medication and exposure review**<br> - NSAIDs, antibiotics, herbal medications, lithium, gold, penicillamine<br> - Important because some drugs are linked to membranous nephropathy or minimal change disease<br><br>8. **Malignancy review**<br> - Weight loss, anorexia, cough, smoking history, GI symptoms, cancer screening history<br> - Important in an older adult with possible secondary membranous nephropathy<br><br>9. **Thrombotic symptoms**<br> - Unilateral leg pain/swelling, pleuritic chest pain, dyspnea<br> - Nephrotic syndrome increases risk of venous thromboembolism |

| | |
|---|---|
| | ### Physical examination<br>1. **Vital signs, especially blood pressure**<br>- Helps assess renal disease severity and possible chronic hypertension<br><br>2. **Edema assessment**<br>- Distribution and severity of peripheral edema; look for anasarca<br>- Helps estimate severity of hypoalbuminemia/fluid retention<br><br>3. **Volume overload assessment**<br>- JVP, lung crackles, pleural effusion, ascites<br>- Distinguishes systemic fluid overload from isolated peripheral edema<br><br>4. **Examination for DVT**<br>- Asymmetric leg swelling, calf tenderness<br>- Important because nephrotic syndrome is prothrombotic<br><br>5. **Skin, joints, and neurologic exam**<br>- Rash, purpura, synovitis, neuropathy<br>- Suggests autoimmune disease, vasculitis, or amyloidosis<br><br>6. **Funduscopic exam**<br>- Retinopathy may support diabetes or chronic hypertension<br><br>7. **Cardiac, liver, lymph node, and abdominal exam**<br>- Looks for alternative causes of edema and signs of malignancy or systemic disease |
| **Phase I – Q4: Plan for investigations/tests** | |
| Baseline LLM (Model A) | # 4. Tests to narrow the diagnosis<br>### Initial laboratory tests<br>1. **Urinalysis with urine sediment**<br>- **Purpose**: confirm proteinuria and look for hematuria/casts.<br>- **Expected result**: heavy proteinuria; bland sediment would support nephrotic syndrome, while RBCs/casts would suggest nephritic disease.<br><br>2. **Quantification of proteinuria**: spot urine protein-creatinine ratio or albumin-creatinine ratio, or 24-hour urine protein<br>- **Purpose**: determine severity and whether nephrotic range.<br>- **Expected result**: likely **nephrotic-range proteinuria**.<br><br>3. **Serum creatinine/eGFR, BUN, electrolytes**<br>- **Purpose**: assess kidney function and urgency.<br>- **Expected result**: may be normal or mildly impaired early on.<br><br>4. **Serum albumin and total protein**<br>- **Purpose**: confirm nephrotic syndrome.<br>- **Expected result**: likely **hypoalbuminemia**.<br><br>5. **Lipid profile**<br>- **Purpose**: nephrotic syndrome often causes hyperlipidemia.<br>- **Expected result**: elevated cholesterol/triglycerides.<br><br>### Tests for cause<br>6. **HbA1c/glucose** |

| | |
|---|---|
| | - **Purpose**: assess for diabetic kidney disease.<br>- **Expected result**: may be normal if no diabetes.<br><br>7. **Immunologic and serologic workup**: ANA, complements, hepatitis B/C, HIV, serum/urine protein electrophoresis ± immunofixation, serum free light chains<br>- **Purpose**: evaluate lupus, infection-related disease, amyloidosis, monoclonal gammopathy.<br>- **Expected result**: often negative in primary membranous nephropathy, but important to exclude secondary causes.<br><br>8. **Anti-PLA2R antibody**<br>- **Purpose**: supports primary membranous nephropathy.<br>- **Expected result**: may be positive if primary membranous nephropathy.<br><br>### Imaging and further evaluation<br>9. **Renal ultrasound**<br>- **Purpose**: assess kidney size and exclude structural disease.<br>- **Expected result**: usually normal-sized kidneys in glomerular disease.<br><br>10. **Age-appropriate malignancy screening**<br>- **Purpose**: older adult with possible membranous nephropathy may have malignancy-associated disease.<br>- **Expected result**: usually negative, but must be checked.<br><br>11. **Kidney biopsy**<br>- **Purpose**: definitive diagnosis of glomerular disease in an adult with new significant proteinuria/nephrotic syndrome.<br>- **Expected result**: if membranous nephropathy, subepithelial immune deposits and GBM thickening; biopsy also distinguishes minimal change disease, FSGS, amyloidosis, etc.<br><br>Overall, the diagnostic strategy should first **confirm nephrotic syndrome and assess kidney function**, then **differentiate primary from secondary glomerular disease**, with **kidney biopsy as the key test** in most adults with newly developed significant proteinuria. |
| With Static RAG (Model B) | 4. **Tests to order**<br><br>**Initial laboratory evaluation**<br>- **Urinalysis with sediment**<br>- Purpose: confirm proteinuria and check for hematuria/casts.<br>- Expected: heavy proteinuria; sediment may be bland in nephrotic syndrome.<br>- **Spot urine protein-to-creatinine ratio or albumin-to-creatinine ratio** (or 24-hour urine protein if needed)<br>- Purpose: quantify proteinuria.<br>- Expected: possibly nephrotic-range proteinuria.<br>- **Serum creatinine, eGFR, electrolytes**<br>- Purpose: assess kidney function and urgency.<br>- Expected: may be normal or impaired depending on disease severity.<br>- **Serum albumin, total protein, lipid profile**<br>- Purpose: evaluate for nephrotic syndrome.<br>- Expected: hypoalbuminemia and hyperlipidemia if nephrotic syndrome is present.<br>- **CBC, liver panel**<br>- Purpose: assess systemic illness and alternative causes of edema. |

| | |
|---|---|
|  | **Evaluation for secondary causes**<br>- **HbA1c/glucose**<br> - Purpose: screen for diabetic kidney disease.<br>- **ANA, complements (C3/C4), anti-dsDNA if indicated**<br> - Purpose: evaluate autoimmune causes.<br>- **Hepatitis B, hepatitis C, HIV serologies**<br> - Purpose: identify infection-associated glomerular disease.<br>- **SPEP/UPEP with immunofixation and serum free light chains**<br> - Purpose: screen for monoclonal gammopathy/amyloidosis in an older adult with new proteinuria.<br>- **Anti-PLA2R antibody**<br> - Purpose: support primary membranous nephropathy.<br> - Expected: may be positive if primary membranous nephropathy.<br><br>**Imaging**<br>- **Renal ultrasound**<br> - Purpose: assess kidney size, chronicity, and exclude structural disease.<br> - Expected: usually normal-sized kidneys in primary glomerular disease.<br><br>**Definitive diagnostic test**<br>- **Kidney biopsy**<br> - Purpose: establish the exact glomerular diagnosis and distinguish primary from secondary patterns.<br> - Expected: particularly important in an adult with new likely nephrotic syndrome and no obvious cause.<br><br>**Additional age-appropriate malignancy screening**<br>- Based on symptoms and standard screening (e.g., chest imaging, colon cancer screening if not up to date)<br> - Purpose: evaluate for malignancy-associated membranous nephropathy or other secondary causes. |
| With Static RAG and Mult-agent framework (Model C) | # 4. Testing plan to narrow the diagnosis<br><br>### A. First, confirm the syndrome<br>1. **Urinalysis with microscopy/sediment**<br> - Confirms proteinuria and evaluates for hematuria, casts, oval fat bodies<br> - A bland sediment would support a nephrotic-pattern disorder<br><br>2. **Quantify proteinuria**<br> - **Spot UPCR or UACR** or **24-hour urine protein**<br> - Determines whether proteinuria is in the nephrotic range<br><br>3. **Serum chemistry including creatinine/eGFR and serum albumin**<br> - Establishes kidney function and confirms hypoalbuminemia<br> - **Serum albumin is central** to confirming nephrotic syndrome<br><br>4. **Lipid profile**<br> - Hyperlipidemia supports nephrotic syndrome<br><br>### B. Then evaluate the cause<br>5. **PLA2R antibody**<br> - Supports primary membranous nephropathy if positive |

6. **HBsAg, HCV testing, HIV testing**
 - Screens for important secondary infectious causes

7. **ANA, anti-dsDNA, complement levels**
 - Evaluates for lupus or immune-complex disease

8. **SPEP/UPEP with immunofixation and serum free light chains**
 - Screens for monoclonal gammopathy and amyloidosis

9. **HbA1c or fasting glucose**
 - Assesses for occult diabetes, but is lower priority than the above based on current information

#### C. Supportive/structural assessment
10. **Renal ultrasound**
 - Assesses kidney size and excludes structural disease

11. **Consider coagulation/thrombotic risk assessment if nephrotic syndrome is confirmed**
 - Clinically relevant because of increased VTE risk, especially with severe hypoalbuminemia

12. **Age-appropriate malignancy screening guided by history/exam**
 - Important if membranous nephropathy or amyloidosis is suspected

#### D. Definitive test
13. **Kidney biopsy**
 - Usually indicated in an adult with new significant proteinuria and suspected glomerular disease unless a clear alternative explanation emerges
 - Can distinguish membranous nephropathy, FSGS, minimal change disease, amyloidosis, diabetic nephropathy, and others

#### Most efficient diagnostic sequence
1. **Confirm nephrotic syndrome**: urine microscopy, UPCR/UACR or 24-hour protein, **serum albumin**, creatinine/eGFR, lipids
2. **Screen for major causes**: PLA2R, hepatitis/HIV tests, ANA/complements, SPEP/UPEP/free light chains, HbA1c
3. **Proceed to kidney biopsy** if no obvious alternative explanation is established

---

### Changes from the initial version
1. **Problem representation revised**
 - Changed from implying likely nephrotic syndrome to explicitly stating **“suspected nephrotic syndrome”** pending confirmation with **protein quantification and serum albumin**.

2. **Differential diagnosis reprioritized**
 - **Diabetic kidney disease was moved lower**
 - **FSGS/minimal change disease** and **amyloidosis/secondary causes** were given greater prominence
 - **Lupus nephritis** was de-emphasized, though still considered in appropriate screening

| | |
|---|---|
|  | 3. **History and exam refined**<br> - Added **hypertension history**, **prior kidney disease**, **recent infections/vaccinations**, and a more structured **drug review**<br> - Added exam emphasis on **DVT**, **pleural effusion**, and signs of **cardiac/liver/systemic disease**<br><br>4. **Testing plan reorganized**<br> - Now clearly separated into:<br> - **confirmation of nephrotic syndrome**<br> - **etiologic workup**<br> - **definitive biopsy**<br> - **Serum albumin** was made explicit as central<br> - Added **urine microscopy details** and **coagulation/thrombotic risk relevance**<br><br>If you want, I can also convert this into a **shorter ward-style assessment and plan** or a **teaching-case discussion format**. |
| **Phase II – Q5: Interpreting test results** | |
| Baseline LLM (Model A) | # 5. Interpretation of the test results<br>- **Serum albumin 2.5 g/dL + UPCR 5.0 g/gCr + edema + frothy urine**: This confirms **nephrotic syndrome**. The hypoalbuminemia explains the peripheral edema, and the heavy proteinuria is consistent with glomerular injury.<br>- **LDL cholesterol 216 mg/dL**: Marked hypercholesterolemia is a typical metabolic consequence of nephrotic syndrome and further supports the syndrome clinically.<br>- **Creatinine 0.65 mg/dL**: Renal function is **preserved at this stage**, which is favorable prognostically and suggests the disease is currently limited mainly to the glomerular filtration barrier rather than causing advanced chronic damage.<br>- **No hematuria**: A **bland urine sediment** supports a nephrotic rather than nephritic process, making proliferative glomerulonephritis less likely.<br>- **Kidney ultrasound without renal mass**: This does not prove absence of secondary causes overall, but it excludes an obvious renal structural lesion as the cause of proteinuria.<br>- **Kidney biopsy findings**:<br> - **Light microscopy**: capillary wall deposits are compatible with **membranous nephropathy (MN)**.<br> - **IF: granular IgG and C3 along capillary walls**: this is the classic immune-complex pattern of MN.<br> - **IgG4-predominant staining**: strongly favors **primary MN** over many secondary forms.<br> - **PLA2R positivity**: this is the most important finding; it strongly supports **PLA2R-associated primary membranous nephropathy**.<br> - **No significant tubulointerstitial fibrosis/inflammation**: suggests limited chronic irreversible damage and a better chance of response to treatment.<br> - **EM unavailable**: this is a limitation, but the diagnosis remains well supported by LM + IF + PLA2R positivity. |
| With Static RAG (Model B) | **5. Interpretation of the test results**<br><br>- **Albumin 2.5 g/dL + UPCR 5.0 g/gCr + edema + frothy urine**: This confirms **nephrotic syndrome**. The low albumin explains the edema, and the heavy proteinuria indicates a glomerular permeability disorder. |

| | |
|---|---|
| | - **LDL 216 mg/dL**: Marked hypercholesterolemia is typical of nephrotic syndrome and supports the clinical severity of protein loss.<br>- **Creatinine 0.65 mg/dL**: Kidney function is currently **preserved**, which is favorable prognostically and relevant when choosing treatment intensity.<br>- **No hematuria**: A **bland urine sediment** is more consistent with a nephrotic process such as membranous nephropathy than with inflammatory/nephritic glomerulonephritis.<br>- **Ultrasound without renal mass**: Helps exclude an obvious structural renal cause; it does not rule out all secondary causes, but supports a primary glomerular process.<br>- **Kidney biopsy findings**:<br> - **Light microscopy** showing capillary wall deposits is compatible with **membranous nephropathy (MN)**.<br> - **IF: granular IgG and C3 along glomerular capillary walls** is the classic immune-complex pattern of MN.<br> - **IgG4-predominant deposition** and **PLA2R positivity** are particularly important because they strongly favor **primary (autoimmune) MN** rather than secondary MN.<br> - **Minimal tubulointerstitial damage** suggests limited chronic injury, which generally predicts better renal outcome.<br> - **EM unavailable** is a limitation, but the diagnosis is still well supported by LM + IF + PLA2R positivity. |
| With Static RAG and Mult-agent framework (Model C) | # 5. Interpretation of the test results<br>These results **confirm nephrotic syndrome** and **strongly support membranous nephropathy (MN), specifically PLA2R-associated disease most consistent with primary MN**.<br><br>- **Serum albumin 2.5 g/dL**: This degree of hypoalbuminemia explains his **pitting edema** and is a central feature of nephrotic syndrome.<br>- **UPCR 5.0 g/gCr**: This indicates **nephrotic-range proteinuria**, matching his **frothy urine** and supporting a glomerular cause of heavy protein loss.<br>- **LDL cholesterol 216 mg/dL**: Marked hypercholesterolemia is typical of nephrotic syndrome and further supports the syndrome clinically.<br>- **Creatinine 0.65 mg/dL**: This suggests **preserved kidney function at present**, although renal function should ideally be interpreted with **eGFR and overall clinical context**.<br>- **No hematuria**: A **bland urine sediment** is compatible with MN and makes inflammatory/nephritic glomerulonephritides less likely.<br>- **Ultrasound without renal mass**: This lowers concern for an obvious renal structural lesion, but it does **not exclude secondary causes** such as malignancy elsewhere or infection-related disease.<br>- **Kidney biopsy**:<br> - **Light microscopy** showing capillary wall deposits is compatible with MN.<br> - **IF showing diffuse granular IgG and C3 along glomerular capillary walls** is classic for MN.<br> - **IgG4-predominant deposition** favors **primary** over many secondary forms.<br> - **PLA2R positivity** strongly supports **primary PLA2R-associated MN**.<br> - **No significant tubulointerstitial fibrosis or inflammation** suggests little chronic damage so far.<br><br>- **EM unavailable**: This is a limitation because EM is useful for confirming subepithelial deposits and assessing ultrastructural features. However, the diagnosis of **MN is still well supported pathologically** by the available LM and IF findings |

| | |
|---|---|
| | plus **PLA2R positivity**.<br><br>**Overall interpretation:**<br>This patient has **nephrotic syndrome** due to **biopsy-proven membranous nephropathy**, with findings **strongly favoring primary PLA2R-associated MN**, preserved renal function, and minimal chronic tubulointerstitial injury. Secondary causes are now less likely, but they are **not completely excluded** and should still be assessed as clinically indicated. |
| **Phase II – Q6: Reconsidering the differential diagnosis** | |
| Baseline LLM (Model A) | # 6. Updated diagnostic thinking / Working diagnosis<br>The differential diagnosis narrows substantially. Before biopsy, causes of nephrotic syndrome in this age group included MN, minimal change disease, FSGS, and secondary glomerular diseases. After biopsy:<br><br>1. **Primary membranous nephropathy** — now overwhelmingly most likely<br>2. Secondary MN — much less likely given **PLA2R positivity** and **IgG4 predominance**<br>3. Other nephrotic syndromes (minimal change disease, FSGS, amyloidosis) — unlikely based on pathology<br><br>**Working Diagnosis:**<br>**Primary PLA2R-associated membranous nephropathy presenting with nephrotic syndrome, with preserved kidney function.** |
| With Static RAG (Model B) | **6. Updated diagnostic thinking / Working diagnosis**<br><br>The differential diagnosis is now substantially narrowed.<br>- **Primary membranous nephropathy** becomes overwhelmingly the leading diagnosis because of:<br> - nephrotic syndrome,<br> - bland urine sediment,<br> - biopsy pattern of MN,<br> - **PLA2R positivity**,<br> - **IgG4-dominant immune deposits**.<br><br>Other causes such as minimal change disease, FSGS, diabetic kidney disease, and secondary MN are now much less likely. Secondary MN is not completely impossible, but the biopsy pattern strongly shifts probability toward **primary PLA2R-associated MN**.<br><br>**Working Diagnosis:**<br>**Primary (PLA2R-associated) membranous nephropathy presenting with nephrotic syndrome, with preserved renal function.** |
| With Static RAG and Mult-agent framework (Model C) | ### 6. Updated diagnostic thinking / Working diagnosis<br>The differential diagnosis is now substantially narrowed.<br><br>## **Working diagnosis: Membranous nephropathy, PLA2R-positive, most consistent with primary MN, presenting with nephrotic syndrome**<br><br>### Why this is now the most probable diagnosis<br>- **Nephrotic syndrome is established** by:<br> - **nephrotic-range proteinuria** (UPCR 5.0 g/gCr) |

| | |
|---|---|
| | - **hypoalbuminemia** (2.5 g/dL)<br>- **hyperlipidemia** (LDL 216 mg/dL)<br>- edema and frothy urine on history<br>- The **decisive findings are the biopsy and immunopathology**:<br>- capillary wall deposits on light microscopy<br>- granular **IgG and C3** along the capillary walls on IF<br>- **IgG4 predominance**<br>- **PLA2R positivity**<br>- These findings are highly characteristic of **primary MN**, rather than FSGS, minimal change disease, diabetic kidney disease, amyloidosis, or lupus nephritis.<br><br>### Diagnostic refinement<br>- The diagnosis is best framed as **pathologically established MN with features favoring primary disease**.<br>- **Absence of hematuria** and **preserved kidney function** support the pattern but are **not the decisive features**.<br>- Because **EM was unavailable**, and because even PLA2R-positive disease does not absolutely eliminate secondary causes, **appropriate screening for secondary causes should still be completed as clinically indicated**, especially malignancy and chronic infection in a 60-year-old patient.<br><br>### Changes from the initial diagnostic impression<br>- The earlier broad differential has now narrowed to a **biopsy-based diagnosis of MN**.<br>- **Primary PLA2R-associated MN** has moved from a leading possibility to the **clear leading diagnosis**.<br>- Competing diagnoses such as **FSGS, minimal change disease, amyloidosis, diabetic kidney disease, and lupus nephritis** are now much less likely because the **biopsy/IF pattern is specific for MN**.<br>- Diagnostic certainty is stronger than before, but still expressed as **“strongly supports primary MN”** rather than absolute certainty because **EM was not available** and secondary causes still merit appropriate review. |
| **Phase II – Q7: Treatment planning** | |
| Baseline LLM (Model A) | ### 7. Treatment plan<br>#### Main treatment plan<br>1. **Supportive/conservative therapy for all patients**<br>- **RAS blockade (ACE inhibitor or ARB)** if tolerated<br>- Reduces intraglomerular pressure and proteinuria.<br>- **Salt restriction** and **loop diuretic as needed**<br>- For edema control.<br>- **Statin therapy**<br>- Reasonable for severe LDL elevation in nephrotic syndrome.<br>- **Monitor renal function, serum albumin, and proteinuria**<br>- To assess spontaneous remission vs persistent active disease.<br>- **Assess thrombotic risk carefully**<br>- MN with nephrotic syndrome carries increased risk of venous thromboembolism, especially when albumin is low.<br><br>2. **Consider immunosuppressive therapy because this is nephrotic primary MN**<br>- Since he has **nephrotic-range proteinuria (UPCR 5.0 g/gCr)** and **albumin 2.5 g/dL**, he is at meaningful risk of ongoing disease, although kidney function is still preserved.<br>- **Rituximab** is a strong evidence-based first-line option for primary PLA2R- |

| | |
|---|---|
| | associated MN because it is effective and generally safer than alkylating agents.<br>- If risk is judged lower or early spontaneous remission is considered possible, a period of optimized supportive care with close follow-up may also be reasonable before immunosuppression.<br><br>#### Rationale<br>- Treatment in primary MN is based on balancing the chance of spontaneous remission against the risk of persistent nephrotic syndrome, kidney function decline, and thrombosis.<br>- This patient has **clear nephrotic syndrome**, but also **preserved renal function** and **minimal chronic histologic damage**, which are favorable features.<br>- Thus, **aggressive supportive care is mandatory**, and **rituximab is an appropriate preferred immunosuppressive option** if proteinuria remains significant or if his overall risk is considered moderate to high.<br><br>#### Other possible options<br>- **Cyclophosphamide plus glucocorticoids**: effective, especially in higher-risk disease, but with greater toxicity.<br>- **Calcineurin inhibitors (cyclosporine or tacrolimus)**: can reduce proteinuria, but relapse after discontinuation is common.<br>- **Observation with supportive therapy alone**: acceptable initially in selected lower-risk patients with stable kidney function, provided close follow-up is possible.<br><br>In summary, the patient most likely has **primary PLA2R-associated membranous nephropathy causing nephrotic syndrome**, and management should center on **supportive nephrotic-syndrome care plus consideration of rituximab-based immunotherapy depending on risk and clinical course**. |
| With Static RAG (Model B) | **7. Treatment plan**<br><br>**Initial treatment plan**<br>1. **Supportive therapy for all patients**<br>- **RAS blockade** (ACE inhibitor or ARB), if tolerated<br>- To reduce intraglomerular pressure and proteinuria.<br>- **Salt restriction** and **diuretics** as needed<br>- To control edema.<br>- **Statin therapy**<br>- Reasonable given LDL 216 mg/dL and nephrotic dyslipidemia.<br>- Monitor:<br>- blood pressure,<br>- body weight/edema,<br>- serum creatinine,<br>- potassium,<br>- albumin,<br>- UPCR.<br><br>2. **Risk assessment for need of immunosuppression**<br>- He has **nephrotic-range proteinuria (5 g/gCr)** and hypoalbuminemia, but **normal kidney function** and little chronic damage on biopsy.<br>- Based on accepted management of primary MN, a short period of optimized supportive care with close monitoring can be considered, but persistent nephrotic syndrome would favor **immunosuppressive therapy**.<br><br>3. **Preferred disease-specific option if treatment escalation is needed** |

| | |
|---|---|
| | - **Rituximab** is a strong evidence-based option for primary PLA2R-associated MN.<br>- Rationale:<br>- effective for inducing remission,<br>- relatively favorable safety profile compared with alkylating-agent regimens,<br>- particularly suitable in PLA2R-positive primary MN.<br><br>**Alternative options**<br>- **Cyclophosphamide plus glucocorticoids**<br>- Effective, especially in higher-risk disease, but with greater toxicity.<br>- **Calcineurin inhibitors** (cyclosporine or tacrolimus)<br>- Can reduce proteinuria, but relapse after discontinuation is more common; often used when rituximab is unsuitable.<br>- **Observation with supportive care alone**<br>- Possible initially in selected patients because some undergo spontaneous remission, but continued heavy proteinuria and hypoalbuminemia would argue for active treatment if persistent.<br><br>**Overall plan at this point**<br>- Start **supportive nephrotic syndrome management immediately**.<br>- Follow proteinuria, albumin, renal function, and ideally **PLA2R antibody titer** if available.<br>- If nephrotic syndrome persists or risk is judged moderate/high, proceed with **rituximab-based therapy** for **primary PLA2R-associated MN**. |
| With Static RAG and Mult-agent framework (Model C) | ### 7. Treatment plan<br>## **Initial management plan**<br><br>### 1. Supportive therapy for nephrotic syndrome<br>- Start **RAS blockade** with an **ACE inhibitor or ARB** if tolerated, to reduce proteinuria<br>- **Salt restriction** and **diuretics** for edema control<br>- Treat marked hyperlipidemia with a **statin**<br>- Monitor:<br>- **blood pressure**<br>- **body weight and edema**<br>- **serum creatinine/eGFR**<br>- **potassium**<br>- **serum albumin**<br>- **proteinuria** (e.g., UPCR)<br><br>### 2. Risk stratification and close follow-up<br>Management should be guided by **risk of progression**, not by nephrotic syndrome alone.<br><br>Follow closely:<br>- **proteinuria trend**<br>- **serum albumin**<br>- **creatinine/eGFR**<br>- **blood pressure**<br>- **anti-PLA2R antibody level**, if available, because it can help assess immunologic activity and response<br><br>The biopsy shows **little chronic tubulointerstitial injury** and creatinine is currently |

preserved, which are favorable features, but he already has clinically significant nephrotic syndrome and therefore requires careful surveillance.

#### 3. Assess thromboembolic risk
This is especially important because:
- he has **nephrotic syndrome**
- **albumin is 2.5 g/dL**
- MN is associated with increased **venous thromboembolism risk**

He should have **individualized assessment for VTE prophylaxis/anticoagulation**, balancing:
- serum albumin level
- severity of proteinuria
- mobility and other thrombosis risks
- bleeding risk

#### 4. Consider immunosuppressive therapy if risk is moderate/high or disease persists
In primary MN, immunosuppression is generally considered when there is:
- **persistent nephrotic-range proteinuria despite optimized supportive care**
- **declining kidney function**
- severe or prolonged nephrotic syndrome
- other features suggesting higher risk of progression

**Rituximab** is a common evidence-based first-line option in many patients with primary MN.

#### Alternative immunosuppressive options
- **Cyclophosphamide plus glucocorticoids**
  - effective, especially in higher-risk disease
  - but associated with greater toxicity
- **Calcineurin inhibitors** (cyclosporine or tacrolimus)
  - can reduce proteinuria
  - but relapse after withdrawal and nephrotoxicity are concerns

### **Rationale**
- He has **biopsy-proven MN**, currently **preserved kidney function**, and **minimal chronic scarring**, so **supportive care and structured risk assessment are essential first steps**.
- However, because he already has **established nephrotic syndrome** with substantial proteinuria and hypoalbuminemia, spontaneous remission cannot be assumed.
- **Rituximab** is attractive in PLA2R-associated MN because it targets the immune process and avoids some toxicities of alkylating therapy.
- The decision to start immunosuppression should depend on the **trajectory** of proteinuria, kidney function, serum albumin, and immunologic activity rather than a single snapshot alone.

### **Additional important measures**
- Continue evaluation for **secondary causes** as clinically appropriate, especially:
  - **age-appropriate malignancy screening**
  - **infection screening** if not already done
- Educate the patient about signs of:
  - worsening edema
  - reduced urine output

| | |
|---|---|
| | - thrombosis (leg swelling, chest pain, dyspnea)<br><br>## **Changes from the initial plan**<br>- The plan is now **more specific to PLA2R-positive membranous nephropathy** rather than a general nephrotic syndrome workup.<br>- **Biopsy results now drive management**, making alternative diagnoses much less relevant.<br>- Greater emphasis is placed on:<br>- **risk-based follow-up**<br>- **anti-PLA2R monitoring if available**<br>- **individualized thromboembolism assessment**<br>- deciding on immunosuppression based on **persistence/severity of proteinuria and risk of progression**, not nephrotic syndrome alone<br><br>**In summary:**<br>The most appropriate current approach is **optimized supportive therapy, KDIGO-style risk stratification, close monitoring, individualized VTE risk assessment, and consideration of rituximab-based immunosuppression if the disease remains active or risk of progression is judged moderate to high**. |
| **Phase III – Q8: Evaluation of treatment** | |
| Baseline LLM (Model A) | ## 8. Monitoring treatment effectiveness, safety, risks, and countermeasures<br><br>### A. Effectiveness monitoring<br>Monitor both **IgG4-RD activity** and **MN activity**, because both improved after prednisolone.<br><br>1. **Kidney disease response**<br>- **Urinalysis and UPCR**: follow regularly to confirm maintenance of complete remission and detect relapse early.<br>- **Serum albumin and lipid profile**: improvement supports sustained nephrotic syndrome remission.<br>- **Serum creatinine/eGFR**: confirm preserved kidney function.<br><br>2. **IgG4-RD response**<br>- **Serum IgG4 (and IgG)**: useful for trend monitoring; the marked fall after treatment supports response, though clinical/imaging correlation is essential.<br>- **Imaging of pancreas and kidneys**:<br>- **CT/MRI/MRCP as appropriate** to confirm continued regression/no re-enlargement of renal masses and pancreatic lesion, and no recurrent ductal abnormality.<br>- **Clinical symptoms/signs** of pancreatitis, obstructive jaundice, or other organ involvement should be reviewed at each visit.<br><br>3. **Glucocorticoid taper monitoring**<br>- During dose reduction, watch for **rebound proteinuria**, rising **IgG4**, recurrent **renal masses/pancreatic swelling**, or worsening kidney function.<br><br>### B. Safety monitoring: specific risks and countermeasures<br>1. **Infection risk from prednisolone**<br>- Important because he previously had **asymptomatic esophageal candidiasis**.<br>- **Monitor**: fever, oral/esophageal symptoms, respiratory symptoms, urinary symptoms; CBC if clinically indicated.<br>- **Countermeasures**: |

| | |
|---|---|
| | - Use the **lowest effective maintenance dose** and taper carefully.<br>- Maintain vigilance for candidiasis recurrence; evaluate promptly if suggestive symptoms appear.<br>- Avoid unnecessary additional immunosuppression unless relapse or steroid-refractory disease occurs.<br><br>2. **Steroid-related metabolic adverse effects**<br>- **Monitor**: blood pressure, body weight, edema, fasting glucose/HbA1c, lipids.<br>- **Countermeasures**:<br>- Dietary counseling (salt restriction, weight control).<br>- Treat hypertension, diabetes, or dyslipidemia if they emerge or worsen.<br><br>3. **Bone and musculoskeletal toxicity**<br>- **Monitor**: risk of osteoporosis/fracture and myopathy during prolonged steroid therapy.<br>- **Countermeasures**:<br>- Bone protection according to standard practice for chronic glucocorticoid use (e.g., calcium/vitamin D and fracture-risk-based therapy as appropriate).<br>- Encourage exercise/physical activity as tolerated.<br><br>4. **Gastrointestinal adverse effects**<br>- **Monitor**: dyspepsia, GI bleeding symptoms.<br>- **Countermeasures**:<br>- Gastroprotection if clinically indicated, especially if other GI risk factors are present.<br><br>5. **Relapse of IgG4-RD or MN**<br>- **Monitor**: UPCR, urinalysis, creatinine/eGFR, serum IgG4, and periodic imaging.<br>- **Countermeasures**:<br>- If mild recurrence, reconsider steroid dose/taper speed.<br>- If repeated relapse or steroid toxicity occurs, consider steroid-sparing therapy based on overall disease behavior.<br><br>6. **Need to avoid missing malignancy**<br>- Renal and pancreatic lesions were biopsied and improved with steroids, supporting IgG4-RD, but continued follow-up is still important.<br>- **Monitor**: interval imaging and clinical course.<br>- **Countermeasures**:<br>- If lesions fail to regress, enlarge, or show atypical behavior, reassess for malignancy rather than assuming IgG4-RD alone. |
| With Static RAG (Model B) | 8. **Monitoring of treatment effectiveness, safety, risks, and countermeasures**<br><br>**A. Effectiveness monitoring**<br><br>- **Kidney response (MN)**<br>- Follow **urinalysis and UPCR** regularly to confirm sustained remission or detect relapse early.<br>- Monitor **serum albumin, creatinine, and eGFR**.<br>- In this case, remission was supported by the fall in proteinuria to **UPCR 0.17 then 0.08 g/gCr**, normalization of albumin, and stable kidney function.<br><br>- **IgG4-RD activity** |

- Follow **serum IgG4 and total IgG** trends.
- Assess for recurrence of organ involvement using **symptoms, labs, and imaging**.
- For this patient, **CT/MRCP** were useful because they documented improvement in **renal masses, pancreatic mass, and pancreatic duct irregularity** after prednisolone.

- **Practical follow-up plan**
  - During steroid taper and early maintenance: periodic checks of
    - **urinalysis/UPCR**
    - **serum Cr/eGFR**
    - **albumin**
    - **IgG4**
  - Repeat **abdominal imaging** if relapse is suspected or to confirm continued stability of pancreatic/renal lesions.

**B. Safety monitoring for prednisolone**

Main risks and countermeasures:

1. **Infection**
   - Important because he had **esophageal candidiasis** before steroid use.
   - Watch for fever, cough, dysphagia/odynophagia, urinary symptoms, or other new infectious symptoms.
   - Perform routine clinical review and basic labs as needed.
   - **Countermeasures:** use the **lowest effective steroid dose**, taper appropriately, and promptly evaluate/treat suspected infection. Particular attention should be paid to recurrence/worsening of fungal infection.

2. **Steroid-induced hyperglycemia**
   - Especially relevant because pancreatic disease and glucocorticoids both can impair glucose tolerance.
   - Monitor **fasting glucose or HbA1c** periodically.
   - **Countermeasures:** diet advice, closer glucose monitoring after steroid initiation/escalation, and start antihyperglycemic therapy if needed.

3. **Hypertension / fluid retention**
   - Monitor **blood pressure, body weight, edema**.
   - **Countermeasures:** continue salt restriction, adjust antihypertensive/diuretic therapy as needed.

4. **Bone toxicity (osteoporosis/fracture)**
   - Relevant with prolonged glucocorticoid therapy.
   - **Countermeasures:** assess fracture risk, consider **calcium/vitamin D** and guideline-based osteoporosis prevention/treatment if prolonged therapy continues.

5. **Gastrointestinal adverse effects**
   - Steroids may aggravate GI symptoms.
   - **Countermeasures:** monitor symptoms; consider gastroprotection if clinically indicated.

6. **Dyslipidemia**
   - Nephrotic dyslipidemia improved with remission, but steroids can worsen metabolic parameters.

| | |
|---|---|
| | - Monitor **lipid profile** periodically.<br>- **Countermeasures:** lifestyle management and lipid-lowering therapy if needed.<br><br>7. **Adrenal suppression / relapse during taper**<br>- Watch for relapse of proteinuria or IgG4-RD lesions when reducing prednisolone.<br>- **Countermeasures:** taper gradually and reassess promptly if disease activity returns.<br><br>**C. Disease-specific risks to watch for**<br>- **Relapse of IgG4-RD** in pancreas or kidney<br>- **Relapse of MN/proteinuria**<br>- **Progressive tubulointerstitial injury** despite preserved creatinine<br>- **Initially missed malignancy**, since pancreatic and renal masses were part of the presentation and were biopsied as non-malignant<br><br>**Countermeasures**<br>- Continue integrated monitoring of **proteinuria + kidney function + IgG4 + imaging**<br>- Re-biopsy or repeat focused imaging if lesions enlarge, atypical features appear, or steroid response becomes inconsistent with IgG4-RD |
| With Static RAG and Mult-agent framework (Model C) | ## 8. Monitoring of treatment effectiveness, safety, risks, and countermeasures<br><br>Because prednisolone was started for **IgG4-RD (AIP and IgG4-RKD)** and this was followed by **complete remission of proteinuria in PLA2R-positive MN**, monitoring should assess both **IgG4-RD organ response** and **renal disease activity**, while also watching for **steroid toxicity** and **ARB-related adverse effects**.<br><br>### A. Monitoring treatment effectiveness<br><br>#### 1) Renal response: highest priority<br>In this case, the most practical markers of ongoing renal response are **serial proteinuria and renal function**, more than serum IgG4 alone.<br><br>- **Urinalysis and UPCR**<br>- Track remission of MN and detect relapse early<br>- Especially important because the patient previously had persistent residual proteinuria on losartan before steroid treatment<br>- **Serum creatinine/eGFR**<br>- Monitor renal function and detect recurrent IgG4-RKD, progressive tubulointerstitial injury, or other kidney complications<br>- **Serum albumin**<br>- Confirms sustained remission of nephrotic syndrome<br>- **Anti-PLA2R antibody level, if available**<br>- May help distinguish ongoing or relapsing **PLA2R-associated MN activity** from IgG4-RD activity<br><br>#### 2) IgG4-RD activity<br>- **Clinical assessment**<br>- Watch for symptoms suggesting recurrent pancreatic or renal disease, such as abdominal pain, jaundice, constitutional symptoms, or new renal findings<br>- **Serum IgG4 and total IgG**<br>- Useful as **supportive trend markers** |

  - In this patient, IgG4 fell promptly after prednisolone, supporting treatment response
  - However, relapse/remission should be judged mainly by **clinical findings, proteinuria/renal parameters, and imaging when indicated**
- **Imaging of pancreas and kidneys**
  - **CT/MRI/MRCP** as clinically indicated to document regression or detect relapse of pancreatic and renal lesions

#### B. Safety monitoring during treatment

##### 1) Prednisolone-related adverse effects

**Infection**
This is particularly important here because glucocorticoids were initially deferred due to **asymptomatic esophageal candidiasis**.

- **Monitor**
  - Fever
  - oral or esophageal symptoms such as odynophagia or dysphagia
  - cough, urinary symptoms, other focal infectious symptoms
  - CBC and other infection-related labs as clinically indicated
- **Countermeasures**
  - Low threshold to evaluate for recurrent fungal infection if symptoms recur
  - Prompt targeted investigation and treatment if infection is suspected
  - Taper to the lowest effective maintenance dose

**Metabolic and cardiovascular toxicity**
- **Monitor**
  - blood pressure
  - body weight
  - fasting glucose or **HbA1c**
  - lipid profile
- **Countermeasures**
  - salt restriction and lifestyle measures
  - treat hypertension, diabetes, or dyslipidemia if needed

**Bone toxicity**
- **Monitor**
  - fracture risk, back pain, height loss, osteoporosis risk factors
- **Countermeasures**
  - standard bone protection for prolonged steroid therapy as appropriate

**GI toxicity and adrenal suppression**
- **Monitor**
  - dyspepsia/GI symptoms
  - symptoms of adrenal insufficiency during taper
- **Countermeasures**
  - GI protection if clinically indicated
  - gradual tapering

##### 2) Losartan-related monitoring
Because the patient remains on **losartan**, routine monitoring should include:
- **serum creatinine/eGFR**
- **serum potassium**
- blood pressure

<table>
<tr><td></td><td>This helps detect ARB-related hyperkalemia or a hemodynamic fall in GFR.<br><br>### C. Specific risks in this case<br>1. **Relapse of IgG4-RD** during steroid taper or maintenance<br>2. **Relapse of proteinuria/MN**, despite current complete remission<br>3. **Progression of tubulointerstitial injury** even if serum creatinine remains initially preserved<br>4. **Recurrent infection**, especially fungal infection<br>5. **Long-term steroid toxicity**<br>6. **ARB-related hyperkalemia or renal function change**<br><br>### D. Practical follow-up plan<br>A practical follow-up schedule would be:<br><br>- **More frequent visits early after steroid initiation and during taper**<br>- Then **less frequent follow-up once stable on maintenance therapy**<br><br>At follow-up visits, monitor:<br>- blood pressure, weight, edema, symptoms of infection<br>- **urinalysis and UPCR**<br>- **serum creatinine/eGFR and potassium**<br>- serum albumin<br>- fasting glucose or **HbA1c** as appropriate<br>- periodic lipid profile<br>- **periodic serum IgG4/IgG**<br>- **anti-PLA2R antibody**, if available and clinically useful<br><br>Repeat **CT/MRI/MRCP**:<br>- to document sustained response when clinically needed<br>- or promptly if relapse of pancreatic/renal lesions is suspected<br><br>### Changes from the initial version<br>- Greater emphasis is now placed on **serial UPCR, albumin, and renal function** as the main markers of response<br>- **Serum IgG4** is now described as a **supportive trend marker**, not the primary determinant of remission<br>- Added explicit monitoring of **potassium and creatinine/eGFR while on losartan**<br>- Added more specific steroid safety monitoring, including **HbA1c/glucose** and **bone protection**<br>- Added **anti-PLA2R antibody monitoring if available**, given the persistent **PLA2R-positive MN phenotype**</td></tr>
<tr><td colspan="2">Phase III – Q9: Formulating plans for worsening conditions</td></tr>
<tr><td>Baseline LLM (Model A)</td><td>## 9. If the condition does not improve as expected or changes suddenly: reassessment and action plan<br><br>### A. First step: identify what changed<br>Rapidly determine whether the problem is:<br>1. **Relapse/progression of IgG4-RD**<br>2. **Relapse/worsening of MN**<br>3. **Treatment complication**, especially infection or steroid toxicity<br>4. **Alternative diagnosis**, including malignancy or another kidney disease</td></tr>
</table>

| | |
|---|---|
| | ### B. Practical reassessment by scenario<br>1. **If proteinuria rises again**<br>- Recheck **urinalysis, UPCR, albumin, creatinine/eGFR**.<br>- Consider:<br>- relapse of MN,<br>- relapse of IgG4-RKD,<br>- reduced treatment adherence/effect of losartan,<br>- emergence of another glomerular process.<br>- If accompanied by rising IgG4 or recurrent masses/pancreatic abnormalities, relapse of IgG4-RD becomes more likely.<br><br>2. **If creatinine rises or kidney function worsens**<br>- Urgently distinguish:<br>- active IgG4-related TIN,<br>- hemodynamic/drug-related decline,<br>- obstruction,<br>- another AKI cause.<br>- Repeat labs and **kidney imaging**; if uncertainty remains or findings are discordant, reconsider **repeat kidney biopsy**.<br><br>3. **If renal or pancreatic masses enlarge or fail to improve**<br>- Do **not** assume refractory IgG4-RD automatically.<br>- Reconsider **malignancy**, especially because mass lesions were an important part of the presentation.<br>- Repeat targeted imaging and obtain **repeat tissue diagnosis** if needed.<br><br>4. **If fever or systemic symptoms develop after starting steroids**<br>- Prioritize evaluation for **infection** before escalating immunosuppression.<br>- Given prior esophageal candidiasis, fungal relapse is one concern, but common bacterial/viral infections must also be assessed.<br><br>### C. How treatment would be revised<br>- **If clear steroid-responsive IgG4-RD relapse**: adjust steroid dose and taper more cautiously.<br>- **If relapse is frequent or steroid toxicity becomes problematic**: consider a **steroid-sparing strategy**.<br>- **If MN relapses independently of IgG4-RD activity**: reassess whether MN behavior is still linked to IgG4-RD or whether management should shift toward standard primary MN-directed therapy.<br>- **If malignancy or another diagnosis is suspected**: pause anchoring on the original diagnosis and redirect evaluation/treatment accordingly.<br><br>### Bottom line<br>The key is **serial integrated monitoring** of proteinuria, kidney function, serum IgG4, and imaging, while remaining alert to **infection, steroid toxicity, relapse, and missed malignancy**. If the course becomes atypical, the diagnosis must be **re-opened promptly**, with repeat imaging and, when necessary, repeat biopsy rather than assuming all changes are due to IgG4-RD alone. |
| With Static RAG (Model B) | 9. **If the condition does not improve as expected or changes suddenly: reassessment and action plan**<br><br>**A. Reassess the problem by pattern of change** |

1. **If proteinuria worsens or MN relapses**
  - Reconsider whether the proteinuria reflects:
    - relapse of **primary PLA2R-positive MN**,
    - relapse of **IgG4-RD activity**,
    - or both.
  - Check:
    - **UPCR/urinalysis**
    - **serum albumin**
    - **Cr/eGFR**
    - **serum IgG4**
  - If available in practice, **PLA2R antibody** can help assess activity of primary MN.
  - If the pattern is unclear or atypical, consider whether repeat kidney biopsy is needed.

2. **If renal function declines**
  - Think beyond MN alone, because MN typically causes proteinuria more than abrupt creatinine rise.
  - Rapid reassessment should include:
    - volume status,
    - drug effects,
    - obstruction,
    - active IgG4-related TIN,
    - other superimposed kidney injury.
  - Repeat labs and renal imaging promptly.

3. **If pancreatic or renal masses enlarge, or response to steroids is poor**
  - Reconsider the diagnosis, especially **malignancy** or another inflammatory disease.
  - This is crucial because mass-like lesions can mimic cancer.
  - Repeat **CT/MRI/MRCP**, and if findings are discordant with expected IgG4-RD behavior, consider **repeat tissue sampling**.

4. **If fever or systemic deterioration occurs after steroids**
  - Prioritize evaluation for **infection**, including recurrence of candidiasis, before assuming disease flare.

**B. How to revise treatment**

- **If relapse occurs during taper but disease remains steroid-responsive**
  - Adjust prednisolone to the minimum dose needed for control, then taper more cautiously.

- **If repeated relapse, steroid toxicity, or inadequate response occurs**
  - Consider adding or switching to a **steroid-sparing immunosuppressive strategy** used for IgG4-RD and/or MN according to the dominant process and overall risk.

- **If findings suggest that MN is behaving independently of IgG4-RD**
  - Reassess as **persistent primary PLA2R-associated MN** and treat according to standard MN risk-based management.

**C. Practical principle**
- Do not assume every new abnormality is explained by the known diagnosis.
- In this case, because both **primary PLA2R-positive MN** and **IgG4-RD (AIP

| | |
|---|---|
|  | + TIN/renal masses)** coexist, any unexpected change should trigger a structured reassessment of:<br> 1. **disease relapse**,<br> 2. **treatment toxicity**,<br> 3. **infection**, and<br> 4. **missed malignancy or alternative diagnosis**. |
| With Static RAG and Mult-agent framework (Model C) | ## 9. Reassessment and action plan if the condition does not improve as expected or changes suddenly<br><br>### A. First define the pattern of change<br>The main abnormal scenarios in this patient would be:<br>1. **Proteinuria increases again while kidney function remains stable**<br>2. **Serum creatinine rises**<br>3. **Renal or pancreatic masses fail to regress, recur, or enlarge**<br>4. **New signs of infection appear**<br>5. **Serologic changes such as rising IgG4 or, if available, anti-PLA2R activity**<br><br>### B. Differential reassessment: keep multiple mechanisms in parallel<br>If the course changes unexpectedly, I would not assume a single explanation. The key possibilities are:<br><br>- **Relapse or insufficient control of IgG4-RD**<br>- **Persistent or relapsing PLA2R-associated MN**, which may not be fully explained by IgG4-RD alone<br>- **Missed malignancy**, especially if mass lesions persist or enlarge despite steroid therapy<br>- **Treatment-related complications**, including steroid-related infection or drug-related renal dysfunction<br><br>### C. Action plan organized by clinical pattern<br><br>#### 1) If proteinuria worsens but creatinine is stable<br>- Repeat:<br> - urinalysis<br> - **UPCR**<br> - serum albumin<br> - creatinine/eGFR<br>- Review:<br> - adherence to losartan and supportive measures<br> - blood pressure control<br>- Reassess **anti-PLA2R antibody** if available<br>- Consider that MN may be relapsing even if IgG4-RD appears otherwise controlled<br><br>#### 2) If creatinine rises<br>Promptly assess for:<br>- volume/hemodynamic causes<br>- drug effects<br>- hyperkalemia-associated issues<br>- urinary obstruction<br>- recurrent or progressive **IgG4-RKD/TIN**<br>- other causes of AKI<br><br>Then obtain: |

- repeat renal labs
- urinalysis
- renal imaging as indicated

##### 3) If renal or pancreatic lesions fail to shrink, recur, or enlarge
This should raise renewed concern for **malignancy**, because both pancreatic and renal IgG4-RD lesions can mimic cancer.

- Repeat appropriate imaging
- Reconsider tissue diagnosis
- Maintain a **low threshold for repeat biopsy or oncologic evaluation** if the response is not as expected

##### 4) If infection is suspected
- Assess urgently
- Look specifically for recurrence of **fungal infection** as well as common bacterial infections
- Consider temporary modification of immunosuppression depending on severity
- Start targeted antimicrobial therapy promptly

#### D. How the diagnosis and treatment plan would be revised
- **If IgG4-RD relapse is likely**
  - increase glucocorticoid intensity as needed
  - reconsider the maintenance strategy and possible steroid-sparing therapy
- **If MN remains active or relapses despite control of IgG4-RD**
  - reassess this as persistent or relapsing **PLA2R-associated MN**
  - consider MN-directed immunosuppressive therapy
- **If steroid toxicity becomes limiting**
  - reduce steroid exposure if possible
  - consider alternative steroid-sparing approaches
- **If malignancy becomes a concern**
  - prioritize repeat tissue confirmation and oncologic workup

#### E. Core reasoning
This patient's pancreatic lesion, renal masses, serum IgG4 elevation, renal TIN with IgG4-positive plasma cells, and strong response to prednisolone support true **IgG4-RD**. At the same time, the glomerular lesion remained **PLA2R-positive**, so the MN may represent either an overlapping process or a disease modified by IgG4-RD treatment. Therefore, if the clinical course changes, reassessment should simultaneously address:
- **IgG4-RD activity**
- **MN activity**
- **treatment toxicity**
- **alternative diagnoses, especially malignancy**

#### Changes from the initial version
- The reassessment plan is now more clearly organized by **clinical change pattern**
- It now more explicitly separates:
  - worsening **proteinuria with stable renal function**
  - rising **creatinine**
  - **non-resolving/recurrent mass lesions**
  - **infection signs**
- It adds clearer emphasis that **persistent or enlarging lesions despite steroids should trigger renewed concern for malignancy and possible repeat biopsy**

| | |
|---|---|
| | - It also adds reassessment of **supportive therapy and anti-PLA2R status** when proteinuria relapses |

| | |
|---|---|
| **Case 2: A case of posterior and reversible encephalopathy syndrome in a patient previously undiagnosed with lupus nephritis** [10] | |
| **Phase I – Q1: Summarizing medical problems** | |
| Baseline LLM (Model A) | 1. **Problem representation**<br><br>A previously healthy **37-year-old woman** presents with **acute visual impairment and headache** in the setting of **severe hypertension**, after a **3-month history of newly diagnosed hypertension** and a **2-month history of leg edema with proteinuria**. She has also been taking a **low-estrogen oral contraceptive for 3 years**. This suggests a **hypertensive emergency with target-organ involvement (especially ocular/CNS)** superimposed on a recent syndrome of **hypertension + edema + proteinuria**, raising concern for **secondary hypertension and renal disease**. |
| With Static RAG (Model B) | 1. **Problem representation**<br><br>A **37-year-old woman** with **newly diagnosed hypertension over 3 months**, followed by **leg edema and proteinuria 2 months earlier**, now presents with **sudden visual impairment, headache, and markedly elevated blood pressure**, concerning for **acute severe hypertension with target-organ damage (hypertensive emergency), especially hypertensive retinopathy/encephalopathy**, on a background suggesting possible **secondary hypertension with renal involvement**. Her use of a **low-estrogen oral contraceptive** is a relevant thrombotic risk factor. |
| With Static RAG and Mult-agent framework (Model C) | 1. **Problem representation**<br><br>A **37-year-old woman** with **new-onset hypertension over 3 months**, followed by **leg edema and proteinuria 2 months earlier**, now presents with **sudden visual impairment, headache, and severe hypertension**, consistent with a **hypertensive emergency with target-organ damage**. In a previously healthy young woman, the combination of **acute severe hypertension plus renal involvement (edema, proteinuria)** raises concern for **secondary hypertension**, especially **renal parenchymal disease, renovascular disease, or pregnancy-related hypertensive disease**, while **thrombotic microangiopathy** must also be considered. Her **oral contraceptive use** is relevant because it may contribute to **hypertension and thrombotic risk**.<br><br>**Change from initial version:**<br>I broadened the framing from “secondary renal/renovascular cause” to **“hypertensive emergency with renal involvement in a young woman”**, explicitly keeping **intrinsic renal disease** and **pregnancy-related disease** in the front line before narrowing to renovascular causes. |
| **Phase I – Q2: Differential diagnoses and rationale** | |
| Baseline LLM (Model A) | 2. **Differential diagnosis**<br><br>**Top 3 differential diagnoses:**<br><br>1. **Hypertensive emergency due to glomerular/renal parenchymal disease** |

| | |
|---|---|
| | - Most likely because the patient had **recent hypertension, edema, and proteinuria**, which strongly suggest intrinsic renal disease causing volume expansion and worsening BP.<br>- Acute visual symptoms and headache fit **severe hypertensive retinopathy, choroidopathy, or encephalopathy/PRES**.<br><br>2. **Thrombotic microangiopathy (TMA), including malignant hypertension-associated TMA**<br>- Must consider because severe hypertension with possible renal injury and visual/CNS symptoms can occur with **malignant hypertension**, which may also cause hemolysis, thrombocytopenia, and acute kidney injury.<br>- Important because delay in recognition may be life-threatening.<br><br>3. **Secondary hypertension due to renovascular disease or endocrine causes**<br>- Examples: **renal artery stenosis/fibromuscular dysplasia**, less likely primary aldosteronism or pheochromocytoma.<br>- Young woman with recent-onset hypertension raises concern for a secondary cause, though **proteinuria and edema** make renal parenchymal disease more likely than purely endocrine hypertension.<br><br>**Most likely diagnosis:**<br>**Hypertensive emergency caused by renal parenchymal disease (likely glomerular disease) with hypertensive retinopathy/encephalopathy.**<br><br>**Must not miss:**<br>**Hypertensive encephalopathy/PRES and TMA**, because both can rapidly lead to irreversible neurologic, renal, and visual damage. |
| With Static RAG (Model B) | 2. **Differential diagnosis**<br><br>**Top 3 differential diagnoses**<br>1. **Hypertensive emergency due to secondary hypertension, likely renal/parenchymal or renovascular cause**<br>- The combination of **recent-onset hypertension, edema, proteinuria, headache, and visual symptoms** strongly suggests severe hypertension with end-organ injury.<br>- Proteinuria and edema raise concern for **kidney disease as either the cause or target of hypertension**.<br><br>2. **Thrombotic microangiopathy (TMA), including malignant hypertension-associated TMA or TTP**<br>- Sudden severe hypertension with neurologic/visual symptoms and renal involvement can occur in **malignant hypertension** and may overlap with **TMA**.<br>- Must consider because delayed recognition can be dangerous.<br><br>3. **Cerebral venous sinus thrombosis (CVST) or other thrombotic cerebrovascular event related to oral contraceptive use**<br>- **Headache + visual disturbance** in a woman taking oral contraceptives should prompt consideration of **intracranial thrombosis**.<br>- Severe hypertension may be reactive or coexist, though this does not fully explain proteinuria/edema.<br><br>**Most likely diagnosis**<br>- **Hypertensive emergency, probably malignant hypertension, likely on the basis of |

| | |
|---|---|
| | underlying renal disease or secondary hypertension.**<br>- Reason: This best integrates the **subacute course of new hypertension**, **proteinuria/edema**, and the **acute onset of headache and visual impairment**.<br><br>**Must not be missed**<br>- **Hypertensive encephalopathy / posterior reversible encephalopathy syndrome (PRES)**<br>- **TTP/TMA**<br>- **Cerebrovascular event including CVST or intracranial hemorrhage**<br><br>These are immediately dangerous and can present with **headache, visual symptoms, severe hypertension, and renal abnormalities**. |
| With Static RAG and Mult-agent framework (Model C) | 2. **Prioritized differential diagnosis**<br><br>### A. Immediate clinical syndrome causing the current emergency<br>1. **Hypertensive emergency / malignant hypertension**<br>- Sudden **headache, visual impairment, and severe hypertension** strongly suggest acute end-organ injury involving the **retina, brain, and kidneys**.<br><br>2. **Hypertensive encephalopathy / PRES**<br>- Important because severe hypertension with **headache and visual symptoms** may reflect cerebral edema or posterior reversible encephalopathy.<br><br>3. **Thrombotic microangiopathy (TMA)**<br>- Can present with **severe hypertension, renal injury, neurologic/visual symptoms**, and may either be caused by malignant hypertension or be a primary process (less likely TTP/HUS).<br><br>### B. Likely underlying etiology of the hypertension<br>4. **Renal parenchymal disease (glomerular disease; nephritic/nephrotic spectrum)**<br>- Supported by **proteinuria and edema** preceding this presentation.<br><br>5. **Renovascular hypertension**<br>- Important in a **young woman with recent severe hypertension**, especially possible **fibromuscular dysplasia**.<br><br>6. **Pregnancy-related hypertensive disease (preeclampsia/eclampsia spectrum)**<br>- Must be excluded in any reproductive-age woman with **hypertension, proteinuria, headache, and visual symptoms**.<br><br>7. **Medication-related contribution**<br>- **Oral contraceptives** can worsen hypertension and increase thrombotic risk.<br><br>**Most likely diagnosis:**<br>**Hypertensive emergency (malignant hypertension) with retinal/CNS/renal involvement, likely due to an underlying secondary renal cause.**<br><br>**Must not miss diagnoses:**<br>- **Pregnancy-related hypertensive disease**<br>- **Thrombotic microangiopathy**<br>- **Hypertensive encephalopathy/PRES**<br>- **Intracranial hemorrhage or stroke** |

| | |
|---|---|
| | **Change from initial version:**<br>I separated the differential into **(1) the acute emergency syndrome** and **(2) the underlying cause**, added **pregnancy-related hypertensive disease** explicitly, and broadened renal causes to include **renal parenchymal/glomerular disease** rather than emphasizing renovascular disease too early. |
| **Phase I – Q3: Necessary physical examinations and reasons** | |
| Baseline LLM (Model A) | 3. **Additional history and physical examination**<br><br>**History**<br>- **Severity and timing of visual symptoms**: blurring vs field defect vs transient loss<br> - Helps distinguish hypertensive retinopathy/choroidopathy, retinal vascular event, or neurologic cause.<br>- **Neurologic symptoms**: confusion, seizure, focal deficits, nausea/vomiting<br> - Evaluates for hypertensive encephalopathy or PRES.<br>- **Renal/nephritic features**: gross hematuria, foamy urine, oliguria<br> - Supports glomerular disease.<br>- **Systemic symptoms**: fever, rash, arthralgia, oral ulcers, Raynaud symptoms<br> - Screens for autoimmune glomerulonephritis (e.g., lupus, vasculitis).<br>- **Pregnancy/gynecologic history**: current pregnancy possibility, recent postpartum state<br> - Important because preeclampsia-related disease must be excluded in women of childbearing age.<br>- **Drug history**: NSAIDs, sympathomimetics, supplements, cocaine, adherence to telmisartan<br> - Identifies medication/substance-related hypertension or renal injury.<br>- **Family history**: kidney disease, hypertension, autoimmune disease<br> - May suggest hereditary or immune-mediated disease.<br><br>**Physical examination**<br>- **Repeat BP in both arms, pulse, temperature**<br> - Confirms severity and assesses for secondary causes.<br>- **Fundoscopy**<br> - Look for papilledema, retinal hemorrhages, cotton-wool spots, exudates: evidence of hypertensive emergency.<br>- **Focused neurologic exam**<br> - Detects encephalopathy, focal deficits, seizure sequelae.<br>- **Volume status**: edema, JVP, lung crackles<br> - Assesses fluid overload from renal disease.<br>- **Cardiovascular exam**<br> - Looks for S4, heart failure, hypertensive target-organ damage.<br>- **Skin/joint exam**<br> - Rash, purpura, synovitis may suggest vasculitis or connective tissue disease.<br>- **Abdominal/bruit exam**<br> - Renal artery bruit may suggest renovascular hypertension. |
| With Static RAG (Model B) | 3. **Additional history and physical examination**<br><br>**History**<br>- **Blood pressure course/severity**: highest home/clinic BP, medication adherence, response to telmisartan<br> - To assess chronicity and severity.<br>- **Visual symptoms**: blurred vision, field defect, diplopia, transient vision loss<br> - To distinguish retinopathy, encephalopathy, stroke, or papilledema. |

| | |
|---|---|
|  | - **Neurologic symptoms**: confusion, seizure, focal weakness, dysarthria<br> - To evaluate hypertensive encephalopathy, PRES, stroke, CVST.<br>- **Renal symptoms**: foamy urine, oliguria, gross hematuria<br> - To assess glomerular disease or acute kidney injury.<br>- **Systemic symptoms**: fever, weight loss, rash, arthralgia<br> - To screen for vasculitis, lupus, systemic disease.<br>- **Thrombotic risk factors**: smoking, migraine, thrombosis history, family history, recent immobility<br> - Relevant to OCP-associated thrombosis/CVST.<br>- **Secondary hypertension clues**:<br> - Episodic palpitations/sweating → pheochromocytoma<br> - Muscle weakness/cramps → hyperaldosteronism<br> - Snoring/daytime sleepiness → sleep apnea<br> - Flank pain/abdominal bruit history → renovascular disease<br>- **Pregnancy possibility** despite OCP use<br> - Important because preeclampsia must be excluded in women of childbearing age.<br><br>**Physical examination**<br>- **Repeat BP in both arms, pulse, temperature, oxygen saturation**<br> - Confirm severity; inter-arm difference may suggest vascular disease.<br>- **Funduscopic examination**<br> - Look for hemorrhages, exudates, papilledema, retinal changes of malignant hypertension.<br>- **Full neurologic examination**<br> - Detect focal deficits, encephalopathy, seizure-related findings.<br>- **Volume status/cardiopulmonary exam**<br> - Edema, JVP, crackles, S3 for heart failure/volume overload.<br>- **Abdominal exam and auscultation for renal artery bruit**<br> - Suggest renovascular hypertension.<br>- **Skin exam**<br> - Rash, purpura, livedo for vasculitis/TMA/connective tissue disease. |
| With Static RAG and Mult-agent framework (Model C) | 3. **Purposeful history and physical examination**<br><br>### A. Immediate bedside priorities<br>- **Confirm blood pressure severity and repeat measurements**<br> - To verify true hypertensive emergency.<br>- **Assess mental status, confusion, seizure activity, focal deficits**<br> - To identify **hypertensive encephalopathy, stroke, or PRES**.<br>- **Ask about pregnancy possibility / last menstrual period**<br> - Essential because **preeclampsia/eclampsia** changes diagnosis and management.<br><br>### B. Symptoms clarifying target-organ damage<br>- **Headache details**: onset, severity, progression, associated nausea/vomiting<br> - Helps assess encephalopathy or intracranial hemorrhage.<br>- **Visual symptoms**: blurring, field loss, transient loss, diplopia<br> - Helps distinguish **retinopathy, retinal vascular disease, or CNS involvement**.<br>- **Neurologic symptoms**: seizures, weakness, numbness, altered behavior<br> - Screens for CNS end-organ injury.<br>- **Urinary symptoms**: oliguria, hematuria, foamy urine<br> - Helps identify renal injury or glomerular disease.<br>- **Dyspnea/chest pain**<br> - Screens for heart failure, pulmonary edema, or cardiac ischemia. |

| | |
|---|---|
|  | ### C. Clues to underlying secondary cause<br>- **Timeline of hypertension and prior BP readings**<br> - Rapid onset supports secondary hypertension.<br>- **Medication history**<br> - Telmisartan use, adherence, NSAIDs, decongestants, supplements, and oral contraceptive details.<br>- **Systemic symptoms**<br> - Fever, rash, arthralgia, weight loss may suggest autoimmune or vasculitic renal disease.<br>- **Family history**<br> - Kidney disease, autoimmune disease, severe early-onset hypertension.<br><br>### D. Physical examination<br>- **Fundoscopy**<br> - Look for **papilledema, retinal hemorrhages, cotton-wool spots, exudates**.<br>- **Full neurologic examination**<br> - Detect encephalopathy, stroke, seizure-related findings.<br>- **Volume status**<br> - Edema, JVP, lung crackles for renal/cardiac involvement.<br>- **Cardiovascular exam**<br> - S4, signs of LVH, heart failure.<br>- **Abdominal exam with renal artery bruit**<br> - Suggests renovascular disease.<br>- **Skin exam**<br> - Petechiae, purpura, livedo, rash may suggest **TMA, vasculitis, autoimmune disease**.<br><br>**Change from initial version:**<br>I reorganized this section into **emergency stabilization**, **target-organ-organ assessment**, and **secondary-cause clues**, with stronger emphasis on **pregnancy**, **encephalopathy/seizure risk**, and **funduscopic findings** as immediate bedside priorities. |
| **Phase I – Q4: Plan for investigations/tests** | |
| Baseline LLM (Model A) | 4. **Tests to order**<br><br>**Urgent initial tests**<br>- **Basic metabolic panel, creatinine, BUN, electrolytes**<br> - Purpose: assess kidney injury and electrolyte abnormalities.<br> - Expected: possible elevated creatinine if renal involvement is significant.<br>- **Urinalysis with microscopy and urine protein quantification**<br> - Purpose: characterize renal disease (proteinuria, hematuria, casts).<br> - Expected: significant proteinuria; possible dysmorphic RBCs/RBC casts if glomerulonephritis.<br>- **CBC, platelet count, LDH, haptoglobin, bilirubin, peripheral smear**<br> - Purpose: evaluate for TMA/hemolysis.<br> - Expected: if TMA, anemia, thrombocytopenia, schistocytes, elevated LDH, low haptoglobin.<br>- **Funduscopic examination by ophthalmology**<br> - Purpose: confirm hypertensive retinopathy/choroidopathy and assess severity.<br> - Expected: retinal hemorrhages/exudates/papilledema if malignant hypertension.<br>- **ECG**<br> - Purpose: assess cardiac target-organ damage. |

| | |
|---|---|
| | - Expected: possible LVH or ischemic changes.<br><br>**Imaging**<br>- **Brain MRI (or CT initially if urgent)**<br>- Purpose: evaluate headache/visual symptoms for PRES, hemorrhage, stroke.<br>- Expected: PRES would show posterior vasogenic edema on MRI.<br>- **Chest X-ray**<br>- Purpose: assess pulmonary edema/cardiomegaly if volume overloaded.<br>- **Renal ultrasound**<br>- Purpose: assess kidney size, chronicity, obstruction; useful baseline renal evaluation.<br>- Expected: may be normal-sized kidneys in acute/subacute intrinsic renal disease.<br><br>**Evaluation for etiology of renal disease/secondary hypertension**<br>- **Pregnancy test**<br>- Purpose: rule out pregnancy-related hypertensive disease.<br>- **Autoimmune/glomerulonephritis serologies**: ANA, anti-dsDNA, complements, ANCA, anti-GBM, hepatitis B/C as indicated<br>- Purpose: identify immune-mediated glomerular disease.<br>- Expected: depends on cause; may reveal lupus nephritis or vasculitis if present.<br>- **Renin/aldosterone and renal artery imaging (e.g., Doppler/CTA/MRA) if suggested**<br>- Purpose: evaluate secondary hypertension such as fibromuscular dysplasia.<br>- Expected: pursued especially if renal findings do not fully explain the presentation.<br><br>**Likely next diagnostic step if renal involvement is confirmed**<br>- **Kidney biopsy** (after stabilization)<br>- Purpose: establish the exact glomerular/vascular renal diagnosis and guide treatment.<br>- Expected: may show glomerulonephritis or vascular changes of malignant hypertension, depending on the underlying process. |
| With Static RAG (Model B) | 4. **Tests to narrow the diagnosis**<br><br>**Immediate tests**<br>1. **Basic metabolic panel, creatinine, BUN, electrolytes**<br>- Purpose: assess kidney function, AKI, electrolyte clues to secondary hypertension.<br>- Expected: possible **elevated creatinine** if renal injury is present.<br><br>2. **Urinalysis with microscopy and urine protein quantification**<br>- Purpose: confirm severity/type of renal involvement.<br>- Expected: **proteinuria**, possibly hematuria/casts if glomerular disease exists.<br><br>3. **CBC with smear, LDH, bilirubin, haptoglobin, platelets**<br>- Purpose: evaluate for **hemolysis/TMA/TTP**.<br>- Expected: in TMA, **anemia, thrombocytopenia, schistocytes, high LDH, low haptoglobin**.<br><br>4. **Funduscopic examination by ophthalmology if needed**<br>- Purpose: document **hypertensive retinopathy/papilledema**.<br>- Expected: retinal hemorrhages/exudates/papilledema in malignant hypertension.<br><br>5. **Brain imaging: non-contrast head CT initially; brain MRI if stable**<br>- Purpose: exclude **intracranial hemorrhage, stroke**, and assess for |

| | |
|---|---|
| | **PRES/CVST**.<br>- Expected:<br>- CT may exclude hemorrhage.<br>- MRI may show **posterior white matter edema** in PRES.<br>- MR venography if CVST is suspected.<br><br>**Tests for underlying cause**<br>6. **Pregnancy test**<br>- Purpose: exclude pregnancy-related hypertensive disease.<br>- Expected: usually negative, but essential not to miss.<br><br>7. **ECG and chest X-ray**<br>- Purpose: assess cardiac end-organ damage/heart failure.<br>- Expected: possible LVH or pulmonary congestion if severe longstanding hypertension.<br><br>8. **Renal ultrasound**<br>- Purpose: evaluate kidney size/asymmetry, obstruction, chronic parenchymal disease.<br>- Expected: may show evidence of parenchymal disease or asymmetry suggesting renovascular disease.<br><br>9. **Renin/aldosterone testing, plasma metanephrines, autoimmune serologies (ANA, complements, ANCA), as guided by initial results**<br>- Purpose: investigate **secondary hypertension** and systemic renal disease.<br>- Expected: depends on initial findings; these are not first-line before stabilization but are important once the patient is stabilized.<br><br>**Overall plan**<br>- The priority is to confirm **hypertensive emergency and end-organ injury**, while urgently excluding **PRES, stroke/CVST, and TMA**.<br>- Given the prior **proteinuria and edema**, renal disease is likely central to the case and should be evaluated early with kidney function testing and urine studies. |
| With Static RAG and Mult-agent framework (Model C) | 4. **Testing plan**<br><br>### A. Emergent tests needed immediately<br>1. **Pregnancy test**<br>- Must rapidly exclude **pregnancy-related hypertensive disease**.<br><br>2. **Serum creatinine, BUN, electrolytes**<br>- Assess kidney injury and guide acute BP management.<br><br>3. **Urinalysis with microscopy and urine protein quantification**<br>- Clarifies renal involvement; looks for **hematuria/casts** suggesting glomerular disease.<br><br>4. **CBC with platelet count**<br>- Looks for anemia and thrombocytopenia suggesting **TMA**.<br><br>5. **Hemolysis evaluation: peripheral smear, LDH, haptoglobin, bilirubin, reticulocyte count**<br>- Evaluates for **microangiopathic hemolytic anemia**. |

| | |
|---|---|
| | 6. **Funduscopic examination**<br>- Confirms hypertensive retinopathy/papilledema as target-organ damage.<br><br>7. **ECG**<br>- Assesses cardiac strain/ischemia.<br><br>8. **Brain imaging if neurologic symptoms are significant or diagnosis is unclear**<br>- **Head CT first** if concern for hemorrhage; **MRI** if evaluating for **PRES**.<br><br>### B. Early inpatient tests after initial stabilization<br>9. **Chest X-ray**<br>- If dyspnea or concern for pulmonary edema/heart failure.<br><br>10. **Renal ultrasound**<br>- Low-risk first test for kidney size, asymmetry, obstruction.<br><br>11. **Renal artery imaging (Doppler US, CTA, or MRA)**<br>- If suspicion remains high for renovascular hypertension, especially **fibromuscular dysplasia**.<br><br>### C. Tests deferred until stabilized and guided by initial results<br>12. **Autoimmune/glomerulonephritis workup**<br>- ANA, complements, anti-dsDNA, ANCA, anti-GBM, hepatitis serologies if urine sediment/renal function suggests intrinsic renal disease.<br><br>13. **Renin/aldosterone testing**<br>- Not an early priority because results are **confounded by acute illness and ARB therapy**.<br><br>**Change from initial version:**<br>I now clearly separate **emergent tests** from **post-stabilization evaluation**, moved **pregnancy test, renal function, urinalysis, CBC/hemolysis labs, fundus exam, and brain imaging when indicated** to the top priority, and **de-emphasized renin/aldosterone testing** in the acute setting because it is low yield and confounded by telmisartan use use.<br><br>---<br><br>### Summary<br>This patient most likely has a **hypertensive emergency with retinal, neurologic, and renal involvement**. The leading underlying causes are **renal parenchymal disease**, **renovascular hypertension**, and **pregnancy-related hypertensive disease**, while **TMA** must be urgently excluded. The immediate approach is to confirm end-organ damage, assess for CNS and retinal complications, evaluate renal injury, and rapidly rule out pregnancy and hemolysis. |
| **Phase II – Q5: Interpreting test results** | |
| Baseline LLM (Model A) | **5. Interpretation of the test results**<br><br>- **Hypertensive emergency with PRES:** BP **236/122 mmHg**, hypertensive retinopathy on funduscopy, and MRI showing **posterior vasogenic edema** (FLAIR high signal, increased ADC) with tiny hemorrhages are classic for **PRES due to severe hypertension**. Improvement after BP reduction strongly supports this |

| | |
|---|---|
| | mechanism.<br>- **Evidence of active SLE:** The **butterfly rash**, **low complements (C3/C4/CH50)**, and strongly positive **ANA** and **anti-dsDNA** indicate **immune-complex active lupus**. Anti-SSA/SSB support systemic autoimmunity.<br>- **Renal involvement is significant and active:** **Proteinuria** (UPCR 3.71, then 7.17 g/gCr), **hematuria**, **hypoalbuminemia (2.3 g/dL)**, and worsening **kidney function** indicate active glomerular disease, not just hypertensive nephrosclerosis.<br>- **Kidney biopsy confirms severe lupus nephritis:** Diffuse proliferative GN with **endocapillary proliferation**, **wire-loop lesions**, **full-house immunofluorescence**, and subendothelial/subepithelial/mesangial electron-dense deposits are classic for **lupus nephritis class IV-G (A)**. This is a highly active, severe form that explains the proteinuria, hematuria, renal dysfunction, and likely the hypertension.<br>- **Associated renal vasculitis:** **Fibrinoid necrosis in small vessels** indicates **vasculitic vascular injury**, which can contribute to severe hypertension and organ ischemia.<br>- **TMA/TTP is less likely:** No schistocytes, normal platelets, and normal **ADAMTS13** argue against TTP. No thrombi were seen on biopsy. Mildly elevated LDH alone is nonspecific.<br>- **Antiphospholipid antibodies:** **Anticardiolipin IgG positive** suggests possible antiphospholipid autoimmunity, but there is **no histologic thrombosis** and anti-β2GPI is negative, so APS nephropathy is not established from the provided data. |
| With Static RAG (Model B) | **5. Interpretation of the test results**<br><br>- **BP 236/122 mmHg with headache, visual impairment, hypertensive retinopathy, and MRI findings** indicates a **hypertensive emergency**.<br> - MRI showing **FLAIR hyperintensity in bilateral occipital lobes/cerebellum**, **increased ADC**, and small hemorrhages is typical of **PRES**, reflecting **vasogenic edema from failed cerebral autoregulation/endothelial injury**.<br> - Improvement after BP reduction strongly supports this mechanism.<br><br>- **Butterfly rash**, **low complements (C3/C4/CH50)**, and **positive ANA/anti-dsDNA** are highly consistent with **active SLE**, not incidental autoimmunity.<br> - Low complement plus high anti-dsDNA particularly suggests **immune-complex active lupus**, including renal involvement.<br><br>- **Proteinuria, hematuria, hypoalbuminemia (albumin 2.3 g/dL), edema, and declining eGFR** indicate **active glomerular disease**, clinically consistent with **lupus nephritis**.<br> - UPCR rising from **3.71 to 7.17 g/gCr** suggests severe and worsening renal inflammation/injury.<br><br>- **Kidney biopsy** is decisive:<br> - **Diffuse proliferative glomerulonephritis with endocapillary proliferation and wire-loop lesions** is classic for **active class IV lupus nephritis**.<br> - **Full-house immunofluorescence** and **subendothelial/mesangial/subepithelial electron-dense deposits** confirm **immune-complex lupus nephritis**.<br> - **Fibrinoid necrosis in small vessels** indicates **renal vasculitis associated with lupus**.<br> - Absence of **luminal thrombi**, absence of schistocytes, normal platelets, and |

| | |
|---|---|
|  | normal ADAMTS13 argue **against TTP/TMA as the main process**.<br> - Negative ANCA makes primary ANCA-associated vasculitis less likely.<br><br>- **Anticardiolipin IgG positivity** is important because it suggests possible **antiphospholipid antibody positivity**, but biopsy did **not** show thrombotic microangiopathy or vascular thrombosis, so **antiphospholipid syndrome is not the main explanation here** based on current data.<br><br>**Particular points of interest**<br>- PRES in this case is most plausibly secondary to **severe hypertension and endothelial injury from active SLE/lupus nephritis**.<br>- The pathology shows **lupus nephritis with vasculitis**, which can itself contribute to severe hypertension and renal dysfunction.<br>- This is not just uncomplicated hypertension; it is **systemic autoimmune disease with major renal and neurologic target-organ involvement**. |
| With Static RAG and Mult-agent framework (Model C) | 5. **Interpretation of the test results**<br><br>- **BP 236/122 mmHg with retinal hemorrhages/exudates and MRI-proven PRES** confirms a true **hypertensive emergency with target-organ injury** involving the **brain, eyes, and kidneys**. The MRI pattern—**bilateral occipital/cerebellar FLAIR hyperintensity with increased ADC**—is classic for **vasogenic edema**, supporting **PRES** rather than infarction. The **minute occipital hemorrhages** indicate severe endothelial/vascular injury during the hypertensive crisis. The fact that symptoms and MRI abnormalities improved after BP lowering strongly supports **hypertension-triggered PRES**.<br><br>- The **butterfly rash**, **markedly low complement levels**, **positive ANA**, and **very high anti-dsDNA** indicate **active systemic lupus erythematosus** rather than isolated essential hypertension or a purely vascular disorder. These serologies fit well with **immune-complex disease activity**.<br><br>- **Proteinuria, hematuria, hypoalbuminemia (2.3 g/dL), edema, and reduced eGFR** indicate clinically significant **glomerular renal disease**. The worsening **UPCR from 3.71 to 7.17 g/gCr** and decline in kidney function after admission support **active progressive nephritis**.<br><br>- The **kidney biopsy is diagnostic**:<br> - **Diffuse proliferative glomerulonephritis with endocapillary proliferation and wire-loop lesions** is characteristic of **active class IV lupus nephritis**.<br> - **Full-house immunofluorescence** and **electron-dense deposits in mesangial, subendothelial, and subepithelial areas** strongly support **immune-complex lupus nephritis**.<br> - **Small-vessel fibrinoid necrosis** shows **vascular involvement**. This can reflect **lupus-associated vascular injury/vasculitic change** in the full clinical context.<br> - **Mesangiolytic changes** may also be seen with **severe endothelial injury**, so the pathology should be interpreted together with the malignant hypertension and lupus activity.<br><br>- **Normal platelets, absence of schistocytes, normal ADAMTS13, and no intraluminal thrombi on biopsy** make **TTP and other primary thrombotic microangiopathies less likely**. The mild **LDH elevation** is nonspecific and does not establish microangiopathic hemolysis. |

| | |
|---|---|
| | - **Positive anticardiolipin IgG** suggests an **antiphospholipid antibody profile**, but **APS is not established** from the provided data because there is **no documented thrombosis**, **anti-β2GPI is negative**, and persistence over time is not shown.<br><br>**Key point:** The results support a **bidirectional relationship** between **active lupus nephritis with renal vascular injury** and **malignant hypertension**. Severe lupus renal disease likely contributed to the hypertension, while the hypertensive emergency also likely worsened renal and endothelial injury. PRES was precipitated by the hypertensive emergency.<br><br>**Changes from the initial interpretation:** I made the causal language more cautious, emphasizing a **bidirectional relationship** between nephritis/vascular injury and malignant hypertension rather than saying lupus nephritis alone drove the hypertension. I also clarified that **anticardiolipin positivity alone does not diagnose APS** and noted that **mesangiolytic change can occur with severe endothelial injury**. |
| **Phase II – Q6: Reconsidering the differential diagnosis** | |
| Baseline LLM (Model A) | **6. Updated diagnostic thinking / Working diagnosis**<br><br>The differential narrows substantially. The most likely unifying diagnosis is:<br><br>**Working diagnosis:**<br>**Systemic lupus erythematosus with active lupus nephritis, ISN/RPS class IV-G (A), accompanied by renal small-vessel vasculitis, presenting as hypertensive emergency with PRES and hypertensive retinopathy.**<br><br>Why this is now most likely:<br>- SLE is supported clinically and serologically.<br>- Kidney biopsy definitively shows **active diffuse proliferative lupus nephritis** with **vasculitic changes**.<br>- This renal inflammatory process plausibly caused the **worsening hypertension, edema, proteinuria, and renal dysfunction**, which then precipitated **PRES**.<br>- TMA, primary CNS disease, isolated malignant hypertension, and APS nephropathy become less likely as primary explanations because the biopsy and serology provide a clearer cause and there is no convincing evidence of thrombotic microangiopathy. |
| With Static RAG (Model B) | **6. Updated diagnostic thinking / Working diagnosis**<br><br>The differential diagnosis narrows substantially after serology, MRI, and kidney biopsy.<br><br>**Most likely working diagnosis:**<br>**Systemic lupus erythematosus with active lupus nephritis class IV-G (A) and renal vasculitis, complicated by hypertensive emergency causing PRES.**<br><br>**Why this is now most likely**<br>- SLE is supported by **malar rash, hypocomplementemia, positive ANA, strongly positive anti-dsDNA**, and renal findings.<br>- The kidney biopsy definitively shows **active diffuse proliferative lupus nephritis** with **vasculitic vascular lesions**.<br>- PRES is explained by the combination of **severe hypertension** and **systemic |

| | |
|---|---|
| | endothelial injury** in active lupus.<br>- TTP/TMA became less likely because there are **no schistocytes, no thrombocytopenia, normal ADAMTS13, and no biopsy thrombi**.<br>- Antiphospholipid syndrome remains a consideration in the background because of anticardiolipin positivity and prior oral contraceptive use, but current evidence does **not** make it the primary diagnosis. |
| With Static RAG and Mult-agent framework (Model C) | 6. **Updated diagnostic thinking / Working diagnosis**<br><br>### **Most likely diagnosis**<br>**Active systemic lupus erythematosus with biopsy-proven class IV-G (A) lupus nephritis with small-vessel fibrinoid necrosis/vascular involvement, complicated by hypertensive emergency causing PRES and hypertensive retinopathy.**<br><br>### **Why this is now the leading diagnosis**<br>- **SLE is established** clinically and serologically.<br>- **Kidney biopsy definitively shows active diffuse proliferative lupus nephritis**.<br>- The renal disease explains the **proteinuria, hematuria, edema, hypoalbuminemia, and reduced eGFR**.<br>- The **hypertensive emergency** explains the **PRES** and **hypertensive retinopathy**, and the improvement after BP control supports this.<br>- The renal vascular lesion is best described cautiously as **small-vessel fibrinoid necrosis/vascular involvement** in active lupus, rather than overcommitting to a single vascular mechanism beyond what the biopsy proves.<br><br>### **Less likely alternatives still in the differential**<br>- **Primary TMA/TTP:** less likely because platelets are normal, schistocytes are absent, ADAMTS13 is normal, and biopsy showed no thrombi.<br>- **APS nephropathy:** remains a **background consideration** because anticardiolipin IgG is positive, but it is **not established** and is not the main explanation of the current renal lesion.<br>- **Renovascular hypertension/fibromuscular dysplasia:** now much less likely as the unifying diagnosis given the lupus-specific serology and biopsy.<br><br>**Changes from the initial working diagnosis:** I refined the wording from “lupus nephritis with renal vasculitis” to **“class IV-G (A) lupus nephritis with small-vessel fibrinoid necrosis/vascular involvement”** to better match the pathology without overstating certainty. I also explicitly state that **PRES was caused by the hypertensive emergency**, not directly by SLE itself. |
| **Phase II – Q7: Treatment planning** | |
| Baseline LLM (Model A) | **7. Treatment plan**<br><br>### A. Immediate/ongoing supportive management<br>- **Strict blood pressure control**<br>- Continue appropriate antihypertensives to prevent recurrent PRES and further kidney injury.<br>- Rationale: BP reduction already improved neurologic symptoms and MRI lesions.<br>- Reasonable agents include **long-acting calcium channel blocker** plus **ARB** if tolerated and renal function/potassium permit.<br><br>- **General supportive care**<br>- Salt restriction, edema management, close monitoring of urine protein, creatinine, electrolytes, and neurologic status. |

| | |
|---|---|
|  | ### B. Induction therapy for active class IV lupus nephritis<br>- **High-dose glucocorticoids**<br> - Typically pulse **IV methylprednisolone** followed by high-dose oral prednisone.<br> - Rationale: rapid suppression of active immune-mediated glomerular and vascular inflammation.<br><br>- **Add a major immunosuppressive agent for induction**<br> - **Mycophenolate mofetil (MMF)** or **intravenous cyclophosphamide** are standard evidence-based options for class III/IV lupus nephritis.<br> - In this patient, given **class IV-G (A)** with worsening renal function and vasculitic changes, many clinicians would favor **aggressive induction**; **cyclophosphamide** may be particularly considered in severe disease, while **MMF** is also a standard first-line option.<br> - Choice depends on disease severity, fertility concerns, adverse-effect profile, and local practice.<br><br>- **Hydroxychloroquine**, unless contraindicated<br> - Rationale: recommended in SLE because it improves overall disease control and reduces flares.<br><br>### C. Adjunctive renoprotective therapy<br>- **Continue RAAS blockade** if tolerated<br> - Helps reduce proteinuria and protect kidney function.<br>- **Monitor for nephrotic-range complications**<br> - Hypoalbuminemia and heavy proteinuria increase risk of thrombosis and infection.<br><br>### D. Considerations related to antiphospholipid antibodies and contraception<br>- **Stop estrogen-containing oral contraceptive**<br> - Rationale: lupus plus anticardiolipin positivity increases thrombotic risk; estrogen is undesirable in this setting.<br>- **Anticoagulation is not automatically indicated**<br> - Because no thrombosis or APS nephropathy is proven from the provided information.<br> - If persistent antiphospholipid antibody positivity is confirmed and/or thrombosis occurs, management would need reassessment.<br><br>### E. Alternatives / escalation options<br>- If response is inadequate or toxicity limits treatment:<br> - Switch between **MMF** and **cyclophosphamide**<br> - Consider **rituximab** or **calcineurin inhibitor-based regimens** in selected refractory cases<br>- If PRES recurs or neurologic status worsens:<br> - Reassess BP control, exclude seizures, hemorrhage, and other CNS lupus complications.<br><br>Overall, the priority is **combined control of hypertensive emergency and prompt induction treatment for severe active lupus nephritis with vasculitic involvement**, because this is the most likely driver of both the renal disease and the PRES. |
| With Static RAG (Model B) | **7. Treatment plan**<br><br>### A. Immediate/acute management<br>1. **Strict blood pressure control** |

- Continue appropriate antihypertensive therapy to prevent recurrence/worsening of **PRES**, intracranial hemorrhage, and further renal injury.
- The observed response to **nicardipine/nifedipine plus ARB** is appropriate.
- Rationale: PRES treatment depends primarily on correcting the trigger, here **severe hypertension**.

2. **Stop estrogen-containing oral contraceptive**
- Rationale: it may increase **thrombotic risk**, especially with **anticardiolipin antibody positivity**.

#### B. Disease-directed therapy for lupus nephritis class IV-G (A) with vasculitis
1. **Induction immunosuppression**
- **High-dose glucocorticoids** (typically pulse methylprednisolone followed by oral prednisone/prednisolone) plus
- **Either mycophenolate mofetil (MMF) or intravenous cyclophosphamide**
- Rationale: Class IV active lupus nephritis requires prompt induction therapy to preserve kidney function and suppress systemic inflammation.

2. **Choice of agent**
- Given **severe active proliferative nephritis**, worsening kidney function, and **vasculitic lesions**, **cyclophosphamide-based induction** is a strong option.
- **MMF** is also evidence-based and may be considered depending on fertility concerns, toxicity profile, and treating team preference.
- Rationale: both are standard-of-care induction options for proliferative lupus nephritis.

3. **Hydroxychloroquine**
- Unless contraindicated, this should be added as background SLE therapy.
- Rationale: improves lupus control and reduces flares.

#### C. Supportive renal management
1. **RAAS blockade** if tolerated
- Continue ARB cautiously as renal function and potassium allow.
- Rationale: helps reduce **proteinuria** and supports BP control.

2. **Edema/proteinuria management**
- Salt restriction and diuretics as clinically needed.
- Rationale: symptomatic management of nephritic/nephrotic features.

3. **Monitoring**
- Serial **creatinine/eGFR, urine protein, urine sediment, complement, anti-dsDNA, CBC, liver function, and BP**.
- Rationale: assesses treatment response and drug toxicity.

#### D. Thrombosis/APS considerations
1. **Reassess antiphospholipid antibodies later**
- A single positive anticardiolipin test does not establish APS.
- Rationale: persistent positivity is required for classification, and there is no current biopsy evidence of thrombotic nephropathy.

2. **Anticoagulation**
- Not routinely indicated solely from the provided data, especially with **minute cerebral hemorrhage on MRI**.
- Rationale: risk-benefit does not favor empiric anticoagulation without confirmed

| | |
|---|---|
|  | thrombosis/APS indication.<br><br>### E. Alternative/adjunctive options<br>- If refractory or intolerant to standard induction:<br> - **Rituximab** may be considered in selected cases.<br> - **Calcineurin inhibitor-based combinations** may be considered in some lupus nephritis settings, though caution is needed with hypertension.<br>- For maintenance after response:<br> - **MMF or azathioprine**, with tapering glucocorticoids.<br><br>**Overall treatment goal:**<br>Control the **hypertensive emergency/PRES**, then aggressively treat **active class IV lupus nephritis with vasculitis** to preserve renal function, reduce proteinuria, prevent relapse, and limit further systemic organ damage. |
| With Static RAG and Mult-agent framework (Model C) | 7. **Treatment plan**<br><br>### A. Immediate and ongoing management of hypertensive emergency / PRES<br>1. **Careful blood pressure control**<br> - Continue strict BP control because this is essential for **PRES recovery**, prevention of recurrent cerebral injury, retinal protection, and kidney protection.<br> - The acute use of **IV nicardipine** followed by oral **nifedipine** was appropriate.<br> - BP should be lowered **gradually rather than excessively fast** to avoid ischemic complications.<br> - Once stable, continuing a regimen such as **dihydropyridine calcium channel blocker plus ARB** is reasonable, with close monitoring.<br><br>2. **Neurologic and ophthalmologic follow-up**<br> - Monitor for recurrence of headache, visual symptoms, seizures, or worsening neurologic signs.<br> - Follow retinal findings and visual recovery as indicated.<br><br>### B. Disease-specific treatment for active class IV lupus nephritis<br>3. **Induction immunosuppressive therapy**<br> - This is **organ-threatening proliferative lupus nephritis**, so standard evidence-based induction is:<br> - **High-dose glucocorticoids**<br> - often **pulse methylprednisolone** first if clinically appropriate, followed by **oral prednisone**, plus<br> - either **mycophenolate mofetil (MMF)** or **IV cyclophosphamide**.<br><br>4. **Choosing MMF vs cyclophosphamide**<br> - **MMF** is an accepted first-line option for proliferative lupus nephritis.<br> - **Cyclophosphamide** is also appropriate and may be favored in **more severe/aggressive disease** or if vascular involvement is clinically concerning.<br> - The choice should consider **severity, fertility goals, infection risk, tolerance, and kidney function**.<br><br>5. **Hydroxychloroquine**<br> - Should generally be used unless contraindicated because it reduces SLE activity and future flares.<br> - Arrange **baseline ophthalmologic assessment** and dose appropriately for body weight and renal function. |

#### C. Supportive kidney and general care
6. **Renoprotective measures**
  - **RAS blockade** may help reduce proteinuria, but because kidney function has been unstable, **creatinine and potassium must be monitored closely**.
  - Salt restriction and edema management are appropriate.
  - Dose-adjust medications according to kidney function.

7. **Monitoring disease activity and treatment toxicity**
  - Serial follow-up of:
    - **Creatinine/eGFR**
    - **Urinalysis and UPCR**
    - **CBC**
    - **Liver function**
    - **Complements and anti-dsDNA**
    - **Blood pressure**
  - These help assess both renal response and adverse effects of therapy.

#### D. Prevention and safety with immunosuppression
8. **Steroid and immunosuppression safety measures**
  - Consider **bone protection** and **GI protection** when indicated with high-dose steroids.
  - Review **vaccination status** and infection risk before/while giving major immunosuppression.
  - Consider **Pneumocystis prophylaxis** when risk is high based on steroid dose and immunosuppressive regimen.
  - If cyclophosphamide is used, discuss **fertility/gonadal toxicity** and provide appropriate counseling.

9. **Contraception and estrogen exposure**
  - Stop the **estrogen-containing oral contraceptive**, given severe hypertension and antiphospholipid antibody positivity.
  - Provide **safe contraception counseling**, especially before starting teratogenic agents such as MMF or cyclophosphamide.

#### E. Antiphospholipid antibody follow-up
10. **Repeat antiphospholipid antibody testing**
  - Repeat at the appropriate interval to assess persistence.
  - **Anticoagulation is not automatically indicated** on the current data because there is **no documented thrombosis** and no thrombotic lesion on biopsy.

#### F. Longer-term plan
11. **Maintenance therapy after induction**
  - If remission is achieved, typical maintenance would be **MMF or azathioprine**, with steroid taper and continued hydroxychloroquine.

12. **Alternatives if refractory or intolerant**
  - If response is inadequate or treatment is not tolerated, alternatives may include **rituximab**, **calcineurin inhibitor-based regimens**, **belimumab**, or **voclosporin**, depending on clinical context and availability.

**Overall rationale:** The priority is to continue controlling the **hypertensive emergency/PRES** while promptly treating the underlying **active proliferative lupus nephritis with vascular involvement**, because this is the main driver of

| | |
|---|---|
| | ongoing renal risk and future morbidity.<br><br>**Changes from the initial treatment plan:** I added more explicit **guideline-based supportive care and safety measures**, including **gradual BP targets**, **pulse steroids then oral steroids if appropriate**, **MMF vs cyclophosphamide selection based on severity/fertility/infection risk**, **baseline eye assessment for hydroxychloroquine**, **close creatinine/potassium monitoring with ARB use**, and **bone/GI protection, infection prophylaxis, vaccination review, and contraception/fertility counseling**. |
| **Phase III – Q8: Evaluation of treatment** | |
| Baseline LLM (Model A) | 8. **Monitoring effectiveness, risks, and countermeasures**<br><br>**Effectiveness**<br>- **Blood pressure:** frequent measurement during the acute phase, then regular outpatient monitoring. Goal is stable control to prevent recurrent PRES and further kidney injury.<br>- **Neurologic/ophthalmologic status:** monitor headache, visual symptoms, and consider repeat brain MRI and eye evaluation if symptoms recur or fail to normalize.<br>- **Renal response:** serial **serum creatinine/eGFR**, **urinalysis**, and **urine protein quantification (UPCR)**. These are key markers because lupus nephritis activity and treatment response are reflected by renal function, hematuria, and proteinuria.<br><br>- **SLE activity:** follow **complement levels (C3/C4/CH50)** and **anti-dsDNA titers**, together with clinical findings such as rash and edema.<br>- **General status:** body weight and edema to assess volume status and nephrotic activity.<br><br>**Safety / specific risks to watch for**<br>- **High-dose steroids (mPSL/PSL):** infection, hyperglycemia, hypertension worsening, psychiatric symptoms, gastrointestinal bleeding, osteoporosis.<br> - **Countermeasures:** regular CBC and glucose checks, monitor BP closely, screen for fever/infectious symptoms, use gastric and bone protection as appropriate, and taper steroids as disease control allows.<br>- **MMF:** leukopenia, liver dysfunction, gastrointestinal toxicity, infection, teratogenicity.<br> - **Countermeasures:** periodic **CBC** and **liver tests**, assess GI tolerance, counsel strict pregnancy avoidance, and reduce/hold MMF if significant cytopenia or infection occurs.<br>- **Belimumab:** infusion reactions and infection risk.<br> - **Countermeasures:** monitor during/after infusion and watch for delayed infectious complications.<br>- **Renal disease-related risks:** persistent proteinuria, CKD progression, edema.<br> - **Countermeasures:** continue renoprotective BP control, RAAS blockade if tolerated, and adjust immunosuppression according to renal response.<br>- **Possible TMA/vascular complication:** occasional fragmented RBCs raise concern, even without thrombocytopenia.<br> - **Countermeasures:** trend **CBC, platelets, LDH, haptoglobin, bilirubin, creatinine, urinalysis**, and repeat peripheral smear if hemolysis or kidney function worsens. Reassess for malignant hypertension-related TMA, APS-associated microangiopathy, or active vascular lupus.<br>- **Thrombosis risk:** SLE, heavy proteinuria, and anticardiolipin positivity increase risk. |

| | |
|---|---|
| | - **Countermeasures:** monitor for clinical thrombosis; avoid estrogen exposure; reassess antiphospholipid antibody profile if clinically indicated. |
| With Static RAG (Model B) | **8. List specific measures for evaluating treatment effectiveness and potential side effects or complications, and explain the monitoring and management plan for them.**<br><br>### A. Monitoring treatment effectiveness<br>- **Blood pressure and neurologic/visual symptoms**<br>- Follow home/clinic BP, headache, and visual symptoms.<br>- Goal: prevent **PRES recurrence** and further vascular/renal injury.<br>- If symptoms recur, reassess urgently with neurologic evaluation and brain imaging.<br><br>- **Renal response**<br>- Monitor **serum creatinine/eGFR**, **urinalysis**, urine sediment, and **UPCR/proteinuria trend** at regular intervals.<br>- Improvement in renal function and reduction of proteinuria indicate response; persistent proteinuria means only partial renal response and requires close follow-up.<br><br>- **SLE activity**<br>- Follow **complement levels (C3/C4/CH50)** and **anti-dsDNA antibody titers**, together with clinical findings such as rash and edema.<br>- Rising complement and falling anti-dsDNA support control of lupus activity.<br><br>### B. Safety monitoring and key risks<br>- **Glucocorticoids (mPSL/PSL)**<br>- Risks: infection, hyperglycemia, hypertension/fluid retention, psychiatric symptoms, gastrointestinal bleeding, osteoporosis.<br>- Monitoring: temperature/symptoms of infection, CBC, glucose, BP, body weight/edema.<br>- Countermeasures: gradual taper as disease allows; infection vigilance; bone and gastric protection according to standard practice.<br><br>- **MMF**<br>- Risks: leukopenia, liver dysfunction, gastrointestinal toxicity, infection, teratogenicity.<br>- Monitoring: **CBC**, liver enzymes, renal function, GI symptoms.<br>- Countermeasures: dose reduction/interruption if cytopenia or significant adverse effects occur; strict contraception counseling.<br><br>- **Belimumab**<br>- Risks: infusion reactions and infection.<br>- Monitoring: infusion-related symptoms and infectious symptoms during follow-up.<br>- Countermeasures: hold treatment and evaluate promptly if serious infection is suspected.<br><br>### C. Specific complications to watch for in this patient<br>- **Residual or worsening lupus nephritis**<br>- Because proteinuria did not remit, continue close renal monitoring.<br>- If proteinuria or kidney function worsens, reconsider disease activity, adherence, drug dose adequacy, and need to revise immunosuppression.<br><br>- **TMA / secondary microangiopathic process**<br>- Fragmented RBCs were intermittently present, although thrombocytopenia was absent. |

| | |
|---|---|
| | - Monitor **CBC/platelets**, smear, **LDH**, hemoglobin, creatinine, and BP.<br>- Countermeasure: if hemolysis, thrombocytopenia, or rapid renal deterioration appears, promptly reassess for malignant hypertension-related TMA, APS-associated nephropathy, or other causes.<br><br>- **Thrombotic risk**<br>- She has **anticardiolipin antibody positivity** and prior estrogen exposure.<br>- Monitor clinically for venous/arterial thrombosis; avoid estrogen-containing agents.<br>- Repeat antiphospholipid antibody testing later if clinically indicated. |
| With Static RAG and Mult-agent framework (Model C) | **8. List specific measures for evaluating treatment effectiveness and potential side effects or complications, and explain the monitoring and management plan for them.**<br><br>### A. Monitoring treatment effectiveness<br><br>- **Blood pressure control**<br>- Monitor BP closely because control of hypertension is essential to prevent **recurrent PRES**, further retinal injury, and worsening kidney damage.<br>- In the early post-discharge period, BP should be checked frequently, including home BP if feasible.<br>- Recurrence of severe hypertension should prompt reassessment of adherence, volume status, renal disease activity, and antihypertensive regimen.<br><br>- **Renal response**<br>- Follow **serum creatinine/eGFR**, **urinalysis**, and **UPCR** serially.<br>- Improvement in **renal function, hematuria, edema, and proteinuria** indicates response to treatment.<br>- Because **proteinuria did not reach remission**, persistent proteinuria should be interpreted carefully as possibly reflecting **ongoing active lupus nephritis or chronic residual kidney damage**, so continued close follow-up is required.<br><br>- **SLE activity**<br>- Monitor **complement levels (C3/C4/CH50)** and **anti-dsDNA antibody** as laboratory markers of lupus activity.<br>- Also assess clinical improvement in **butterfly rash, edema, and other systemic symptoms**.<br><br>- **Neurologic and ophthalmologic recovery**<br>- Reassess for recurrence of **headache, visual disturbance, confusion, or seizures**.<br>- Repeat **brain MRI** or ophthalmologic assessment if neurologic or visual symptoms recur or do not continue to improve.<br><br>### B. Safety monitoring and complications<br><br>- **Glucocorticoid toxicity (mPSL/PSL)**<br>- Major risks: **infection, hyperglycemia, hypertension worsening, psychiatric symptoms, gastrointestinal injury, osteoporosis**.<br>- Monitoring: regular symptom review, **BP**, glucose, and general infection surveillance.<br>- Management: taper steroids as disease control permits; consider **GI protection** and **bone protection** as appropriate.<br><br>- **MMF toxicity**<br>- Major risks: **leukopenia, infection, liver dysfunction, gastrointestinal symptoms, |

teratogenicity**.
  - Monitoring: regular **CBC** and **liver function tests**.
  - Management: dose reduction or interruption if significant cytopenia or toxicity develops; ensure **effective contraception**.

- **Belimumab toxicity**
  - Major risks: **infection, infusion reaction, hypersensitivity**, and rarely **depression or mood change**.
  - Monitoring: observe during/after infusion, monitor for infectious symptoms, and assess mood/neuropsychiatric symptoms during follow-up.

- **Persistent or evolving microangiopathic/endothelial injury**
  - Because **fragmented red blood cells (0.5–10%) were intermittently detected**, ongoing endothelial injury or secondary **TMA-like process** cannot be ignored even though **thrombocytopenia was absent** and initial studies did not support primary TMA.
  - Monitoring: serial **CBC/platelets**, peripheral smear, **LDH**, bilirubin, haptoglobin, **creatinine**, and urinalysis.
  - Management trigger: if **schistocytes increase**, **LDH rises**, **platelets fall**, or **kidney function worsens rapidly**, urgently reassess for **secondary TMA, malignant hypertension-related endothelial injury, or APS-related vascular disease**.

- **Infection risk from combined immunosuppression**
  - Risk is increased by **high-dose steroids + MMF + belimumab**.
  - Monitoring: check for fever and localizing symptoms at every visit.
  - Management: maintain a low threshold for infectious evaluation and treatment; review preventive measures as appropriate.

#### C. Practical follow-up plan

- **Early after discharge**
  - Frequent outpatient follow-up is needed because she had **class IV lupus nephritis**, recent **PRES**, persistent **proteinuria**, and intermittent **fragmented red blood cells**.
  - At follow-up, check:
    - **Blood pressure**
    - **CBC with platelets**
    - **Creatinine/eGFR, electrolytes**
    - **Liver function tests**
    - **Urinalysis and UPCR**
    - **Complements and anti-dsDNA**

- **How treatment success is judged**
  - Judge by **trends over time**, not a single value.
  - Improvement would include:
    - stable or improved **BP**
    - improving **creatinine/eGFR**
    - falling **UPCR**
    - less active urine sediment
    - improving **complements/anti-dsDNA**
    - no recurrence of **PRES-related symptoms**
  - If proteinuria improves only partially, continue therapy with cautious steroid taper while monitoring closely.

| | |
|---|---|
| | - If proteinuria plateaus or worsens, reassess whether this reflects **persistent active nephritis versus chronic damage**.<br><br>**Changes from the initial version:**<br>I made the monitoring plan more specific by adding **follow-up intensity**, clearer treatment response measures, and explicit recognition that **persistent proteinuria may reflect either ongoing activity or chronic damage**. I also strengthened the section on intermittent **fragmented red blood cells**, defining them as a reason for repeated reassessment for **TMA or malignant hypertension-related endothelial injury**, even without thrombocytopenia. |
| **Phase III – Q9: Formulating plans for worsening conditions** | |
| Baseline LLM (Model A) | 9. **If the condition does not improve or changes suddenly: reassessment and action plan**<br><br>- **First, identify what is worsening:** BP, neurologic symptoms, kidney function, proteinuria, hemolysis markers, platelets, or systemic lupus features.<br>- **Reconsider the cause rather than assuming refractory lupus nephritis alone.** Main possibilities include:<br>1. **Insufficient control of active lupus nephritis**<br>2. **Recurrent/uncontrolled hypertension causing recurrent PRES**<br>3. **TMA or APS-related vascular disease**, especially if schistocytes increase, LDH rises, anemia worsens, or platelets fall<br>4. **Drug toxicity or infection** related to immunosuppression<br>5. Less likely, another superimposed renal or neurologic process<br><br>**Concrete actions**<br>- **If neurologic symptoms recur:** urgent BP reassessment, focused neurologic exam, and repeat brain imaging to evaluate recurrent PRES, hemorrhage, or another CNS event.<br>- **If kidney function or proteinuria worsens:** repeat urine studies, creatinine, complement, anti-dsDNA, hemolysis panel, and review BP control and medication adherence/tolerance.<br>- **If hemolysis/TMA is suspected:** repeat smear, CBC, platelets, LDH, haptoglobin, bilirubin, coagulation studies, and reassess for APS/TMA; involve hematology if needed.<br>- **If infection is possible:** obtain cultures/imaging as indicated and reduce or hold immunosuppression depending on severity.<br>- **If response is inadequate despite appropriate treatment:** reconsider the pathologic driver and revise therapy, for example by intensifying or changing induction immunosuppression based on overall clinical reassessment.<br>- **Throughout:** make treatment changes promptly and multidisciplinary, because this patient is at risk for irreversible neurologic and renal damage if relapse or an alternative vascular process is missed. |
| With Static RAG (Model B) | **9. Explain the thought process and action plan for rapidly reassessing the cause in response to changes in the patient's condition, and for flexibly revising the diagnosis and treatment plan.**<br><br>### A. If the condition does not improve as expected<br>- First, determine whether the problem is:<br>1. **Persistent active lupus nephritis**,<br>2. **Treatment toxicity/infection**, or<br>3. **Another overlapping vascular process** such as TMA or APS-related disease. |

| | |
|---|---|
| | - Reassess with:<br>- **BP trend**<br>- **Creatinine/eGFR, UPCR, urinalysis**<br>- **CBC with platelets, smear, LDH**<br>- **Complement and anti-dsDNA**<br>- Review of medication adherence and tolerance<br><br>- Interpretation:<br>- **Low complement/high anti-dsDNA with active urine findings** suggests ongoing lupus activity.<br>- **Cytopenia, hemolysis, rising LDH, schistocytes, thrombocytopenia** suggests TMA.<br>- **Fever or inflammatory symptoms** raises concern for infection rather than flare.<br><br>### B. If the condition suddenly changes<br>- **Neurologic or visual worsening / severe headache**<br>- Treat as emergency: check BP immediately and repeat brain imaging to evaluate for **recurrent PRES, hemorrhage, or stroke**.<br><br>- **Rapid renal deterioration or increased schistocytes**<br>- Urgently reassess for **malignant hypertension-related TMA, APS nephropathy, or progressive lupus nephritis**.<br>- Repeat hemolysis studies, platelet count, and consider whether repeat kidney-directed evaluation is needed.<br><br>- **Fever or systemic decline during immunosuppression**<br>- Assume infection must be excluded first.<br>- Obtain appropriate cultures/imaging and consider temporarily modifying immunosuppression depending on severity.<br><br>### C. How to revise treatment<br>- If evidence supports **persistent active lupus nephritis despite current therapy**, consider intensifying or changing induction therapy based on response and toxicity.<br>- If findings instead support **TMA/APS or uncontrolled hypertension**, redirect treatment toward that mechanism rather than simply escalating lupus therapy.<br>- If adverse effects are the main issue, reduce or switch the responsible drug while maintaining control of lupus as safely as possible.<br><br>**Core principle:** in this patient, any lack of improvement or abrupt change should trigger rapid reassessment of three competing categories—**active SLE/lupus nephritis, vascular complications (hypertension/TMA/APS), and treatment-related infection or toxicity**—and management should be revised according to the dominant mechanism. |
| With Static RAG and Mult-agent framework (Model C) | **9. Explain the thought process and action plan for rapidly reassessing the cause in response to changes in the patient's condition, and for flexibly revising the diagnosis and treatment plan.**<br><br>### A. Reassessment framework<br><br>If the patient does not improve as expected, I would reconsider whether the problem is: |

1. **Persistent active class IV lupus nephritis**,
2. **Residual/chronic kidney damage after severe nephritis and hypertensive injury**,
3. **Recurrent or insufficiently controlled hypertension causing renewed endothelial injury/PRES risk**, or
4. **A superimposed process**, especially **secondary TMA, APS-related vascular disease, infection, or drug toxicity**.

This framework is especially important here because the patient had:
- **biopsy-proven class IV lupus nephritis**
- prior **PRES**
- **positive anticardiolipin antibody**
- intermittent **fragmented red blood cells**, despite no thrombocytopenia

#### B. Immediate reassessment steps

If her condition worsens, I would promptly re-evaluate:
- **BP trend**
- **Serum creatinine/eGFR**
- **Urinalysis, urine sediment, and UPCR**
- **CBC with platelet count**
- **Hemolysis markers**: smear, LDH, bilirubin, haptoglobin
- **Complements and anti-dsDNA**
- **Medication adherence and adverse effects**
- Symptoms suggesting **infection**

#### C. Concrete triggers that should change management

- **Recurrent headache, visual symptoms, confusion, or seizure**
  - Treat urgently as possible **recurrent hypertensive emergency/PRES**
  - Immediate BP reassessment and urgent neurologic evaluation, with repeat **brain imaging** as indicated

- **Worsening creatinine, increasing proteinuria, or more active urine sediment**
  - Suggests inadequate control of **active lupus nephritis**, uncontrolled hypertension, or superimposed vascular injury
  - Reassess whether the current immunosuppressive regimen is sufficient

- **Increasing schistocytes, rising LDH, or new thrombocytopenia**
  - Raises urgent concern for **secondary TMA or APS-related vascular disease**
  - This should trigger prompt hematologic reassessment and review of the kidney pathology/clinical course

- **Fever, leukopenia, new cytopenia, or systemic symptoms**
  - Consider **infection** or **drug toxicity** before escalating immunosuppression

#### D. How the diagnosis and treatment plan would be revised

- **If hypertension is again uncontrolled**
  - Intensify antihypertensive treatment and evaluate for renewed end-organ injury.

- **If active lupus nephritis appears to persist**
  - Reassess adequacy of induction therapy and consider modifying or intensifying immunosuppression based on efficacy and safety.

| | |
|---|---|
|  | - **If TMA or APS-related disease becomes more likely**<br> - Re-evaluate hematologic data and vascular pathology urgently, because this would significantly affect management.<br><br>- **If infection or medication toxicity is suspected**<br> - Temporarily adjust immunosuppressive therapy as needed while treating the alternative cause.<br><br>- **If the picture remains unclear**<br> - Reconsider the original assumptions and, if necessary, repeat kidney or other organ evaluation to distinguish **active inflammation, chronic damage, or vascular/endothelial complications**.<br><br>### E. Core principle<br><br>The key principle is to **reassess the syndrome first, then the mechanism**:<br>- first determine what is worsening: **BP, kidney function, urine findings, CNS symptoms, or hematologic markers**<br>- then determine whether the cause is **active SLE**, **chronic damage**, **recurrent hypertensive endothelial injury/PRES**, **TMA/APS-related disease**, **infection**, or **treatment toxicity**<br>- and revise treatment accordingly rather than assuming all deterioration is due to lupus nephritis alone<br><br>**Changes from the initial version:**<br>I made the reassessment plan more concrete by defining specific findings that would trigger immediate action in this patient: **recurrent neurologic symptoms**, **worsening creatinine/proteinuria/active sediment**, and **increasing schistocytes/LDH or falling platelets**. I also more explicitly integrated her actual risk factors—**class IV lupus nephritis, prior PRES, anticardiolipin positivity, and intermittent fragmented red blood cells**—and emphasized that **infection and drug toxicity must be excluded before intensifying immunosuppression**. |

| **Case 3: Anti-factor H antibody-positive C3 glomerulonephritis secondary to poststreptococcal acute glomerulonephritis with diabetic nephropathy** [11] | |
|---|---|
| **Phase I – Q1: Summarizing medical problems** | |
| Baseline LLM (Model A) | 1. **Problem representation**<br><br>A **49-year-old Japanese man** with a **7-year history of type 2 diabetes complicated by diabetic retinopathy and prior diabetic foot gangrene**, and previously documented **proteinuria (1.77 g/day) with near-normal creatinine**, now presents with a **subacute decline in renal function over days to weeks** after a **febrile episode treated as UTI**, with **serum creatinine rising rapidly from about 1.0 to 4.3 mg/dL** and new **bilateral leg edema**.<br>Key features are:<br>- Established **microvascular diabetic complications**, making diabetic kidney disease plausible<br>- But **rapid AKI/subacute kidney injury** is atypical for uncomplicated diabetic nephropathy<br>- **Pyuria/preceding infection** raises concern for urinary tract/interstitial/infectious renal disease<br>- **Edema** suggests significant proteinuria and/or reduced kidney function |

| | |
|---|---|
| With Static RAG (Model B) | ## 1. Problem representation<br>A **49-year-old Japanese man** with a **7-year history of type 2 diabetes complicated by retinopathy and prior diabetic foot gangrene**, and previously documented **proteinuria (1.77 g/day) with near-normal creatinine (0.9–1.2 mg/dL)**, now presents with a **1-month subacute decline in renal function** after a **febrile episode treated as UTI with ceftriaxone**, with **serum creatinine rising rapidly to 4.3 mg/dL within days** and **new bilateral leg edema**.<br><br>### Key points<br>- He already had **microvascular diabetic complications** and prior proteinuria, so **underlying diabetic kidney disease (DKD)** is plausible.<br>- However, the course is **too rapid** for typical isolated DKD progression.<br>- The combination of **recent fever/pyuria**, **AKI**, and **edema** raises concern for a **superimposed intrinsic renal process** rather than simple chronic diabetic nephropathy alone. |
| With Static RAG and Mult-agent framework (Model C) | ## 1) Problem representation<br><br>A **49-year-old man** with **type 2 diabetes complicated by retinopathy and prior diabetic foot gangrene**, and prior evidence of **proteinuric chronic kidney disease likely due to diabetic kidney disease** (**proteinuria 1.77 g/day** with **creatinine 0.9–1.2 mg/dL** 3 years earlier), now presents with a **subacute-to-acute severe decline in renal function**: after a **febrile illness with pyuria** treated empirically with **ceftriaxone**, his **creatinine rose rapidly to 4.3 mg/dL within days** and he developed **bilateral leg edema**.<br><br>This is best framed as **probable underlying diabetic kidney disease with a superimposed acute kidney injury**, rather than diabetic nephropathy alone. The key diagnostic question is whether the new process is **glomerular, tubulointerstitial, infectious, obstructive, or hemodynamic/ischemic**. |
| **Phase I – Q2: Differential diagnoses and rationale** | |
| Baseline LLM (Model A) | 2. **Differential diagnosis**<br><br>**Top 3 differential diagnoses**<br>1. **Acute interstitial nephritis (AIN), possibly drug-induced (e.g., ceftriaxone) or infection-associated**<br>2. **Rapid progression of diabetic kidney disease with superimposed acute kidney injury**<br>3. **Infection-related glomerulonephritis / other nephritic glomerulonephritis**<br><br>**Most likely: Acute interstitial nephritis or another superimposed intrinsic AKI on chronic diabetic kidney disease**<br>**Must not miss: Rapidly progressive glomerulonephritis (RPGN) and obstructive pyelonephritis/postrenal obstruction**<br><br>**Reasoning**<br>- **AIN**: The creatinine rose markedly within days after treatment of a febrile urinary illness. Pyuria supports tubulointerstitial inflammation, and antibiotic exposure is a classic trigger.<br>- **Diabetic kidney disease with superimposed AKI**: He already had substantial proteinuria and advanced diabetic complications, so underlying diabetic nephropathy is very likely. However, diabetic nephropathy alone usually does **not** cause such |

| | |
|---|---|
|  | a rapid rise in creatinine.<br>- **Glomerulonephritis**: Edema and renal dysfunction could reflect a glomerular process, especially if hematuria, active urine sediment, or heavy proteinuria is present. Infection-related GN is possible after a recent infection.<br>- **Must not miss**<br> - **RPGN** because delay risks irreversible kidney failure.<br> - **Obstruction/complicated pyelonephritis** because it is treatable and may present after presumed UTI. |
| With Static RAG (Model B) | ## 2. Differential diagnosis<br>### Top 3 differential diagnoses<br>1. **Acute interstitial nephritis (AIN), possibly drug-induced (including ceftriaxone) or infection-related**<br>2. **Rapid worsening of diabetic kidney disease with superimposed acute kidney injury**<br>3. **Infection-related glomerulonephritis / rapidly progressive glomerulonephritis (RPGN spectrum)**<br><br>### Most likely<br>**Acute interstitial nephritis (AIN)** is the most likely.<br><br>### Must not be missed<br>**Rapidly progressive glomerulonephritis (RPGN)** must not be missed.<br><br>### Reasoning<br>#### 1) AIN<br>- The patient had **fever and pyuria**, then received **ceftriaxone**, after which **creatinine rose rapidly over days**.<br>- This time course is compatible with **drug-associated AIN**.<br>- Pyuria can occur in both UTI and AIN, so the original urinary findings may not have represented simple infection alone.<br><br>#### 2) Diabetic kidney disease with AKI<br>- He has strong evidence of longstanding diabetic microvascular disease: **retinopathy, foot gangrene, prior significant proteinuria**.<br>- DKD likely forms the **background kidney disease**.<br>- However, **creatinine from ~1 mg/dL to 4.3 mg/dL within days** is atypically rapid for DKD alone, suggesting a superimposed insult.<br><br>#### 3) Infection-related GN / RPGN<br>- A preceding infectious episode followed by AKI and edema could indicate **glomerulonephritis**.<br>- Edema may reflect **worsening proteinuria and salt/water retention**.<br>- This is particularly important because **crescentic GN** requires urgent recognition and treatment. |
| With Static RAG and Mult-agent framework (Model C) | ## 2) Prioritized differential diagnosis<br><br>### Most important differential diagnoses<br>1. **Superimposed glomerular disease**<br> - Includes **rapidly progressive glomerulonephritis** and **infection-related glomerulonephritis**.<br> - Why high priority: the **rapid creatinine rise**, edema, and prior proteinuria raise concern for an active intrinsic renal process beyond baseline diabetic kidney disease. |

| | |
|---|---|
| | - Must not miss because delayed treatment may lead to irreversible kidney failure.<br><br>2. **Acute interstitial nephritis (AIN)**<br>- Could be **drug-related** or **infection-associated**.<br>- Why considered: AKI developed after a febrile illness and antibiotic exposure, and pyuria can be seen in AIN.<br>- However, it should **not be considered the single most likely diagnosis prematurely**, because pyuria and ceftriaxone exposure are nonspecific.<br><br>3. **Acute tubular injury (ATI/ATN)**<br>- Could follow sepsis/infection, ischemia, or nephrotoxic exposure.<br>- Why considered: the abrupt rise in creatinine over days is compatible with acute tubular injury, especially if there was systemic illness.<br><br>4. **Ongoing urinary infection/acute pyelonephritis**<br>- Why considered: fever and pyuria preceded the AKI.<br>- Important to confirm because infection can directly worsen kidney function and may coexist with another renal process.<br><br>5. **Obstructive uropathy**<br>- Why considered: obstruction is a reversible cause of AKI and should always be excluded early, especially when the diagnosis is uncertain.<br><br>### Background diagnosis<br>6. **Underlying diabetic kidney disease**<br>- Strongly supported by diabetic retinopathy, prior foot gangrene, and significant prior proteinuria.<br>- However, **diabetic kidney disease alone is unlikely to explain such an abrupt creatinine rise over days**. |
| **Phase I – Q3: Necessary physical examinations and reasons** | |
| Baseline LLM (Model A) | 3. **Additional history and physical examination**<br><br>**History**<br>- **Urinary symptoms**: dysuria, frequency, urgency, flank pain, weak stream, incomplete emptying<br>- To assess persistent UTI, pyelonephritis, or obstruction<br>- **Timing of ceftriaxone and other drugs**: NSAIDs, PPIs, herbal medicines, antibiotics<br>- To evaluate drug-induced AIN<br>- **Gross hematuria, foamy urine, oliguria**<br>- To distinguish glomerular disease, nephrotic-range proteinuria, or severe AKI<br>- **Systemic symptoms**: rash, arthralgia, fever recurrence, weight loss<br>- To support AIN, vasculitis, or systemic inflammatory disease<br>- **Recent skin/throat infections, endocarditis risk factors**<br>- To assess infection-related GN<br>- **Past kidney history, prior urinalyses, hypertension history**<br>- To define chronicity and baseline CKD<br><br>**Physical examination**<br>- **Volume status**: BP, JVP, lung crackles, degree of edema<br>- To assess severity of fluid overload and need for urgent management<br>- **Skin exam for rash**<br>- Suggestive of drug-induced AIN or vasculitis |

| | |
|---|---|
|  | - **Costovertebral angle tenderness / abdominal exam / bladder distention**<br> - For pyelonephritis or obstruction<br>- **Fundoscopy if available**<br> - Severe diabetic retinopathy supports diabetic kidney disease, though does not exclude non-diabetic disease<br>- **Cardiac exam and signs of infection**<br> - To look for endocarditis or other infectious sources |
| With Static RAG (Model B) | ## 3. Additional history and physical examination<br>### Additional history<br>1. **Urinary symptoms**: dysuria, frequency, urgency, flank pain, gross hematuria, decreased urine volume<br> - To distinguish **true UTI/pyelonephritis**, **obstruction**, and **glomerular disease**.<br><br>2. **Drug exposure history**: exact timing of ceftriaxone, NSAIDs, PPIs, herbal drugs, recent antibiotics or contrast<br> - To assess for **drug-induced AIN** or other nephrotoxins.<br><br>3. **Systemic symptoms**: rash, fever persistence, arthralgia, eosinophilia-related symptoms<br> - To support **AIN** or systemic inflammatory disease.<br><br>4. **Nephritic features**: cola-colored urine, foamy urine, sudden hypertension<br> - To evaluate for **GN** and degree of proteinuria.<br><br>5. **Volume status symptoms**: orthopnea, dyspnea, rapid weight gain<br> - To assess severity of **fluid overload**.<br><br>6. **Diabetes history**: glycemic control over time, neuropathy, known nephropathy, prior hypertension<br> - To estimate likelihood of **baseline DKD**.<br><br>### Physical examination<br>1. **Blood pressure, weight, volume status**<br> - Evaluate severity of AKI-related fluid retention and nephritic/nephrotic physiology.<br><br>2. **Skin examination for rash**<br> - Suggestive of **AIN** or vasculitis.<br><br>3. **Peripheral edema and jugular venous pressure**<br> - To characterize fluid overload.<br><br>4. **Costovertebral angle tenderness**<br> - To assess for ongoing **pyelonephritis**.<br><br>5. **Fundoscopy if available / confirmation of diabetic retinopathy history**<br> - Strong retinopathy supports **DKD background**, though does not exclude another kidney disease.<br><br>6. **Cardiopulmonary exam**<br> - To detect pulmonary edema or findings suggesting systemic vasculitis. |
| With | ## 3) Additional history and physical examination |

| | |
|---|---|
| Static RAG and Mult-agent framework (Model C) | ## Focused history<br>- **Urine output (oliguria/anuria)**<br>- Helps assess AKI severity and raises concern for obstruction or severe intrinsic renal injury.<br>- **Gross hematuria or cola-colored urine**<br>- Suggests glomerulonephritis.<br>- **Dysuria, frequency, urgency, flank pain**<br>- Supports ongoing UTI or pyelonephritis.<br>- **Medication and nephrotoxin exposure**<br>- Timing of **ceftriaxone**, and use of **NSAIDs, PPIs, other antibiotics, contrast, herbal medications**.<br>- Important for AIN or toxic tubular injury.<br>- **Systemic symptoms**<br>- **Rash, arthralgia, fever recurrence, sinus/pulmonary symptoms, neuropathy, weight loss**<br>- May suggest AIN, vasculitis, or systemic inflammatory disease.<br>- **Recent blood pressure change, headache, dyspnea, orthopnea, weight gain**<br>- Helps assess fluid overload, hypertensive renal injury, and heart failure.<br>- **Voiding difficulty, weak stream, retention history**<br>- Screens for postrenal obstruction.<br>- **Recent infections**<br>- Skin, throat, dental, foot, or other sources that could trigger infection-related GN.<br><br>## Focused physical examination<br>- **Vital signs, especially blood pressure and temperature**<br>- Hypertension may accompany glomerular disease or volume overload; fever suggests persistent infection/inflammation.<br>- **Volume status**<br>- Peripheral edema, JVP, lung crackles, daily weight.<br>- Evaluates fluid overload from AKI/nephrotic or nephritic processes.<br>- **Skin and joints**<br>- Rash, purpura, ulcers, synovitis may point to AIN or vasculitis.<br>- **Cardiopulmonary exam**<br>- Looks for pulmonary edema or heart failure.<br>- **Abdominal/bladder exam**<br>- Distended bladder suggests retention/obstruction.<br>- **Costovertebral angle tenderness**<br>- Supports pyelonephritis.<br>- **Funduscopic/diabetic complication review if available**<br>- Severe retinopathy supports baseline diabetic kidney disease, though not the acute process. |
| **Phase I – Q4: Plan for investigations/tests** | |
| Baseline LLM (Model A) | 4. **Testing plan**<br><br>1. **Urinalysis with microscopy**<br>- **Purpose**: Distinguish glomerular vs tubulointerstitial vs infectious process<br>- **Expected findings**:<br>- **AIN**: pyuria, WBC casts, mild-moderate proteinuria<br>- **GN**: hematuria, dysmorphic RBCs, RBC casts<br>- **Diabetic kidney disease**: albuminuria/proteinuria with relatively bland sediment |

2. **Urine protein quantification (spot protein/Cr or albumin/Cr; or 24-hour protein if needed)**
  - **Purpose**: Assess severity of proteinuria and nephrotic contribution to edema
  - **Expected findings**: Significant proteinuria likely; nephrotic-range proteinuria would broaden glomerular causes

3. **Blood tests: CBC, CRP, electrolytes, BUN/Cr, albumin, liver panel**
  - **Purpose**: Evaluate inflammation/infection, eosinophilia, renal severity, hypoalbuminemia
  - **Expected findings**:
    - Possible **eosinophilia** in AIN
    - **Low albumin** if heavy protein loss
    - Electrolyte abnormalities from AKI

4. **Renal ultrasound with bladder assessment**
  - **Purpose**: Exclude hydronephrosis/obstruction; assess kidney size and chronicity
  - **Expected findings**:
    - **Hydronephrosis** if postrenal cause
    - Normal-to-enlarged kidneys may be seen in diabetic kidney disease; small kidneys would suggest chronic irreversible disease

5. **Urine culture and blood cultures if clinically indicated**
  - **Purpose**: Confirm persistent or complicated urinary infection
  - **Expected findings**: Usually negative if infection resolved, but positive culture would support ongoing infection

6. **Serologic workup for glomerulonephritis if urine sediment is active**
  - Tests: **C3/C4, ANA, anti-dsDNA, ANCA, anti-GBM, hepatitis B/C**
  - **Purpose**: Evaluate immune-mediated or infection-related GN
  - **Expected findings**:
    - Often normal in AIN/diabetic kidney disease
    - Abnormal complement or positive autoantibodies would support specific GN

7. **Consider urine eosinophils only cautiously**
  - **Purpose**: Historically used for AIN, but limited accuracy
  - **Expected findings**: May be positive in AIN, but not reliable enough to confirm/exclude

8. **Kidney biopsy**
  - **Purpose**: Definitive diagnosis if cause remains unclear, especially because the rise in creatinine is rapid and atypical for diabetic nephropathy alone
  - **Expected findings**:
    - **AIN**: interstitial inflammatory infiltrates/tubulitis
    - **Diabetic kidney disease**: mesangial expansion, GBM thickening, nodular sclerosis
    - **GN**: disease-specific glomerular lesions

**Overall diagnostic approach:**
First confirm whether the urine sediment is **bland, inflammatory, or nephritic**, while simultaneously excluding **obstruction** and **ongoing infection**. Because this patient has known diabetic microvascular disease but an **unusually rapid renal decline**, a **superimposed non-diabetic kidney disease is strongly suspected**, and **kidney biopsy should be considered early** if initial studies do not clearly identify

| | |
|---|---|
| | a reversible cause. |
| With Static RAG (Model B) | ## 4. Tests to order<br>### 1) Urinalysis with microscopy<br>**Purpose:** Differentiate tubular/interstitial vs glomerular process.<br>**Expected findings:**<br>- **AIN**: pyuria, WBC casts, possibly mild hematuria/proteinuria<br>- **GN**: dysmorphic RBCs, RBC casts<br>- **DKD**: albuminuria/proteinuria, usually bland sediment unless superimposed disease<br><br>### 2) Quantify proteinuria (spot protein-creatinine ratio or 24-hour urine)<br>**Purpose:** Assess severity of glomerular involvement and explain edema.<br>**Expected findings:** Significant proteinuria may reflect **DKD**, but sudden marked increase could suggest **superimposed GN**.<br><br>### 3) Serum chemistry: BMP/CMP, albumin, CBC, eosinophils<br>**Purpose:** Define AKI severity, nephrotic contribution to edema, and clues to AIN/inflammation.<br>**Expected findings:**<br>- Elevated creatinine/BUN<br>- Low albumin if heavy protein loss<br>- Possible eosinophilia in **AIN** (not sensitive)<br><br>### 4) Urine culture and repeat infection evaluation<br>**Purpose:** Confirm or exclude persistent UTI/pyelonephritis.<br>**Expected findings:** May be negative if initial diagnosis was not true infection or if antibiotics already sterilized urine.<br><br>### 5) Renal ultrasound<br>**Purpose:** Exclude **postrenal obstruction**, assess kidney size and chronicity.<br>**Expected findings:** Usually no obstruction; kidneys may be normal/enlarged in diabetes.<br><br>### 6) Serologic workup for glomerulonephritis<br>- **C3/C4**<br>- **ANA**<br>- **ANCA**<br>- **anti-GBM antibody**<br>- **HBV/HCV as indicated**<br>**Purpose:** Evaluate for **RPGN/infection-related or immune-mediated GN**.<br>**Expected findings:** Usually normal in AIN/DKD; abnormalities would support specific GN.<br><br>### 7) Consider urinary eosinophils<br>**Purpose:** Supportive, though not definitive, evidence for AIN.<br>**Expected findings:** May be positive in AIN, but limited diagnostic value.<br><br>### 8) Kidney biopsy<br>**Purpose:** Definitive diagnosis if urinalysis/serologies do not clearly explain the rapid AKI, especially given known diabetes but atypically rapid decline.<br>**Expected findings:**<br>- **AIN**: interstitial inflammatory infiltrates and tubulitis<br>- **DKD**: diabetic glomerulosclerosis |

| | |
|---|---|
| | - **GN**: crescentic or immune-complex lesions depending on cause<br><br>## Bottom line<br>This patient most likely has **underlying diabetic kidney disease with a superimposed acute intrinsic renal injury**, and **drug-induced AIN after ceftriaxone** is the leading diagnosis. However, because the renal decline was **rapid**, **RPGN or another glomerulonephritis must be urgently excluded**, and **urine microscopy, renal ultrasound, serologies, and likely kidney biopsy** are key to narrowing the diagnosis. |
| With Static RAG and Mult-agent framework (Model C) | ## 4) Logical testing plan<br><br>### Immediate first-line tests<br>1. **Urinalysis with urine sediment microscopy**<br>- Highest-yield early test.<br>- Looks for:<br>- **Dysmorphic RBCs/RBC casts** → glomerulonephritis<br>- **WBCs/WBC casts** → interstitial nephritis or pyelonephritis<br>- **Granular casts** → tubular injury<br>- **Bacteriuria** → infection<br><br>2. **Quantification of proteinuria**<br>- Spot **protein-creatinine** or **albumin-creatinine ratio**.<br>- Helps determine whether nephrotic-range proteinuria or a major glomerular process is present.<br><br>3. **Basic blood tests**<br>- **BMP/CMP, BUN/creatinine, potassium, bicarbonate, albumin**<br>- **CBC**<br>- **CRP/ESR**<br>- These assess AKI severity, electrolyte complications, inflammation, anemia, and hypoalbuminemia.<br><br>4. **Renal ultrasound**<br>- Early, noninvasive, and cost-effective.<br>- Essential to exclude **hydronephrosis/obstruction** and assess kidney size.<br><br>5. **Urine culture** and **blood cultures if febrile**<br>- To confirm persistent urinary infection or bacteremia.<br><br>### Second-line tests if intrinsic renal disease is suspected<br>6. **Serologic evaluation for glomerulonephritis**<br>- **C3/C4, ANA, ANCA, anti-GBM**, and infection-related studies as clinically indicated.<br>- Useful if urine sediment suggests glomerular disease or AKI remains unexplained.<br><br>### Definitive test if diagnosis remains unclear or sediment is active<br>7. **Kidney biopsy**<br>- Indicated if there is **unexplained intrinsic AKI**, especially with active urine sediment or concern for glomerulonephritis/AIN.<br>- Can distinguish:<br>- diabetic kidney disease with superimposed lesion<br>- crescentic GN<br>- infection-related GN |

| | |
|---|---|
| | - AIN<br>- acute tubular injury<br><br>### Lower-priority tests<br>- **Urine eosinophils / fractional excretion indices**<br>- Limited diagnostic value and should not be relied upon.<br><br>---<br><br>## Changes from the initial version<br><br>1. **Problem representation revised**<br>- I more clearly separated **baseline likely diabetic kidney disease** from the **new superimposed AKI**.<br>- I avoided assuming the prior episode was definitely a **UTI**, since **pyuria alone does not prove infection**.<br><br>2. **Differential diagnosis re-ranked more cautiously**<br>- I no longer present **AIN as the most likely diagnosis**.<br>- I now prioritize **superimposed glomerular disease** and explicitly include **acute tubular injury** and **underlying diabetic kidney disease with an acute insult**.<br><br>3. **History and exam refined**<br>- I added emphasis on **urine output**, **gross hematuria**, **recent blood pressure change**, **pulmonary edema/heart failure**, and **signs of vasculitis**.<br><br>4. **Testing plan prioritized**<br>- I now emphasize **urinalysis with sediment**, **protein quantification**, **basic labs**, **urine culture**, and **renal ultrasound** as the immediate first steps.<br>- I reduced emphasis on **eosinophiluria** and **fractional excretion indices**, since they are less useful diagnostically. |
| **Phase II – Q5: Interpreting test results** | |
| Baseline LLM (Model A) | **5. Interpretation of the test results**<br><br>- **Nephrotic syndrome is clearly present**: albumin **1.9 g/dL**, marked edema, and urinary protein **7.33 g/day** (spot protein/Cr 12.8 g/gCr) indicate heavy glomerular protein loss.<br>- **There is also an active nephritic process**: creatinine **3.35 mg/dL**, hematuria (**11–30 RBC/HPF**), hypertension, and low **C3 (35 mg/dL)** with low CH50 suggest acute inflammatory glomerular injury rather than diabetic kidney disease alone.<br>- **The infection link is strong**: **ASO 1424 IU/mL** and elevated **CRP 6.3 mg/dL** support a recent streptococcal infection or streptococcal-related immune response. Blood cultures being negative does not exclude a preceding infection-related GN.<br>- **Biopsy findings are highly characteristic of infection-related/endocapillary proliferative GN**:<br>- **LM**: diffuse **endocapillary hypercellularity**, many **neutrophils**, lobulation, and **subepithelial humps**<br>- **IF**: **C3-dominant** granular staining with only mild IgG<br>- **EM**: **subepithelial humps** and intramembranous/subendothelial electron- |

| | |
|---|---|
|  | dense deposits<br> These are classic for **postinfectious/infection-related glomerulonephritis**, especially streptococcal-associated GN.<br>- **NAPlr/plasmin staining positivity** supports a **streptococcal infection-related mechanism** and is an important clue favoring poststreptococcal/infection-related GN.<br>- **Diabetic kidney disease is also present in the background**:<br> - prior diabetic retinopathy and neuropathy<br> - previous proteinuria<br> - biopsy showing **GBM thickening (650–970 nm)** and **polar vasculosis**<br> These findings support **coexistent diabetic nephropathy**, but the acute presentation is better explained by superimposed infection-related GN.<br>- Mild chronic damage (**5/25 globally sclerosed glomeruli, <15% IFTA**) suggests there is still meaningful reversible kidney injury. |
| With Static RAG (Model B) | ### 5. Interpretation of the test results<br>- **Nephrotic syndrome is clearly present**: albumin **1.9 g/dL**, marked leg edema, and **heavy proteinuria** (24-hour urine protein **7.33 g**, spot urine protein **12.8 g/gCr**). This degree of protein loss explains the edema and suggests major glomerular injury rather than isolated tubulointerstitial disease.<br>- **AKI is real and clinically significant**: creatinine **3.35 mg/dL** (eGFR **16.9 mL/min**) with BUN **42 mg/dL**. The rapid rise from prior baseline is not typical for uncomplicated diabetic kidney disease alone.<br>- **Urine sediment is nephritic rather than purely diabetic**: **11–30 RBC/HPF** with very few WBCs on admission argues against active pyelonephritis/AIN as the main cause and supports glomerular inflammation.<br>- **Low complement, especially low C3 (35 mg/dL), with elevated inflammatory markers** strongly supports an **infection-related immune-complex glomerulonephritis**, particularly a post-streptococcal pattern. C4 is not low, which fits alternative complement pathway activation often seen in post-infectious GN.<br>- **Markedly elevated ASO (1424 IU/mL)** is important evidence of **recent streptococcal infection**, even though blood cultures were negative. Negative cultures do not exclude a post-infectious process because the infection may already have resolved when nephritis appears.<br>- **Biopsy findings are highly characteristic of acute post-infectious / post-streptococcal GN**:<br> - **Diffuse endocapillary hypercellularity with many neutrophils**<br> - **Lobulated glomeruli**<br> - **Subepithelial humps** on LM/EM<br> - **C3-dominant granular deposits** on IF, with only mild IgG<br> These are classic features of infection-related GN.<br>- **NAPlr positivity** further supports **streptococcal infection-related glomerulonephritis**, because NAPlr is a recognized marker associated with post-streptococcal/infection-related GN.<br>- **GBM thickening (650–970 nm), enlarged glomeruli, and polar vasculosis** indicate **underlying diabetic nephropathy** as a background lesion. Thus, the patient likely has pre-existing diabetic kidney disease with a superimposed acute infection-related GN.<br>- The combination of **severe proteinuria/nephrotic syndrome** and **post-infectious GN pathology** is notable because post-streptococcal GN usually presents with nephritic syndrome, but in adults—especially with underlying diabetic glomerular changes—proteinuria can be much heavier.<br>- The later improvement in **creatinine, CRP, ASO, and complement** in the table is consistent with a **resolving post-infectious process**, which supports the |

| | |
|---|---|
| | interpretation above. |
| With Static RAG and Mult-agent framework (Model C) | ## 5. Interpretation of the test results<br><br>- **Nephrotic syndrome with superimposed AKI:** The patient has marked edema, **albumin 1.9 g/dL**, and **heavy proteinuria** (**24-hour urine protein 7.33 g/day**, urine protein **12.8 g/gCr**), which establish nephrotic syndrome. The simultaneous **creatinine 3.35 mg/dL** and **eGFR 16.9 mL/min** indicate a superimposed acute intrinsic renal process rather than stable diabetic kidney disease alone.<br><br>- **Active glomerular inflammation rather than tubulointerstitial disease:** **Hematuria (11–30 RBC/HPF)** with **<1 WBC/HPF** at admission supports a glomerular process. The earlier pyuria (>100 WBC/HPF) likely reflects a different time point during the preceding febrile illness and does not, by itself, establish AIN or persistent pyelonephritis.<br><br>- **Serology supports infection-related immune complex GN, likely streptococcal:** **Low C3 (35 mg/dL)** and **low CH50 (37 U/mL)** with relatively preserved/high **C4 (34 mg/dL)** fit complement consumption typical of infection-related GN, often through predominant alternative pathway activation. The markedly elevated **ASO 1424 IU/mL** strongly supports a recent streptococcal infection.<br><br>- **Biopsy is highly characteristic of infection-related glomerulonephritis:**<br> - **Light microscopy:** diffuse **endocapillary hypercellularity**, many **neutrophils**, mesangial expansion, swollen endothelial cells, and lobulated glomeruli are classic for exudative infection-related GN.<br> - **Immunofluorescence:** **C3-dominant** granular deposits with only weak **IgG** are typical of infection-related GN.<br> - **Electron microscopy:** **subepithelial humps** and intramembranous electron-dense deposits strongly support this diagnosis.<br><br>- **NAPlr/plasmin-related staining further supports bacterial infection-related GN:** Positivity for **NAPlr** and plasmin activity is an important pathologic clue favoring **streptococcal/bacterial infection-related glomerulonephritis**.<br><br>- **Underlying diabetic kidney disease is present as background pathology:** **GBM thickening (650–970 nm)** and **polar vasculosis** are consistent with diabetic glomerular change. These background diabetic changes may also contribute to the marked proteinuria, but they do not explain the acute nephritic/inflammatory lesion seen on biopsy.<br><br>- **Inflammatory state is present:** **CRP 6.3 mg/dL** supports recent systemic inflammation/infection. Negative blood cultures do not exclude a recently treated or already resolved infection.<br><br>- **Severe anemia is likely multifactorial:** **Hemoglobin 7.2 g/dL** may reflect inflammation, kidney dysfunction, and hemodilution from fluid retention. |
| **Phase II – Q6: Reconsidering the differential diagnosis** | |
| Baseline LLM (Model A) | **6. Updated diagnostic thinking / Working diagnosis**<br><br>The differential now shifts strongly away from isolated diabetic nephropathy or drug-induced interstitial nephritis. |

| | |
|---|---|
| | **Most likely working diagnosis:**<br>**Poststreptococcal acute glomerulonephritis (PSAGN) / streptococcal infection-related glomerulonephritis causing nephrotic syndrome and AKI, superimposed on underlying diabetic kidney disease.**<br><br>**Why this is most likely**<br>- Recent febrile illness<br>- Markedly elevated **ASO**<br>- **Low C3**<br>- Nephritic urine sediment with heavy proteinuria<br>- Biopsy showing **diffuse endocapillary proliferative exudative GN with subepithelial humps and C3-dominant deposits**<br>- **NAPlr positivity**, which strongly supports streptococcal infection-related GN<br><br>**Lower-priority alternatives**<br>- Other forms of infection-related GN<br>- C3 glomerulopathy or MPGN pattern disease<br>These are less likely because the clinical context and biopsy pattern are much more typical of poststreptococcal/infection-related GN. |
| With Static RAG (Model B) | ### 6. Updated diagnostic thinking / Working diagnosis<br>The differential has shifted substantially away from AIN or isolated diabetic kidney disease.<br><br>#### Most likely diagnosis<br>**Acute post-streptococcal glomerulonephritis (PSGN) / streptococcal infection-related glomerulonephritis, superimposed on underlying diabetic nephropathy.**<br><br>#### Why this is now the leading diagnosis<br>- Recent febrile illness<br>- **Very high ASO**<br>- **Low C3**<br>- **Hematuria with heavy proteinuria**<br>- **Diffuse endocapillary proliferative/exudative GN with neutrophils**<br>- **Subepithelial humps**<br>- **C3-dominant IF**<br>- **NAPlr positivity**<br><br>These findings together are much more specific for **streptococcal infection-related GN** than for other causes.<br><br>#### Revised prioritization of differential diagnosis<br>1. **Post-streptococcal / streptococcal infection-related GN** — overwhelmingly most likely<br>2. **Other infection-related GN** — less likely given strong streptococcal serology/NAPlr support<br>3. **Underlying diabetic nephropathy** — present as a chronic background condition, but not sufficient to explain the acute syndrome by itself<br><br>AIN is now unlikely as the main diagnosis because the biopsy and urine findings do not support it. |
| With | ## 6. Updated diagnostic thinking / Working diagnosis |

| | |
|---|---|
| Static RAG and Mult-agent framework (Model C) | The new data shift the differential strongly toward **infection-related glomerulonephritis**, most likely **streptococcal infection-related GN / APSGN-spectrum disease**, superimposed on **underlying diabetic kidney disease**.<br><br>### Working diagnosis<br>**Streptococcal infection-related glomerulonephritis (APSGN-spectrum disease), superimposed on diabetic kidney disease.**<br><br>### Why this is now the most probable diagnosis<br>- Recent **febrile illness**<br>- Markedly elevated **ASO**<br>- **Low C3/low CH50** with relatively preserved C4<br>- **Hematuria + nephrotic-range proteinuria + AKI**<br>- Biopsy showing **diffuse endocapillary proliferative/exudative GN with neutrophils**, **C3-dominant deposits**, and **subepithelial humps**<br>- **NAPlr positivity**, which supports bacterial, especially streptococcal, infection-related GN<br><br>### Revised differential priority<br>1. **Streptococcal infection-related GN / APSGN-spectrum disease** — most likely<br>2. **Other bacterial infection-related GN** — still important in an adult, though less favored by the high ASO and NAPlr findings<br>3. **Diabetic kidney disease with superimposed GN** — present as background disease, not sufficient alone to explain the presentation<br>4. **AIN or pyelonephritis** — now much less likely given the admission urine sediment and the biopsy findings |
| Phase II – Q7: Treatment planning | |
| Baseline LLM (Model A) | **7. Treatment plan**<br><br>### Main treatment approach<br>**Supportive management plus eradication/control of infection if any focus remains; avoid routine immunosuppression initially.**<br><br>#### a) Volume and blood pressure control<br>- **Salt and fluid restriction**<br>- **Loop diuretic** for edema<br>- Careful BP control, as BP is **178/84 mm Hg**<br>- Consider **RAS blockade later**, once kidney function is stable enough and acute phase improves, because it may reduce residual proteinuria, but it should be used cautiously during active AKI.<br><br>**Rationale:** In PSAGN/infection-related GN, supportive care is the cornerstone; edema and hypertension are major immediate problems.<br><br>#### b) Search for and treat any residual streptococcal or other infection<br>- Evaluate for ongoing infection focus and give appropriate **antibiotic therapy** if present; if streptococcal infection is suspected or not fully treated, penicillin-class therapy is commonly used unless contraindicated.<br><br>**Rationale:** Antibiotics do not reverse established immune injury rapidly, but they help eliminate persistent antigen exposure and prevent recurrence/transmission. |

| | |
|---|---|
|  | #### c) Monitor kidney complications closely<br>- Serial **creatinine/eGFR**, electrolytes, urine protein, complement, CRP, and ASO trend<br>- Monitor for indications for **dialysis**: refractory fluid overload, hyperkalemia, severe acidosis, or uremic complications<br><br>**Rationale:** AKI from PSAGN is often self-limited but may require temporary renal replacement support.<br><br>#### d) Avoid nephrotoxins<br>- Avoid NSAIDs, unnecessary antibiotics, radiocontrast, and other nephrotoxic exposures.<br><br>### Role of immunosuppression<br>- **Steroids are not first-line** for typical infection-related GN/PSAGN.<br>- They might be considered only in selected severe situations (e.g., crescentic rapidly progressive disease or persistent severe inflammation after infection is controlled), but such findings are **not provided here**.<br><br>**Rationale:** Evidence for routine steroid use in PSAGN is limited, and immunosuppression may be harmful if infection is still active.<br><br>### Additional long-term management<br>- After recovery phase, optimize **CKD and diabetic kidney disease care**:<br> - BP reduction<br> - proteinuria-lowering strategy<br> - ongoing diabetic complication management<br> - cardiovascular risk reduction<br><br>**Summary treatment plan:**<br>This patient should be managed primarily as **streptococcal infection-related GN with AKI and nephrotic syndrome**, using **supportive care (diuretics, salt restriction, BP control), assessment/treatment of residual infection, close renal monitoring, and dialysis if needed**, while recognizing coexisting **diabetic kidney disease** in the background. |
| With Static RAG (Model B) | ### 7. Treatment plan<br>#### Main treatment approach<br>**Supportive care for infection-related GN, plus management of nephrotic syndrome/AKI and confirmation that infection has been eradicated.**<br><br>#### Core components<br>1. **Treat or ensure eradication of streptococcal infection**<br> - Use an appropriate anti-streptococcal antibiotic if active/recent infection has not been definitively treated.<br> - Rationale: antibiotics do not directly reverse GN once established, but they help eliminate residual infection and prevent ongoing antigen exposure/transmission.<br><br>2. **Volume and blood pressure control**<br> - **Salt restriction**<br> - **Loop diuretics** for edema<br> - Antihypertensive therapy as needed<br> - Rationale: the patient has marked edema and hypertension, both of which worsen kidney stress and symptoms. |

| | |
|---|---|
|  | 3. **Careful AKI management**<br> - Monitor creatinine, urine output, potassium, acid-base status<br> - Avoid nephrotoxins<br> - Adjust drug doses for renal function<br> - Rationale: most adult post-infectious GN is managed conservatively unless severe complications occur.<br><br>4. **Proteinuria management**<br> - Consider **RAS blockade** only after renal function and volume status stabilize sufficiently<br> - Rationale: may reduce proteinuria long term, but initiation during unstable AKI requires caution.<br><br>5. **Monitor complement, ASO, CRP, urinary findings, and renal function over time**<br> - Rationale: normalization supports resolution; persistent abnormalities would prompt reassessment.<br><br>#### Role of immunosuppression<br>- **Routine corticosteroids are not first-line** for typical PSGN/infection-related GN.<br>- They may be considered only in selected severe situations (for example, crescentic disease or progressive renal failure not improving), but that is not established by the provided biopsy description.<br>- Rationale: in infection-related GN, treating supportively and addressing infection is standard; immunosuppression may be harmful if infection is ongoing.<br><br>#### Renal replacement therapy<br>- **Dialysis** only if indicated for refractory fluid overload, severe hyperkalemia, uremic complications, or severe acidosis.<br><br>#### Other considerations<br>- Because biopsy suggests **coexisting diabetic nephropathy**, long-term care should also include:<br> - strict BP control<br> - ongoing diabetes surveillance even if HbA1c is currently normal<br> - nephrology follow-up for residual CKD/proteinuria<br><br>#### Alternative options<br>- If subsequent course were atypical (persistent hypocomplementemia, worsening renal function, or evidence of another infection), reevaluate for **other infection-related GN** or **C3 glomerulopathy**.<br>- If active infection were proven elsewhere, management should prioritize **source control and targeted antibiotics**.<br><br>**In summary:** the best current approach is **supportive management of post-streptococcal infection-related GN with aggressive edema/BP control, confirmation of infection eradication, and close renal follow-up**, while recognizing underlying diabetic nephropathy as a chronic coexisting disease. |
| With Static RAG and Mult- | ## 7. Treatment plan<br><br>### Main treatment approach: supportive care plus evaluation for any ongoing infection focus |

| | |
|---|---|
| agent framework (Model C) | 1. **Supportive renal management**<br>- **Sodium and fluid restriction**<br>- **Loop diuretics** for edema and volume overload<br>- Close monitoring of **weight, urine output, blood pressure, creatinine, electrolytes**<br>- This is the cornerstone of treatment for infection-related GN<br><br>2. **Blood pressure control**<br>- Necessary because **BP is 178/84 mm Hg**<br>- Antihypertensive therapy should be chosen according to volume status and renal function<br>- **RAAS blockade should be deferred initially** during active AKI and reconsidered only after renal stabilization<br><br>3. **Evaluate carefully for an active or incompletely treated infection**<br>- Antibiotics should **not be given automatically**<br>- If an ongoing infectious source is identified, treat it appropriately and pursue **source control** if needed<br>- Negative blood cultures do not exclude a prior infection, but they do reduce support for active bacteremia<br><br>4. **Monitor disease course**<br>- Follow **creatinine, urine protein, urine sediment, C3/CH50, albumin, and ASO**<br>- Recovery of complement levels and improvement in kidney function/proteinuria would support the diagnosis and response<br><br>5. **Manage complications**<br>- Evaluate and treat **severe anemia**<br>- Monitor for indications for **dialysis**, including refractory volume overload, hyperkalemia, severe acidosis, or uremic complications<br><br>### Why immunosuppression is not first-line<br>- In typical **infection-related GN**, treatment is primarily **supportive** and focused on controlling infection if still present<br>- **Steroids are not standard initial therapy** and may worsen occult infection<br>- Immunosuppression would generally be reserved only for unusual situations, such as severe crescentic/progressive disease after infection has been excluded or controlled<br><br>## Changes from the initial results<br><br>- I **reframed the diagnosis more cautiously** as **streptococcal infection-related GN / APSGN-spectrum disease**, rather than stating definitively that it is classic APSGN.<br>- I **retained other bacterial infection-related GN in the differential**, especially because this is an adult patient.<br>- I clarified that **low C3 with relatively preserved C4** is compatible with infection-related GN.<br>- I added that the **nephrotic-range proteinuria may be accentuated by underlying diabetic kidney disease** and GBM thickening, although the acute lesion is clearly infection-related GN.<br>- I softened the earlier interpretation of urinary WBCs by noting that the **pyuria discrepancy likely reflects different time points**, rather than using it as strong evidence for or against AIN.<br>- I revised the treatment plan to state that **antibiotics should be used only if there is |

| | |
|---|---|
| | evidence of active or incompletely treated infection**, and that **RAAS blockade should be deferred initially** during AKI. |
| **Phase III – Q8: Evaluation of treatment** | |
| Baseline LLM (Model A) | 8. **Monitoring effectiveness, safety, risks, and countermeasures**<br><br>**Effectiveness monitoring**<br>- **Clinical status:** body weight, leg edema, blood pressure, urine volume, symptoms of uremia<br> - Goal: reduction in fluid overload and stable BP.<br>- **Renal function:** serial serum creatinine, BUN, eGFR<br> - Goal: stabilization or improvement; detect relapse or progression.<br>- **Proteinuria/urinalysis:** spot urine protein/Cr or 24-hour protein, urine sediment (hematuria)<br> - Goal: falling proteinuria and less active sediment.<br>- **Inflammation/complement activity:** CRP, C3/C4/CH50<br> - In PSAGN, normalization is expected; persistent C3-dominant activity suggests ongoing complement-mediated disease such as C3GN.<br>- **If available in follow-up of C3GN:** trend of **anti-factor H antibody** level may help assess ongoing complement dysregulation, though treatment decisions are mainly clinical/pathologic.<br><br>**Safety monitoring / specific risks**<br>1. **Fluid overload and uncontrolled hypertension**<br> - Risk: pulmonary edema, heart failure, faster CKD progression<br> - Monitoring: daily weight, edema, BP, intake/output<br> - Countermeasures: strict salt restriction, fluid restriction, adjust diuretics and antihypertensives promptly<br><br>2. **Overdiuresis / prerenal worsening**<br> - Risk: excessive volume depletion, AKI, hypotension<br> - Monitoring: BP, creatinine, electrolytes, orthostatic symptoms<br> - Countermeasures: titrate diuretics carefully; reassess if creatinine rises after aggressive fluid removal<br><br>3. **Electrolyte abnormalities**<br> - Risk: hypo-/hyperkalemia, hyponatremia<br> - Monitoring: serial electrolytes<br> - Countermeasures: adjust diuretics, diet, and medications according to abnormalities<br><br>4. **Progressive glomerular disease despite apparent resolution of infection**<br> - Risk: missed transition from PSAGN to **C3GN**<br> - Monitoring: persistent or recurrent nephrotic proteinuria, persistent C3-dominant deposits, complement abnormalities, worsening pathology<br> - Countermeasures: early repeat evaluation, including **repeat kidney biopsy** when the course is atypical<br><br>5. **Infection**<br> - Risk: recurrent or persistent infection, especially if immunosuppression is considered later<br> - Monitoring: fever, CRP, cultures as indicated<br> - Countermeasures: exclude active infection before any immunosuppressive therapy; treat documented infection promptly |

| | |
|---|---|
| | 6. **Progression to ESRD**<br>- Monitoring: long-term trend in creatinine/eGFR, proteinuria, BP control, symptoms<br>- Countermeasures: CKD care optimization, nephrology follow-up, timely dialysis planning if progression continues |
| With Static RAG (Model B) | ## 8. Monitoring effectiveness, safety, risks, and countermeasures<br><br>### A. How to monitor treatment effectiveness<br>1. **Clinical status**<br>- **Body weight, edema, blood pressure, urine volume**<br>- These directly reflect control of nephrotic fluid retention and hypertension.<br>- In this case, improvement in weight, edema, proteinuria, and creatinine after salt/water restriction and diuretics showed initial treatment response.<br><br>2. **Kidney function**<br>- **Serum creatinine, eGFR, BUN**<br>- Monitor for recovery, stabilization, or renewed decline.<br><br>3. **Proteinuria and urine sediment**<br>- **Spot urine protein/creatinine or 24-hour urine protein**<br>- **Urinalysis for hematuria**<br>- Falling proteinuria/hematuria suggests resolving glomerular inflammation; recurrent or worsening nephrotic-range proteinuria suggests relapse, progression, or an alternative diagnosis such as C3GN.<br><br>4. **Inflammatory and complement markers**<br>- **CRP, C3, CH50 (and C4 as needed), ASO**<br>- In typical resolving PSAGN, inflammation subsides and complement normalizes.<br>- Persistent or recurrent glomerular disease despite normalized infection-related markers should prompt reconsideration of the diagnosis.<br><br>5. **If atypical course persists**<br>- Reassess for **complement-mediated disease**, including **C3 glomerulopathy-related testing** when available (as ultimately done here with **aFH antibody** and **C3NeF**).<br><br>### B. Safety monitoring and specific risks<br>1. **Overdiuresis / volume depletion**<br>- Risks: worsening AKI, hypotension, prerenal azotemia.<br>- Monitoring: daily weight, BP, orthostatic symptoms, creatinine, electrolytes.<br>- Countermeasure: adjust diuretic dose, liberalize fluid restriction if excessive, reassess volume status carefully.<br><br>2. **Electrolyte disturbances from diuretics**<br>- Risks: hypokalemia, hyponatremia, metabolic alkalosis.<br>- Monitoring: serial **Na, K, bicarbonate, creatinine**.<br>- Countermeasure: dose adjustment, electrolyte replacement, change diuretic strategy if needed.<br><br>3. **Uncontrolled hypertension / persistent fluid overload**<br>- Risks: heart failure, accelerated CKD progression.<br>- Monitoring: BP, weight trend, edema, dyspnea. |

| | |
|---|---|
| | - Countermeasure: intensify salt restriction, optimize diuretics and antihypertensives, consider hospitalization if severe.<br><br>4. **Progressive kidney failure**<br>- Risks: ESRD, uremic complications.<br>- Monitoring: creatinine/eGFR trend, potassium, acid-base status, symptoms of uremia.<br>- Countermeasure: urgent nephrology reassessment, repeat biopsy if needed, prepare for renal replacement therapy when indicated.<br><br>5. **Missed transition from resolving PSAGN to C3GN**<br>- Risk: delayed diagnosis and treatment revision.<br>- Monitoring: discordance between clinical course and expected PSAGN recovery, especially:<br>- persistent or recurrent heavy proteinuria<br>- ongoing C3-dominant deposits<br>- persistent active urinary abnormalities<br>- progressive sclerosis on repeat biopsy<br>- Countermeasure: early repeat evaluation, including **repeat biopsy** and **complement pathway investigation**.<br><br>6. **Adherence failure**<br>- Risk: recurrent fluid retention, uncontrolled BP, delayed recognition of progression.<br>- Monitoring: missed visits, abrupt weight gain, return of edema/proteinuria.<br>- Countermeasure: reinforce follow-up, provide clear self-monitoring instructions (weight, edema, BP), and arrange close outpatient follow-up. |
| With Static RAG and Mult-agent framework (Model C) | 8. **Specific measures for evaluating treatment effectiveness and potential side effects or complications, and the monitoring/management plan**<br><br>### A. Monitoring treatment effectiveness<br><br>- **Volume status and congestion**<br>- Follow **body weight, edema, blood pressure**, and ideally urine output.<br>- In this case, supportive therapy initially appeared effective because **body weight decreased from 85.6 kg to 71.7 kg**, with improvement in edema.<br>- However, improvement in weight/edema should be interpreted as **control of fluid overload**, not necessarily remission of the glomerular disease.<br><br>- **Kidney function**<br>- Serial **serum creatinine/eGFR, BUN, and electrolytes** should be monitored.<br>- Initial improvement of **creatinine from 3.35 to 2.01 mg/dL** suggested partial recovery, but persistent renal vulnerability remained.<br><br>- **Proteinuria and urine findings**<br>- Repeated **quantitative proteinuria** and urinalysis are essential.<br>- Proteinuria improved initially (**12.8 to 2.97 g/day**) but later worsened markedly (**13.99 g/day**), which is a key sign that the disease process was not simply resolving PSAGN.<br>- Persistent or recurrent **nephrotic-range proteinuria** after apparent clinical improvement should trigger prompt reassessment.<br><br>- **Markers of post-infectious activity**<br>- Follow **CRP, C3, C4, CH50, and ASO**. |

|  |  |
|---|---|
|  | - In this case, later **normalization of CRP, C3, CH50, and ASO** supported resolution of the acute post-streptococcal phase.<br>- Therefore, recurrent edema and heavy proteinuria after serologic normalization should not be attributed automatically to persistent PSAGN.<br><br>- **Monitoring for transition from PSAGN to C3 glomerulopathy**<br>- Because the initial diagnosis was **PSAGN with coexisting diabetic nephropathy**, follow-up should also have been explicitly directed toward detecting **persistent complement-mediated glomerular injury**.<br>- Warning signs include:<br>- persistent or recurrent **heavy proteinuria**<br>- failure of renal/urinary findings to normalize despite resolution of infection-related markers<br>- pathologic evolution on repeat biopsy<br>- In this case, the second biopsy showed:<br>- persistent **C3-dominant deposits**<br>- minimal **IgG**<br>- disappearance of **subepithelial humps**<br>- clearer **subendothelial and mesangial electron-dense deposits**<br>- **GBM duplication**<br>- negative **NAPlr/plasmin receptor** staining<br>- These findings shifted concern away from uncomplicated resolving PSAGN and toward **C3GN secondary to PSAGN**.<br><br>- **Role of diabetic nephropathy in monitoring**<br>- Background **diabetic nephropathy class 1/2a** may have contributed to baseline proteinuria, but it did **not adequately explain the later relapse**, biopsy evolution, or progression to ESRD.<br>- Thus, recurrent nephrotic-range proteinuria should not be explained by diabetic nephropathy alone when complement-mediated pathology is evolving.<br><br>### B. Monitoring treatment safety and complications<br><br>- **Diuretics**<br>- Risks: **volume depletion, prerenal azotemia, hyponatremia, hypokalemia**, hyperuricemia.<br>- Monitor: **daily weight, blood pressure, creatinine, sodium, potassium, uric acid**, symptoms of dizziness or dehydration.<br>- Management: adjust dose to avoid overdiuresis; replace electrolytes when necessary.<br><br>- **Antihypertensive therapy**<br>- Risks: **hypotension, decreased renal perfusion, electrolyte abnormalities** depending on agent.<br>- Monitor: **blood pressure, creatinine, potassium**.<br>- Management: titrate carefully and reassess if renal function worsens after intensification.<br><br>- **Strict salt and water restriction**<br>- Risks: **dehydration**, excessive thirst, poor adherence.<br>- Monitor: **weight trend, sodium, creatinine**, symptoms of orthostasis or thirst.<br>- Management: individualize restrictions and reinforce dietary education.<br><br>- **Missed follow-up / poor adherence** |

| | |
|---|---|
| | - This was a major safety issue in this case because the patient interrupted follow-up repeatedly.<br>- Monitor: attendance, home records of **weight and BP** if available.<br>- Management: emphasize the need for regular nephrology review and educate about warning signs such as **rapid weight gain, worsening edema, rising BP, and recurrent heavy proteinuria**.<br><br>- **Long-term progression**<br>- Monitor for **CKD progression**, including eGFR, proteinuria, BP, anemia, and CKD-related complications.<br>- Because this condition progressed to **ESRD**, early planning for renal replacement therapy is important when deterioration continues. |
| **Phase III – Q9: Formulating plans for worsening conditions** | |
| Baseline LLM (Model A) | 9. **If the condition does not improve or suddenly changes: reassessment and revised action plan**<br><br>**How to think**<br>- If renal function, edema, or proteinuria **do not improve as expected**, do not assume prolonged PSAGN alone.<br>- Reconsider:<br>1. **Poor adherence / inadequate volume or BP control**<br>2. **Persistent or new infection**<br>3. **Underlying complement-mediated disease**, especially **C3GN**<br>4. Less likely but important: another superimposed renal process or medication-related injury<br><br>**What to do**<br>1. **Reassess clinically and biochemically**<br>- Repeat weight, BP, edema assessment<br>- Repeat creatinine/eGFR, albumin, proteinuria, urine sediment<br>- Recheck CRP and complement levels<br><br>2. **Look for clues that the diagnosis should be revised**<br>- Persistent or recurrent **heavy proteinuria**<br>- Failure of renal recovery<br>- Persistent **C3-dominant** findings or atypical histologic evolution<br>- Resolution of infection markers but ongoing glomerular activity<br><br>3. **Repeat kidney biopsy when the course is atypical**<br>- This is especially important if nephrotic syndrome worsens again or renal function behaves inconsistently with resolving PSAGN.<br>- In this case, the second biopsy appropriately helped reveal evolution toward **C3GN**.<br><br>4. **Evaluate the alternative mechanism directly**<br>- If C3 glomerulopathy is suspected, assess complement pathway abnormalities such as **C3NeF** and **anti-factor H antibodies**.<br><br>5. **Revise treatment according to the updated diagnosis**<br>- For resolving PSAGN: continue supportive care<br>- If **C3GN** is identified or strongly suspected: shift management toward a **complement-mediated glomerulopathy framework**, with closer specialized follow-up and consideration of disease-specific therapy according to availability and |

| | |
|---|---|
| | accepted practice<br>- Continue CKD risk reduction and prepare for renal replacement therapy if progression continues<br><br>**Practical principle**<br>- In infection-related GN, early improvement with supportive care is reassuring, but **relapse, persistent nephrotic syndrome, or persistent C3-dominant pathology should trigger prompt re-evaluation for C3GN rather than simple observation alone.** |
| With Static RAG (Model B) | ## 9. If the condition does not improve as expected or changes suddenly: reassessment and action plan<br><br>### A. How to think<br>If the patient does not follow the expected recovery course for PSAGN, I would first ask:<br>1. **Is this still resolving PSAGN, or is there another glomerular disease?**<br>2. **Is there a superimposed problem** such as recurrent infection, medication/volume-related worsening, or progression of diabetic kidney disease?<br>3. **Are the pathology and laboratory trends now more consistent with C3 glomerulopathy?**<br><br>In this case, the key warning signs were:<br>- recurrent **massive proteinuria**<br>- persistent glomerular abnormalities despite normalization of **CRP, ASO, and complement**<br>- repeat biopsy showing disappearance of humps but **persistent C3 deposits**, more evident **subendothelial/mesangial EDDs**, and **GBM duplication**<br>These are atypical for simple resolved PSAGN and should shift thinking toward **C3GN**.<br><br>### B. What actions to take<br>1. **Rapid clinical reassessment**<br>- Check weight, edema, BP, urine output, and current kidney function.<br>- Rule out immediately reversible factors such as excess salt/water intake, medication nonadherence, nephrotoxins, or hemodynamic causes.<br><br>2. **Repeat laboratory evaluation**<br>- Creatinine/eGFR, albumin, urinalysis, urine protein quantification<br>- CRP, C3/C4/CH50, ASO<br>- This helps determine whether the process is inflammatory/infectious, resolving, or evolving into complement-mediated disease.<br><br>3. **Repeat kidney biopsy when the course is atypical**<br>- Appropriate when nephrotic syndrome worsens again or pathology may have changed.<br>- In this case, the second biopsy was the correct step because it revealed features raising suspicion for C3GN.<br><br>4. **Revise the differential diagnosis rather than anchoring on the original diagnosis**<br>- Move from “resolving PSAGN only” to “PSAGN-triggered or secondary **C3GN**,” especially when C3-dominant deposits persist and morphology evolves.<br><br>5. **Perform complement-pathway workup**<br>- Test for **aFH antibodies, C3NeF**, and related abnormalities when available. |

| | |
|---|---|
| | - In this case, persistent elevation of **aFH antibodies** supported the final diagnosis of **aFH antibody-positive C3GN secondary to PSAGN**.<br><br>6. **Revise treatment/follow-up intensity**<br>- Tighten follow-up frequency.<br>- Continue supportive nephrotic/CKD management.<br>- If C3GN is confirmed or strongly suspected, management should be reconsidered in that framework and long-term renal prognosis discussed.<br><br>### C. Practical principle<br>When a presumed PSAGN patient shows **recurrent nephrotic syndrome, persistent C3-dominant pathology, or progressive renal dysfunction despite improvement of infection-related markers**, do not assume simple slow recovery. Reassess promptly, repeat biopsy if needed, investigate the alternative complement pathway, and revise the diagnosis and management accordingly. |
| With Static RAG and Mult-agent framework (Model C) | 9. **Thought process and action plan for rapid reassessment and flexible revision of diagnosis/treatment**<br><br>### A. Reassessment when the course changes<br><br>If a patient with presumed **PSAGN with diabetic nephropathy** improves initially with supportive therapy but later develops **recurrent edema or nephrotic-range proteinuria**, I would immediately reconsider whether:<br>1. the original diagnosis was incomplete,<br>2. the post-infectious process has transitioned into **C3 glomerulopathy**,<br>3. the worsening is due only to **fluid/salt nonadherence**, or<br>4. another superimposed renal insult has appeared.<br><br>In this case, the key point is that **improvement in edema, body weight, and creatinine did not prove immunologic remission**. Supportive therapy may relieve congestion while **ongoing complement-mediated glomerular injury** continues.<br><br>### B. Immediate action plan<br><br>- Reassess:<br>- **body weight, edema, BP**<br>- **serum creatinine/eGFR, albumin, electrolytes**<br>- **quantitative proteinuria and urine sediment**<br>- **CRP, C3/C4, CH50, ASO**<br>- Review reversible contributors:<br>- nonadherence to **salt/fluid restriction**<br>- uncontrolled hypertension<br>- nephrotoxic medications<br>- recurrent infection<br><br>### C. Threshold for revising the diagnosis<br><br>Once the patient had:<br>- recurrent **nephrotic-range proteinuria**<br>- recurrent edema<br>- normalization of **CRP, C3, CH50, and ASO**<br>- and biopsy evolution away from classic acute PSAGN |

the working diagnosis should shift from “resolving PSAGN” to **suspected C3 glomerulopathy**, specifically **C3GN secondary to PSAGN**.

This threshold should be explicit: in **PSAGN with diabetic nephropathy**, persistent or recurrent heavy proteinuria after normalization of infection-related serologies warrants:
- **early repeat kidney biopsy**
- evaluation of the **alternative complement pathway**

#### D. Diagnostic revision based on the second biopsy

The second biopsy provided major clues favoring **C3GN**:
- persistent **C3-dominant deposits**
- only minimal **IgG**
- disappearance of classic **humps**
- more evident **subendothelial and mesangial EDDs**
- **GBM duplication**
- loss of **NAPlr/plasmin receptor** staining

These changes are not well explained by diabetic nephropathy and are more consistent with an evolving **C3 glomerulopathy** after the acute post-infectious phase.

#### E. Complement-focused evaluation

When C3 glomerulopathy is suspected, assessment of the **alternative complement pathway** is required, including:
- **C3 nephritic factor (C3NeF)**
- **anti-factor H (aFH) autoantibodies**

In this case:
- **C3NeF was normal**
- **aFH antibodies were persistently elevated** at the first biopsy, second biopsy, and later follow-up

These results support the final diagnosis of **aFH antibody-positive C3GN secondary to PSAGN**.

#### F. Treatment-plan revision

- Continue optimal **supportive therapy** for volume overload, BP control, and proteinuria surveillance.
- But once the disease pattern no longer fits resolving PSAGN, management should be revised around the possibility of **complement-mediated glomerular disease**, rather than assuming prolonged recovery.
- Diabetic nephropathy should be recognized as a **coexisting background lesion**, not the main explanation for relapse.
- Because the disease can progress to **ESRD**, ensure:
  - close nephrology follow-up
  - monitoring for CKD complications
  - timely dialysis planning if progression continues

#### Changes from the initial results

- I made the monitoring plan more explicit about the need to detect **transition from

| | |
|---|---|
| | PSAGN to C3 glomerulopathy**, not just track edema and creatinine.<br>- I clarified that improvement in **weight/edema** reflects **better volume control**, which may not equal true disease remission.<br>- I placed greater emphasis on **persistent/recurrent nephrotic-range proteinuria after normalization of CRP/complement/ASO** as a trigger for **early repeat biopsy** and **alternative complement pathway testing**.<br>- I revised the reassessment framework so that, after the second biopsy findings, **C3GN secondary to PSAGN** is prioritized over "resolving PSAGN."<br>- I also clarified that **diabetic nephropathy coexisted** but did not explain the biopsy shift, relapse, or progression to ESRD. |

| **Case 4: Early administration of caplacizumab combined with plasma exchange for thrombotic microangiopathy due to malignant hypertension: a case report** [12] | |
|---|---|
| **Phase I – Q1: Summarizing medical problems** | |
| Baseline LLM (Model A) | 1. **Problem representation**<br><br>A 51-year-old woman with a **subacute 3–4 week course** of **severe hypertension** (initially **225/150 mmHg**) developed **visual disturbance, headache, fatigue, nausea, oliguria**, and was found to have **KW grade III hypertensive retinopathy**, **renal dysfunction**, **anemia**, and **thrombocytopenia**. This is a **hypertensive emergency with target-organ damage** involving the **retina, kidneys, and likely microvasculature**, raising strong concern for **thrombotic microangiopathy (TMA) associated with malignant hypertension**. |
| With Static RAG (Model B) | ## 1. Problem representation<br>A 51-year-old woman with a subacute 25-day course of **hypertensive emergency/malignant hypertension** presents with **very severe BP elevation (225/150 mmHg)**, **grade III hypertensive retinopathy with papilledema/retinal hemorrhage**, progressive **headache, visual deterioration, nausea, fatigue, oliguria**, and is now found to have **acute kidney injury/renal dysfunction**, **anemia**, and **thrombocytopenia**.<br>The key clinical problem is **severe hypertension with end-organ damage** involving the **retina, kidney, and likely microvasculature**, raising concern for **malignant hypertension with thrombotic microangiopathy (TMA)**, while also requiring exclusion of other primary TMAs and secondary causes of severe hypertension. |
| With Static RAG and Mult-agent framework (Model C) | ## 1. Problem representation<br>A 51-year-old woman with a **subacute 25-day course** of **severe hypertension** (initial BP **225/150 mmHg**) presents with **visual decline, headache, general fatigue, nausea, oliguria, renal dysfunction, anemia, and thrombocytopenia**, with funduscopic evidence of **Keith–Wagener class III hypertensive retinopathy** (retinal hemorrhage and papilledema). This is most consistent with **hypertensive emergency/malignant hypertension with retinal and renal end-organ injury**, plus **bicytopenia suspicious for thrombotic microangiopathy (TMA)**; however, **microangiopathic hemolytic anemia still requires confirmation** by smear and hemolysis studies. |
| **Phase I – Q2: Differential diagnoses and rationale** | |
| Baseline LLM (Model A) | 2. **Differential diagnoses** |

| | |
|---|---|
| | **Top 3 differential diagnoses**<br>1. **Malignant hypertension / hypertensive emergency with secondary TMA**<br>- Most likely because she has markedly elevated BP, hypertensive retinopathy with papilledema/hemorrhage, renal dysfunction, and cytopenias compatible with mechanical hemolysis and platelet consumption from vascular injury.<br>2. **Primary TMA syndrome, especially thrombotic thrombocytopenic purpura (TTP)**<br>- Consider because of anemia + thrombocytopenia + neurologic-type symptoms (headache) + renal dysfunction. Severe hypertension can occur secondarily, though the retinal findings and very high BP favor malignant hypertension.<br>3. **Hemolytic uremic syndrome / complement-mediated TMA (atypical HUS)**<br>- Renal dysfunction with anemia and thrombocytopenia fits; severe hypertension may be a consequence or trigger. Important if renal injury is disproportionate or if findings are not fully explained by hypertension alone.<br><br>**Most likely diagnosis**<br>- **Malignant hypertension causing hypertensive emergency with renal TMA.**<br><br>**Must not be missed**<br>- **TTP**, because delayed plasma exchange can be fatal.<br>- Also important not to miss **secondary causes of severe hypertension** (e.g., scleroderma renal crisis, pheochromocytoma, renovascular disease), but based on the given data TTP is the most urgent alternate diagnosis. |
| With Static RAG (Model B) | ## 2. Differential diagnosis<br>### Top 3 differential diagnoses<br>1. **Malignant hypertension causing secondary TMA**<br>- Most likely.<br>- Rationale: Extreme hypertension, hypertensive retinopathy with papilledema, renal dysfunction, and cytopenias strongly fit malignant hypertension with endothelial injury and microangiopathic hemolysis.<br><br>2. **Thrombotic thrombocytopenic purpura (TTP) / primary TMA**<br>- Must be considered because anemia + thrombocytopenia + renal dysfunction can reflect TMA.<br>- Rationale: TTP can present with neurologic symptoms, renal injury, thrombocytopenia, and hemolytic anemia. Severe hypertension may be secondary rather than primary.<br><br>3. **Atypical hemolytic uremic syndrome (aHUS) / complement-mediated TMA**<br>- Important alternative.<br>- Rationale: Renal dysfunction with thrombocytopenia and anemia is compatible with complement-mediated TMA; hypertension may be severe in this setting.<br><br>### Most likely diagnosis<br>**Malignant hypertension with hypertensive emergency causing renal injury and secondary TMA.**<br>This is supported by the prominent hypertensive target-organ findings, especially **KW class III retinopathy**, severe BP elevation, and progressive kidney-related symptoms.<br><br>### Must not be missed<br>**TTP** must not be missed because delayed treatment can be fatal and may require urgent plasma exchange. |

| | |
|---|---|
| | Also important not to miss **secondary causes of severe hypertension** such as scleroderma renal crisis, pheochromocytoma, renal artery stenosis, or glomerulonephritis, depending on supporting findings. |
| With Static RAG and Mult-agent framework (Model C) | ## 2. Prioritized differential diagnosis<br>### 1. Malignant hypertension with secondary TMA and acute kidney injury<br>- Most likely.<br>- Strongly supported by **markedly elevated BP**, **advanced hypertensive retinopathy**, **headache/visual symptoms**, and **progressive renal dysfunction with oliguria**.<br>- Severe hypertension can itself cause endothelial injury leading to **anemia and thrombocytopenia**.<br><br>### 2. Thrombotic thrombocytopenic purpura (TTP)<br>- Must not be missed.<br>- Anemia + thrombocytopenia + neurologic-type symptoms + renal dysfunction can fit TTP.<br>- However, the **very severe hypertension with grade III hypertensive retinopathy** makes malignant hypertension more likely than primary TTP.<br><br>### 3. Complement-mediated TMA / atypical HUS<br>- Important alternative if hemolysis is confirmed and the course is not fully explained by hypertension alone.<br>- Can present with **AKI, thrombocytopenia, anemia, and severe hypertension**.<br><br>### 4. Intrinsic renal disease causing severe/secondary hypertension<br>- Examples: chronic glomerular disease, rapidly progressive glomerulonephritis, advanced CKD.<br>- Important because severe renal parenchymal disease may present with **HT, renal dysfunction, and urinary abnormalities**, and may either trigger or coexist with malignant hypertension.<br><br>### 5. Secondary hypertension, especially renovascular disease<br>- Should be considered in severe new hypertension with renal dysfunction.<br>- Other secondary causes are less directly suggested here but still possible depending on history/exam.<br><br>### Most likely<br>**Malignant hypertension with secondary renal injury and possible secondary TMA**<br><br>### Must not miss<br>**TTP**, because urgent plasma exchange may be lifesaving. |
| **Phase I – Q3: Necessary physical examinations and reasons** | |
| Baseline LLM (Model A) | 3. **Additional history and physical examination**<br><br>**Targeted history**<br>- **Neurologic symptoms**: confusion, focal deficits, seizures<br> - To assess severity of hypertensive emergency and screen for TTP/CNS involvement.<br>- **Bleeding/purpura**<br> - To characterize thrombocytopenia severity and support TMA.<br>- **Fever, diarrhea, recent infection**<br> - To evaluate TTP/HUS triggers. |

| | |
|---|---|
|  | - **Drug history**: quinine, calcineurin inhibitors, antiplatelet drugs, chemotherapy, oral contraceptives, illicit drugs<br> - To identify drug-induced TMA or causes of severe hypertension.<br>- **Autoimmune symptoms**: Raynaud phenomenon, skin thickening, arthralgia, rash<br> - To screen for scleroderma renal crisis or lupus-related TMA.<br>- **Symptoms of secondary hypertension**: episodic palpitations/sweating, muscle weakness, sleep apnea symptoms<br> - To assess pheochromocytoma, hyperaldosteronism, etc.<br>- **Urinary history**: hematuria, frothy urine, urine volume trend<br> - To define renal involvement.<br>- **Previous BP history** and any prior pregnancy-related hypertension<br> - To judge chronicity and predisposition.<br><br>**Physical examination**<br>- **Accurate BP in both arms**, repeated measurements<br> - To confirm severity and assess for vascular disease.<br>- **Detailed neurologic exam**<br> - To detect hypertensive encephalopathy or TTP-related CNS findings.<br>- **Funduscopic confirmation**<br> - To document malignant hypertensive changes.<br>- **Volume status**: edema, JVP, mucous membranes, lung crackles<br> - To guide antihypertensive and renal management.<br>- **Cardiovascular exam**: S4, murmurs, signs of heart failure<br> - To evaluate end-organ damage.<br>- **Skin exam** for purpura, livedo, sclerodactyly<br> - To support TMA, vasculopathy, or connective tissue disease. |
| With Static RAG (Model B) | ## 3. Additional history and physical examination<br>### Additional questions<br>- **Prior hypertension history**: any previous BP measurements, headaches, longstanding symptoms<br> - To distinguish chronic untreated HT from abrupt secondary HT.<br>- **Neurologic symptoms**: confusion, focal deficits, seizures<br> - To assess hypertensive encephalopathy or TTP-related CNS involvement.<br>- **Hemolysis/TMA symptoms**: dark urine, jaundice, bruising, bleeding<br> - To support microangiopathic hemolytic anemia.<br>- **Volume/renal symptoms**: edema, dyspnea, urine volume trend, gross hematuria, foamy urine<br> - To characterize AKI and possible intrinsic renal disease.<br>- **Autoimmune symptoms**: Raynaud phenomenon, skin thickening, rash, oral ulcers, arthralgia<br> - To assess for scleroderma renal crisis, lupus, vasculitis.<br>- **Drug/exposure history**: NSAIDs, oral contraceptives, cocaine/amphetamines, herbal agents<br> - To identify contributors to hypertensive crisis or TMA.<br>- **Endocrine/secondary HT symptoms**: episodic palpitations, sweating, weight loss, muscle weakness<br> - To screen for pheochromocytoma, hyperthyroidism, primary aldosteronism.<br>- **Infectious/diarrheal prodrome**<br> - To assess HUS.<br>- **Pregnancy possibility**<br> - Low likelihood at 51, but hypertensive/TMA syndromes related to pregnancy should be excluded if relevant. |

| | |
|---|---|
|  | ### Physical examination<br>- **Repeat BP in both arms, pulse, orthostatic status**<br> - To confirm severity and evaluate for vascular disease.<br>- **Full neurologic exam**<br> - To detect encephalopathy, stroke, or TTP-related deficits.<br>- **Funduscopic confirmation**<br> - To document malignant hypertensive retinopathy severity.<br>- **Cardiopulmonary exam**<br> - To assess heart failure, pulmonary edema, LV strain from severe HT.<br>- **Volume status exam**<br> - To guide BP lowering and renal management.<br>- **Skin exam**<br> - Purpura/ecchymosis suggest thrombocytopenia; skin thickening may suggest scleroderma.<br>- **Abdominal/vascular exam**<br> - Bruits may suggest renovascular hypertension. |
| With Static RAG and Mult-agent framework (Model C) | ## 3. Focused additional history and physical examination<br>### Additional history<br>1. **Previous blood pressure history**<br> - To determine whether this is longstanding unrecognized hypertension versus a new abrupt process.<br><br>2. **Baseline kidney-related history**<br> - Prior edema, foamy urine, hematuria, nocturia, or known kidney disease would support underlying renal parenchymal disease.<br><br>3. **Medication and substance exposure**<br> - NSAIDs, oral contraceptives, decongestants, stimulants, cocaine, herbal agents, and adherence to recent antihypertensives may contribute to severe HT or kidney injury.<br><br>4. **Urine output trend and volume-overload symptoms**<br> - Oliguria duration, dyspnea, orthopnea, and leg edema help assess AKI severity and complications.<br><br>5. **Neurologic symptoms**<br> - Confusion, focal deficits, seizure, or altered mental status would suggest hypertensive encephalopathy, PRES, stroke, or TTP-related CNS involvement.<br><br>6. **Bleeding/bruising or hemolysis symptoms**<br> - Petechiae, easy bruising, dark urine, jaundice can support thrombocytopenia or hemolysis.<br><br>7. **Secondary hypertension clues**<br> - Episodic palpitations/sweating, muscle weakness, sleep apnea symptoms, or flank/abdominal vascular symptoms.<br><br>### Focused physical examination<br>1. **Repeat BP, ideally in both arms**<br> - Confirms severity and helps assess for vascular disease.<br><br>2. **Detailed neurologic examination** |

| | |
|---|---|
| | - Looks for encephalopathy, stroke, or seizure-related findings.<br><br>3. **Funduscopic examination**<br>- Confirms severity of hypertensive retinal injury.<br><br>4. **Cardiopulmonary examination**<br>- Evaluates for heart failure, pulmonary edema, or chronic hypertensive heart disease.<br><br>5. **Volume assessment**<br>- JVP, edema, lung crackles, mucous membranes; important for BP and renal management.<br><br>6. **Skin examination**<br>- Petechiae or purpura may support thrombocytopenia/TMA.<br><br>7. **Abdominal/vascular examination**<br>- Renal artery bruits or other vascular findings may suggest renovascular hypertension. |
| **Phase I – Q4: Plan for investigations/tests** | |
| Baseline LLM (Model A) | 4. **Tests to narrow the diagnosis**<br><br>**Essential initial tests**<br>- **CBC with peripheral smear, reticulocyte count**<br>- Purpose: confirm anemia/thrombocytopenia and look for **schistocytes** suggesting microangiopathic hemolytic anemia.<br>- Expected: anemia, thrombocytopenia, schistocytes if TMA is present.<br>- **Hemolysis panel: LDH, haptoglobin, indirect bilirubin**<br>- Purpose: determine whether anemia is hemolytic.<br>- Expected: high LDH, low haptoglobin, elevated indirect bilirubin in TMA.<br>- **Basic metabolic panel, creatinine, BUN, electrolytes**<br>- Purpose: assess severity of kidney injury and complications.<br>- Expected: renal dysfunction, possibly hyperkalemia or metabolic acidosis.<br>- **Urinalysis with sediment and urine protein quantification**<br>- Purpose: characterize renal injury (proteinuria, hematuria, casts).<br>- Expected: proteinuria/hematuria are common in malignant hypertension.<br>- **Coagulation tests (PT, aPTT, fibrinogen, D-dimer)**<br>- Purpose: distinguish TMA from DIC.<br>- Expected: usually relatively normal in TMA; abnormal values would suggest DIC.<br>- **Direct Coombs test**<br>- Purpose: exclude immune hemolysis.<br>- Expected: negative in TMA.<br>- **ADAMTS13 activity/inhibitor**<br>- Purpose: evaluate for TTP.<br>- Expected: severe deficiency would support TTP; normal/moderately reduced would favor hypertension-associated TMA.<br>- **Pregnancy test only if relevant biologically**<br>- Usually low likelihood here, but pregnancy-associated TMA should be excluded if appropriate.<br><br>**Assessment of hypertensive end-organ damage**<br>- **ECG and echocardiography**<br>- Purpose: detect LVH, ischemia, heart failure. |

| | |
|---|---|
| | - Expected: chronic pressure overload findings may be present.<br>- **Brain imaging if neurologic symptoms/signs are significant**<br>- Purpose: rule out intracranial hemorrhage, PRES, stroke.<br>- **Chest X-ray**<br>- Purpose: assess cardiomegaly/pulmonary edema.<br><br>**Tests for secondary causes / alternative TMA etiologies**<br>- **ANA, complements, anti-dsDNA, antiphospholipid antibodies**<br>- Purpose: evaluate autoimmune causes.<br>- **Renin/aldosterone, catecholamine testing, renal artery imaging**<br>- Purpose: investigate secondary hypertension if clinically suspected.<br>- **Stool Shiga toxin testing** if diarrheal prodrome exists<br>- Purpose: assess typical HUS.<br><br>**Kidney biopsy**<br>- Consider **after stabilization** if diagnosis remains uncertain.<br>- Purpose: distinguish malignant nephrosclerosis/TMA from primary glomerular disease or other renal pathology.<br>- Expected in malignant hypertension: arteriolar fibrinoid necrosis, onion-skin intimal thickening, ischemic glomerular changes, TMA features.<br><br>**Overall expected conclusion from testing**<br>- The most likely pattern would be **microangiopathic hemolytic anemia with thrombocytopenia and acute kidney injury in the setting of malignant hypertension**, with **ADAMTS13 not severely deficient**, supporting **hypertension-induced TMA** over TTP. |
| With Static RAG (Model B) | ## 4. Tests to narrow the diagnosis<br>### Essential initial tests<br>1. **CBC with smear, reticulocyte count**<br>- Purpose: characterize anemia/thrombocytopenia; look for schistocytes.<br>- Expected: anemia, thrombocytopenia, schistocytes if TMA is present.<br><br>2. **Hemolysis panel: LDH, haptoglobin, indirect bilirubin**<br>- Purpose: confirm microangiopathic hemolytic anemia.<br>- Expected: high LDH, low haptoglobin, elevated indirect bilirubin.<br><br>3. **Basic metabolic panel, creatinine, BUN, electrolytes**<br>- Purpose: define severity of kidney injury and complications.<br>- Expected: renal dysfunction; possible hyperkalemia or metabolic acidosis.<br><br>4. **Urinalysis with microscopy and urine protein quantification**<br>- Purpose: evaluate intrinsic kidney injury, proteinuria, hematuria, casts.<br>- Expected: proteinuria and hematuria may be present in malignant hypertension; heavy active sediment would raise concern for glomerulonephritis.<br><br>5. **Coagulation profile (PT, aPTT, fibrinogen, D-dimer)**<br>- Purpose: distinguish TMA from DIC.<br>- Expected: often relatively normal in TMA.<br><br>### Key tests for TMA subtype<br>6. **ADAMTS13 activity/inhibitor**<br>- Purpose: evaluate for TTP.<br>- Expected: severe deficiency would support TTP; normal/moderately reduced |

| | |
|---|---|
| | favors secondary TMA.<br><br>7. **Complement studies (C3, C4, possibly soluble C5b-9/genetic workup if needed)**<br>- Purpose: assess complement-mediated TMA/aHUS.<br>- Expected: may be low or normal; not diagnostic alone.<br><br>8. **Direct Coombs test**<br>- Purpose: exclude autoimmune hemolysis.<br>- Expected: negative in MAHA/TMA.<br><br>### Evaluation of hypertensive emergency and secondary causes<br>9. **ECG, chest X-ray, echocardiography**<br>- Purpose: detect cardiac end-organ damage.<br>- Expected: LVH and/or strain; possible heart failure findings.<br><br>10. **Renal ultrasound**<br>- Purpose: assess kidney size, chronicity, obstruction.<br>- Expected: no obstruction; kidney size may help judge chronicity.<br><br>11. **Renal artery imaging if indicated (Doppler/CTA/MRA)**<br>- Purpose: evaluate renovascular hypertension.<br>- Expected: usually normal unless renovascular disease is present.<br><br>12. **Autoimmune serologies if suggested clinically (ANA, anti-dsDNA, ANCA, anti-Scl-70/RNA polymerase III)**<br>- Purpose: assess lupus, vasculitis, scleroderma renal crisis.<br>- Expected: guided by clinical suspicion rather than routine positivity.<br><br>### Possible definitive test<br>13. **Kidney biopsy** if diagnosis remains unclear and platelet count/BP allow<br>- Purpose: distinguish malignant nephrosclerosis from primary TMA or glomerulonephritis.<br>- Expected in malignant hypertension: arteriolar fibrinoid necrosis, onion-skin intimal hyperplasia, ischemic glomerular changes.<br><br>## Bottom line<br>The leading diagnosis is **malignant hypertension with hypertensive emergency causing renal injury and secondary TMA**. The immediate priority is to confirm **MAHA/TMA**, assess the extent of end-organ damage, and urgently exclude **TTP**, the key life-threatening alternative. |
| With Static RAG and Mult-agent framework (Model C) | ## 4. Logical testing plan<br>### Immediate tests<br>1. **CBC with peripheral smear and reticulocyte count**<br>- Confirms severity of anemia/thrombocytopenia and looks for **schistocytes**, which would support TMA.<br><br>2. **Hemolysis studies: LDH, haptoglobin, indirect bilirubin**<br>- Determines whether **microangiopathic hemolytic anemia** is actually present.<br><br>3. **Basic metabolic panel, BUN, creatinine, electrolytes**<br>- Assesses severity of AKI and urgent complications. |

| | |
|---|---|
| | 4. **Urinalysis with urine sediment and protein quantification**<br>- Helps distinguish hypertensive renal injury from intrinsic glomerular disease.<br><br>5. **Coagulation studies: PT/INR, aPTT, fibrinogen, D-dimer**<br>- Helps separate TMA from DIC.<br><br>6. **ADAMTS13 activity/inhibitor**<br>- Essential to evaluate for **TTP**.<br><br>### Early end-organ and structural assessment<br>7. **Renal ultrasound**<br>- Low-risk test to assess kidney size, chronicity, and exclude obstruction.<br><br>8. **ECG and echocardiography**<br>- Evaluates cardiac target-organ damage such as LVH or heart failure.<br><br>9. **Brain imaging if neurologic findings are significant**<br>- To assess for hemorrhage, infarction, or PRES.<br><br>### Deferred/targeted tests after stabilization<br>10. **Complement studies (C3/C4 and further complement workup if indicated)**<br>- Consider if TMA is confirmed and the course is atypical for hypertension alone.<br><br>11. **Secondary hypertension evaluation guided by suspicion**<br>- Such as renin/aldosterone testing or renal artery imaging, especially if clinical clues emerge.<br><br>12. **Kidney biopsy**<br>- **Selective**, not routine.<br>- Consider only after stabilization and if thrombocytopenia/bleeding risk permit, especially if uncertainty remains between malignant nephrosclerosis, primary renal disease, or complement-mediated TMA.<br><br>## Changes from the initial version<br>- **Problem representation refined** to emphasize the **subacute course** and to avoid assuming proven microangiopathic involvement before hemolysis is confirmed.<br>- **Differential broadened** to include **intrinsic renal disease** and **secondary hypertension (especially renovascular disease)**, while keeping malignant hypertension first and TTP as the key dangerous alternative.<br>- **History/exam reprioritized** toward **baseline hypertension history, medication/substance exposure, urine output, dyspnea/edema, neurologic exam, and volume/cardiac assessment**; lower-yield items were de-emphasized.<br>- **Testing plan reorganized** into **immediate**, **early**, and **deferred** steps, with emphasis on urgent confirmation of hemolysis/TMA and exclusion of TTP.<br>- **Kidney biopsy reframed** as a **selective later test**, not an early routine step.<br><br>## Bottom line<br>The leading diagnosis is **malignant hypertension causing retinal injury, AKI, and possible secondary TMA**, but **TTP must be urgently excluded**, and **underlying renal disease or secondary hypertension** should be considered if the presentation is not fully explained after initial evaluation. |
| **Phase II – Q5: Interpreting test results** | |
| Baseline | **5. Interpretation of the test results** |

| | |
|---|---|
| LLM (Model A) | - **Severe kidney failure dominates the presentation:** BUN **290.6 mg/dL** and creatinine **24.4 mg/dL** indicate profound renal failure, consistent with her oliguria and uremic symptoms. **Normal-sized kidneys** suggest an **acute or subacute process**, not long-standing end-stage chronic kidney disease.<br>- **Urinalysis supports intrinsic renal microvascular/glomerular injury:** **Proteinuria 1.8 g/day** and **10–19 RBC/HPF** indicate renal parenchymal injury, but this is not a nephrotic or highly inflammatory urinary pattern. In the setting of severe hypertension, this fits **hypertensive nephrosclerosis/TMA-related renal injury**.<br>- **Anemia + thrombocytopenia suggest TMA, but not classic fulminant MAHA/TTP:** Hb **6.2 g/dL** and platelets **15,000/µL** are severe. However, **no schistocytes**, **normal bilirubin**, only **mild LDH elevation**, and **haptoglobin 45 mg/dL** do not show strong ongoing hemolysis. Thus, if TMA is present, it appears **kidney-dominant and relatively less hemolytic**, which is more compatible with **malignant hypertension-associated TMA** than classic TTP.<br>- **Low reticulocyte count (0.3%)** is notable: despite severe anemia, marrow response is poor. This may reflect **uremia-related suppression**, severe illness, or reduced erythropoietin production from acute renal failure, rather than brisk peripheral hemolysis.<br>- **High immature platelet fraction (15.3%)** suggests preserved marrow platelet production with **peripheral platelet consumption**, supporting a consumptive microangiopathic process rather than marrow failure.<br>- **Coagulation studies are not consistent with DIC:** PT/aPTT/fibrinogen are essentially preserved; D-dimer is only modestly elevated. This favors **TMA over DIC**.<br>- **Hypertension-mediated vascular injury is supported by end-organ findings:** prior **KW grade III hypertensive retinopathy** plus reduced intrarenal arterial blood flow on ultrasound strongly support **malignant hypertension with vascular damage**.<br>- **RAAS activation** (plasma renin activity **15.6 ng/mL**) fits **severe renal ischemia from malignant hypertension** and may perpetuate BP elevation.<br>- **Electrolyte abnormalities** (hyponatremia, severe hyperphosphatemia, hypocalcemia) are typical consequences of advanced renal failure.<br>- **Inflammatory/infectious and autoimmune causes are not supported:** CRP is normal, cultures are negative, and ANA/ANCA/anti-GBM/antiphospholipid-related testing are negative.<br>- **Typical HUS is unlikely:** no diarrhea or bloody stool.<br>- **TTP becomes less likely:** severe thrombocytopenia is compatible, but the overall pattern—marked malignant hypertension, severe renal failure out of proportion to hemolysis, retinal injury, and no clear MAHA picture—argues against classic TTP.<br>- **Complement-mediated TMA (aHUS) remains possible but less favored:** low **C3 (59 mg/dL)** raises interest, but C4 is preserved, FH-related assay is not supportive, and the clinical picture is strongly explained by malignant hypertension. Complement activation may also be **secondary** to endothelial injury. |
| With Static RAG (Model B) | ## 5. Interpretation of the test results<br>- **Severe renal failure dominates the presentation**: BUN **290.6 mg/dL** and creatinine **24.4 mg/dL** indicate extremely advanced kidney injury. The **normal-sized kidneys** suggest this may be an acute/subacute process superimposed on not-yet-atrophic kidneys rather than long-standing small contracted CKD.<br>- **Hypertensive renal vascular injury is strongly supported**: prior malignant-range BP, grade III hypertensive retinopathy, and **reduced interlobar arterial flow on ultrasound** fit **malignant hypertension-associated renal ischemic injury**. The |

| | |
|---|---|
| | **high plasma renin activity (15.6 ng/mL)** is also consistent with renal ischemia/activation of the renin-angiotensin system.<br>- **TMA is possible, but the pattern is not classic for TTP**:<br> - Severe **thrombocytopenia** (platelets **1.5 × $10^4$/mm$^3$**) and anemia (**Hb 6.2 g/dL**) raise concern for TMA.<br> - However, **no schistocytes**, **bilirubin is normal**, **LDH only mildly elevated**, and **haptoglobin is not clearly depleted**, so there is not strong evidence of brisk microangiopathic hemolysis.<br> - **Coagulation studies are essentially preserved** (PT/aPTT/fibrinogen near normal), arguing **against DIC**.<br> - **Immature platelet fraction 15.3%** suggests peripheral platelet consumption/destruction rather than marrow failure.<br>- **Anemia is severe but not fully explained by hemolysis alone**:<br> - **Normocytic anemia** with **very low reticulocytes (0.3%)** indicates poor marrow response, which can occur in severe uremia and critical illness; it is less typical of active MAHA, where reticulocytes usually rise.<br> - **WT1 mRNA negative** argues against TTP according to the marker.<br>- **Urinalysis supports intrinsic renal injury**:<br> - **Proteinuria 1.8 g/day** and **microscopic hematuria** are compatible with malignant nephrosclerosis/TMA-related renal injury, though not specific.<br>- **Complement findings are nonspecific but not strongly supportive of aHUS**:<br> - **Low C3 (59 mg/dL)** may reflect complement activation, but **C4/CH50 are not markedly depressed**, and the assay did **not confirm FH-related complement regulatory defects**. This does not completely exclude aHUS, but it weakens it.<br>- **Autoimmune, vasculitic, anti-GBM, antiphospholipid, infectious, and diarrheal HUS clues are absent**:<br> - Serologies are negative, cultures are negative, and there is no diarrhea/bloody stool.<br>- **Electrolyte abnormalities reflect advanced renal failure**:<br> - **Hyponatremia**, **severe hyperphosphatemia**, and **hypocalcemia** are typical of profound kidney dysfunction.<br> - Potassium is not elevated, but this does not lessen the severity of renal failure. |
| With Static RAG and Mult-agent framework (Model C) | ## 5. Interpretation of the test results<br>- **Severe renal failure is the dominant abnormality.**<br> BUN **290.6 mg/dL** and creatinine **24.4 mg/dL** indicate profound kidney dysfunction, matching her **oliguria, nausea, appetite loss, and hypertensive emergency history**. The **normal-sized kidneys** suggest an acute or acute-on-chronic process rather than long-standing end-stage atrophic CKD.<br><br>- **Urinary findings support intrinsic renal microvascular/parenchymal injury.**<br> Proteinuria (**1.8 g/day**) and microscopic hematuria (**10–19 RBC/HPF**) are compatible with **malignant hypertension-related renal injury / nephrosclerosis with possible TMA**, though they are not specific enough to exclude other renal parenchymal disease.<br><br>- **Anemia and thrombocytopenia are both severe.**<br> Hb **6.2 g/dL** and platelets **15,000/µL** are major abnormalities. The **immature platelet fraction of 15.3%** suggests increased peripheral platelet consumption rather than pure marrow failure. The **reticulocyte count of 0.3%** is lower than expected for active hemolysis and may reflect **inadequate marrow response, severe uremia, or a later presentation**, so it does not exclude TMA.<br><br>- **Evidence for overt microangiopathic hemolysis is limited, but TMA is not excluded.** |

| | |
|---|---|
| | The absence of **schistocytes**, normal bilirubin, only mildly low haptoglobin, and only modest LDH elevation argue **against florid MAHA/TTP**, but **absence of schistocytes does not rule out TMA**, especially in a renal-predominant presentation.<br><br>- **Coagulation is largely preserved, arguing against DIC.**<br>PT, aPTT, fibrinogen, and antithrombin are not consistent with consumptive coagulopathy. Mild **D-dimer elevation** is nonspecific.<br><br>- **Complement results raise but do not prove complement-mediated TMA.**<br>**Low C3 with normal C4** suggests alternative pathway activation. This can be seen in **complement-mediated TMA**, but it is **supportive rather than diagnostic**. The negative FH-related hemolytic assay does **not rule out aHUS/complement-mediated TMA**.<br><br>- **Autoimmune, infectious, and immune cytopenic workup is unrevealing.**<br>Negative ANA, ANCA, anti-GBM, antiphospholipid-related antibody, Coombs, and anti-platelet antibody tests, together with **negative cultures** and **no diarrheal illness**, make vasculitis, lupus, immune hemolysis, typical HUS, and overt infection-related processes less likely.<br><br>- **The hypertension-related renal injury hypothesis is strengthened.**<br>**Elevated plasma renin activity** and **reduced interlobar arterial blood flow** on ultrasound fit **renal ischemia from malignant hypertension / malignant nephrosclerosis**.<br><br>- **Electrolyte abnormalities reflect advanced uremia.**<br>Hyponatremia, severe **hyperphosphatemia (16.4 mg/dL)**, and marked **hypocalcemia (6.2 mg/dL)** are consistent with advanced kidney failure and support the need for **urgent renal replacement therapy**. |
| **Phase II – Q6: Reconsidering the differential diagnosis** | |
| Baseline LLM (Model A) | **6. Updated diagnostic thinking / Working diagnosis**<br><br>The differential now shifts away from primary hematologic TMA and toward **malignant hypertension causing secondary thrombotic microangiopathy with severe acute kidney injury**.<br><br>**Most likely working diagnosis:**<br>**Malignant hypertension (hypertensive emergency) with hypertension-associated renal TMA / malignant nephrosclerosis causing severe AKI, anemia, and thrombocytopenia.**<br><br>**Why this is now most likely:**<br>- Clear antecedent **severe hypertension with hypertensive retinopathy**<br>- **Renal failure is extremely severe** and central to the case<br>- TMA-like thrombocytopenia is present, but **hemolysis is limited**<br>- Coagulation is not consistent with DIC<br>- Autoimmune, infectious, and shiga-toxin HUS clues are absent<br>- TTP is less likely because the pattern is **kidney-predominant with severe hypertension**, not classic systemic MAHA/CNS-predominant TTP<br><br>**Still important alternative:** **complement-mediated TMA (aHUS)**, especially because of low C3 and severe renal involvement, but based on the provided information it is **less probable than hypertension-associated TMA**. |

| | |
|---|---|
| With Static RAG (Model B) | ## 6. Updated diagnostic thinking / Working diagnosis<br>The differential now shifts further toward:<br><br>1. **Malignant hypertension with severe renal involvement and secondary TMA-like thrombocytopenia**<br>2. **Primary TMA (especially TTP)** becomes less likely<br>3. **Complement-mediated TMA/aHUS** remains a consideration but is less favored<br><br>### Why malignant hypertension is now most likely<br>- The clinical picture is anchored by **documented malignant hypertension with hypertensive retinopathy/papilledema**, followed by profound renal failure.<br>- The renal vascular findings and high renin support **hypertensive/ischemic renal injury**.<br>- Although thrombocytopenia is severe, the laboratory evidence for **active MAHA is weaker than expected for TTP**: no schistocytes, only mild LDH rise, normal bilirubin, and non-suppressed haptoglobin.<br>- DIC, autoimmune hemolysis, vasculitis, anti-GBM disease, and infectious causes are not supported.<br><br>### Working diagnosis<br>**Malignant hypertension (hypertensive emergency) causing severe renal failure, most likely malignant nephrosclerosis / hypertension-associated renal TMA, with secondary thrombocytopenia and anemia.** |
| With Static RAG and Mult-agent framework (Model C) | ## 6. Updated diagnostic thinking / Working diagnosis<br>The results shift the differential further **away from DIC and primary TTP**, because:<br>- coagulation studies are preserved,<br>- there is no infectious or diarrheal trigger,<br>- hemolysis evidence is incomplete rather than classic,<br>- and the clinical context remains dominated by **preceding extreme hypertension with hypertensive retinopathy and severe renal failure**.<br><br>### Working diagnosis<br>**Malignant hypertension / malignant nephrosclerosis with severe AKI and suspected secondary TMA**<br><br>### Why this is now the leading diagnosis<br>- prior **BP 225/150 mmHg**<br>- **Keith–Wagener class III hypertensive retinopathy**<br>- severe **renal failure with oliguria**<br>- proteinuria and hematuria<br>- **elevated renin** and reduced intrarenal arterial flow<br>- severe thrombocytopenia compatible with **secondary microvascular platelet consumption**<br><br>### Important competing diagnoses that remain<br>1. **Complement-mediated TMA (aHUS)**<br>Still an important alternative because of:<br>- **low C3**<br>- severe renal-predominant disease<br>- marked thrombocytopenia<br>Current data do not confirm it, but they also do **not exclude** it. |

| | |
|---|---|
| | 2. **TTP**<br>Now less likely than malignant hypertension-associated TMA, but not completely dismissed because platelet count is very low (**15,000/µL**). **ADAMTS13 testing is still needed urgently** before concern for TTP can be fully relaxed.<br><br>3. **Primary renal parenchymal disease with malignant hypertension**<br>Less likely based on current serologies and overall pattern, but still should remain in reserve until follow-up or biopsy clarifies the pathology. |
| **Phase II – Q7: Treatment planning** | |
| Baseline LLM (Model A) | **7. Treatment plan**<br><br>### A. Immediate management<br>1. **Urgent renal replacement therapy (hemodialysis)**<br>- Indicated by extreme azotemia, oliguria, uremic symptoms, and major electrolyte/mineral derangements.<br>- This is the most urgent supportive treatment.<br><br>2. **Careful blood pressure control**<br>- Continue/adjust antihypertensive therapy with a goal of **gradual reduction**, avoiding overly rapid lowering that may worsen renal/brain ischemia.<br>- In a hypertensive emergency, standard practice is to reduce mean arterial pressure cautiously over hours to days.<br>- Choice of agents depends on real-time status; IV drugs are used if unstable, while oral agents may be sufficient if already partly controlled and no acute neurologic/cardiac instability is present.<br><br>3. **Red blood cell transfusion**<br>- Reasonable because Hb is **6.2 g/dL** and she is symptomatic/critically ill.<br><br>### B. Management of suspected TMA<br>4. **Close reassessment for primary TMA**<br>- Send/confirm **ADAMTS13 activity** if not already available.<br>- If clinical suspicion for TTP remains substantial, **plasma exchange should be considered without delay** because missing TTP is dangerous.<br>- However, based on current data, the leading strategy is to prioritize **BP control + dialysis**, as malignant hypertension-associated TMA is most likely.<br><br>5. **Consider complement-mediated TMA only if the course is atypical**<br>- If thrombocytopenia/renal failure do not improve with BP control, or biopsy/labs later support complement-mediated TMA, **eculizumab/ravulizumab** would become a reasonable option.<br>- At present, evidence from the provided data is insufficient to make this first-line therapy.<br><br>### C. Further evaluation once stabilized<br>6. **Kidney biopsy when safe**<br>- Helpful to distinguish **malignant nephrosclerosis/hypertension-associated TMA** from other causes of intrinsic renal disease or complement-mediated TMA.<br>- Must wait until platelet count and bleeding risk are acceptable.<br><br>### D. Other supportive care<br>7. **Manage CKD/AKI complications** |

| | |
|---|---|
| | - Correct calcium/phosphate imbalance, monitor potassium, nutrition, and volume status.<br>8. **Smoking cessation and long-term BP management**<br>- Essential for secondary prevention.<br><br>### Rationale and alternatives<br>- The current evidence best supports **secondary TMA from malignant hypertension**, so the most evidence-based immediate treatment is **controlled BP lowering plus dialysis and supportive care**.<br>- **Plasma exchange** is the key alternative if TTP cannot be confidently excluded.<br>- **Complement inhibition** is an alternative if the clinical course suggests **aHUS** rather than hypertension alone. |
| With Static RAG (Model B) | ## 7. Treatment plan<br>### Immediate treatment priorities<br>1. **Urgent renal replacement therapy (hemodialysis)**<br>- Rationale: She has **extreme azotemia**, oliguria, and major electrolyte/mineral abnormalities from severe renal failure. Even without hyperkalemia, this degree of uremia strongly supports dialysis.<br><br>2. **Careful blood pressure control**<br>- Use titratable antihypertensive therapy initially if needed, then oral agents.<br>- Aim to lower BP **gradually**, not abruptly, to avoid worsening renal/brain ischemia.<br>- Rationale: This is hypertensive emergency physiology; renal and retinal injury are ongoing.<br><br>3. **Supportive transfusion management**<br>- **Red blood cell transfusion** for symptomatic severe anemia.<br>- Platelet transfusion only if there is active bleeding or an invasive procedure is required, since platelet consumption may be ongoing in TMA states.<br><br>### Further diagnostic/therapeutic steps<br>4. **Kidney biopsy when safe**<br>- Rationale: If platelet count and BP can be stabilized, biopsy would help distinguish **malignant nephrosclerosis/hypertensive TMA** from primary TMA or other intrinsic renal diseases, and guide prognosis.<br><br>5. **Monitor for TMA evolution**<br>- Serial CBC, LDH, bilirubin, haptoglobin, smear, renal parameters.<br>- If future data strongly suggest TTP (e.g., ADAMTS13 severe deficiency or evolving MAHA/CNS findings), treatment should be escalated urgently.<br><br>### Role of plasma exchange / complement inhibition<br>- **Plasma exchange**: not the leading choice based on current data, because TTP is not strongly supported. However, if clinical suspicion remains high or the patient deteriorates before exclusion of TTP, empiric plasma exchange can be justified.<br>- **Eculizumab**: considered only if **complement-mediated TMA/aHUS** becomes more convincing after further evaluation; present data do not make it the first choice.<br><br>### Overall rationale<br>The most evidence-based initial strategy is to treat this as **malignant hypertension with dialysis-requiring renal failure**, while continuing to reassess for a primary TMA |

<table>
<tr><td></td><td>that would require disease-specific therapy.</td></tr>
<tr><td>With Static RAG and Mult-agent framework (Model C)</td><td>## 7. Treatment plan<br>### Immediate treatment<br>1. **Urgent hemodialysis / renal replacement therapy**<br>- Strongly indicated by:<br>- extreme azotemia,<br>- oliguria,<br>- severe hyperphosphatemia,<br>- hypocalcemia,<br>- likely symptomatic uremia.<br>- Dialysis access planning and nephrology co-management are essential.<br><br>2. **Careful blood pressure control**<br>- Continue management as **hypertensive emergency with renal end-organ injury**.<br>- BP should be lowered **gradually**, avoiding overly rapid reduction that could worsen renal ischemia.<br>- Use titratable therapy initially, then transition to oral multidrug treatment as stable.<br><br>3. **Transfusion support**<br>- **Red blood cell transfusion** is appropriate for Hb **6.2 g/dL**, especially if symptomatic.<br>- **Platelet transfusion** should generally be reserved for **active bleeding or an invasive procedure**, because thrombocytopenia appears consumptive/TMA-related.<br><br>### Urgent additional evaluation and monitoring<br>4. **Immediate hematology and nephrology co-management**<br>- The combination of severe thrombocytopenia, AKI, and possible TMA warrants joint management.<br><br>5. **Send urgent ADAMTS13 testing**<br>- TTP is less likely, but still too important to ignore with platelets at **15,000/µL**.<br><br>6. **Serial monitoring for TMA evolution**<br>- Repeat:<br>- CBC with smear,<br>- LDH,<br>- bilirubin,<br>- haptoglobin,<br>- reticulocyte count,<br>- renal function,<br>- neurologic assessment.<br>- This helps detect evolving MAHA or clarify response to BP control.<br><br>7. **Correct renal failure-related metabolic abnormalities**<br>- Manage **calcium/phosphate abnormalities** and other electrolyte issues in conjunction with dialysis and nephrology guidance.<br><br>### Disease-specific escalation options<br>8. **Plasma exchange**<br>- Not automatic from the current data, because classic TTP features are weak.<br>- However, the threshold should depend on **overall clinical suspicion**, not only on unmistakable later deterioration.</td></tr>
</table>

| | |
|---|---|
| | - Escalate if:<br>- ADAMTS13 shows severe deficiency,<br>- neurologic findings emerge,<br>- hemolysis becomes clearer,<br>- or thrombocytopenia fails to improve as expected.<br><br>9. **Kidney biopsy**<br>- **Defer for now** because platelet count is severely low.<br>- Consider only after stabilization and safer hemostatic conditions.<br>- It may help distinguish **malignant nephrosclerosis/secondary TMA** from **complement-mediated TMA** or another intrinsic renal disease.<br><br>10. **Eculizumab / complement blockade**<br>- Not first-line at this stage.<br>- Consider if the course remains strongly suspicious for **complement-mediated TMA**, especially if renal failure and thrombocytopenia do not improve despite BP control and supportive care, or if biopsy/expanded complement evaluation supports that diagnosis.<br><br>## Changes from the initial interpretation<br>- I now state more explicitly that the picture supports **malignant hypertension with possible secondary TMA**, but **does not fully exclude complement-mediated TMA**.<br>- I have **reduced overconfidence** in using absent schistocytes and limited hemolysis to move away from TMA; **absence of schistocytes does not rule it out**.<br>- I revised the interpretation of the **low reticulocyte count** to note that it may reflect **inadequate marrow response or later presentation**, not just uremia.<br>- I reframed the diagnosis as **malignant hypertension/malignant nephrosclerosis with AKI and suspected secondary TMA**, while keeping **aHUS as a meaningful competing diagnosis**.<br>- I added that **ADAMTS13 testing is urgently needed** before concern for TTP can be fully relaxed.<br>- I strengthened the treatment section by specifying **urgent nephrology/hematology co-management**, **serial hemolysis monitoring**, **dialysis access/metabolic management**, and clearer triggers for escalation to **plasma exchange** or **complement blockade**. |
| **Phase III – Q8: Evaluation of treatment** | |
| Baseline LLM (Model A) | **8. Monitoring of treatment effectiveness, safety, risks, and countermeasures**<br><br>### A. Effectiveness monitoring<br>- **Blood pressure**<br>- Monitor frequently during the acute phase, then daily trends.<br>- Goal: stable BP reduction without organ hypoperfusion.<br>- A favorable sign is improvement in urine output, platelet count, and renal function after BP control.<br><br>- **Renal status**<br>- Daily: **urine volume, serum creatinine/BUN, electrolytes, acid-base status, body weight, volume status**.<br>- Assess readiness for dialysis withdrawal by increasing urine output and improving biochemical parameters.<br>- Long-term: outpatient **sCr/eGFR, proteinuria, urinalysis**, because chronic kidney dysfunction may persist. |

- **TMA/hematologic activity**
  - Serial **platelet count, hemoglobin, LDH, haptoglobin, bilirubin, peripheral smear**, and if available **VWF-related markers**.
  - Response is indicated by rising platelet count, stabilization of Hb, and reduction of hemolysis markers.

- **Clinical end-organ recovery**
  - Follow **neurologic symptoms, visual symptoms, and general condition**.
  - Ophthalmologic follow-up is appropriate because hypertensive retinopathy was present.

#### B. Safety monitoring and specific risks

##### 1. Risks from BP lowering
- **Risk:** overly rapid BP reduction causing cerebral, cardiac, or renal ischemia.
- **Monitoring:** frequent BP checks, mental status, chest symptoms, urine output, renal function.
- **Countermeasure:** lower BP gradually and titrate IV antihypertensives carefully.

##### 2. Dialysis/CHDF-related risks
- **Risks:** hypotension, bleeding from anticoagulation, catheter infection, electrolyte shifts.
- **Monitoring:** hemodynamics during dialysis, catheter site, fever, serial electrolytes.
- **Countermeasure:** adjust ultrafiltration, careful anticoagulation strategy, strict catheter care, prompt culture/workup if infection is suspected.

##### 3. Plasma exchange (PE)-related risks
- **Risks:** allergic reactions, hypocalcemia from citrate, catheter complications, infection, coagulopathy, loss of immunoglobulins.
- **Monitoring:** vital signs during PE, calcium levels/symptoms, CBC/coagulation, catheter status, serum IgG if repeated sessions are needed.
- **Countermeasure:** premedication/supportive treatment if reactions occur, calcium supplementation when needed, aseptic catheter management, reassess need for further PE promptly once TTP is excluded.

##### 4. Caplacizumab-related risks
- **Main risk:** **bleeding**, because caplacizumab interferes with VWF-platelet interaction.
- **Monitoring:** clinical bleeding (mucosal, GI, urinary, injection-site), Hb trend, platelet count; extra caution before invasive procedures such as renal biopsy.
- **Countermeasure:** stop caplacizumab once TTP is excluded, avoid unnecessary invasive procedures while on treatment, and if major bleeding occurs, discontinue the drug and provide supportive hemostatic management.

##### 5. Transfusion-related risks
- **Risks:** transfusion reaction, volume overload, hyperkalemia.
- **Monitoring:** symptoms during/after transfusion, volume status, potassium.
- **Countermeasure:** transfuse only as needed, monitor closely, and manage reactions immediately.

##### 6. Residual or progressive CKD
- **Risk:** incomplete renal recovery despite acute improvement.

| | |
|---|---|
| | - **Monitoring:** long-term renal function, BP, proteinuria.<br>- **Countermeasure:** strict long-term BP control, nephrology follow-up, CKD complication management. |
| With Static RAG (Model B) | **8. Monitoring effectiveness, safety, risks, and countermeasures**<br><br>**Effectiveness monitoring**<br>- **Blood pressure control:** frequent BP monitoring, especially during ICU/IV antihypertensive treatment, aiming for steady control without excessive rapid lowering.<br>- **Renal recovery:** daily urine output, body weight/volume status, serial **sCr/BUN/electrolytes**, and dialysis requirement. Key signs of success are rising urine output, improving azotemia, and ability to discontinue HD.<br>- **TMA/hematologic response:** serial **platelet count, hemoglobin, LDH, bilirubin, haptoglobin, and smear**. Improvement in **Plt/Hb** and reduction of VWF-related activity support response.<br>- **End-organ injury:** follow symptoms/signs of hypertensive emergency such as headache, visual symptoms, and neurologic status.<br>- **After biopsy/discharge:** monitor **renal function, urinalysis/proteinuria, and BP** long term, since residual CKD remained.<br><br>**Safety monitoring / key risks**<br>- **Over-rapid BP reduction** causing renal, cerebral, or myocardial hypoperfusion.<br>- **Countermeasure:** use titratable IV agents, gradual BP lowering, and close neurologic/cardiovascular/renal monitoring.<br>- **Dialysis/CHDF complications**: hypotension, disequilibrium, catheter infection/thrombosis, electrolyte shifts.<br>- **Countermeasure:** close hemodynamic monitoring, sterile catheter care, frequent electrolyte checks, individualized ultrafiltration/dialysis prescription.<br>- **Caplacizumab-related bleeding** (major specific risk).<br>- **Countermeasure:** monitor for mucosal, gastrointestinal, urinary, and procedure-related bleeding; follow Hb and clinical bleeding signs; avoid unnecessary invasive procedures/antithrombotics; stop caplacizumab if significant bleeding occurs.<br>- **Plasma exchange complications**: allergic reactions, hypocalcemia from citrate, infection/line complications, coagulation factor loss.<br>- **Countermeasure:** monitor vitals during PE, calcium and coagulation status as needed, line care, and watch for transfusion reactions.<br>- **FFP exposure risks**: volume overload, allergic reactions, TRALI.<br>- **Countermeasure:** assess respiratory/volume status and stop or modify treatment if reactions occur.<br>- **Anemia/thrombocytopenia-related complications**: tissue hypoxia or bleeding.<br>- **Countermeasure:** serial CBC, transfuse RBCs as clinically needed, and reserve platelet transfusion for active bleeding or procedures.<br>- **Renal biopsy complications**: bleeding after biopsy, especially in a patient with recent thrombocytopenia and severe HT.<br>- **Countermeasure:** perform only after BP and platelets improve; monitor post-biopsy vitals, Hb, hematuria, and flank pain. |
| With Static RAG and Mult-agent framework | ### 8. Monitoring of treatment effectiveness and safety<br><br>**Effectiveness**<br><br>- **Primary marker of response: blood pressure control with renal recovery**<br>- In this case, treatment effectiveness should be judged **mainly by response to BP |

| | |
|---|---|
| (Model C) | reduction and renal support**, not by assuming benefit from caplacizumab/PE alone.<br> - Monitor **frequent BP**, avoiding both persistent severe hypertension and excessive hypotension.<br><br>- **Renal recovery**<br> - Follow **daily urine output, fluid balance, body weight, serum creatinine/BUN, electrolytes, and dialysis requirement**.<br> - In this patient, meaningful improvement would be:<br> - gradual increase in urine volume,<br> - improvement in azotemia,<br> - transition from **CHDF to intermittent HD**,<br> - and eventual **withdrawal from HD**, as occurred on hospital day 19.<br><br>- **Hematologic/TMA activity**<br> - Track **serial CBC**, especially **Hb and platelet count**, together with **peripheral smear, LDH, haptoglobin, and bilirubin**.<br> - Improvement is suggested by:<br> - rising platelet count,<br> - stabilization/improvement of Hb beyond transfusion effect,<br> - and reduction in laboratory evidence of hemolysis/endothelial injury.<br> - Because the initial hemolysis profile was limited, these markers should be interpreted longitudinally rather than relying on any single value.<br><br>- **Supportive adjunctive markers**<br> - **VWF-related activity** and **complement levels** may be followed if available, but they should be treated as **supportive rather than decisive** markers.<br> - In this case, disappearance of VWF activation and normalization of complement after treatment may support improvement, but they do not by themselves establish which therapy caused it.<br><br>- **After biopsy-confirmed MHT-related chronic TMA**<br> - Long-term follow-up should include **BP, serum creatinine, proteinuria, and urinalysis**, because residual CKD is likely.<br> - This fits the actual course, with persistent renal impairment (sCr around 3.5 mg/dL at 10 months).<br><br>**Safety / complications to monitor**<br><br>- **Overly rapid BP lowering**<br> - Risk: worsening renal ischemia or cerebral hypoperfusion.<br> - Plan: ICU-level monitoring with **titratable IV antihypertensives**, especially early in the course.<br><br>- **Dialysis-related complications**<br> - Risk: hypotension, catheter-related infection, bleeding, disequilibrium, electrolyte shifts.<br> - Plan: close hemodynamic and electrolyte monitoring, catheter care, and dialysis modality adjustment, as was done from **CHDF to intermittent HD**.<br><br>- **Red cell transfusion complications**<br> - Risk: volume overload, transfusion reaction, hyperkalemia.<br> - Plan: monitor vitals, potassium, and volume status; transfuse only as needed.<br><br>- **Plasma exchange complications** |

| | |
|---|---|
| | - Risk: allergic reaction, hypocalcemia, coagulopathy, infection from central access, and **immunoglobulin reduction**.<br>- Plan: monitor **calcium, coagulation, access complications, and IgG levels**, as was done after PE.<br><br>- **Caplacizumab-related bleeding**<br>- This is a major safety issue.<br>- Plan: monitor for **mucosal, gastrointestinal, urinary, catheter-site, and procedure-related bleeding**.<br>- This is especially important **before and after renal biopsy**, since biopsy increases bleeding risk.<br>- Once **ADAMTS13 activity was 42% with no inhibitor**, stopping caplacizumab/PE was appropriate to reduce unnecessary bleeding risk because **iTTP was excluded**.<br><br>- **Missed alternative TMA or persistent active disease**<br>- If thrombocytopenia, anemia/hemolysis, oliguria, or dialysis dependence persist despite BP improvement, reconsider whether MHT alone explains the course.<br>- Continue to integrate **ADAMTS13 results, complement data, biopsy findings, and clinical trajectory**.<br><br>**Changes from my initial version**<br>- I now emphasize more clearly that **effectiveness should be judged primarily by BP control and renal support**, since the observed recovery may not be attributable mainly to caplacizumab/PE.<br>- I strengthened the plan for **serial hemolysis monitoring** with CBC/smear/LDH/haptoglobin/bilirubin.<br>- I added more explicit **bleeding surveillance**, especially around **caplacizumab use and renal biopsy**.<br>- I now frame **VWF/complement trends as adjunctive, not diagnostic or decisive**, markers. |
| **Phase III – Q9: Formulating plans for worsening conditions** | |
| Baseline LLM (Model A) | **9. If the condition does not improve as expected or changes suddenly: reassessment and action plan**<br><br>### A. Reassess the working diagnosis quickly<br>If thrombocytopenia, anemia, or renal failure **do not improve after BP control and dialysis**, or if there is sudden worsening, I would reconsider whether this is truly only **MHT-associated TMA**.<br><br>Key alternative diagnoses to re-open:<br>- **TTP** if platelet count falls again, neurologic symptoms appear, or hemolysis worsens.<br>- **Complement-mediated TMA (aHUS)** if renal dysfunction and TMA persist despite BP improvement.<br>- **DIC/sepsis** if coagulation abnormalities, fever, or infection signs appear.<br>- Other secondary causes of TMA or severe hypertension if the course becomes atypical.<br><br>### B. Immediate actions depending on the change<br><br>#### 1. If hematologic TMA worsens<br>- Repeat **CBC, smear, LDH, bilirubin, haptoglobin, coagulation tests**. |

| | |
|---|---|
| | - Recheck/expedite **ADAMTS13** if not yet definitive.<br>- If TTP cannot be excluded clinically, **restart or continue plasma exchange urgently**, with TTP-directed treatment as needed.<br><br>#### 2. If renal recovery does not occur<br>- Reassess volume status, dialysis adequacy, BP control, and urinary findings.<br>- Reconsider **complement-mediated TMA** or another intrinsic kidney disease.<br>- Use **kidney biopsy** findings, when safely available, to revise the diagnosis.<br><br>#### 3. If sudden neurologic, cardiac, or visual deterioration occurs<br>- Treat as recurrent or uncontrolled **hypertensive emergency** until proven otherwise.<br>- Perform urgent organ-specific evaluation (for example, brain imaging if neurologic symptoms develop).<br>- Intensify BP management while avoiding excessive BP reduction.<br><br>#### 4. If bleeding develops<br>- Suspect **caplacizumab-related bleeding**, especially around procedures.<br>- Stop caplacizumab, assess severity, support hemostasis, and reconsider the balance of benefit and risk.<br><br>### C. How the diagnosis/treatment plan should be revised flexibly<br>- **If ADAMTS13 is severely deficient/inhibitor positive:** manage as **iTTP**, continue PE/caplacizumab and add immunosuppression.<br>- **If TTP is excluded and biopsy supports chronic hypertensive TMA:** stop TTP-specific therapy and focus on **BP control, renal support, and CKD follow-up**.<br>- **If complement-mediated TMA becomes more likely:** consider **complement inhibition** after excluding infection and reviewing pathology/clinical course.<br><br>In short, the response to treatment itself is diagnostic: **improvement with BP control supports MHT-associated TMA**, while persistent or relapsing TMA should trigger rapid reconsideration of **TTP, aHUS, infection/DIC, or another secondary process**. |
| With Static RAG (Model B) | **9. If the condition does not improve or suddenly changes: reassessment and action plan**<br><br>**How to think**<br>- Reassess whether the working diagnosis remains correct:<br>1. **Persistent malignant hypertension-related TMA**,<br>2. **Missed primary TMA** such as **TTP** or **complement-mediated TMA/aHUS**,<br>3. Other causes of AKI/cytopenias such as bleeding, infection, DIC, drug effect, or another intrinsic renal disease.<br>- Use the pattern of change:<br>- **Platelets not rising / hemolysis worsening / neurologic symptoms appearing** → think more strongly about **TTP/ongoing TMA**.<br>- **Renal recovery absent despite BP control** → reconsider **aHUS, irreversible chronic vascular injury, or another renal diagnosis**.<br>- **Sudden respiratory/hemodynamic change** → look for dialysis/PE/transfusion complications, pulmonary edema, infection, or bleeding.<br><br>**What to do**<br>- **Repeat urgent labs:** CBC, smear, LDH, bilirubin, haptoglobin, coagulation |

| | |
|---|---|
|  | profile, sCr/BUN, electrolytes, urinalysis.<br>- **Recheck ADAMTS13/inhibitor results and complement data** if not yet available or if the course is atypical.<br>- **Reassess BP trend and volume status** to confirm adequate but safe BP control.<br>- **Search for treatment complications**: bleeding from caplacizumab/biopsy, line infection, transfusion reaction, dialysis-related hypotension.<br>- **Escalate disease-specific therapy if new evidence supports it**:<br> - If TTP becomes more likely, continue/intensify **PE-based therapy** and TTP-directed treatment.<br> - If complement-mediated TMA becomes more likely, consider complement-targeted therapy after specialist reassessment.<br>- **Use kidney biopsy findings** when available to revise the diagnosis; in this case, biopsy supported **chronic TMA/glomerular microangiopathy due to malignant hypertension**, allowing discontinuation of unnecessary iTTP-directed immunosuppression.<br>- **Revise prognosis and long-term plan** if recovery is incomplete: prepare for CKD follow-up, strict BP control, and monitoring for recurrent TMA features. |
| With Static RAG and Mult-agent framework (Model C) | ### 9. Reassessment strategy if the condition does not improve as expected or suddenly changes<br><br>**How to think**<br><br>- If the patient had continued to have **thrombocytopenia, anemia, oliguria, or dialysis dependence despite BP improvement**, I would not assume these were all due to malignant hypertension alone.<br>- Reassessment should first focus on whether the problem is due to:<br> 1. **Uncontrolled or ongoing malignant hypertension-related injury**<br> 2. **Complement-mediated TMA / aHUS**<br> 3. **Another superimposed renal disease**<br> 4. **Treatment-related complications** such as bleeding, infection, or dialysis-related instability<br><br>- **TTP concern should be de-escalated once ADAMTS13 results are available.**<br> - In this case, **ADAMTS13 activity of 42% with no inhibitor essentially argues against immune TTP**.<br> - Therefore, any later nonresponse would shift concern more toward:<br> - persistent MHT-related vascular injury,<br> - complement-mediated TMA,<br> - chronic vascular/glomerular injury,<br> - or complications of dialysis/PE/caplacizumab,<br> - rather than reflexively restarting TTP-directed therapy.<br><br>**How to act**<br><br>- **Immediate reassessment**<br> - Repeat:<br> - BP trend and hemodynamic status<br> - neurologic examination<br> - volume status<br> - CBC with smear<br> - LDH, bilirubin, haptoglobin<br> - coagulation studies<br> - creatinine, BUN, electrolytes |

- urinalysis
- Also reassess dialysis adequacy and access-related issues.

- **If BP remains uncontrolled or fluctuates**
  - Intensify antihypertensive strategy, because ongoing hypertension itself may sustain renal and endothelial injury.

- **If thrombocytopenia/hemolysis persists after BP improvement and ADAMTS13 is not severely reduced**
  - Shift attention toward **complement-mediated TMA/aHUS** or another renal microangiopathic process.
  - Expand complement-focused evaluation and consider disease-specific therapy only if the overall picture supports it.

- **If fever or instability develops**
  - Search for **catheter-related infection**, PE-related infection risk, or other nosocomial complications.

- **If bleeding occurs**
  - Consider **caplacizumab-related bleeding**, PE-associated coagulopathy, uremic platelet dysfunction, or post-biopsy bleeding.
  - Stop or reverse the offending treatment as appropriate and provide supportive hemostatic care.

- **Use kidney biopsy once safe**
  - In this case, biopsy after stabilization was decisive.
  - It helped distinguish **chronic MHT-related TMA and glomerular microangiopathy** from iTTP and from other primary renal diseases.
  - Therefore, if the course is atypical and platelet count/bleeding risk permit, biopsy is a key step in revising the diagnosis.

**Bottom line**
Treatment response should be judged mainly by **stable BP control, increasing urine output, improving renal function, reduced dialysis dependence, and recovery of platelet count/Hb**. If the course is atypical, reassessment should prioritize **BP trajectory, serial hemolysis markers, infection/bleeding, and dialysis-related issues first**, while using **ADAMTS13 results to de-escalate TTP concern** and **kidney biopsy to clarify persistent diagnostic uncertainty**.

**Changes from my initial version**
- I now state more explicitly that after **ADAMTS13 42% with no inhibitor**, **iTTP is essentially ruled out**, so later deterioration should not automatically trigger renewed PE/caplacizumab.
- I reprioritized the contingency plan to first reassess **BP control, hemolysis profile, infection/bleeding, and dialysis-related complications**.
- I made **complement-mediated TMA** a more relevant alternative than TTP once ADAMTS13 results are known.
- I clarified that **kidney biopsy after platelet recovery/stabilization** is central for revising the diagnosis when the course remains unexplained.

**Case 5: Effective calcineurin inhibitor treatment in adult-onset steroid-resistant nephrotic syndrome with a novel splice donor site variant of TRPC6: a case report** [13]

| **Phase I – Q1: Summarizing medical problems** | |
|---|---|
| Baseline LLM (Model A) | 1. **Problem representation**<br><br>A **44-year-old woman** with **RA, SLE, and Sjögren's syndrome**, previously with **normal kidney function and normal urinalysis**, developed **acute-onset edema, 4-kg weight gain, oliguria, severe proteinuria, and hypoalbuminemia** in September 2021, consistent with **new-onset nephrotic syndrome**. Key issues are the **abrupt onset**, the background of **systemic autoimmune disease**, and the need to distinguish **glomerular disease related to SLE or SjS**, **drug-related disease**, and other primary causes of nephrotic syndrome. |
| With Static RAG (Model B) | ## 1. Problem representation<br>A **44-year-old woman** with **RA, SLE, and Sjögren's syndrome**, previously with **normal renal function and normal urinalysis**, developed **acute-onset leg edema, 4 kg weight gain, oliguria, severe proteinuria, and hypoalbuminemia** in **September 2021**, consistent with a **new nephrotic syndrome**. Her autoimmune diseases were reportedly **clinically stable**, with RA controlled on **abatacept**, and SLE/Sjögren's not requiring active treatment.<br><br>**Key points**<br>- Acute onset nephrotic syndrome<br>- Prior normal kidney function/urinalysis suggests a **new renal process**<br>- Important background of **systemic autoimmune disease**<br>- Exposure to **biologic therapy (abatacept)**<br>- Need to distinguish **glomerular disease related to SLE/Sjögren's**, **drug-related disease**, and other causes of nephrotic syndrome |
| With Static RAG and Mult-agent framework (Model C) | ## 1. Problem representation<br>A 44-year-old woman with autoimmune overlap syndrome (RA, SLE, and Sjögren's syndrome), previously with **normal renal function and normal urinalysis**, developed **acute nephrotic syndrome** in September 2021, characterized by **leg edema, 4 kg weight gain, oliguria, severe proteinuria, and hypoalbuminemia**, with apparently preserved baseline creatinine. This presentation suggests a **new glomerular process causing nephrotic syndrome**, and the cause requires **biopsy-based diagnosis**. |
| **Phase I – Q2: Differential diagnoses and rationale** | |
| Baseline LLM (Model A) | 2. **Differential diagnosis**<br><br>**Top 3 differential diagnoses**<br>1. **Lupus nephritis, especially membranous lupus nephritis (class V)**<br>- Most likely because she has established SLE and now presents with nephrotic syndrome.<br>- Class V lupus nephritis often presents with **heavy proteinuria, edema, and preserved creatinine** early on.<br>- Even if extrarenal SLE seems clinically stable, renal involvement can newly appear.<br><br>2. **AA amyloidosis secondary to RA**<br>- RA can cause AA amyloidosis with **nephrotic-range proteinuria**.<br>- Less likely here because her RA is reportedly **well controlled**, but it remains important given the chronic inflammatory background.<br><br>3. **Minimal change disease or membranous nephropathy, including secondary forms** |

| | |
|---|---|
| | - Acute nephrotic syndrome with preserved renal function can be seen in **minimal change disease**.<br>- **Membranous nephropathy** may be primary or secondary to autoimmune disease.<br>- Drug-related podocytopathy should also be considered, though no clear offending agent is provided.<br><br>**Most likely diagnosis:**<br>- **Lupus nephritis, particularly class V (membranous lupus nephritis).**<br><br>**Must not be missed:**<br>- **Proliferative lupus nephritis (class III/IV, with or without class V)** because it may require urgent immunosuppressive treatment to prevent irreversible kidney damage.<br>- Also important not to miss **renal vein thrombosis or thromboembolic complications** associated with nephrotic syndrome. |
| With Static RAG (Model B) | ## 2. Differential diagnosis<br>### Top 3 differential diagnoses<br>1. **Lupus nephritis, especially membranous lupus nephritis (Class V)**<br>- SLE is a major risk factor for nephrotic-range proteinuria.<br>- Class V lupus nephritis often presents with **preserved creatinine and nephrotic syndrome**.<br>- Even if extrarenal SLE seems quiescent, renal disease can still newly appear.<br><br>2. **Membranous nephropathy (primary or secondary, including autoimmune-associated)**<br>- Nephrotic syndrome with preserved kidney function is a classic presentation.<br>- Secondary membranous nephropathy may be associated with **SLE, Sjögren's, or other autoimmune disease**.<br><br>3. **Minimal change disease or other podocytopathy, including drug-associated disease**<br>- Acute onset edema, oliguria, severe proteinuria, and previously normal urinalysis fit a podocytopathy.<br>- Autoimmune disease and biologic exposure raise the possibility of **secondary minimal change disease** or another immune-mediated podocyte injury.<br><br>### Most likely<br>**Membranous lupus nephritis (Class V lupus nephritis)** is most likely, because:<br>- She has established **SLE**<br>- Presentation is dominated by **nephrotic syndrome with initially preserved creatinine**<br>- This pattern is highly compatible with **membranous lupus nephritis**<br><br>### Must not be missed<br>**Proliferative lupus nephritis (Class III/IV ± V)** must not be missed, because:<br>- It may initially coexist with nephrotic syndrome<br>- It has major implications for **urgent immunosuppressive treatment and prognosis**<br>- Missing active inflammatory lupus nephritis could lead to irreversible kidney damage<br><br>A second important "must not miss" consideration is **renal vein thrombosis / thromboembolic complications of nephrotic syndrome**, especially if severe |

| | |
|---|---|
| | hypoalbuminemia is present. |
| With Static RAG and Mult-agent framework (Model C) | ## 2. Prioritized differential diagnosis<br>### 1. **Membranous lupus nephritis (class V lupus nephritis)**<br>- Most likely given the history of SLE and the nephrotic presentation with previously preserved renal function.<br>- Class V lupus nephritis often presents with **heavy proteinuria, edema, and relatively normal creatinine**.<br>- Renal lupus activity may occur even when extrarenal SLE appears clinically quiet.<br><br>### 2. **Primary or secondary membranous nephropathy**<br>- Membranous nephropathy is a classic cause of nephrotic syndrome in adults.<br>- It may be **primary** or **secondary** to autoimmune disease, drugs, infection, or other conditions.<br>- This deserves explicit consideration rather than attributing the presentation automatically to SLE.<br><br>### 3. **Minimal change disease, including drug-related disease**<br>- Acute onset nephrotic syndrome with preserved kidney function is compatible with minimal change disease.<br>- Drug exposure, especially **NSAIDs**, can trigger minimal change disease or membranous nephropathy.<br>- Less common than membranous lupus nephritis, but clinically plausible.<br><br>### 4. **AA amyloidosis secondary to chronic inflammatory disease**<br>- RA can lead to AA amyloidosis with nephrotic syndrome.<br>- However, this is **less likely** here because the RA has reportedly been well controlled and the presentation is acute rather than insidious.<br><br>### 5. **Sjögren-associated renal disease**<br>- Sjögren's syndrome more commonly causes **tubulointerstitial nephritis** or **renal tubular acidosis** than nephrotic syndrome.<br>- Glomerular disease can occur, but it is less typical than the diagnoses above.<br><br>### Must not miss<br>- **Proliferative lupus nephritis with active urinary sediment**, because it may progress rapidly and requires urgent treatment.<br>- **Thromboembolic complications of nephrotic syndrome** such as renal vein thrombosis, DVT, or PE. |
| **Phase I – Q3: Necessary physical examinations and reasons** | |
| Baseline LLM (Model A) | 3. **Additional history and physical examination**<br><br>**History**<br>- **Urine characteristics**: foamy urine, gross hematuria<br> - To assess severity of proteinuria and possible nephritic features.<br>- **Symptoms suggesting active SLE**: rash, oral ulcers, serositis, fever, alopecia, arthralgia<br> - To evaluate whether renal disease is part of systemic flare.<br>- **Timeline and medications**: recent biologics, NSAIDs, antibiotics, herbal drugs<br> - To assess for drug-induced nephrotic syndrome or interstitial disease.<br>- **Symptoms of thrombosis**: unilateral leg pain/swelling, dyspnea, chest pain<br> - Nephrotic syndrome increases thrombotic risk.<br>- **Infection symptoms** |

| | |
|---|---|
| | - Important before considering immunosuppression and because infection can trigger or mimic glomerular disease.<br>- **Blood pressure history and prior urinalysis trends**<br>- To determine acuity and evidence of evolving nephritis.<br><br>**Physical examination**<br>- **Blood pressure, volume status, extent of edema, weight**<br>- To assess nephrotic severity and complications.<br>- **Skin and mucosal exam** for rash, purpura, oral ulcers<br>- To look for active connective tissue disease or vasculitis.<br>- **Cardiopulmonary exam**<br>- To detect pleural effusion, pulmonary edema, or serositis.<br>- **Abdominal exam**<br>- For ascites related to severe hypoalbuminemia.<br>- **Examination for DVT**<br>- Because nephrotic syndrome predisposes to thrombosis. |
| With Static RAG (Model B) | ## 3. Additional history and physical examination<br>### Additional questions<br>- **Timeline of edema, weight gain, and urine reduction**<br>- To confirm acuity and severity<br>- **Foamy urine, gross hematuria, dark urine**<br>- Suggests glomerular disease; hematuria would support lupus nephritis or other glomerulonephritis<br>- **Recent SLE symptoms**: rash, oral ulcers, serositis, fever, alopecia, arthralgia worsening<br>- To assess whether renal involvement parallels systemic lupus activity<br>- **Recent infections**<br>- Infection-related glomerular disease or trigger of autoimmune flare<br>- **Medication history**, especially recent changes:<br>- NSAIDs, biologics, antibiotics, herbal drugs<br>- NSAIDs can cause minimal change disease/membranous nephropathy; biologics may rarely trigger immune renal disease<br>- **Thrombotic symptoms**: flank pain, dyspnea, chest pain, unilateral leg pain<br>- To screen for nephrotic syndrome complications such as renal vein thrombosis or PE<br>- **History suggesting malignancy**<br>- Because membranous nephropathy can be secondary to malignancy, though less likely at this age<br>- **Blood pressure history**<br>- Hypertension may suggest more active glomerular inflammation<br><br>### Physical examination<br>- **Volume assessment**: peripheral edema, sacral edema, ascites, JVP, lung crackles<br>- To assess severity of fluid retention<br>- **Blood pressure**<br>- Helps gauge renal involvement severity and urgency<br>- **Skin/mucosa/joints**<br>- Rash, oral ulcers, synovitis, alopecia suggesting active SLE/RA<br>- **Cardiopulmonary exam**<br>- Evaluate fluid overload and thromboembolic complications<br>- **Abdominal/flank exam**<br>- Flank tenderness may raise concern for renal vein thrombosis<br>- **Signs of DVT** |

| | |
|---|---|
| | - Important because nephrotic syndrome increases thrombosis risk |
| With Static RAG and Mult-agent framework (Model C) | ## 3. Additional questions and physical examination<br>### Focused history<br>- **Urine findings**: foamy urine, cola-colored urine, gross hematuria<br>- Helps distinguish pure nephrotic syndrome from a nephritic component.<br>- **Timing and progression of edema and oliguria**<br>- Clarifies acuity and severity.<br>- **History of hypertension** or recent blood pressure change<br>- Supports possible nephritic/glomerular inflammation.<br>- **Symptoms of lupus activity**: rash, oral ulcers, serositis, fever, alopecia, joint symptoms<br>- Assesses whether renal disease may reflect lupus activity, though renal involvement can occur despite systemic quiescence.<br>- **Prior complement and anti-dsDNA trends**<br>- Helpful for comparison with current results.<br>- **Detailed medication history**, especially recent NSAIDs, antibiotics, biologics, supplements, or herbal drugs<br>- Important for drug-induced nephrotic syndromes.<br>- **RA inflammatory symptoms**<br>- Relevant because uncontrolled chronic inflammation would make AA amyloidosis more plausible.<br>- **Thrombotic symptoms**: unilateral leg pain/swelling, flank pain, dyspnea, pleuritic chest pain<br>- Screens for nephrotic complications.<br>- **Infectious history**<br>- Some infections can cause secondary membranous nephropathy.<br><br>### Physical examination<br>- **Blood pressure and volume assessment**: edema distribution, ascites, pleural effusions, JVP<br>- Determines severity and whether a nephritic component or fluid overload is present.<br>- **Skin/mucosal exam**: malar rash, oral ulcers, purpura<br>- Looks for lupus or vasculitic activity.<br>- **Joint exam**<br>- Assesses current inflammatory disease activity.<br>- **Cardiopulmonary exam**<br>- Evaluates effusions, pulmonary edema, or alternate causes of edema.<br>- **Abdominal exam**<br>- Looks for ascites and organomegaly.<br>- **Examination for thrombosis**<br>- Important because nephrotic syndrome markedly increases thrombotic risk. |
| **Phase I – Q4: Plan for investigations/tests** | |
| Baseline LLM (Model A) | 4. **Tests to narrow the diagnosis**<br><br>1. **Urinalysis with microscopy and urine protein quantification (spot protein/creatinine ratio or 24-hour urine)**<br>- Purpose: confirm nephrotic-range proteinuria and assess for hematuria/casts.<br>- Expected findings:<br>- Heavy proteinuria.<br>- If lupus nephritis is proliferative, may see dysmorphic RBCs or casts.<br>- Bland sediment would fit membranous disease or minimal change disease. |

| | |
|---|---|
| | 2. **Serum chemistry: creatinine, eGFR, albumin, total protein, electrolytes, lipid profile**<br>- Purpose: evaluate kidney function and nephrotic syndrome severity.<br>- Expected findings:<br>- Hypoalbuminemia, hyperlipidemia; creatinine may still be normal.<br><br>3. **Immunologic tests: anti-dsDNA, complements (C3, C4, CH50), ANA if needed**<br>- Purpose: assess SLE activity.<br>- Expected findings:<br>- Elevated anti-dsDNA and low complement would support active lupus nephritis, though class V may show less dramatic abnormalities.<br><br>4. **CBC, CRP/ESR**<br>- Purpose: assess inflammation, cytopenias, and infection/inflammatory activity.<br>- Expected findings:<br>- Cytopenias may support lupus activity; inflammatory markers may help evaluate RA activity or amyloidosis context.<br><br>5. **Tests for secondary nephrotic causes as indicated: hepatitis B/C, HIV**<br>- Purpose: exclude secondary membranous nephropathy and important mimics before immunosuppression.<br>- Expected results:<br>- Often negative, but necessary to check.<br><br>6. **Renal ultrasound**<br>- Purpose: assess kidney size, exclude obstruction, provide procedural information before biopsy.<br>- Expected findings:<br>- Usually normal-sized kidneys in glomerular disease.<br><br>7. **Kidney biopsy**<br>- Purpose: definitive diagnosis; essential to distinguish **class V lupus nephritis**, **proliferative lupus nephritis**, **AA amyloidosis**, **minimal change disease**, or other pathology.<br>- Expected findings:<br>- Most likely: features of **membranous lupus nephritis**.<br>- Must determine whether there is any **class III/IV component**, which would change treatment and prognosis.<br>- Congo red positivity would support amyloidosis; podocyte foot process effacement would support minimal change disease.<br><br>Overall, the diagnostic priority is to **confirm nephrotic syndrome, assess for active lupus nephritis, and obtain a kidney biopsy**, because management depends critically on the histologic diagnosis. |
| With Static RAG (Model B) | ## 4. Tests to narrow the diagnosis<br>### Urine studies<br>1. **Urinalysis with microscopy**<br>- Purpose: detect hematuria, casts, lipiduria<br>- Expected findings:<br>- Heavy proteinuria confirms nephrotic syndrome<br>- Dysmorphic RBCs/RBC casts would support inflammatory glomerulonephritis such as lupus nephritis<br>- Bland sediment would fit membranous nephropathy or minimal change disease |

2. **Quantification of proteinuria**: spot urine protein/creatinine ratio or 24-hour urine protein
   - Purpose: confirm nephrotic-range proteinuria and monitor disease burden
   - Expected: nephrotic-range proteinuria

#### Blood tests
3. **Serum creatinine, BUN, electrolytes, albumin, total protein, lipid profile**
   - Purpose: assess kidney function, nephrotic severity, and complications
   - Expected: preserved or near-preserved creatinine, low albumin, hyperlipidemia

4. **Complement levels (C3, C4) and anti-dsDNA**
   - Purpose: assess lupus activity
   - Expected:
     - Low complements and elevated anti-dsDNA would support active lupus nephritis
     - Normal results would not fully exclude Class V lupus nephritis but may make active proliferative disease less likely

5. **ANA profile and immunologic testing if needed**
   - Purpose: characterize autoimmune background, though diagnosis of SLE/Sjögren's is already established

#### Secondary nephrotic syndrome evaluation
6. **HBV, HCV, HIV serologies**
   - Purpose: exclude infection-associated secondary membranous nephropathy or other glomerular disease
   - Expected: likely negative, but important before immunosuppression

7. **Anti-PLA2R antibody**
   - Purpose: help distinguish primary membranous nephropathy from secondary causes
   - Expected:
     - Positive would support primary membranous nephropathy
     - Negative would be more compatible with secondary membranous disease, including lupus-associated disease

#### Imaging
8. **Renal ultrasound**
   - Purpose: assess kidney size, exclude obstruction, provide baseline
   - Expected: usually normal-sized kidneys

9. **If thrombosis suspected: Doppler/CT imaging**
   - Purpose: evaluate renal vein thrombosis or DVT/PE
   - Expected: only if symptoms/signs raise suspicion

#### Definitive test
10. **Kidney biopsy**
   - Purpose: essential to distinguish among **Class V lupus nephritis, proliferative lupus nephritis, membranous nephropathy, minimal change disease, amyloidosis, or other glomerular disease**
   - Expected:
     - **Class V lupus nephritis**: membranous pattern with immune complex deposition, often "full-house" immunofluorescence
     - **Primary membranous nephropathy**: subepithelial deposits, often PLA2R-

| | |
|---|---|
| | related, without lupus-type full-house pattern<br>- **Minimal change disease**: diffuse foot process effacement with minimal light microscopy changes<br><br>## Bottom line<br>This patient has **new-onset nephrotic syndrome** on a background of **SLE/Sjögren's/RA**, and the leading concern is **membranous lupus nephritis**, while **proliferative lupus nephritis must be excluded urgently**. The most important next step is a **focused nephrotic workup plus kidney biopsy**, because management depends on the exact histologic diagnosis. |
| With Static RAG and Mult-agent framework (Model C) | ## 4. Testing plan<br>### Initial essential tests<br>1. **Urinalysis with urine microscopy**<br>- To confirm proteinuria and assess for **microscopic hematuria, dysmorphic RBCs, and casts**.<br>- Active sediment would raise concern for proliferative lupus nephritis.<br><br>2. **Quantification of proteinuria**<br>- Spot urine protein-creatinine ratio or 24-hour urine protein.<br>- Confirms nephrotic-range proteinuria and helps assess severity.<br><br>3. **Serum chemistry**<br>- Creatinine, BUN, electrolytes, albumin, total protein, lipid profile.<br>- Evaluates kidney function and consequences of nephrotic syndrome.<br><br>4. **CBC**<br>- Assesses cytopenias and overall systemic involvement.<br><br>5. **Lupus activity markers**<br>- **C3, C4, anti-dsDNA**<br>- These are supportive, not diagnostic.<br>- Low complement and high anti-dsDNA would support lupus nephritis, but normal values do not exclude class V disease.<br>- **ANA is not particularly useful here for current renal assessment**, since SLE is already established.<br><br>6. **Inflammatory markers**<br>- ESR and CRP.<br>- Persistent inflammation would make AA amyloidosis more plausible, though this remains less likely.<br><br>### Additional targeted tests<br>7. **Anti-PLA2R antibody**<br>- Helpful if membranous nephropathy is suspected, especially to evaluate for primary membranous nephropathy.<br><br>8. **Renal ultrasound**<br>- Excludes obstruction, confirms kidney size, and prepares for biopsy.<br><br>9. **Secondary workup if indicated**<br>- Hepatitis B/C serologies and serum/urine protein electrophoresis.<br>- Considered if the clinical picture suggests alternative causes of membranous disease or paraproteinemia. |

<table>
<tr><td></td><td>### Decisive test<br>10. **Kidney biopsy**<br>- This is the **key and urgent test** because substantial nephrotic syndrome in this setting cannot be accurately classified by serology alone.<br>- It is required to distinguish among:<br>  - **Class V lupus nephritis**<br>  - Primary/secondary membranous nephropathy<br>  - Minimal change disease<br>  - AA amyloidosis<br>  - Other less common glomerular lesions<br><br>## Changes from the initial version<br>- **Problem representation revised** to emphasize **acute nephrotic syndrome with preserved baseline renal function** and the need for a **biopsy-based diagnosis**.<br>- **Differential diagnosis reprioritized**:<br>  - Kept **class V lupus nephritis** first<br>  - Moved **primary/secondary membranous nephropathy** higher<br>  - Kept **minimal change disease** as a major consideration<br>  - **De-emphasized AA amyloidosis** because the RA was well controlled and the presentation was acute<br>  - Added clearer note that **Sjögren's usually causes tubulointerstitial rather than nephrotic glomerular disease**<br>- **History/physical expanded** to include:<br>  - **Hypertension**<br>  - **Microscopic hematuria/nephritic features**<br>  - **Prior complement/anti-dsDNA trends**<br>  - **Exact medication timing**<br>- **Testing plan refined** by:<br>  - Adding **urine microscopy detail**<br>  - Adding **anti-PLA2R testing**<br>  - Clarifying that **ANA is not helpful for current lupus renal activity**<br>  - Framing **kidney biopsy as central and urgent**.</td></tr>
<tr><td colspan="2">Phase II – Q5: Interpreting test results</td></tr>
<tr><td>Baseline LLM (Model A)</td><td>**5. Interpretation of the test results**<br><br>- The patient has **full nephrotic syndrome**: massive proteinuria (**19.29 g/day**, PCR **19.34 g/g**), **severe hypoalbuminemia** (**1.6 g/dL**), edema, weight gain, oliguria, and **elevated LDL** (**159 mg/dL**). This explains her fluid retention and is clinically significant because it implies high risk of **thrombosis, infection, and progressive kidney injury**.<br>- **Serum creatinine 1.65 mg/dL** represents a clear decline from her baseline (**0.57 mg/dL**), indicating **new acute kidney dysfunction**. This is important because pure nephrotic syndromes such as minimal change disease or membranous nephropathy may preserve kidney function early, whereas the creatinine rise raises concern for more active glomerular injury, hemodynamic effects from severe nephrosis, or less likely another superimposed process.<br>- Urine microscopy shows only **5–9 RBCs/HPF**, i.e., **mild hematuria**. This suggests a **glomerular process**, but the sediment is not strongly nephritic. In context, this fits better with a **membranous pattern** (including class V lupus nephritis) than with highly inflammatory proliferative GN.<br>- The immunologic data are especially important: **anti-dsDNA is negative/minimally positive and C3/C4 are normal**, arguing **against active proliferative lupus</td></tr>
</table>

| | |
|---|---|
| | nephritis**. Similarly, there is no serologic evidence of active vasculitis or anti-GBM disease.<br>- Strong **anti-SS-A positivity** confirms underlying Sjögren's serology but does **not by itself explain massive nephrotic-range proteinuria**. Sjögren's kidney disease more often causes tubulointerstitial nephritis than severe nephrotic syndrome.<br>- Negative **HBsAg/HCV antibody** reduces the likelihood of common infection-related secondary membranous nephropathy.<br><br>**Overall meaning:** the results strongly support a **severe nephrotic glomerular disease**, with the pattern favoring a **non-proliferative immune-mediated lesion**, especially **membranous lupus nephritis (class V)**, although biopsy is still essential. |
| With Static RAG (Model B) | **5. Interpretation of the test results**<br><br>- The patient has **full nephrotic syndrome**:<br> - **Massive proteinuria** (UPCR 19.34 g/g; 24-h urine protein 19.29 g/day)<br> - **Marked hypoalbuminemia** (albumin 1.6 g/dL)<br> - **Edema and weight gain**<br> - **Hyperlipidemia** (LDL 159 mg/dL)<br> These findings explain her leg edema and oliguria and indicate severe glomerular capillary permeability injury.<br><br>- **Serum creatinine 1.65 mg/dL** represents **new acute kidney dysfunction** compared with her baseline 0.57 mg/dL. This is important because her presentation is not just isolated nephrotic syndrome; there is also renal function decline, which broadens concern beyond a purely benign podocytopathy.<br><br>- **5–9 RBCs/HPF** indicates **microscopic hematuria**, supporting a **glomerular process** rather than a purely noninflammatory cause of edema. However, the hematuria is relatively mild, so the picture is still dominated by nephrotic-range proteinuria.<br><br>- The **normal C3/C4** and **negative/minimally positive anti-dsDNA** make **active immune-complex lupus flare, especially proliferative lupus nephritis, less likely**, though they do **not exclude lupus-related renal disease**, particularly **membranous lupus nephritis (Class V)**, which may occur with relatively quiet serologies.<br><br>- Strong **anti-SS-A positivity** confirms the Sjögren background but does not by itself explain this degree of nephrotic syndrome. Sjögren renal disease more often causes **tubulointerstitial nephritis** than massive nephrotic proteinuria.<br><br>- Negative **HBV/HCV**, **ANCA**, and **anti-GBM** reduce the likelihood of major alternative secondary causes of glomerulonephritis.<br><br>**Particular points of interest**<br>1. **Proteinuria is extremely heavy (~19 g/day)**, which strongly points to a **podocyte/membranous-type glomerular lesion**.<br>2. **Renal dysfunction is new**, so this is a clinically significant active renal disease, not just laboratory nephrosis.<br>3. **Serologies do not support an active systemic lupus flare**, shifting attention away from proliferative lupus nephritis and toward **membranous lupus nephritis or another nonproliferative nephrotic glomerulopathy**. |

| | |
|---|---|
| With Static RAG and Mult-agent framework (Model C) | ## 5. Interpretation of the test results<br><br>- The patient has **full nephrotic syndrome**:<br> - **Massive proteinuria**: urine protein/creatinine ratio **19.34 g/g** and 24-hour urine protein **19.29 g/day**.<br> - **Severe hypoalbuminemia**: albumin **1.6 g/dL**.<br> - **Edema** and **hyperlipidemia**: LDL **159 mg/dL**.<br> These findings explain her **leg edema** and **weight gain** and confirm that this is not mild proteinuria but a severe nephrotic presentation.<br><br>- **Renal function is newly impaired**:<br> - Creatinine rose to **1.65 mg/dL** from a baseline of **0.57 mg/dL**, indicating **new kidney injury**.<br> - In this setting, the rise in creatinine broadens concern beyond isolated nephrotic syndrome to include **AKI related to the nephrotic state**, **intravascular volume depletion**, or **superimposed active glomerular inflammation**.<br><br>- **Urine microscopy shows mild microscopic hematuria**:<br> - **5–9 RBCs/HPF** is **compatible with glomerular disease**, but by itself is **not specific** without dysmorphic RBCs or casts.<br> - The overall picture still remains **predominantly nephrotic rather than overtly nephritic**.<br><br>- **The serologic pattern does not support active proliferative lupus nephritis**:<br> - **Anti-dsDNA is negative/minimally positive** and **C3/C4 are normal**.<br> - This makes **active proliferative lupus nephritis less likely**, but **does not exclude class V lupus nephritis**, which can present with nephrotic syndrome and relatively inactive serologies.<br><br>- **RA and Sjögren serologies do not indicate active systemic inflammatory disease**:<br> - Strong **anti-SS-A** confirms the Sjögren background but does **not by itself explain massive nephrotic syndrome**.<br> - Reportedly stable RA makes **AA amyloidosis less likely**, though not impossible.<br><br>- **Negative hepatitis B/C, ANCA, and anti-GBM testing**:<br> - These results make several important secondary or vasculitic causes less likely and narrow the differential toward **membranous-pattern disease or another primary glomerular lesion**.<br><br>**Key point:**<br>This is **severe nephrotic syndrome with new kidney dysfunction and mild microscopic hematuria, without strong serologic evidence of active proliferative lupus**. The pattern most strongly suggests a **membranous-pattern glomerulopathy**, but biopsy is required. |
| **Phase II – Q6: Reconsidering the differential diagnosis** | |
| Baseline LLM (Model A) | **6. Updated diagnostic thinking / Working diagnosis**<br><br>**Revised differential diagnosis priority**<br>1. **Membranous lupus nephritis (class V lupus nephritis)** — now most likely<br> - Nephrotic syndrome is very prominent. |

| | |
|---|---|
| | - SLE is established.<br>- Normal complements and negative/low anti-dsDNA make **proliferative lupus nephritis less likely**, while class V can occur with relatively quiescent serology.<br>- Mild hematuria is compatible.<br><br>2. **Primary or secondary membranous nephropathy**<br>- Also compatible with heavy proteinuria, edema, and relatively bland urine sediment.<br>- Autoimmune disease could also be a secondary trigger.<br><br>3. **Minimal change disease / podocytopathy**<br>- Massive proteinuria fits, but the hematuria and autoimmune background make this less likely than membranous disease.<br><br>**Less likely now**<br>- **AA amyloidosis**: possible with RA history, but less favored because RA is reportedly well controlled and the presentation is more suggestive of immune glomerular disease.<br>- **Proliferative lupus nephritis (class III/IV)**: not excluded, but now lower priority because serologic activity is absent and urinary sediment is not strongly nephritic.<br>- **ANCA-associated GN / anti-GBM disease**: unlikely given negative serology and nephrotic-predominant presentation.<br><br>**Working diagnosis:**<br>**Nephrotic syndrome most likely due to membranous lupus nephritis (class V lupus nephritis), pending kidney biopsy confirmation.** |
| With Static RAG (Model B) | **6. Updated diagnostic thinking / Working diagnosis**<br><br>The differential now shifts as follows:<br><br>1. **Membranous lupus nephritis (Class V lupus nephritis)** — **most likely**<br>- Fits the combination of **very heavy nephrotic proteinuria**, **hypoalbuminemia**, **edema**, and **relatively quiet lupus serology**.<br>- Mild hematuria can be seen.<br>- Lupus nephritis can newly appear even when extrarenal SLE seems inactive.<br><br>2. **Secondary membranous nephropathy related to autoimmune disease (including SjS/SLE overlap)**<br>- Also compatible with severe nephrotic syndrome and relatively bland serologic inflammatory activity.<br><br>3. **Minimal change disease / podocytopathy, possibly drug-related**<br>- Acute nephrotic syndrome is compatible, but the presence of **microscopic hematuria** and **renal dysfunction** makes this somewhat less straightforward.<br><br>4. **Proliferative lupus nephritis (Class III/IV ± V)**<br>- Now less favored because **complements are normal** and **anti-dsDNA is not elevated**, and the presentation is more nephrotic than nephritic; however, it still cannot be excluded without biopsy.<br><br>**Working diagnosis:**<br>**Nephrotic syndrome due to glomerular disease, most likely membranous lupus nephritis (Class V lupus nephritis), pending kidney biopsy.** |

| | |
|---|---|
| With Static RAG and Mult-agent framework (Model C) | ## 6. Updated diagnostic thinking / Working diagnosis<br><br>### Revised prioritization of the differential diagnosis<br><br>1. **Membranous-pattern nephropathy: either class V lupus nephritis or primary/secondary membranous nephropathy** — **highest priority**<br>- This is now the leading category because the presentation is dominated by:<br>- **very heavy proteinuria**<br>- **severe hypoalbuminemia**<br>- **edema**<br>- **mild hematuria**<br>- **weak serologic evidence of active proliferative lupus**<br>- On the available data, **class V lupus nephritis and primary/secondary membranous nephropathy should be considered nearly co-equal**, because serologies alone cannot reliably distinguish them.<br><br>2. **Minimal change disease**<br>- Still possible because it can cause abrupt, severe nephrotic syndrome.<br>- However, the **creatinine rise** and **microscopic hematuria** make it somewhat less typical than membranous disease.<br><br>3. **FSGS or another primary glomerular disease**<br>- Also possible in an adult with nephrotic syndrome and renal dysfunction.<br>- Not favored over membranous disease on the current data, but should remain in the biopsy-based differential.<br><br>4. **AA amyloidosis**<br>- Less likely because the RA is reportedly well controlled and the presentation appears relatively acute rather than chronic and progressive.<br><br>5. **Sjögren-associated glomerular disease**<br>- Possible but less typical, since Sjögren's more commonly causes **tubulointerstitial nephritis** than severe nephrotic syndrome.<br><br>### Working diagnosis<br>**Severe nephrotic syndrome due to a membranous-pattern glomerular disease, most likely either class V lupus nephritis or primary/secondary membranous nephropathy, pending kidney biopsy.** |
| **Phase II – Q7: Treatment planning** | |
| Baseline LLM (Model A) | **7. Treatment plan**<br><br>**Immediate management of nephrotic syndrome and AKI**<br>- **Hospital-based supportive care**<br>- Salt restriction and careful fluid management.<br>- **Loop diuretics** for edema, with close monitoring of renal function and volume status.<br>- **RAAS blockade (ACE inhibitor or ARB)** if blood pressure, potassium, and kidney function allow<br>- To reduce intraglomerular pressure and proteinuria.<br>- **Statin therapy** may be considered for hyperlipidemia, especially if nephrotic syndrome persists.<br>- **Assess thrombosis risk carefully** |

| | |
|---|---|
| | - Severe hypoalbuminemia (**1.6 g/dL**) and massive proteinuria place her at high thrombotic risk; prophylactic anticoagulation may be considered after balancing bleeding risk.<br>- Monitor creatinine, urine output, electrolytes, albumin, body weight, and complications.<br><br>**Disease-specific management**<br>- **Kidney biopsy should be performed promptly** because treatment depends on histology.<br>- If biopsy confirms **class V lupus nephritis with nephrotic syndrome**, standard evidence-based options include:<br>- **Glucocorticoids plus mycophenolate mofetil**, or<br>- **Glucocorticoids plus a calcineurin inhibitor** (tacrolimus/cyclosporine), depending on severity and patient factors.<br>- **Hydroxychloroquine** is generally recommended in SLE unless contraindicated, as background therapy because it improves renal and overall lupus outcomes.<br><br>**Rationale**<br>- Supportive therapy addresses the immediate consequences of nephrotic syndrome and AKI.<br>- Immunosuppressive therapy is justified if biopsy confirms immune-mediated lupus nephritis, especially with **very heavy proteinuria** and declining renal function.<br>- Biopsy is critical because treatment differs markedly if the lesion is **membranous lupus nephritis, proliferative lupus nephritis, amyloidosis, or minimal change disease**.<br><br>**Other possible options depending on biopsy**<br>- If **proliferative lupus nephritis** is found: more intensive induction therapy such as **high-dose glucocorticoids plus MMF or cyclophosphamide**.<br>- If **primary membranous nephropathy**: treatment may shift toward membranous nephropathy-specific therapy.<br>- If **minimal change disease**: glucocorticoid-based therapy would be typical.<br>- If **AA amyloidosis**: the main strategy would be strict control of underlying inflammatory disease rather than lupus-directed immunosuppression.<br><br>**In summary:** begin supportive nephrotic syndrome management immediately, obtain a **renal biopsy urgently**, and plan **lupus nephritis-directed immunosuppression** if biopsy confirms **class V lupus nephritis**. |
| With Static RAG (Model B) | **7. Treatment plan**<br><br>## Immediate/supportive management<br>- **Hospital-based monitoring** of body weight, urine output, blood pressure, serum creatinine, electrolytes.<br>- **Salt restriction** and **loop diuretics** for symptomatic edema.<br>- Rationale: relieve fluid overload while monitoring for worsening intravascular depletion/AKI.<br>- Consider cautious **albumin plus loop diuretic** only if edema is severe and diuretic response is poor.<br>- **RAAS blockade (ACE inhibitor/ARB)** may help reduce proteinuria, but initiation should be cautious because she already has **AKI**.<br>- **Statin therapy** can be considered for nephrotic-associated hyperlipidemia, especially if proteinuria persists.<br>- Because albumin is **1.6 g/dL**, assess **thrombotic risk** carefully; nephrotic |

| | |
|---|---|
|  | syndrome markedly increases risk of venous thrombosis. Prophylactic anticoagulation depends on bleeding risk and overall clinical assessment.<br><br>## Disease-specific plan<br>- **Kidney biopsy is essential before definitive immunosuppressive treatment**, because treatment differs substantially among:<br> - Class V lupus nephritis<br> - Proliferative lupus nephritis<br> - Primary/secondary membranous nephropathy<br> - Minimal change disease<br><br>## If biopsy confirms membranous lupus nephritis (most likely)<br>- Start **glucocorticoid-based therapy plus an immunosuppressive agent**, commonly:<br> - **Mycophenolate mofetil (MMF)**, or<br> - **Calcineurin inhibitor** (tacrolimus/cyclosporine), depending on severity and patient factors.<br>- Rationale:<br> - Current evidence and guidelines support immunosuppression for **nephrotic-range Class V lupus nephritis**, especially with severe proteinuria and renal dysfunction.<br> - MMF is widely used because of efficacy in lupus nephritis and steroid-sparing potential.<br> - Calcineurin inhibitors are alternatives, especially when rapid antiproteinuric effect is desired.<br><br>## If biopsy shows proliferative lupus nephritis<br>- More intensive induction therapy would be needed, typically:<br> - **High-dose glucocorticoids + MMF or cyclophosphamide**.<br><br>## If biopsy shows minimal change disease/podocytopathy<br>- **High-dose corticosteroids** would be the main treatment, with review of possible drug triggers.<br><br>## Other options / considerations<br>- If membranous nephropathy unrelated to lupus is found, management would depend on whether it is **primary or secondary**.<br>- Review current medications, including **abatacept**, for possible temporal relationship to renal disease, although current data provided do not establish causality.<br>- Continue close surveillance for complications of nephrotic syndrome: **infection, thrombosis, worsening AKI, and severe edema**.<br><br>**Practical conclusion:**<br>At this stage, the best plan is **supportive nephrotic syndrome management plus urgent kidney biopsy**, with the leading expectation that treatment will likely require **immunosuppression for membranous lupus nephritis**. |
| With Static RAG and Mult-agent framework (Model C) | ## 7. Treatment plan<br><br>### Immediate priorities<br>- **Prompt kidney biopsy**<br> - This should be prioritized because the distinction among **class V lupus nephritis**, **membranous nephropathy**, **minimal change disease**, **FSGS**, and **amyloidosis** cannot be made reliably from serologies alone.<br> - **Empiric disease-specific immunosuppression should ideally be deferred until |

biopsy**, unless clinical deterioration makes immediate treatment unavoidable.

#### Supportive management of severe nephrotic syndrome
- **Salt restriction** and **careful loop diuretic therapy** for edema.
  - Because creatinine is elevated, diuresis should be **cautious**, with close monitoring for **intravascular depletion**, worsening AKI, weight change, urine output, and electrolytes.

- **Monitor renal function and volume status closely**
  - Serial creatinine, BUN, electrolytes, daily weight, edema, and urine output.

- **Assess thrombotic risk urgently**
  - Albumin is **1.6 g/dL** with massive proteinuria, placing her at **high risk for venous thromboembolism**.
  - She should be assessed for **DVT/PE/renal vein thrombosis symptoms**, and the need for prophylactic anticoagulation should be considered based on overall bleeding risk and local practice.

- **RAAS blockade**
  - May help reduce proteinuria, but should be used **cautiously** because of the **acute rise in creatinine**.

- **Treat hyperlipidemia**
  - A **statin** is reasonable if nephrotic syndrome persists.

#### Biopsy-directed treatment options
- **If class V lupus nephritis is confirmed**:
  - Use **glucocorticoid-based therapy plus an immunosuppressive agent**.
  - Common options include:
    - **mycophenolate mofetil + glucocorticoids**
    - or a **calcineurin inhibitor-based regimen** in selected cases.
  - This is justified by the **massive nephrotic syndrome** and impaired renal function.

- **If primary membranous nephropathy is confirmed**:
  - Continue optimized supportive care and consider immunosuppression according to risk.
  - Options may include **rituximab**, calcineurin inhibitor, or cyclophosphamide-based therapy in high-risk disease.

- **If minimal change disease is confirmed**:
  - **High-dose glucocorticoids** are standard first-line therapy.

- **If FSGS is confirmed**:
  - Treatment depends on subtype, but often includes supportive care and, in selected primary forms, immunosuppression.

- **If AA amyloidosis is confirmed**:
  - Focus on control of the underlying inflammatory disease.

#### Rationale
The major change in management emphasis is that this patient is now recognized as having **very high-risk nephrotic syndrome with AKI**, so supportive care must include not only edema treatment but also **careful avoidance of overdiuresis, explicit

| | |
|---|---|
|  | thrombosis risk assessment, and biopsy before targeted immunosuppression whenever possible**.<br><br>## Changes from the initial version<br>- I **softened the interpretation of hematuria**: 5–9 RBCs/HPF is now described as **compatible with but not specific for glomerular disease**, rather than clearly proving a glomerular process.<br>- I **removed overstatement about decreased urine output being established by the labs**, since oliguria comes from the clinical history, not the admission laboratory data alone.<br>- I **broadened the explanation for the creatinine rise** to include **AKI from nephrotic syndrome, intravascular depletion, or superimposed nephritis**, not just new glomerular disease.<br>- I **revised the differential** so that **class V lupus nephritis and primary/secondary membranous nephropathy are near co-leaders**, instead of making class V lupus nephritis alone clearly dominant.<br>- I **expanded the biopsy-based differential** to include **FSGS** and acknowledged that **Sjögren-associated glomerular disease** is possible but less typical.<br>- I **strengthened the treatment plan** by explicitly prioritizing:<br> - **biopsy before disease-specific immunosuppression**<br> - **cautious diuresis in the setting of AKI**<br> - **venous thromboembolism risk assessment**<br> - **cautious use of RAAS blockade** because creatinine is already elevated. |
| **Phase III – Q8: Evaluation of treatment** | |
| Baseline LLM (Model A) | **8. Monitoring effectiveness, safety, risks, and countermeasures**<br><br>**A. Effectiveness monitoring**<br>- **Renal function:** serum creatinine/eGFR, BUN, urine output, daily weight during acute phases; then regular outpatient follow-up.<br> - Goal: maintain renal recovery and detect relapse early.<br>- **Proteinuria:** spot urine protein-to-creatinine ratio (or equivalent) serially.<br> - Goal: assess remission status; note that in this case renal function improved more than proteinuria.<br>- **Edema/volume status:** physical exam, body weight, blood pressure.<br> - Goal: judge clinical response even when proteinuria remains high.<br>- **Serum albumin and lipids:** albumin, LDL cholesterol.<br> - Goal: track nephrotic syndrome activity and cardiovascular/thrombotic risk.<br>- **If on CNI:** therapeutic drug monitoring<br> - **CyA:** C2 level.<br> - **TAC:** trough level.<br> - Goal: keep within target range and avoid underexposure or toxicity.<br><br>**B. Safety monitoring and concrete countermeasures**<br>1. **CNI nephrotoxicity (CyA/TAC)**<br> - Monitor: creatinine/eGFR, potassium, blood pressure, drug levels; consider repeat biopsy if the course is unclear.<br> - Risks: afferent vasoconstriction, chronic tubulointerstitial injury.<br> - Countermeasures: dose adjustment, switch CNI if needed, avoid other nephrotoxins, reassess if creatinine rises after dose increase or despite stable disease.<br><br>2. **Steroid toxicity**<br> - Monitor: glucose, blood pressure, infection signs, psychiatric symptoms, muscle |

| | |
|---|---|
| | weakness, GI symptoms.<br>- Risks: infection, hyperglycemia, osteoporosis, myopathy, mood changes.<br>- Countermeasures: taper promptly when possible, infection surveillance, gastric and bone protection as appropriate, treat hyperglycemia if it appears.<br><br>3. **Infection due to immunosuppression**<br>- Monitor: fever, respiratory/urinary symptoms, CBC, inflammatory markers as clinically indicated.<br>- Risks: high because of steroids, CNI use, and baseline autoimmune disease/immunocompromised state.<br>- Countermeasures: low threshold for cultures/imaging, temporarily reduce immunosuppression if severe infection is suspected, avoid unnecessary additional immunosuppressants.<br><br>4. **Relapse/rebound after reduction of immunosuppression**<br>- Monitor: urine output, edema, body weight, creatinine, proteinuria soon after dose changes.<br>- Risk: already demonstrated after CyA dose reduction.<br>- Countermeasures: taper very cautiously, close early follow-up after any dose reduction, reinstate/intensify therapy promptly if relapse occurs.<br><br>5. **Nephrotic syndrome complications**<br>- **Thrombosis:** monitor for leg swelling, pain, dyspnea, chest pain.<br>- Countermeasure: prompt diagnostic evaluation and anticoagulation if thrombosis is confirmed.<br>- **Severe edema/AKI:** monitor urine output, weight, creatinine.<br>- Countermeasure: adjust diuretics/volume management and start renal replacement therapy if indicated, as previously required.<br>- **Hyperlipidemia:** monitor LDL.<br>- Countermeasure: lipid-lowering therapy as appropriate; LDL apheresis may improve cholesterol but is not reliable for proteinuria control.<br><br>6. **Persistent nephrotic proteinuria despite preserved creatinine**<br>- Monitor long-term trend in proteinuria, albumin, renal function.<br>- Risk: chronic kidney disease progression despite apparent short-term renal recovery.<br>- Countermeasures: continue the most effective tolerated antiproteinuric strategy; RAAS blockade is standard but may be limited here by low blood pressure.<br><br>7. **Drug-specific TAC adverse effects**<br>- Monitor: trough level, creatinine, potassium, glucose, neurologic symptoms, blood pressure.<br>- Countermeasures: dose adjustment and supportive treatment if adverse effects emerge. |
| With Static RAG (Model B) | ## 8. Monitoring effectiveness, safety, risks, and countermeasures<br><br>### A. Effectiveness monitoring<br>**Main treatment goals**<br>- Preserve kidney function<br>- Maintain adequate urine output and volume status<br>- Reduce edema<br>- Reduce proteinuria, even if complete remission is difficult<br>- Avoid dialysis re-initiation |

| | |
|---|---|
| | **What to monitor**<br>1. **Renal function**<br>- Serum creatinine, BUN, eGFR: initially frequent, then regular outpatient follow-up<br>- Watch for rising creatinine as a sign of relapse, CNI toxicity, volume depletion, or progression<br><br>2. **Proteinuria**<br>- Spot urine protein/creatinine ratio serially<br>- Complete remission may be unlikely in TRPC6-associated disease, but trends still matter<br><br>3. **Urine output / body fluid status**<br>- Daily urine volume during unstable periods<br>- Body weight, edema, blood pressure<br>- Helps assess response even when proteinuria lags behind<br><br>4. **Need for renal replacement therapy**<br>- Reassess for dialysis indications if oliguria, fluid overload, hyperkalemia, acidosis, or uremia recur<br><br>5. **If clinically indicated, pathology correlation**<br>- If course becomes atypical, reconsider whether evolving FSGS lesions, interstitial damage, or another superimposed process has appeared<br><br>### B. Safety monitoring of current treatment<br>Because she is on **tacrolimus (TAC)** and previously received **high-dose steroids/CNI therapy**, monitoring should focus on:<br><br>1. **CNI nephrotoxicity**<br>- Monitor serum creatinine, potassium, magnesium, blood pressure<br>- Measure **TAC trough level** and keep within target range<br>- Countermeasure:<br>- If creatinine rises, promptly distinguish relapse from CNI toxicity and adjust TAC dose<br>- Consider repeat biopsy if uncertainty persists or kidney injury progresses<br><br>2. **Persistent nephrotic syndrome complications**<br>- **Thrombosis risk**: severe hypoalbuminemia/proteinuria predispose to DVT/PE/renal vein thrombosis<br>- Watch for leg pain, unilateral swelling, chest pain, dyspnea, flank pain<br>- Countermeasure:<br>- Low threshold for urgent imaging if symptoms occur<br>- Supportive management of nephrotic syndrome and mobilization; anticoagulation depends on overall risk-benefit<br><br>3. **Infection**<br>- Risk increased by prior high-dose steroids, CNIs, nephrotic state, and autoimmune disease<br>- Watch for fever, respiratory symptoms, urinary symptoms, skin infection<br>- Countermeasure:<br>- Early culture/imaging when indicated<br>- Reduce immunosuppression if severe infection develops, balancing relapse risk |

| | |
|---|---|
| | 4. **Steroid adverse effects** (especially from prior pulse/ongoing exposure)<br>- Hyperglycemia, psychiatric symptoms, hypertension, infection, osteoporosis, myopathy, GI bleeding<br>- Countermeasure:<br>- Check glucose, monitor symptoms, provide bone and GI protection as appropriate, taper steroid when feasible<br><br>5. **Tacrolimus-specific toxicities**<br>- Neurotoxicity (tremor, headache), hyperglycemia, hypertension, electrolyte abnormalities<br>- Countermeasure:<br>- Regular labs and symptom review; dose adjustment if toxicity appears<br><br>6. **Progressive chronic kidney damage**<br>- Second biopsy already showed interstitial edema/fibrosis and tubular atrophy<br>- Countermeasure:<br>- Avoid unnecessary overexposure to CNI<br>- Continue close longitudinal monitoring of eGFR and urinary findings<br><br>7. **Low blood pressure**<br>- She could not receive renin–angiotensin system blockade because BP was around 100/50 mmHg<br>- Countermeasure:<br>- Continue BP monitoring; only reconsider antiproteinuric RAAS blockade if hemodynamically tolerated<br><br>### C. Practical monitoring plan<br>- **Frequent early follow-up** after any TAC dose change or clinical worsening<br>- At each visit:<br>- Symptoms: edema, urine volume, infection, thrombotic symptoms, neurologic symptoms<br>- Vitals: BP, weight<br>- Labs: creatinine, electrolytes, albumin, glucose, TAC trough, urine protein/Cr<br>- Escalate promptly if:<br>- creatinine rises,<br>- urine output falls,<br>- edema worsens,<br>- TAC level is high,<br>- infection/thrombosis is suspected |
| With Static RAG and Mult-agent framework (Model C) | ### 8. Monitoring of treatment effectiveness, safety, risks, and countermeasures<br><br>#### A. Effectiveness monitoring<br>- **Renal function:** serial **serum creatinine, BUN, eGFR**<br>- Main goal: confirm maintenance of renal recovery, since she had **dialysis-requiring AKI** and later relapse.<br>- **Proteinuria:** regular **spot urine protein/creatinine ratio**<br>- Important because **nephrotic-range proteinuria persists** despite improvement in creatinine.<br>- In this case, treatment success should not be judged by proteinuria alone, because **renal recovery occurred even without complete proteinuria remission**.<br>- **Urine output, edema, body weight, blood pressure**<br>- Essential for detecting early relapse, especially because prior deterioration |

presented with **oliguria and worsening edema**.
- **Serum albumin and lipid profile**
  - Reflect ongoing nephrotic activity and complication risk.
- **Urine sediment**
  - Helpful to detect changes suggesting a different or superimposed process.

##### B. Practical follow-up intensity
- **Especially close follow-up after any tacrolimus/CNI dose change**
  - Because relapse occurred **within 10 days of CyA dose reduction**.
  - Practical approach: reassess **creatinine, tacrolimus level, urine protein, weight/edema, and symptoms within days to 1–2 weeks** after any dose adjustment.
- During stable periods, continue **regular trend-based outpatient monitoring** of renal function, proteinuria, blood pressure, and tacrolimus exposure.

##### C. Safety monitoring for current treatment
1. **Calcineurin inhibitor nephrotoxicity / chronic CNI injury**
   - Important because the second biopsy already showed **interstitial fibrosis/edema, tubular simplification, and mild tubular atrophy**.
   - Monitor:
     - **serum creatinine/eGFR trend**
     - **tacrolimus trough** (target **5–8 ng/mL** as described)
     - **blood pressure**
     - **potassium, magnesium**
   - Countermeasures:
     - avoid excessive tacrolimus exposure
     - adjust dose to target trough
     - review long-term trends rather than isolated values
     - if renal decline is unexplained, consider reassessment, including **repeat biopsy only if clinically necessary**

2. **Persistent nephrotic syndrome complications**
   - Risks: **edema, thrombosis, infection, malnutrition, dyslipidemia**
   - Monitor:
     - **albumin, proteinuria, body weight, edema, lipid profile**
     - symptoms of **DVT/PE/renal vein thrombosis**
   - Countermeasures:
     - **salt restriction** and careful diuretic use as tolerated
     - maintain a **low threshold for VTE evaluation** because proteinuria remains nephrotic and albumin was previously very low
     - lipid-lowering therapy as appropriate
     - nutritional support

3. **Infection due to immunosuppression**
   - Relevant because she received **high-dose steroids**, prior **CyA**, now **tacrolimus**, and has autoimmune disease.
   - Monitor:
     - **fever, respiratory/urinary symptoms**
     - **CBC**
     - clinical examination
   - Countermeasures:
     - use the **minimum effective immunosuppression**
     - promptly evaluate and treat suspected infection
     - avoid unnecessary added immunosuppressive therapy

| | |
|---|---|
| | 4. **Steroid-related toxicity**<br>- Risks: **hyperglycemia, infection, myopathy, psychiatric effects, GI toxicity, bone toxicity**<br>- Monitor:<br>- **glucose/HbA1c**<br>- infection symptoms<br>- muscle weakness, mental status, GI symptoms<br>- Countermeasures:<br>- avoid repeated empiric steroid escalation unless clearly justified<br>- taper/discontinue as feasible<br>- provide standard preventive care as appropriate<br><br>5. **Tacrolimus-specific adverse effects**<br>- Risks: **neurotoxicity/tremor, diabetes, GI symptoms, electrolyte abnormalities, liver test abnormalities**<br>- Monitor:<br>- neurologic symptoms<br>- **glucose/HbA1c**<br>- **magnesium/potassium**<br>- **liver enzymes**<br>- tacrolimus trough level<br>- Countermeasures:<br>- dose reduction or regimen adjustment if toxicity appears<br><br>#### D. Special high-risk issue in this patient<br>- **Relapse precipitated by CNI reduction**<br>- This is a proven risk in this case.<br>- Countermeasure:<br>- avoid rapid tapering<br>- reduce dose only cautiously<br>- perform **early post-change monitoring**<br>- **Persistent proteinuria despite preserved kidney function**<br>- Ongoing nephrotic proteinuria should prompt continued surveillance for long-term complications even when creatinine remains near 1.0 mg/dL.<br><br>#### Changes from the initial version<br>- Added a **specific need for early follow-up after any CNI/tacrolimus dose change**, based on the relapse after CyA reduction.<br>- Expanded routine monitoring to include **CBC, liver enzymes, magnesium, glucose/HbA1c, and urine sediment**.<br>- Made **thrombosis surveillance more explicit**, with a lower threshold for VTE evaluation in persistent severe nephrosis.<br>- More clearly incorporated **long-term surveillance for chronic CNI tubulointerstitial toxicity**, supported by the second biopsy findings.<br>- Clarified that **effectiveness should be judged not only by proteinuria**, because this patient's renal function improved despite persistent nephrotic-range proteinuria. |
| **Phase III – Q9: Formulating plans for worsening conditions** | |
| Baseline LLM (Model A) | **9. If the condition does not improve as expected or suddenly changes: reassessment and action plan**<br><br>**A. Reassess the cause systematically**<br>If renal function worsens, urine output falls, edema increases, or proteinuria suddenly changes, reconsider whether the problem is: |

| | |
|---|---|
|  | 1. **Relapse/progression of podocytopathy**<br>2. **CNI toxicity**<br>3. **Hemodynamic/pre-renal AKI**<br>4. **Acute tubular injury**<br>5. **Infection-related decompensation**<br>6. **Renal vein thrombosis or other thromboembolism**<br>7. **An alternative glomerular diagnosis missed initially**, including evolution to **FSGS lesions**<br><br>**B. Immediate actions**<br>- Repeat:<br> - serum creatinine/electrolytes/albumin<br> - urine protein quantification and urinalysis<br> - CNI level<br> - body weight, blood pressure, urine output<br>- Review recent:<br> - medication changes, especially CNI dose reduction/increase<br> - intercurrent illness or infection<br> - volume status and nephrotoxin exposure<br><br>**C. When to revise diagnosis/treatment**<br>- **If deterioration follows CNI reduction:** suspect relapse of steroid-resistant nephrotic syndrome/podocytopathy and consider restoring effective immunosuppression, as was done.<br>- **If creatinine rises with high CNI exposure or without clear nephrotic worsening:** suspect CNI nephrotoxicity; reduce dose, switch agent, or re-biopsy if necessary.<br>- **If the course is atypical or treatment response diverges from expectation:** repeat kidney biopsy should be considered, especially to detect emerging **FSGS**, chronic scarring, or alternative pathology.<br>- **If infection is suspected:** evaluate and treat infection first, and reconsider the intensity of immunosuppression.<br>- **If oliguria/severe AKI recurs:** provide supportive care promptly, including dialysis when indicated.<br><br>**D. Role of the genetic result in future decisions**<br>- The **likely pathogenic TRPC6 variant** supports a **genetic podocytopathy/FSGS-spectrum disease**, explaining steroid resistance and incomplete proteinuria response.<br>- Therefore, if future deterioration occurs, avoid assuming active autoimmune nephritis without evidence; instead prioritize:<br> - relapse of genetic podocytopathy,<br> - progression to FSGS,<br> - or treatment toxicity.<br>- Treatment should remain pragmatic: preserve kidney function, avoid overtreatment with ineffective immunosuppression, and use biopsy/clinical course to guide changes. |
| With Static RAG (Model B) | ## 9. If the condition does not improve or suddenly changes: reassessment and action plan<br><br>### A. Reassess the cause systematically<br>If kidney function worsens, edema increases, or urine output falls, I would rapidly reconsider whether this is:<br>1. **Relapse/progression of podocytopathy**<br>2. **CNI nephrotoxicity** |

3. **Hemodynamic AKI** from volume issues
4. **Superimposed acute tubular injury/interstitial injury**
5. **Infection-related deterioration**
6. **Thrombotic complication**
7. **Evolution from MGA to FSGS or progression of chronic scarring**

#### B. Immediate actions
1. **Clinical reassessment**
   - Volume status, edema, blood pressure, urine output
   - Symptoms of infection or thrombosis

2. **Urgent laboratory review**
   - Creatinine, electrolytes, albumin
   - Urinalysis and urine protein/Cr
   - **TAC trough level**

3. **Medication review**
   - Recent dose changes in TAC/CyA or diuretics
   - Any new nephrotoxic drugs

4. **Imaging or additional testing if indicated**
   - Ultrasound if obstruction/structural issue is a concern
   - Thrombosis imaging if symptoms suggest DVT/PE/renal vein thrombosis

#### C. How I would revise diagnosis and treatment
- **If TAC level is high or findings suggest CNI toxicity**:
  - Reduce or temporarily hold TAC, depending on severity
  - Reassess kidney function closely

- **If relapse of nephrotic disease is more likely**:
  - Optimize TAC exposure within target
  - Reconsider short-term steroid intensification only if benefit outweighs infection risk

- **If the course is atypical or unexplained**:
  - Consider **repeat kidney biopsy**, as already done in this case, to distinguish persistent podocytopathy from FSGS evolution, chronic damage, or drug toxicity

- **If dialysis indications recur**:
  - Restart renal replacement therapy without delay

- **If infection is identified**:
  - Treat infection promptly and adjust immunosuppression as needed

#### D. Broader interpretation in this case
Given the identified **likely pathogenic TRPC6 variant**, I would keep in mind that:
- Persistent nephrotic proteinuria may reflect **genetic podocytopathy**, so complete remission may be difficult
- Treatment goals should emphasize **renal function preservation and complication prevention**, not only proteinuria normalization
- Apparent “steroid resistance” is biologically plausible, so repeated escalation of immunosuppression should be weighed carefully against toxicity

**Bottom line:**
If she worsens, I would quickly distinguish **disease relapse/progression** from

| | |
|---|---|
| | **treatment toxicity or a superimposed complication**, using symptoms, labs, TAC levels, and—if needed—repeat biopsy, then adjust therapy promptly with priority on preserving kidney function and avoiding avoidable immunosuppressive harm. |
| With Static RAG and Mult-agent framework (Model C) | ### 9. Thought process and action plan for rapid reassessment and flexible revision of diagnosis/treatment<br><br>#### A. Immediate reassessment framework<br>If the patient develops **rising creatinine, reduced urine output, worsening edema, or sudden clinical decline**, I would rapidly reconsider:<br>1. **Relapse of podocytopathy**<br>2. **Tacrolimus/CNI nephrotoxicity**<br>3. **Hemodynamic or volume-related AKI**<br>- e.g., intravascular depletion despite edema, or overdiuresis<br>4. **Progression of tubulointerstitial injury**<br>5. **Thrombotic complication of nephrotic syndrome**<br>6. **Infection or another superimposed acute illness**<br>7. **Need for diagnostic revision**<br>- including evolution to **FSGS** or another glomerular lesion not captured initially<br><br>Importantly, in this case **renal deterioration may recur even if proteinuria does not change much**, so worsening creatinine or oliguria should trigger evaluation even without a major rise in urine protein.<br><br>#### B. Immediate actions<br>- Repeat:<br>- **serum creatinine, BUN, electrolytes, albumin**<br>- **urinalysis/urine sediment and urine protein quantification**<br>- **tacrolimus trough level**<br>- Reassess:<br>- **urine output, weight, edema, blood pressure, volume status**<br>- Review:<br>- any **recent tacrolimus dose change**<br>- diuretics<br>- nephrotoxic drugs<br>- intercurrent illness<br>- Screen for complications:<br>- **infection symptoms/signs**<br>- **VTE symptoms**<br>- severe fluid overload or dehydration<br><br>#### C. Action plan according to cause<br>- **If relapse after CNI reduction or low tacrolimus exposure is suspected:**<br>- restore/optimize tacrolimus to the therapeutic target range<br>- monitor closely over the following days to weeks<br>- avoid abrupt future dose reductions<br><br>- **If tacrolimus toxicity is suspected:**<br>- reduce dose and reassess exposure and kidney function<br>- balance toxicity reduction against relapse risk<br><br>- **If severe edema, oliguria, or AKI recurs:**<br>- use **cautious supportive care** |

| | |
|---|---|
| | - hospitalize if there is significant renal decline, refractory edema, or oliguria<br>- provide dialysis again if indicated<br><br>- **If the course is atypical or renal decline is unexplained:**<br>- consider **repeat kidney biopsy**<br>- Purpose: distinguish relapse, chronic CNI injury, evolving FSGS, or another superimposed lesion<br><br>- **If thrombosis or infection is suspected:**<br>- investigate and treat promptly because either can rapidly worsen renal and overall prognosis<br><br>#### D. How the genetic result changes future management<br>- The **likely pathogenic TRPC6 splice-site variant** supports a **genetic podocytopathy / FSGS-spectrum disorder** and helps explain:<br>- steroid resistance<br>- persistent nephrotic-range proteinuria<br>- limited response of proteinuria despite therapy<br>- Therefore, if the condition does not improve as hoped, future management should favor:<br>- **preserving kidney function**<br>- **minimizing treatment toxicity**<br>- **careful use of tacrolimus**<br>- **biopsy- and course-based reassessment**<br>rather than repeated indefinite escalation of nonspecific immunosuppression.<br><br>- In particular, given this case, I would be **cautious about repeated empiric steroid pulses** unless biopsy findings or the clinical pattern clearly support a reversible immune-mediated relapse.<br><br>#### Changes from the initial version<br>- Clarified that **worsening creatinine or oliguria requires urgent reassessment even if proteinuria is unchanged**, reflecting this patient's actual course.<br>- More strongly emphasized **hospitalization/supportive care thresholds** for recurrent severe edema, oliguria, and AKI.<br>- Incorporated the lesson that **renal recovery may occur without proteinuria remission**.<br>- Made the role of the **TRPC6 finding** more explicit: it shifts future management toward **kidney-function preservation and toxicity minimization**, rather than repeated escalation of empiric immunosuppression.<br>- Reinforced that **abrupt CNI reduction is a major relapse trigger** in this patient and should be avoided. |

| **Case 6: Takayasu arteritis-associated refractory hypertension induces nephrotic syndrome through glomerular microangiopathy** [14] | |
|---|---|
| **Phase I – Q1: Summarizing medical problems** | |
| Baseline LLM (Model A) | ## 1. Problem representation<br>A 69-year-old woman with a **4-year progressive course of proteinuria and microscopic hematuria**, now presenting with **nephrotic syndrome** (**proteinuria 5.0 g/gCr, albumin 1.8 g/dL, massive leg edema**) and **longstanding difficult-to-control hypertension**, with **mild rise in creatinine (0.67 to 0.89 mg/dL)**. The key features are **older age, nephrotic-range proteinuria, hematuria, |

| | |
|---|---|
| | hypertension, and gradual renal dysfunction**, suggesting an **underlying glomerular disease**, possibly chronic and progressive. |
| With Static RAG (Model B) | ## 1. Problem representation<br>A 69-year-old woman with a long history of severe hypertension and chronic urinary abnormalities now presents with **progressive nephrotic syndrome** characterized by **massive proteinuria (5.0 g/gCr, low selectivity), severe leg edema, hypoalbuminemia (1.8 g/dL)**, and **microscopic hematuria**, with a **mild rise in serum creatinine (0.67 to 0.89 mg/dL)** over several years. The course is chronic and progressive, suggesting an **underlying glomerular disease**, especially one associated with both **nephrotic-range proteinuria and hematuria** rather than isolated minimal-change-type disease. Her long-standing hypertension may be either a contributor to kidney injury or a consequence of the renal disease. |
| With Static RAG and Mult-agent framework (Model C) | ## 1. Problem representation<br>A **69-year-old woman** with a **4-year progressive course of proteinuria and microscopic hematuria**, longstanding **severe hypertension**, and mild rise in creatinine (**0.67 → 0.89 mg/dL**) now presents with **nephrotic syndrome**: **massive proteinuria** (urine protein 5.0 g/gCr, **nonselective** with selectivity index 0.33), **severe hypoalbuminemia** (1.8 g/dL), **marked leg edema**, and **general malaise**. She also had **10 kg weight loss** over the past year. This is best framed as **nephrotic syndrome with active urinary sediment in an older adult**, suggesting an underlying **structural glomerular disease** rather than minimal change disease or an isolated hemodynamic cause.<br><br>**Key features**<br>- Older adult with **nephrotic syndrome + microscopic hematuria + hypertension**<br>- **Chronic progressive** course, not acute<br>- **Nonselective proteinuria**, supporting organic glomerular disease<br>- Weight loss may reflect **malignancy/systemic disease** or severe chronic illness from nephrotic syndrome itself<br>- Prior optic neuritis suggests possible past immune disease, but its current relevance is **uncertain** and should not be overinterpreted |
| **Phase I – Q2: Differential diagnoses and rationale** | |
| Baseline LLM (Model A) | ## 2. Differential diagnosis<br>### Top 3 differentials<br>1. **Membranoproliferative glomerulonephritis (MPGN) / C3 glomerulopathy**<br> - Fits the combination of **proteinuria, hematuria, hypertension, and slowly progressive kidney dysfunction**.<br> - Particularly important if there are complement abnormalities or chronic active urinary findings.<br><br>2. **Membranous nephropathy**<br> - Common in older adults with nephrotic syndrome.<br> - However, **persistent microscopic hematuria and hypertension** may suggest a more inflammatory glomerular process than pure membranous nephropathy.<br><br>3. **Focal segmental glomerulosclerosis (FSGS)**<br> - Can cause **nonselective proteinuria, hypertension, and progressive renal dysfunction**.<br> - May be primary or secondary.<br><br>### Most likely |

| | |
|---|---|
| | **A chronic glomerulonephritis with nephrotic syndrome, especially MPGN/C3 glomerulopathy**, because the patient has **both nephrotic and nephritic features**: heavy proteinuria, hematuria, hypertension, and gradual kidney dysfunction.<br><br>### Must not be missed<br>**Secondary causes of nephrotic syndrome/glomerulonephritis**, especially:<br>- **Lupus nephritis or other autoimmune disease**<br>- **Infection-related disease** (hepatitis B, hepatitis C)<br>- **Malignancy-associated membranous nephropathy**<br>These are important because they change treatment and prognosis substantially. |
| With Static RAG (Model B) | ## 2. Differential diagnosis<br>### Top 3 differential diagnoses<br>1. **Membranous nephropathy**<br>- Common in older adults presenting with nephrotic syndrome.<br>- Can cause heavy proteinuria with relatively preserved renal function early on.<br>- Microscopic hematuria may occur.<br>- Low-selectivity proteinuria is compatible.<br><br>2. **Focal segmental glomerulosclerosis (FSGS)**<br>- Causes nonselective nephrotic-range proteinuria, hypertension, hematuria, and progressive kidney dysfunction.<br>- Fits the chronic course and gradual creatinine rise.<br>- Could be primary or secondary, including from long-standing hypertension.<br><br>3. **Membranoproliferative glomerulonephritis (MPGN) / C3 glomerulopathy**<br>- Considered because nephrotic syndrome plus hematuria and hypertension suggests a mixed nephritic-nephrotic pattern.<br>- Chronic progressive course is compatible.<br><br>### Most likely<br>**Membranous nephropathy or FSGS** are most likely from the available data; if forced to choose one, **FSGS** is slightly favored because of the combination of **long-standing hypertension, nonselective heavy proteinuria, hematuria, and gradual decline in kidney function**.<br><br>### Must not be missed<br>**Renal amyloidosis or malignancy-associated secondary membranous nephropathy**<br>- In an older patient with weight loss and nephrotic syndrome, a secondary cause must be excluded.<br>- Missing these would delay treatment of an underlying systemic disease or cancer. |
| With Static RAG and Mult-agent framework (Model C) | ## 2. Prioritized differential diagnosis<br>Because nephrotic syndrome is accompanied by hematuria and hypertension, the differential should remain **broad** until biopsy and serologies are available.<br><br>### Highest-priority diagnoses<br>1. **Membranous nephropathy**<br>2. **FSGS / hypertensive glomerular injury**<br>3. **Immune-complex glomerulonephritis**, including **MPGN**<br>4. **Monoclonal gammopathy–associated renal disease**<br>5. **Amyloidosis** |

| | |
|---|---|
| | ### Why these are prioritized<br>- **Membranous nephropathy**: common in older adults with heavy proteinuria and edema, sometimes with hematuria; may be **primary or secondary**.<br>- **FSGS / hypertensive glomerular injury**: longstanding severe hypertension, chronic proteinuria, hematuria, and slow creatinine rise are compatible; however, marked hypoalbuminemia and full nephrotic syndrome suggest more than uncomplicated hypertensive nephrosclerosis.<br>- **Immune-complex GN / MPGN**: hematuria plus proteinuria plus hypertension fits well, especially if complements are low or if infection/autoimmunity is present.<br>- **Monoclonal gammopathy–associated disease**: important in older adults with nephrotic syndrome and hematuria; may mimic membranous or MPGN patterns.<br>- **Amyloidosis**: can cause heavy proteinuria and edema in older adults, though persistent hematuria is less classic.<br><br>### Must not miss<br>**Secondary glomerular disease from malignancy or monoclonal gammopathy**<br><br>**Why**<br>- Age 69 with weight loss and malaise<br>- Membranous nephropathy can be malignancy-associated<br>- Plasma cell or other monoclonal disorders are important causes of nephrotic syndrome in older adults<br><br>### Less likely but worth considering if supported by history/labs<br>- **Lupus nephritis or other connective tissue disease**<br>- **Diabetic kidney disease**<br>- **Infection-related glomerular disease** (HBV, HCV, HIV)<br><br>**Change from initial impression:**<br>I broadened the differential beyond membranous nephropathy and gave more explicit weight to **monoclonal gammopathy-related disease** and **amyloidosis**, while avoiding making membranous nephropathy appear dominant before biopsy. |
| **Phase I – Q3: Necessary physical examinations and reasons** | |
| Baseline LLM (Model A) | ## 3. Additional history and physical examination<br>### Additional questions<br>- **Systemic symptoms**: fever, rash, arthralgia, oral ulcers, sicca symptoms, Raynaud phenomenon<br> - To assess for autoimmune disease such as SLE or vasculitis.<br>- **Infectious history**: hepatitis risk, chronic infections<br> - To evaluate secondary MPGN or membranous nephropathy.<br>- **Malignancy-related symptoms**: weight loss pattern, appetite loss, night sweats, cancer screening history<br> - Because older age and nephrotic syndrome raise concern for malignancy-associated disease.<br>- **Medication history**: NSAIDs, herbal drugs, recent antibiotics<br> - Some drugs can cause nephrotic syndrome/interstitial or glomerular disease.<br>- **Neurologic/ophthalmologic history** in more detail<br> - To clarify whether prior optic neuritis suggests a broader autoimmune disorder.<br><br>### Physical examination<br>- **Volume status**: severity of edema, pleural effusion/ascites, jugular venous pressure<br> - To assess nephrotic syndrome severity and guide diuretic/antihypertensive |

| | |
|---|---|
|  | management.<br>- **Blood pressure in detail**<br> - To confirm severity and possible target organ risk.<br>- **Skin/joints/mucosa**<br> - To look for autoimmune clues.<br>- **Lymph nodes, breast, abdomen**<br> - To screen for occult malignancy.<br>- **Fundoscopy and cardiovascular exam**<br> - To assess hypertensive end-organ damage. |
| With Static RAG (Model B) | ## 3. Additional history and physical examination<br>### Additional questions<br>- **Time course of edema, foamy urine, oliguria**<br> - To assess severity and progression of nephrotic syndrome.<br>- **Systemic symptoms**: fever, rash, arthralgia, oral ulcers, sicca symptoms, neuropathy<br> - To screen for autoimmune or systemic inflammatory disease.<br>- **Infection history**: hepatitis B/C risk, recurrent infections<br> - To evaluate secondary causes of glomerulopathy.<br>- **Malignancy symptoms**: appetite loss, night sweats, bleeding, breast/gynecologic/GI symptoms<br> - Important given age and weight loss.<br>- **Drug history**: NSAIDs, bisphosphonates, herbal drugs, interferon, lithium<br> - For secondary membranous nephropathy or secondary FSGS/minimal change disease.<br>- **Past autoimmune/neurologic history details**, especially optic neuritis and any relapse<br> - To explore systemic immune disease.<br>- **Family history of kidney disease**<br> - To assess hereditary nephropathy.<br><br>### Physical examination<br>- **Volume status**: edema distribution, JVP, pleural effusion, ascites<br> - To determine severity of fluid overload.<br>- **Blood pressure in both arms, funduscopic exam**<br> - To assess chronic hypertensive damage.<br>- **Skin/joints/oral cavity**<br> - For connective tissue disease or vasculitis clues.<br>- **Cardiopulmonary exam**<br> - To detect heart failure or serositis.<br>- **Abdominal exam and lymph nodes**<br> - To screen for malignancy/systemic disease.<br>- **Neurologic exam**<br> - If amyloidosis or systemic immune disease is suspected. |
| With Static RAG and Mult-agent framework (Model C) | ## 3. Focused additional history and physical examination<br><br>### High-yield history<br>1. **Systemic symptoms**<br> - Fever, night sweats, appetite loss, progression of weight loss<br> - To assess malignancy, chronic infection, inflammatory disease<br><br>2. **Medication timeline**<br> - NSAIDs, herbal drugs, antibiotics, supplements, prior long-term medications |

| | |
|---|---|
| | - To identify drug-related nephrotic syndrome or GN<br><br>3. **Diabetes and vascular history**<br>- Diabetes, retinopathy, cardiovascular disease<br>- To assess diabetic kidney disease and chronic vascular renal injury<br><br>4. **Autoimmune symptoms**<br>- Rash, oral ulcers, photosensitivity, arthralgia, sicca, Raynaud phenomenon, neuropathy<br>- To evaluate lupus, Sjögren syndrome, vasculitis, other immune diseases<br><br>5. **Infection risk**<br>- Hepatitis exposure, transfusions, prior chronic infections<br>- To assess secondary membranous nephropathy/MPGN<br><br>6. **Malignancy history and screening**<br>- Prior cancer, breast/GI/gynecologic symptoms, screening status<br>- To assess secondary nephrotic syndrome in an older adult with weight loss<br><br>7. **Thrombotic symptoms**<br>- Sudden dyspnea, pleuritic pain, unilateral leg swelling, flank pain<br>- Nephrotic syndrome increases venous thromboembolism risk<br><br>8. **Family history**<br>- Kidney disease, autoimmune disease, plasma cell disorders<br>- To detect inherited or systemic predisposition<br><br>### Focused physical examination<br>1. **Volume and edema assessment**<br>- Distribution of edema, JVP, lung crackles, ascites, orthostatic vitals<br>- To distinguish total body fluid overload from intravascular depletion<br><br>2. **Blood pressure and chronic end-organ effects**<br>- Repeated BP measurement, fundoscopy if available<br>- To assess severity of chronic hypertension<br><br>3. **Thrombosis exam**<br>- Asymmetric leg swelling, calf tenderness, signs of PE<br>- Important complication of nephrotic syndrome<br><br>4. **General/systemic exam**<br>- Lymph nodes, abdominal masses, cachexia, skin purpura/rash, joint swelling<br>- To look for malignancy, systemic inflammatory disease, vasculitis<br><br>**Change from initial impression:**<br>I prioritized **diabetes history, medication chronology, family history, thrombotic symptoms, edema distribution, and intravascular volume status**, as these are higher yield for this presentation. |
| **Phase I – Q4: Plan for investigations/tests** | |
| Baseline LLM (Model A) | ## 4. Tests to order<br>### Essential laboratory tests<br>1. **Urinalysis with sediment**<br>- Purpose: confirm hematuria, dysmorphic RBCs, casts. |

| | |
|---|---|
| | - Expected: active urinary sediment supporting glomerular disease.<br><br>2. **Quantification of proteinuria**<br>- Purpose: confirm nephrotic-range proteinuria and monitor severity.<br>- Expected: persistent heavy proteinuria.<br><br>3. **Basic blood tests**: CBC, CMP, creatinine, albumin, lipids<br>- Purpose: assess nephrotic syndrome severity, kidney function, complications.<br>- Expected: hypoalbuminemia, possibly hyperlipidemia, mild renal impairment.<br><br>4. **Immunologic workup**: C3, C4, CH50, ANA, anti-dsDNA, ANCA, anti-GBM, serum immunoglobulins<br>- Purpose: evaluate complement-mediated or autoimmune glomerulonephritis.<br>- Expected: low complement would support MPGN/C3-related disease or lupus.<br><br>5. **Infection screening**: hepatitis B, hepatitis C, HIV<br>- Purpose: identify secondary causes of glomerular disease.<br>- Expected: may be negative, but must be checked.<br><br>6. **Monoclonal protein evaluation**: serum/urine protein electrophoresis, immunofixation, serum free light chains<br>- Purpose: assess for monoclonal gammopathy-related renal disease in an older patient.<br>- Expected: usually negative, but important to exclude.<br><br>7. **Anti-PLA2R antibody**<br>- Purpose: assess for primary membranous nephropathy.<br>- Expected: positive would support membranous nephropathy.<br><br>### Imaging<br>8. **Renal ultrasound**<br>- Purpose: assess kidney size/chronicity and exclude structural disease.<br>- Expected: usually preserved kidney size in glomerular disease unless advanced chronic damage.<br><br>9. **Age-appropriate malignancy screening / CT if indicated**<br>- Purpose: exclude malignancy-associated nephrotic syndrome.<br>- Expected: depends on history and baseline screening status.<br><br>### Definitive test<br>10. **Kidney biopsy**<br>- Purpose: establish the exact glomerular diagnosis and guide treatment.<br>- Expected: necessary because the differential includes MPGN/C3 glomerulopathy, membranous nephropathy, FSGS, and secondary causes, which cannot be reliably distinguished clinically alone.<br><br>Overall, the most appropriate next step to narrow the diagnosis is a **comprehensive serologic evaluation followed by kidney biopsy**, assuming no contraindication. |
| With Static RAG (Model B) | ## 4. Tests to narrow the diagnosis<br>### Core laboratory tests<br>1. **Urinalysis with sediment and urine protein quantification**<br>- Purpose: confirm proteinuria severity and characterize hematuria/casts.<br>- Expected: persistent heavy proteinuria, microscopic hematuria; dysmorphic |

| | |
|---|---|
| | RBCs/casts would support glomerular disease.<br><br>2. **Serum chemistry: creatinine, eGFR, albumin, total protein, lipids, electrolytes**<br>- Purpose: assess nephrotic syndrome severity and kidney function.<br>- Expected: hypoalbuminemia, hyperlipidemia, mild renal dysfunction.<br><br>3. **Immunologic and secondary-cause workup**<br>- **ANA, anti-dsDNA, complements (C3/C4), ANCA, anti-GBM**<br>- **HBsAg, anti-HCV, HIV**<br>- **SPEP/UPEP, serum free light chains**<br>- **PLA2R antibody**<br>- Purpose: identify lupus, complement-mediated disease, vasculitis, infection-related disease, monoclonal gammopathy/amyloidosis, and primary membranous nephropathy.<br>- Expected:<br>- PLA2R positivity would support primary membranous nephropathy.<br>- Low complement would favor MPGN/lupus.<br>- Monoclonal protein would raise concern for AL amyloidosis or MGRS.<br><br>### Imaging<br>4. **Renal ultrasound**<br>- Purpose: assess kidney size, chronicity, and exclude obstructive/urologic disease.<br>- Expected: usually normal-sized kidneys in glomerular disease.<br><br>5. **Age-appropriate malignancy screening / targeted CT if indicated**<br>- Purpose: evaluate secondary membranous nephropathy, amyloidosis, or occult cancer given weight loss.<br>- Expected: often negative, but important not to miss occult malignancy.<br><br>### Definitive test<br>6. **Kidney biopsy**<br>- Purpose: establish the exact glomerular diagnosis and guide therapy.<br>- Expected:<br>- **Membranous nephropathy**: subepithelial deposits, GBM spike formation.<br>- **FSGS**: segmental sclerosis/podocyte injury.<br>- **MPGN**: mesangial/endocapillary proliferation with double contours.<br>- **Amyloidosis**: Congo-red-positive deposits.<br><br>Overall, the diagnostic priority is to **confirm the glomerular lesion by kidney biopsy while simultaneously excluding secondary causes**, especially **malignancy, monoclonal gammopathy/amyloidosis, autoimmune disease, and chronic infection**. |
| With Static RAG and Mult-agent framework (Model C) | ## 4. Testing plan to narrow the diagnosis<br><br>### Essential tests before or alongside biopsy<br>1. **Urinalysis with microscopy**<br>- Confirm hematuria; look for dysmorphic RBCs and casts<br>- Helps establish glomerular origin<br><br>2. **Protein quantification**<br>- Spot urine protein/Cr or 24-hour urine protein<br>- Confirms severity and provides baseline<br><br>3. **Basic blood tests** |

- CBC, creatinine, electrolytes, albumin, liver panel, lipid profile
  - Assess nephrotic burden and kidney function

4. **Serologic workup for secondary causes**
  - ANA, anti-dsDNA, C3/C4
  - ANCA/anti-GBM only if sediment or course suggests more inflammatory GN
  - Helps identify immune-complex/systemic disease

5. **Infection screening**
  - HBsAg, anti-HBc, HCV Ab, HIV
  - Important causes of secondary nephrotic syndrome and MPGN

6. **Monoclonal protein evaluation**
  - **SPEP/UPEP with immunofixation** and **serum free light chains**
  - High-yield in an older adult with nephrotic syndrome

7. **PLA2R antibody** (± THSD7A if available)
  - Useful if membranous nephropathy is suspected
  - But does not replace biopsy here

8. **Renal ultrasound**
  - Kidney size, chronicity, structural disease, exclusion of obstruction

#### Definitive next step
9. **Kidney biopsy**
  - This is especially important because **nephrotic syndrome with hematuria and nonselective proteinuria cannot be reliably classified clinically**
  - It will distinguish membranous nephropathy, FSGS, immune-complex GN, amyloidosis, and paraprotein-related disease

#### Malignancy evaluation
10. **Age-appropriate cancer screening plus targeted workup for weight loss**
  - Routine screening plus targeted chest/abdominal/pelvic evaluation as clinically indicated
  - Best guided by history, exam, basic labs, and biopsy pattern

**Change from initial impression:**
I more clearly separated **high-yield pre-biopsy testing** from broader evaluation and emphasized that **prompt kidney biopsy** is the key diagnostic step in this case.

---

### Bottom line
This is a **69-year-old woman with chronic progressive nephrotic syndrome, microscopic hematuria, and severe hypertension**, most consistent with an **underlying structural glomerular disease**. The differential should remain broad and include **membranous nephropathy, FSGS/hypertensive glomerular injury, immune-complex GN/MPGN, monoclonal gammopathy-related disease, and amyloidosis**. The most important next steps are **urine microscopy, targeted serologic/infectious/paraprotein testing, renal ultrasound, and prompt kidney biopsy**.

### Summary of changes from the initial version
- Reframed the case as **"nephrotic syndrome with active urinary sediment in an older

| | |
|---|---|
| | adult.”**<br>- Reduced emphasis on **optic neuritis** as evidence of current autoimmune disease.<br>- Expanded the differential to explicitly include **monoclonal gammopathy-related disease** and **amyloidosis**.<br>- Avoided making **membranous nephropathy** seem too dominant before biopsy.<br>- Focused history/physical exam more on **diabetes, medication timeline, family history, thrombosis, edema distribution, and intravascular volume status**.<br>- Clarified that **kidney biopsy is essential** because the syndrome cannot be classified confidently on clinical grounds alone. |
| **Phase II – Q5: Interpreting test results** | |
| Baseline LLM (Model A) | **5. Interpretation of the test results**<br><br>- **Inflammatory large-vessel disease is strongly suggested.**<br>The combination of **headache, neck pain, refractory hypertension, >15 mmHg inter-arm systolic BP difference, severe aortic stenotic/arterial wall lesions on contrast CT, left subclavian wall thickening, and carotid intima-media thickening** indicates **large-vessel arteritis involving the aorta and its branches** rather than isolated essential hypertension.<br><br>- **Markedly elevated ESR (129 mm/h) with only mildly elevated CRP (1.17 mg/dL)** is still compatible with chronic inflammatory vasculitis.<br>ESR can be very high in active large-vessel arteritis and may also be accentuated by **hypoalbuminemia/nephrotic syndrome**. The inflammatory signal is therefore significant even though CRP is only modestly elevated.<br><br>- **Severe aortic regurgitation (AR)** is a major clue.<br>In the context of aortic wall disease, AR suggests **aortic root/ascending aortic involvement due to aortitis**, which also explains the **very wide pulse pressure (103 mmHg)** and contributes to systolic hypertension.<br><br>- **Normal renin/aldosterone/catecholamines** make endocrine causes of severe hypertension less likely.<br>Her hypertension is better explained by **vascular disease (large-vessel stenosis), aortic regurgitation, and renal disease/nephrotic syndrome**.<br><br>- **Negative ANCA and anti-GBM antibodies** argue against small-vessel ANCA-associated vasculitis and anti-GBM disease as the main unifying diagnosis.<br>This shifts the focus away from pauci-immune crescentic GN and toward **large-vessel vasculitis with a separate or secondary glomerular lesion**.<br><br>- **Normal complement (C3/C4)** makes classic immune-complex hypocomplementemic diseases such as typical MPGN or active lupus nephritis less likely based on the provided data.<br><br>- **Albumin 2.4 g/dL with preserved creatinine (0.93 mg/dL, eGFR 46.2)** supports **ongoing nephrotic syndrome** with relatively preserved renal function.<br>The nephrotic state explains her edema and may also contribute to elevated ESR and thrombotic risk.<br><br>- **Fibrinogen 1033 mg/dL, mildly elevated FDP/D-dimer** support an inflammatory and hypercoagulable state, which is relevant in nephrotic syndrome and systemic vasculitis. |

| | |
|---|---|
| With Static RAG (Model B) | ### 5. Interpretation of the test results<br>- **Large-vessel findings are the most important clue**: the **>15 mmHg systolic BP difference between arms**, **severe aortic stenosis/wall thickening of the left subclavian artery on contrast CT**, and **increased carotid intima-media thickness** strongly indicate **large-vessel vasculitis** rather than essential hypertension alone. In this patient, these vascular lesions plausibly explain the **refractory hypertension, headache, and neck pain**.<br>- **Severe aortic regurgitation (AR)** is highly significant: it can occur from **aortic root/aortic wall inflammation** in large-vessel vasculitis. The **very wide pulse pressure (103 mmHg)** is consistent with severe AR and helps explain the prominent systolic hypertension.<br>- **Inflammatory markers support active systemic inflammation**: **ESR 129 mm/h** is markedly elevated and more striking than the only mildly elevated **CRP 1.17 mg/dL**. This pattern is compatible with chronic inflammatory vasculitis.<br>- **ANCA and anti-GBM negative**: these results make **ANCA-associated vasculitis** and **anti-GBM disease** unlikely causes of the renal findings and systemic symptoms.<br>- **Normal complement (C3/C4)** argues against immune-complex diseases such as active lupus nephritis or some forms of MPGN as the main unifying diagnosis.<br>- **Albumin 2.4 g/dL with preserved Cr 0.93 mg/dL/eGFR 46.2** supports ongoing **nephrotic syndrome** with relatively preserved kidney function. Thus, the kidney disease appears significant but is not sufficient to explain the major vascular abnormalities.<br>- **Fibrinogen 1033 mg/dL and mild FDP/D-dimer elevation** are consistent with an inflammatory/prothrombotic state, which fits both **active vasculitis** and **nephrotic syndrome**.<br>- **No fragmented RBCs** argues against thrombotic microangiopathy as the main cause of hypertension/renal injury.<br>- The absence of classic cranial symptoms of giant cell arteritis (**temporal headache, scalp tenderness, jaw claudication**) makes cranial GCA less typical, but **does not exclude large-vessel GCA**. |
| With Static RAG and Mult-agent framework (Model C) | ## 5. Interpretation of the test results<br><br>- **The new dominant finding is large-vessel disease.**<br>The combination of a **>15 mmHg systolic BP difference between the arms**, **CT evidence of severe aortic stenotic/narrowing change**, **wall thickening of the left subclavian artery**, and **carotid intima-media thickening** strongly supports **large-vessel vasculitis** rather than essential hypertension alone. These vascular abnormalities provide a plausible explanation for her **refractory hypertension, headache, and neck pain**.<br><br>- **Severe aortic regurgitation is a major clinical consequence of the vascular process.**<br>Echocardiography showed **severe AR** with a **very wide pulse pressure (103 mmHg)**. This suggests clinically important involvement of the **aortic root/ascending aorta or valve apparatus** in the inflammatory process. AR itself can further raise systolic pressure and worsen symptoms.<br><br>- **Inflammatory markers support active systemic inflammation.**<br>**ESR 129 mm/h** is markedly elevated and is highly supportive of an inflammatory large-vessel vasculitic process. **CRP 1.17 mg/dL** is only mildly elevated, but this does not exclude active disease. **Fibrinogen 1033 mg/dL** also supports inflammation and likely contributes to the very high ESR. |

| | |
|---|---|
| | - **The serologic pattern argues against some small-vessel causes, but does not define the renal lesion.**<br>**ANCA negative** and **anti-GBM negative** make ANCA-associated vasculitis and anti-GBM disease less likely as the main unifying diagnosis. **Normal C3/C4** makes a classic hypocomplementemic immune-complex process less likely. However, these results **narrow the differential only**; they do **not establish the cause of the nephrotic syndrome**.<br><br>- **The absence of classic cranial GCA symptoms is helpful but not exclusionary.**<br>She does not have temporal headache, scalp tenderness, or jaw claudication, so **classic cranial GCA is not strongly supported clinically**. However, in a **69-year-old woman** with markedly elevated ESR and aortic/subclavian/carotid involvement, **large-vessel GCA remains a major possibility**.<br><br>- **The renal findings confirm nephrotic syndrome, but the renal pathology remains unresolved.**<br>**Albumin 2.4 g/dL** supports ongoing significant urinary protein loss. **Cr 0.93 mg/dL / eGFR 46.2 mL/min/1.73 m$^2$** indicates only mild-to-moderate renal impairment despite heavy proteinuria. This is compatible with glomerular disease, but the available data do **not** prove that the nephrotic syndrome is secondary to vasculitis. The **renal lesion remains unresolved and still requires separate diagnostic confirmation**, ideally by kidney biopsy if feasible.<br><br>- **Coagulation markers indicate increased thrombotic risk.**<br>Markedly elevated **fibrinogen** with mildly elevated **FDP/D-dimer** suggests a **prothrombotic state**, which is relevant because nephrotic syndrome itself increases thrombosis risk.<br><br>**Change from my initial interpretation:**<br>I now more clearly **separate the large-vessel vasculitis findings from the renal diagnosis**, avoid implying that the nephrotic syndrome is already proven to be vasculitic, and interpret "severe aortic stenosis" more cautiously as **aortic stenotic/narrowing change** rather than definite valvular aortic stenosis. |
| **Phase II – Q6: Reconsidering the differential diagnosis** | |
| Baseline LLM (Model A) | **6. Updated diagnostic thinking / Working diagnosis**<br><br>The differential now changes substantially. A primary isolated glomerular disease is no longer the leading overall explanation.<br><br>- The **most likely systemic diagnosis is large-vessel vasculitis**, specifically **Takayasu arteritis**, because of:<br>- aortic and subclavian involvement,<br>- inter-arm BP difference,<br>- carotid wall thickening,<br>- refractory hypertension,<br>- aortic regurgitation from aortitis,<br>- markedly elevated ESR,<br>- absence of evidence for ANCA/anti-GBM small-vessel disease.<br><br>- **Giant cell arteritis remains a differential**, especially given age 69, but is less favored from the provided information because the classic cranial features are absent and the distribution described is highly compatible with Takayasu-type large-vessel |

| | |
|---|---|
| | disease. Age alone does not outweigh the vascular pattern.<br><br>- For the kidney disease, the current information supports **nephrotic syndrome associated with an underlying glomerulopathy**, but the exact renal histology remains unresolved.<br>Based on the systemic picture, the best working diagnosis at this point is:<br><br>**Working diagnosis: Large-vessel vasculitis, most likely Takayasu arteritis, with aortic/subclavian/carotid involvement causing refractory hypertension and severe AR, accompanied by nephrotic syndrome due to an associated glomerular disease (histology not yet defined).** |
| With Static RAG (Model B) | ### 6. Updated diagnostic thinking / Working diagnosis<br>The differential diagnosis shifts substantially from a primary isolated glomerular disease to a **systemic large-vessel vasculitis with renal involvement**.<br><br>**Most likely working diagnosis:**<br>**Large-vessel giant cell arteritis (large-vessel GCA), causing aortitis with left subclavian involvement and severe aortic regurgitation, associated with nephrotic syndrome likely secondary to vasculitis-related glomerular disease.**<br><br>#### Why this is now most likely<br>- Age **69 years** strongly favors **GCA over Takayasu arteritis**.<br>- Imaging shows **aortic and major branch vessel inflammation/stenotic change**, which is characteristic of **large-vessel vasculitis**.<br>- **Marked ESR elevation**, headache/neck pain, and arm BP asymmetry support this.<br>- **Severe AR** is a recognized complication of **aortitis**.<br>- Lack of typical temporal artery symptoms does **not** exclude **large-vessel GCA**, which may present predominantly with extracranial large-artery disease.<br><br>#### Revised prioritization of differentials<br>1. **Large-vessel GCA** – most likely.<br>2. **Takayasu arteritis** – possible from the vascular pattern, but less likely because of age.<br>3. **Other aortitis** (infectious or IgG4-related disease) – less likely from the given data, but should remain in the broader differential if pathology is atypical.<br><br>Regarding the kidney disease, the nephrotic syndrome may still require biopsy for precise classification, but it is now more likely to be **secondary and linked to the systemic inflammatory disease** rather than a completely separate primary nephropathy. |
| With Static RAG and Mult-agent framework (Model C) | ## 6. Updated diagnostic thinking / revised differential diagnosis<br><br>The new data substantially change the case. The central process is now best understood as **large-vessel vasculitis with major cardiovascular involvement**, based on:<br><br>- refractory hypertension,<br>- inter-arm BP difference,<br>- aortic/subclavian/carotid abnormalities,<br>- severe aortic regurgitation,<br>- markedly elevated ESR. |

| | |
|---|---|
| | ### Most likely working diagnosis<br>**Large-vessel vasculitis, probably large-vessel giant cell arteritis versus Takayasu arteritis, with aortic involvement causing severe aortic regurgitation and difficult-to-control systolic hypertension.**<br><br>### Why this revision is more appropriate<br>- The vascular distribution clearly supports **large-vessel vasculitis**.<br>- However, in a **69-year-old woman**, **large-vessel GCA is favored over Takayasu arteritis by age alone**.<br>- The absence of cranial symptoms does **not exclude GCA**, because GCA can present predominantly as **large-vessel disease**.<br>- Takayasu arteritis remains in the differential because of the **aorta/subclavian/carotid pattern** and inter-arm BP discrepancy, but it is less strongly favored given age.<br><br>### Revised differential diagnosis<br>1. **Large-vessel giant cell arteritis** – most likely<br>2. **Takayasu arteritis** – important alternative<br>3. **Separate primary glomerular disease** causing nephrotic syndrome (e.g., membranous nephropathy, FSGS, immune-complex disease, monoclonal gammopathy-related disease)<br>4. Other systemic inflammatory disease with large-vessel involvement – less likely from current data<br><br>### Renal diagnosis: keep separate<br>The nephrotic syndrome may be related to the systemic inflammatory illness, but this is **not established**. At present, the renal disease should still be treated as an **unresolved glomerular process requiring histologic clarification**.<br><br>**Change from my initial diagnostic impression:**<br>I revised the working diagnosis from **Takayasu arteritis as most likely** to **large-vessel vasculitis, probably large-vessel GCA versus Takayasu arteritis**, because the patient's **age strongly favors GCA** and the absence of cranial symptoms does not rule it out. I also removed the unsupported claim of **vasculitis-associated glomerular involvement** and avoided calling the hypertension definitively renovascular. |
| **Phase II – Q7: Treatment planning** | |
| Baseline LLM (Model A) | **7. Treatment plan**<br><br>### A. Immediate treatment priorities<br>1. **Start systemic glucocorticoid therapy**<br>- Large-vessel vasculitis with major vascular involvement and severe AR requires prompt treatment.<br>- Standard approach: **high-dose prednisolone** (e.g., about **0.8–1 mg/kg/day**) unless contraindicated.<br>- Rationale: suppress vascular inflammation, prevent further arterial damage, and possibly improve constitutional/inflammatory findings.<br><br>2. **Strict blood pressure control**<br>- Continue/intensify antihypertensive therapy, especially agents useful in proteinuric kidney disease such as an **ARB/ACE inhibitor** if tolerated.<br>- Add other agents as needed because BP is driven by **vascular stenosis, AR, and renal disease**.<br>- Rationale: reduces cardiovascular and renal injury. |

| | |
|---|---|
| | 3. **Manage nephrotic syndrome supportively**<br>- **Salt restriction**<br>- **Loop diuretic** for edema<br>- Continue **RAS blockade** if tolerated to reduce proteinuria<br>- Monitor renal function, potassium, volume status<br>- Rationale: symptom relief and kidney protection while defining renal pathology.<br><br>### B. Disease-defining next step<br>4. **Kidney biopsy**, if feasible and safe<br>- Needed to determine the cause of nephrotic syndrome and guide whether treatment beyond steroids is necessary.<br>- Rationale: the renal lesion could be membranous nephropathy, FSGS, amyloidosis, immune-complex GN, or another process associated with vasculitis.<br><br>### C. Organ-specific vascular/cardiac management<br>5. **Urgent cardiovascular/vascular evaluation**<br>- Severe **AR** and major aortic disease require close collaboration with cardiology/cardiovascular surgery.<br>- Rationale: medical control of inflammation alone may not be sufficient if valve dysfunction or critical stenotic lesions are hemodynamically significant.<br><br>### D. Steroid-sparing / alternative options<br>6. If disease is refractory, relapsing, or steroid toxicity is a concern:<br>- Consider **steroid-sparing immunosuppressive therapy** such as **methotrexate, azathioprine, mycophenolate mofetil**, or **tocilizumab** depending on response and final diagnostic confidence.<br>- Rationale: these are accepted options in large-vessel vasculitis to reduce relapse and cumulative steroid exposure.<br><br>### E. Supportive preventive care<br>7. Because prolonged glucocorticoids are likely:<br>- **Bone protection** (calcium/vitamin D ± osteoporosis prophylaxis)<br>- Infection risk monitoring<br>- Consider thrombosis risk carefully in the setting of nephrotic syndrome<br><br>**In summary:** the new data strongly shift the case toward **large-vessel vasculitis, most likely Takayasu arteritis**, with severe cardiovascular involvement. Immediate treatment should focus on **high-dose glucocorticoids, BP/volume control, and multidisciplinary assessment of the aortic valve/aorta**, while **kidney biopsy** is important to clarify the nephrotic syndrome and refine long-term therapy. |
| With Static RAG (Model B) | ### 7. Treatment plan<br>#### Main treatment<br>1. **Start systemic glucocorticoid therapy promptly**<br>- Example: **prednisolone 0.5–1 mg/kg/day**, adjusted to severity.<br>- Rationale: suspected **active large-vessel GCA with major organ involvement** (aorta/subclavian artery, severe AR, systemic inflammation) requires rapid suppression of vascular inflammation to prevent progression.<br><br>2. **Consider steroid pulse therapy first if disease is judged severe**<br>- Especially reasonable if there is concern for rapidly progressive vascular inflammation or critical ischemic complications.<br>- Rationale: she has major vessel disease and significant cardiovascular involvement. |

<table>
<tr><td></td><td>3. **Close cardiovascular management**<br>- **Cardiology/cardiovascular surgery consultation** for **severe AR**.<br>- Control afterload and BP carefully, but recognize that systolic hypertension is partly driven by **AR and large-vessel disease**.<br>- Rationale: AR may become a major determinant of prognosis and may require intervention if heart failure or hemodynamic compromise develops.<br><br>4. **Renal management**<br>- Continue/optimize **RAAS blockade** if tolerated for proteinuria reduction.<br>- Salt restriction and **diuretics** for edema.<br>- Monitor kidney function, albumin, and proteinuria.<br>- Rationale: nephrotic syndrome requires supportive care regardless of cause.<br><br>5. **Kidney biopsy**<br>- If clinically feasible after stabilization.<br>- Rationale: to define the exact glomerular lesion and determine whether additional immunosuppression beyond steroids is needed.<br><br>6. **Steroid-sparing therapy if needed**<br>- Consider **tocilizumab** or **methotrexate** in relapsing disease or when steroid toxicity is a concern.<br>- Rationale: these are accepted options in GCA, particularly for relapse prevention and steroid reduction.<br><br>#### Supportive / preventive measures<br>- **Antiplatelet therapy** may be considered depending on bleeding risk and vascular risk profile.<br>- **Bone and gastric protection** with glucocorticoids.<br>- Monitor for **infection, glucose intolerance, and fluid status**.<br>- Given nephrotic syndrome, assess **thrombotic risk** carefully.<br><br>#### Other options / alternatives<br>- If the course or pathology does not fit GCA, reassess for **Takayasu arteritis, IgG4-related disease, or infectious aortitis**, because treatment would differ, especially if infection is suspected.<br>- If renal biopsy identifies a specific glomerulopathy requiring additional therapy, treatment should be tailored accordingly.</td></tr>
<tr><td>With Static RAG and Mult-agent framework (Model C)</td><td>## 7. Treatment plan<br><br>### A. Immediate management of suspected active large-vessel vasculitis<br>1. **Start high-dose glucocorticoid therapy**<br>- Reasonable because there is strong evidence of **active large-vessel vasculitis** with major cardiovascular involvement<br>- Example: **prednisolone 0.8–1.0 mg/kg/day** or equivalent, depending on specialist judgment<br><br>2. **Consider intravenous steroid pulse therapy if disease is judged organ-threatening**<br>- This may be appropriate if there is rapidly progressive vascular inflammation, severe symptomatic disease, or hemodynamic compromise<br><br>### B. Specialist evaluation and disease mapping</td></tr>
</table>

3. **Urgent rheumatology/vasculitis and cardiology consultation**
  - Needed to classify the vasculitis more accurately and coordinate treatment

4. **Further vascular imaging-based assessment**
  - Define the full extent of aortic and branch-vessel disease and provide a baseline for response monitoring

#### C. Cardiovascular management
5. **Careful blood pressure control**
  - Hypertension may reflect **major arterial disease plus severe AR**, not simply essential hypertension
  - Antihypertensives should be adjusted carefully with attention to hemodynamics

6. **Urgent cardiology/cardiovascular surgery assessment for severe AR**
  - Severe AR can worsen systolic hypertension and lead to heart failure
  - Valve or aortic intervention may become necessary depending on symptoms, hemodynamics, and inflammatory control

#### D. Renal/nephrotic syndrome management
7. **Supportive nephrotic syndrome treatment**
  - Salt restriction
  - Diuretics for edema as tolerated
  - Consider RAAS blockade if blood pressure, renal perfusion, and overall hemodynamics permit
  - Monitor creatinine, albumin, edema, and proteinuria

8. **Evaluate thrombotic risk carefully**
  - Nephrotic syndrome plus systemic inflammation increases thrombosis risk
  - Anticoagulation should be individualized rather than automatic

9. **Kidney biopsy when safe and clinically appropriate**
  - Important because the renal lesion remains unproven
  - This should guide whether the nephrotic syndrome is a separate primary glomerular disease or part of a broader systemic process

#### E. Steroid-sparing therapy
10. **Add steroid-sparing immunosuppression only after classification and specialist input**
  - Options may include **tocilizumab, methotrexate, azathioprine**, or others depending on whether the final diagnosis is GCA or Takayasu arteritis
  - This decision should be guided by disease classification, severity, response to steroids, and comorbidity profile

#### Alternative considerations
- If the final classification favors **large-vessel GCA**, glucocorticoids remain first-line and **tocilizumab** may be an important adjunct.
- If the renal biopsy identifies a **separate primary glomerular disease**, treatment should be modified accordingly.
- Vascular or surgical intervention may be required for large-vessel complications, but timing depends on inflammation control and urgency.

**Change from my initial treatment plan:**
I kept **high-dose glucocorticoids** as the immediate priority, but I now place greater emphasis on **rheumatology/cardiology consultation, imaging-based disease

<table>
<tr><td></td><td>mapping, and kidney biopsy when safe**, and I frame **steroid-sparing therapy as dependent on final classification and specialist input** rather than assuming a specific vasculitic subtype or renal mechanism.<br><br>## Summary of changes from the initial results<br>- Shifted the leading diagnosis from **Takayasu arteritis** to **large-vessel vasculitis, probably large-vessel GCA versus Takayasu arteritis**<br>- More clearly emphasized that **age favors GCA**<br>- Clarified that lack of cranial symptoms **does not exclude GCA**<br>- Separated **large-vessel vasculitis** from the **unresolved renal diagnosis**<br>- Removed the unsupported statement that nephrotic syndrome is likely due to **vasculitic glomerular involvement**<br>- Interpreted the aortic finding more cautiously as **aortic stenotic/narrowing change**<br>- Made the treatment plan more dependent on **specialist input, imaging, and kidney biopsy** rather than premature disease-specific assumptions</td></tr>
<tr><td colspan="2">Phase III – Q8: Evaluation of treatment</td></tr>
<tr><td>Baseline LLM (Model A)</td><td>### 8. Monitoring effectiveness, safety, risks, and countermeasures<br><br>**A. Effectiveness monitoring**<br><br>1. **Takayasu arteritis activity**<br>- Monitor: **symptoms** (headache, neck pain, malaise), **BP in both arms**, pulse deficits, signs of heart failure.<br>- Labs: **CRP, ESR** at regular intervals.<br>- Imaging: periodic **vascular imaging** (CT/MR angiography and/or carotid ultrasound) to assess progression of aortic/subclavian/carotid lesions.<br>- Cardiac follow-up: **echocardiography** for severity of **aortic regurgitation** and cardiac function.<br><br>2. **Renal response**<br>- Monitor: **urine protein/Cr ratio**, urinalysis for **microhematuria**, **serum albumin**, body weight, edema.<br>- Kidney function: **serum creatinine, eGFR**, electrolytes.<br>- BP tracking is essential because the biopsy findings suggest **hypertension-related glomerular endothelial/podocyte injury**.<br><br>3. **Volume status / heart failure**<br>- Monitor: symptoms/signs of congestion, **chest X-ray or BNP/NT-proBNP if clinically needed**, daily weight early in treatment.<br><br>**B. Safety monitoring and major risks**<br><br>1. **Sacubitril/valsartan**<br>- Risks: **hypotension**, **decline in kidney function**, **hyperkalemia**.<br>- Monitoring: BP, creatinine/eGFR, potassium shortly after initiation/dose changes and periodically thereafter.<br>- Countermeasures: reduce dose or hold drug if symptomatic hypotension, significant creatinine rise, or hyperkalemia occurs.<br><br>2. **Prednisolone**<br>- Risks: **infection**, hyperglycemia, osteoporosis, muscle weakness, psychiatric effects, fluid retention.</td></tr>
</table>

| | |
|---|---|
| | - Monitoring: temperature/symptoms of infection, CBC, glucose, BP, weight; long-term bone protection assessment.<br>- Countermeasures: use the **lowest effective dose**, taper when disease control allows, provide **osteoporosis prophylaxis** and infection vigilance.<br><br>3. **Tocilizumab**<br>- Risks: **serious infection**, neutropenia, liver dysfunction, lipid elevation; CRP may be suppressed despite infection.<br>- Monitoring: CBC, liver enzymes, lipid profile, careful clinical assessment for infection.<br>- Countermeasures: educate to report fever or new symptoms promptly; do not rely on CRP alone to exclude infection.<br><br>4. **Persistent vascular/cardiac complications of TAK**<br>- Risks: progression of **aortic stenotic lesions**, worsening **aortic regurgitation**, heart failure, ischemic complications.<br>- Monitoring: serial vascular imaging and echocardiography.<br>- Countermeasures: early cardiology/cardiovascular surgery consultation if hemodynamics worsen.<br><br>5. **Nephrotic syndrome complications**<br>- Risks: **thrombosis**, infection, worsening edema.<br>- Monitoring: clinical signs of venous/arterial thrombosis, albumin level, edema, mobility status.<br>- Countermeasures: prompt evaluation if thrombosis is suspected; optimize remission of proteinuria and volume control. |
| With Static RAG (Model B) | ## 8. Monitoring effectiveness, safety, risks, and countermeasures<br><br>### A. Effectiveness monitoring<br>- **Blood pressure**<br>- Monitor office and preferably home BP closely.<br>- Main target: maintain stable control, as improvement in proteinuria paralleled systolic BP reduction to about **100–120 mmHg**.<br>- If BP rises again, reassess both **TAK activity** and adequacy of antihypertensive therapy.<br><br>- **Proteinuria and urine findings**<br>- Serial **urine protein/creatinine ratio** and **urinalysis for hematuria**.<br>- Key efficacy marker: continued decline of proteinuria and disappearance of microhematuria, as already observed.<br><br>- **Kidney function**<br>- Follow **serum creatinine, eGFR, potassium**, and serum albumin.<br>- Goal: maintain renal function while proteinuria improves.<br><br>- **TAK disease activity**<br>- Track **CRP and ESR**, symptoms (headache, neck pain, malaise), inter-arm BP difference, and signs related to aortic regurgitation or ischemia.<br>- Periodic **vascular imaging** (e.g., CT/MRI/vascular ultrasound as appropriate) to assess progression or stability of large-vessel lesions.<br><br>- **Cardiac/volume status**<br>- Monitor for edema, body weight, chest symptoms, chest X-ray/echocardiography |

| | |
|---|---|
| | as clinically indicated, especially because pulmonary congestion and severe AR were present.<br><br>### B. Safety monitoring and major risks<br>1. **Sacubitril/valsartan**<br>- Risks: **hypotension**, decline in **GFR**, **hyperkalemia**.<br>- Countermeasures:<br>- Check BP, creatinine/eGFR, and potassium soon after initiation/dose changes, then periodically.<br>- If symptomatic hypotension or renal dysfunction occurs, reduce dose, review volume status, and adjust concomitant antihypertensives/diuretics.<br><br>2. **Prednisolone**<br>- Risks: **infection, hyperglycemia, osteoporosis, fluid retention, muscle weakness, psychiatric effects**.<br>- Countermeasures:<br>- Monitor temperature/symptoms of infection, CBC, glucose/HbA1c as needed.<br>- Bone protection and gradual taper guided by disease activity.<br>- Use the lowest effective dose.<br><br>3. **Tocilizumab**<br>- Risks: **serious infection, liver dysfunction, cytopenia, dyslipidemia**; also **CRP may become unreliable** as an inflammation marker.<br>- Countermeasures:<br>- Periodic CBC, liver enzymes, lipid profile.<br>- Rely more on **clinical symptoms and imaging**, not CRP alone, to assess TAK relapse.<br>- Hold treatment and evaluate promptly if infection is suspected.<br><br>4. **Persistent severe hypertension / vascular complications**<br>- Risks: progression of **glomerular endothelial injury**, recurrent thrombus formation, worsening AR, heart failure, stroke.<br>- Countermeasures:<br>- Strict BP control, reassess adherence and drug regimen, and repeat vascular/cardiac evaluation if BP worsens.<br><br>5. **Nephrotic syndrome-related thrombosis**<br>- Risks: venous/arterial thrombosis, especially if heavy proteinuria relapses.<br>- Countermeasures:<br>- Clinical surveillance for thrombotic events; reassess risk if proteinuria increases markedly or serum albumin falls again. |
| With Static RAG and Mult-agent framework (Model C) | ## 8. Monitoring effectiveness, safety, risks, and countermeasures<br><br>### A. Monitoring treatment effectiveness<br>1. **Takayasu arteritis activity**<br>- Follow **symptoms** such as headache, neck pain, and malaise.<br>- Measure **BP in both arms** and track any inter-arm difference.<br>- Follow **CRP/ESR**, but interpret **CRP cautiously once tocilizumab is started**, because IL-6 blockade can suppress CRP despite ongoing disease.<br>- Perform **serial vascular imaging** as appropriate to assess aortic, subclavian, and carotid lesions.<br>- Monitor **aortic regurgitation (AR) and cardiac function** with echocardiography. |

| | |
|---|---|
|  | 2. **Renal response**<br> - Follow **urine protein/Cr ratio**, **urinalysis**, and especially **urine sediment/microhematuria**.<br> - Monitor **serum albumin**, body weight, edema, and signs of pulmonary congestion.<br> - Follow **serum creatinine/eGFR**.<br> - Closely track **BP trajectory**, because in this case the kidney biopsy supports **hypertension/endothelial injury–related glomerular microangiopathy without renal artery stenosis**, and renal improvement paralleled tighter systolic BP control.<br><br>3. **Expected response in this case**<br> - Improvement in **proteinuria, microhematuria, serum albumin, edema, pulmonary congestion, and BP**.<br> - Suppression of inflammatory activity by symptoms, ESR, and imaging, rather than CRP alone under tocilizumab.<br> - Stable renal function.<br><br>### B. Safety risks and concrete countermeasures<br>1. **Excessive BP lowering / hypotension**<br> - Risk with **nifedipine plus sacubitril/valsartan**, especially after inflammation and AR-related hemodynamic stress improve.<br> - Monitor home and clinic BP, dizziness, falls, and renal function.<br> - Titrate antihypertensives to maintain control without symptomatic hypotension.<br><br>2. **Renal function decline or hyperkalemia from sacubitril/valsartan**<br> - Monitor **Cr/eGFR and serum K**, especially soon after initiation or dose changes.<br> - Reduce or hold the drug if creatinine rises significantly or hyperkalemia develops.<br><br>3. **Heart failure / volume overload related to severe AR**<br> - Monitor dyspnea, edema, body weight, chest X-ray if needed, and **serial echocardiography**.<br> - Optimize BP and volume control and obtain cardiology input if AR or congestion worsens.<br><br>4. **Relapse or progression of TAK**<br> - Monitor for recurrent symptoms, worsening BP control, new vascular findings, or rising ESR.<br> - Remember that relapse may appear as **worsening hypertension or proteinuria** even if CRP remains suppressed on tocilizumab.<br> - Reassess disease activity promptly with clinical evaluation and imaging.<br><br>5. **Recurrent endothelial/microangiopathic renal injury**<br> - Monitor **urine sediment, proteinuria, renal function, and BP**, because the biopsy showed thrombus formation, endotheliosis, and glomerular microangiopathy.<br> - Tight hemodynamic control is the main countermeasure.<br><br>6. **Glucocorticoid adverse effects**<br> - Monitor for infection, hyperglycemia, osteoporosis, myopathy, and psychiatric effects.<br> - Taper to the lowest effective dose and continue steroid-sparing therapy when possible. |

| | |
|---|---|
| | 7. **Tocilizumab adverse effects**<br>- Monitor **CBC, liver enzymes, and infection symptoms**.<br>- Because CRP is less reliable under IL-6 blockade, rely more on **symptoms, BP trends, renal findings, and imaging**.<br><br>8. **Residual nephrotic-state thrombosis risk**<br>- Monitor for sudden dyspnea, chest pain, unilateral leg swelling, or other thrombotic symptoms.<br>- Risk should fall as proteinuria remits, but investigate promptly if symptoms arise.<br><br>### Changes from the initial version<br>- I made it explicit that the renal lesion was **biopsy-supported as hypertension/endothelial injury–related glomerular microangiopathy without renal artery stenosis**.<br>- I added closer monitoring of **urine sediment**, **volume status**, **serial Cr/K after sacubitril/valsartan changes**, and **echocardiographic AR surveillance**.<br>- I clarified that under **tocilizumab**, **CRP may be misleading**, so relapse may instead appear through **worsening BP, proteinuria, or vascular/cardiac findings**. |
| **Phase III – Q9: Formulating plans for worsening conditions** | |
| Baseline LLM (Model A) | ### 9. Reassessment and action plan if the course is atypical or worsens<br><br>1. **First, define what is worsening**<br>- Is it **proteinuria/hematuria**, **kidney function**, **BP**, **systemic inflammation**, or **cardiopulmonary status**?<br>- Repeat focused history, examination, and key labs immediately.<br><br>2. **If proteinuria or kidney function worsens**<br>- Reassess for:<br>- uncontrolled **hypertension/volume overload**,<br>- progression of **glomerular microangiopathy**,<br>- drug-related kidney injury,<br>- superimposed causes of AKI (infection, dehydration, heart failure).<br>- Actions:<br>- repeat urinalysis, urine protein quantification, creatinine/eGFR, electrolytes;<br>- reassess BP and volume status;<br>- consider repeat renal imaging if clinically indicated;<br>- if the course becomes inconsistent with the original interpretation, reconsider pathology review and alternative renal diagnoses.<br><br>3. **If inflammatory markers or vascular symptoms worsen**<br>- Consider **relapse or progression of TAK** despite therapy.<br>- Actions:<br>- repeat CRP/ESR and vascular imaging,<br>- reassess adherence/drug exposure,<br>- intensify immunosuppression if active vasculitis is confirmed, in collaboration with relevant specialists.<br><br>4. **If sudden dyspnea, pulmonary edema, chest symptoms, or heart failure occurs**<br>- Consider worsening **aortic regurgitation**, uncontrolled hypertension, or fluid overload.<br>- Actions:<br>- urgent chest imaging/echocardiography, BP control, diuresis, cardiology review. |

| | |
|---|---|
| | 5. **If fever or systemic decline occurs**<br>- Distinguish **infection** from vasculitis flare, especially because **tocilizumab can mask CRP elevation**.<br>- Actions:<br>- low threshold for cultures/imaging and temporary treatment adjustment based on severity.<br><br>6. **Flexible revision of diagnosis/treatment**<br>- If the response is not durable or new findings appear, revisit whether nephrotic syndrome is fully explained by **hypertension-related endothelial/podocyte injury associated with TAK**, or whether another superimposed glomerular disease is present.<br>- Revise therapy according to the dominant mechanism: tighter BP/volume control, escalation of vasculitis treatment, or renewed renal diagnostic review. |
| With Static RAG (Model B) | ## 9. If the condition does not improve or changes suddenly: reassessment and action plan<br><br>### A. Reconsider the mechanism first<br>If proteinuria, hematuria, kidney function, BP, edema, or inflammatory findings worsen unexpectedly, reassess whether the problem is due to:<br>1. **Persistent or relapsing TAK activity**<br>2. **Insufficient BP/volume control**<br>3. **Progression of cardiac disease**, especially severe AR or heart failure<br>4. **Drug-related adverse effects**<br>5. **A different or additional renal process** not fully explained by hypertensive/glomerular microangiopathic injury<br><br>### B. Immediate practical response<br>- Recheck:<br>- **BP trend**, body weight, edema, oxygenation<br>- **Urinalysis, urine protein/Cr, serum creatinine/eGFR, potassium, albumin**<br>- **CRP/ESR** (with caution if on tocilizumab)<br>- Reassess for:<br>- **Pulmonary congestion/heart failure**<br>- **Infection**<br>- **Thrombotic events**<br>- New vascular ischemic symptoms<br><br>### C. Diagnostic revision if needed<br>- If inflammation or vascular symptoms recur despite treatment:<br>- Repeat **vascular imaging** to evaluate progression/relapse of TAK.<br>- If renal findings worsen disproportionately despite good BP and controlled TAK:<br>- Re-examine the kidney biopsy interpretation and consider whether another glomerular disease or superimposed thrombotic microangiopathic process is present.<br>- If creatinine rises after therapy changes:<br>- Distinguish **hemodynamic effect of RAAS/neprilysin blockade** from intrinsic renal worsening.<br><br>### D. Treatment adjustment<br>- **If TAK relapse is suspected**: intensify immunosuppression (e.g., optimize glucocorticoid regimen and reassess biologic therapy).<br>- **If BP remains uncontrolled**: further optimize antihypertensives and volume management. |

| | |
|---|---|
| | - **If heart failure/AR worsens**: urgent cardiology reassessment.<br>- **If adverse drug effects occur**: reduce, hold, or switch the responsible drug and provide supportive care.<br><br>In short, if the course deviates from expectation, the key is to rapidly determine whether the driver is **active vasculitis, hemodynamic stress from uncontrolled BP/AR, treatment toxicity, or an alternative renal pathology**, and then revise therapy accordingly. |
| With Static RAG and Mult-agent framework (Model C) | ## 9. Reassessment when the course worsens or changes suddenly<br><br>### A. Reassess in parallel, but prioritize the mechanisms supported by this case<br>If proteinuria, hematuria, edema, renal function, BP, or symptoms worsen, reassess both:<br>1. **TAK activity/progression**, and<br>2. **Hemodynamic-cardiac causes of renal injury**, which in this case are strongly supported by biopsy and treatment response.<br><br>Because the biopsy showed **endothelial injury, thrombus formation, and glomerular microangiopathy without immune-complex deposition**, and because proteinuria improved markedly after stricter **BP and volume control**, deterioration should first prompt reassessment of:<br>- **loss of BP control**,<br>- **worsening AR or pulmonary congestion**,<br>- **drug-related hemodynamic renal effects**, and<br>- **recurrent endothelial/microangiopathic injury**.<br><br>### B. Key scenarios and actions<br>1. **Proteinuria or hematuria worsens**<br>- First reassess **BP trajectory**, adherence, volume overload, and signs of worsening AR/heart failure.<br>- Repeat **urinalysis/urine sediment**, urine protein quantification, serum albumin, and renal function.<br>- If worsening tracks with poor BP control or congestion, intensify hemodynamic and cardiac management.<br>- Only if urinary abnormalities fail to track with hemodynamic and inflammatory control should a separate glomerular disease be reconsidered.<br><br>2. **Creatinine rises after treatment changes**<br>- Distinguish among:<br>- hemodynamic fall in GFR from **sacubitril/valsartan**,<br>- volume depletion,<br>- worsening heart failure/AR,<br>- persistent severe hypertension with renal injury,<br>- progression of intrinsic kidney disease.<br>- Reassess **BP, volume status, creatinine, potassium, urinalysis**, and medication timing; then adjust treatment promptly.<br><br>3. **BP rises again or becomes difficult to control**<br>- Consider **TAK progression/relapse**, worsening AR, fluid overload, or adherence issues.<br>- Repeat clinical assessment and vascular/cardiac evaluation.<br>- This is especially important because in this case worsening systolic BP was likely a major driver of glomerular endothelial damage. |

| | |
|---|---|
| | 4. **Pulmonary congestion or dyspnea appears**<br>- Urgently evaluate for **worsening AR, heart failure, or volume overload**.<br>- Repeat chest imaging and echocardiography as indicated.<br><br>5. **Inflammatory or vascular features recur**<br>- New headache, neck pain, constitutional symptoms, or new vascular findings should raise concern for **TAK relapse**.<br>- Do not rely on CRP alone if on tocilizumab; use **symptoms, ESR, BP pattern, and imaging**.<br><br>6. **No improvement despite controlled inflammation**<br>- If renal findings do not improve despite good BP control, stable cardiac status, and suppressed TAK activity, revisit the renal diagnosis.<br>- Re-review biopsy findings and consider whether another superimposed glomerular disease is present.<br><br>### C. Practical principle<br>Do not anchor on a single explanation, but in this case **prioritize the mechanisms actually supported by the biopsy and treatment course**:<br>- **hemodynamic stress from uncontrolled systolic BP**,<br>- **severe AR/volume overload**,<br>- **endothelial injury with glomerular microangiopathy**, and<br>- **vasculitis relapse/progression**.<br><br>Treatment should then be revised according to the dominant active mechanism:<br>- **increase vasculitis control** if TAK is relapsing,<br>- **tighten BP/volume and cardiac management** if hemodynamic stress is driving renal injury,<br>- **adjust sacubitril/valsartan or other drugs** if adverse hemodynamic kidney effects occur,<br>- and reconsider another renal disease only if the course no longer fits the biopsy-supported explanation.<br><br>### Changes from the initial version<br>- I now more explicitly prioritize reassessment of **BP control, AR/pulmonary congestion, volume status, creatinine/K, and recurrent endothelial microangiopathic injury** before broadly relabeling the renal disease.<br>- I incorporated the biopsy and response-to-treatment evidence showing that the nephrotic syndrome was most plausibly driven by **uncontrolled systolic hypertension, severe AR/volume stress, and endothelial glomerular injury**, not primary immune-complex glomerulonephritis.<br>- I reserved reconsideration of a separate glomerular disease for cases where **proteinuria/hematuria do not track with hemodynamic and inflammatory control**. |

| **Case 7: Successful use of avacopan in a case of ANCA-associated vasculitis with treatment-resistant medium-sized vessel involvement** [15] | |
|---|---|
| **Phase I – Q1: Summarizing medical problems** | |
| Baseline LLM (Model A) | 1. **Problem representation**<br><br>A **75-year-old woman** with a **4-month subacute systemic illness** developed **purpura/erythema/induration of the lower legs**, later **headache, anorexia, and |

| | |
|---|---|
| | right leg numbness**, was found to have **interstitial pneumonia**, and then over **about 2 weeks** progressed from previously normal urinalysis/renal function to **hematuria, proteinuria, and severe acute kidney failure (Cr 14.93 mg/dL) requiring hemodialysis**, with **rising MPO-ANCA titers** and prior **granulomatous skin biopsy**. This is most concerning for **ANCA-associated small-vessel vasculitis with rapidly progressive glomerulonephritis**, likely a **pulmonary-renal systemic vasculitic process**. |
| With Static RAG (Model B) | ## 1. Problem representation<br>A **75-year-old woman** with a **4-month subacute course** of **purpura/erythema/induration of the lower legs**, followed by **headache, anorexia, and right lower-extremity numbness**, later developing **interstitial pneumonia**, and then **abrupt onset of hematuria, proteinuria, and severe acute kidney injury (Cr 14.93 mg/dL) requiring hemodialysis**, with **rising MPO-ANCA positivity**, presents with **rapidly progressive glomerulonephritis (RPGN) suspicious for systemic ANCA-associated vasculitis**.<br>Key features suggesting a systemic small-vessel vasculitis are:<br>- **Skin involvement**: purpura and biopsy-proven granulomatous inflammation<br>- **Neurologic involvement**: focal lower-extremity numbness suggesting possible vasculitic neuropathy<br>- **Pulmonary involvement**: interstitial pneumonia<br>- **Renal involvement**: new hematuria/proteinuria with rapidly progressive renal failure<br>- **Serology**: rising **MPO-ANCA** |
| With Static RAG and Mult-agent framework (Model C) | ## 1. Problem representation<br><br>A **75-year-old woman** with a **4-month subacute multisystem illness** developed **purpura, erythema, and induration of the lower legs**, followed by **headache, anorexia, and right lower-extremity numbness**, then was found to have **CT-defined interstitial pneumonia of unclear etiology**, and subsequently had **abrupt hematuria, proteinuria, and severe acute kidney injury (Cr 14.93 mg/dL) progressing to dialysis-dependent rapidly progressive glomerulonephritis**, with **rising MPO-ANCA titers** and a prior skin biopsy showing **granulomatous inflammation** that is **supportive but not specific**.<br><br>This is best summarized as a **multisystem inflammatory syndrome highly suspicious for MPO-ANCA-associated vasculitis**, with **renal, skin, possible neurologic, and possible pulmonary involvement**, although the pulmonary and cutaneous findings are **not yet pathologically specific** for AAV. |
| **Phase I – Q2: Differential diagnoses and rationale** | |
| Baseline LLM (Model A) | 2. **Differential diagnoses**<br><br>**Top 3 differential diagnoses**<br>1. **MPO-ANCA–associated vasculitis causing pauci-immune crescentic GN**<br>- Most likely.<br>- Supported by **rapidly progressive GN**, **hematuria/proteinuria**, **MPO-ANCA positivity with rising titer**, **skin lesions compatible with vasculitis**, possible **mononeuritis/peripheral neuropathy**, and **interstitial lung involvement**.<br>- Phenotypically this could fit **microscopic polyangiitis (MPA)** best, although the granulomatous skin finding leaves room for overlap with GPA. |

| | |
|---|---|
| | 2. **Granulomatosis with polyangiitis (GPA)**<br>- Considered because of **granulomatous lesion on skin biopsy**, systemic symptoms, neuropathy, and RPGN.<br>- Less typical because **MPO-ANCA** is more often associated with MPA, and no clear ENT manifestations are provided, but GPA remains plausible.<br><br>3. **Anti-GBM disease / pulmonary-renal syndrome or other crescentic GN mimic**<br>- Must be considered in any patient with **abrupt severe RPGN**.<br>- Less favored because the available data point more toward AAV, but missing it would be dangerous because it requires urgent diagnosis and specific treatment.<br><br>**Most likely diagnosis:**<br>- **MPO-ANCA-associated vasculitis with pauci-immune crescentic glomerulonephritis**, probably **microscopic polyangiitis**.<br><br>**Must not be missed:**<br>- **Life-threatening pulmonary-renal vasculitic syndrome**, especially **diffuse alveolar hemorrhage**, and **anti-GBM disease**.<br>- These can rapidly worsen and change immediate treatment intensity. |
| With Static RAG (Model B) | ## 2. Differential diagnosis<br>### Top 3 differential diagnoses<br>1. **Microscopic polyangiitis (MPA) / MPO-ANCA-associated vasculitis**<br>- Most likely because of **MPO-ANCA positivity**, **RPGN**, **skin lesions**, possible **mononeuritis**, and **interstitial lung disease**, which is well recognized in MPO-AAV.<br><br>2. **Granulomatosis with polyangiitis (GPA)**<br>- Considered because the skin biopsy reportedly showed **granulomatous lesions**, and GPA can cause **multisystem disease** including renal, neurologic, pulmonary, and skin involvement. However, MPO-ANCA and interstitial pneumonia are somewhat more supportive of MPA than classic GPA.<br><br>3. **Secondary systemic vasculitis or vasculitis mimic**<br>- Includes **infection-related GN/vasculitis mimic**, **malignancy-associated vasculitis**, or **anti-GBM disease / overlap syndrome**. These are less likely but important because they can also present with RPGN and systemic features.<br><br>### Most likely diagnosis<br>**MPO-ANCA-associated vasculitis causing RPGN**, most likely **microscopic polyangiitis**, possibly with systemic involvement of **skin, peripheral nerve, and lung**.<br><br>### Must not be missed<br>**Anti-GBM disease (including double-positive disease)** and **infection-related processes** must not be missed.<br>- **Anti-GBM disease** can cause fulminant crescentic GN and pulmonary involvement and requires urgent diagnosis because management is time-sensitive.<br>- **Infection-related endocarditis or chronic infection** can mimic ANCA vasculitis and may worsen if immunosuppression is given inappropriately. |
| With Static RAG and | ## 2. Prioritized differential diagnosis<br><br>### Most likely |

| | |
|---|---|
| Mult-agent framework (Model C) | 1. **Microscopic polyangiitis (MPA) / MPO-ANCA-associated vasculitis**<br>- Best fits the combination of **MPO-ANCA positivity**, **rapidly progressive GN**, **purpura**, possible **vasculitic neuropathy**, and possible **lung involvement**.<br>- The renal presentation is particularly characteristic of **pauci-immune crescentic GN** due to AAV.<br><br>### Must-not-miss urgent alternative<br>2. **Anti-GBM disease, including ANCA/anti-GBM double-positive disease**<br>- Important because it can present with **fulminant RPGN**, possible pulmonary involvement, and requires **urgent diagnosis and treatment**.<br>- Severity and tempo of renal failure make this a critical exclusion.<br><br>### Other important alternatives<br>3. **Infection-related glomerulonephritis / infective endocarditis-associated vasculitic syndrome**<br>- Can mimic AAV with **constitutional symptoms, GN, purpura, positive autoantibodies**, and systemic inflammation.<br>- Must be considered before immunosuppression.<br><br>4. **Drug-induced ANCA-associated vasculitis**<br>- Certain drugs can cause **MPO-ANCA-positive vasculitis** with renal and skin involvement.<br>- Requires careful medication reconciliation.<br><br>5. **Granulomatosis with polyangiitis (GPA)**<br>- Less likely than MPA because **MPO** rather than PR3 is reported, and there are **no clear ENT/destructive upper airway features**.<br>- The skin "granulomatous lesion" alone is **not specific** enough to strongly favor GPA.<br><br>6. **Eosinophilic granulomatosis with polyangiitis (EGPA)**<br>- Less likely because there is **no asthma, eosinophilia, or allergic history** provided.<br><br>7. **Immune-complex vasculitis / connective tissue disease-related GN / cryoglobulinemic vasculitis**<br>- Less supported, but should remain in the differential until serologies and biopsy clarify the process.<br><br>### Most likely diagnosis<br>**MPO-ANCA-associated vasculitis causing pauci-immune crescentic glomerulonephritis, most consistent with MPA.** |
| **Phase I – Q3: Necessary physical examinations and reasons** | |
| Baseline LLM (Model A) | 3. **Additional history and physical examination**<br><br>**History**<br>- **Pulmonary symptoms:** dyspnea, cough, hemoptysis<br>- To assess for **alveolar hemorrhage** or active pulmonary vasculitis.<br>- **ENT symptoms:** chronic sinusitis, nasal crusting/epistaxis, hearing loss<br>- Helps distinguish **GPA** from MPA.<br>- **Neurologic symptoms:** asymmetric weakness, sensory loss, foot drop<br>- To identify **mononeuritis multiplex** from systemic vasculitis. |

| | |
|---|---|
|  | - **Skin symptom evolution:** palpable purpura, ulcers, painful nodules<br> - To characterize **cutaneous vasculitis**.<br>- **Renal syndrome details:** oliguria, edema, gross hematuria<br> - To gauge severity of **GN/AKI**.<br>- **Constitutional symptoms:** fever, weight loss<br> - Support systemic inflammatory disease.<br>- **Medication and exposure history:** hydralazine, PTU, minocycline, cocaine; infection risk factors<br> - To evaluate **drug-induced ANCA vasculitis** or infection-related mimics.<br>- **Autoimmune symptoms:** sicca, arthralgia, Raynaud, oral ulcers<br> - To assess for secondary autoimmune disease.<br><br>**Physical examination**<br>- **Vital signs and oxygen saturation**<br> - Severity assessment; hypoxemia may suggest pulmonary involvement.<br>- **Chest exam**<br> - Crackles/respiratory distress suggesting interstitial disease or hemorrhage.<br>- **ENT exam**<br> - Nasal ulcers, crusting, sinus tenderness, otologic findings for GPA.<br>- **Skin exam**<br> - Distribution and morphology of purpura/nodules/ulcers to support vasculitis.<br>- **Detailed neurologic exam**<br> - Focal deficits suggesting mononeuritis multiplex.<br>- **Volume status / edema / blood pressure**<br> - To assess renal failure consequences.<br>- **Abdominal/back exam**<br> - Severe right back pain raises concern for other pathology such as retroperitoneal, musculoskeletal, or renal complication. |
| With Static RAG (Model B) | ## 3. Additional history and physical examination<br>### Additional history<br>- **Pulmonary symptoms**: hemoptysis, dyspnea, cough<br> - To assess for **diffuse alveolar hemorrhage** or progressive pulmonary vasculitis.<br>- **ENT symptoms**: sinusitis, nasal crusting/bleeding, otitis, hearing changes<br> - To look for features favoring **GPA**.<br>- **Neurologic symptoms**: asymmetric weakness, foot drop, sensory distribution, progression of numbness<br> - To evaluate for **mononeuritis multiplex**.<br>- **Constitutional symptoms**: fever, weight loss, night sweats<br> - To support systemic inflammation or suggest infection/malignancy.<br>- **Drug history**: hydralazine, propylthiouracil, minocycline, allopurinol, cocaine exposure<br> - To exclude **drug-induced ANCA vasculitis**.<br>- **Infectious risk/history**: heart murmur history, dental work, chronic infection, TB exposure<br> - To assess for **vasculitis mimics**.<br>- **Urinary symptoms and timeline**: gross hematuria, oliguria, edema<br> - To characterize severity and tempo of renal involvement.<br><br>### Physical examination<br>- **Skin exam**: palpable purpura, ulcers, nodules, livedo<br> - To characterize vasculitic skin disease and identify biopsy targets.<br>- **Full neurologic exam**: motor strength, sensory pattern, reflexes, gait<br> - To detect **mononeuritis multiplex**. |

| | |
|---|---|
|  | - **Chest exam and oxygen saturation**<br> - To identify pulmonary hemorrhage or interstitial lung disease severity.<br>- **ENT exam**<br> - To look for GPA-type upper airway involvement.<br>- **Volume status / edema / blood pressure**<br> - To assess consequences of severe renal failure.<br>- **Cardiac exam**<br> - To look for murmurs suggesting **infective endocarditis**. |
| With Static RAG and Mult-agent framework (Model C) | ## 3. Additional history and physical examination<br><br>### Focused history<br>1. **Pulmonary symptoms**: dyspnea, cough, hemoptysis<br> - To assess for **diffuse alveolar hemorrhage** or active vasculitic lung disease rather than nonspecific "interstitial pneumonia."<br><br>2. **ENT symptoms**: sinusitis, epistaxis, nasal crusting, otitis, hearing loss<br> - To look for features supporting **GPA**.<br><br>3. **Neurologic symptoms**: foot drop, asymmetric weakness, sensory loss progression<br> - To evaluate for **mononeuritis multiplex** from vasculitis.<br><br>4. **Medication exposure in detail**<br> - Especially **hydralazine, propylthiouracil, minocycline, allopurinol, levamisole-contaminated drugs, recent antibiotics**.<br> - Important for **drug-induced AAV**.<br><br>5. **Infectious history**<br> - Fever, chills, recent dental work, bacteremia risk, murmur history, indwelling lines.<br> - Helps assess for **infection-related GN or endocarditis**.<br><br>6. **Urine output trajectory and timing of renal decline**<br> - Helps define acuity and severity of RPGN.<br><br>7. **Constitutional symptoms**<br> - Weight loss, night sweats, decline in function.<br> - Useful for inflammatory burden and infection/malignancy screening.<br><br>8. **Asthma/allergy/eosinophilic history**<br> - To assess for **EGPA**.<br><br>### Focused physical examination<br>1. **Skin exam**<br> - Look for **active palpable purpura, ulcers, nodules, livedo**, and identify lesions suitable for repeat biopsy.<br><br>2. **Neurologic exam**<br> - Check for **asymmetric distal weakness, foot drop, sensory deficits, reflex changes** suggesting vasculitic neuropathy.<br><br>3. **Chest exam and oxygen saturation**<br> - Evaluate for **hypoxemia, crackles, pulmonary hemorrhage, or volume overload**. |

| | |
|---|---|
| | 4. **ENT exam**<br>- Assess for findings that would support **GPA**.<br><br>5. **Cardiac exam**<br>- Look for **murmur or stigmata of endocarditis**.<br><br>6. **Volume status and blood pressure**<br>- Important in severe AKI and dialysis-dependent illness.<br><br>7. **Back/abdominal exam**<br>- Severe right back pain could reflect **retroperitoneal pathology, renal capsular process, stone, infarction, or other urgent nonvasculitic causes**. |
| **Phase I – Q4: Plan for investigations/tests** | |
| Baseline LLM (Model A) | 4. **Tests to narrow the diagnosis**<br><br>1. **Urinalysis with sediment and urine protein quantification**<br>- Purpose: confirm active glomerulonephritis.<br>- Expected: **dysmorphic RBCs, RBC casts, proteinuria**.<br><br>2. **Basic blood tests: CBC, CMP, CRP/ESR**<br>- Purpose: assess inflammation, anemia, electrolyte derangements, dialysis indication, and possible pulmonary hemorrhage-related anemia.<br>- Expected: inflammatory markers elevated; anemia may be present; severe renal dysfunction already known.<br><br>3. **ANCA testing with antigen specificity (MPO/PR3)**<br>- Purpose: characterize AAV subtype.<br>- Expected: **MPO-ANCA positive**.<br><br>4. **Anti-GBM antibody**<br>- Purpose: exclude **anti-GBM disease**, an important pulmonary-renal emergency.<br>- Expected: likely negative, but must be checked.<br><br>5. **Complement levels (C3/C4), ANA, anti-dsDNA, cryoglobulins, hepatitis B/C serologies**<br>- Purpose: evaluate immune-complex vasculitis or connective tissue disease mimics.<br>- Expected: in AAV, complements are usually **normal**.<br><br>6. **Chest CT and/or chest radiograph**<br>- Purpose: define lung involvement and look for **alveolar hemorrhage**, nodules, or interstitial changes.<br>- Expected: may show previously noted interstitial abnormalities; hemorrhagic changes would alter urgency.<br><br>7. **Renal ultrasound**<br>- Purpose: exclude obstruction and assess kidney size before biopsy.<br>- Expected: no obstruction; kidneys may be normal-sized.<br><br>8. **Kidney biopsy**<br>- Purpose: definitive diagnosis and classification of RPGN; essential for treatment planning if feasible despite dialysis dependence. |

| | |
|---|---|
| | - Expected: **pauci-immune necrotizing crescentic glomerulonephritis** in AAV.<br><br>9. **Review/re-evaluate skin biopsy pathology**<br>- Purpose: determine whether lesions show **vasculitis**, granulomatous inflammation, or another process.<br>- Expected: findings supportive of systemic vasculitis may help phenotype the disease.<br><br>10. **If respiratory symptoms or imaging suggest hemorrhage: bronchoscopy/BAL**<br>- Purpose: confirm **diffuse alveolar hemorrhage** when clinically necessary.<br>- Expected: progressively bloodier lavage if present.<br><br>Overall, the diagnostic plan should prioritize confirming **AAV with RPGN**, excluding **anti-GBM disease and other mimics**, and identifying **immediately life-threatening pulmonary involvement**. |
| With Static RAG (Model B) | ## 4. Tests to narrow the diagnosis<br>### Essential tests<br>1. **Kidney biopsy**<br>- **Purpose**: establish the cause of RPGN and distinguish pauci-immune GN from anti-GBM disease, immune-complex GN, or other mimics.<br>- **Expected result**: **necrotizing crescentic pauci-immune glomerulonephritis** if AAV is correct.<br><br>2. **Urinalysis with sediment and urine protein quantification**<br>- **Purpose**: confirm active glomerulonephritis.<br>- **Expected result**: **dysmorphic RBCs, RBC casts, proteinuria**.<br><br>3. **Repeat ANCA testing with PR3-ANCA and MPO-ANCA titers**<br>- **Purpose**: support AAV subtype.<br>- **Expected result**: persistent **MPO-ANCA positivity**.<br><br>4. **Anti-GBM antibody**<br>- **Purpose**: exclude **anti-GBM disease / double-positive disease**.<br>- **Expected result**: likely negative, but crucial not to miss.<br><br>5. **Basic serologic workup for alternative causes**<br>- **ANA, dsDNA, complements, cryoglobulins, hepatitis B/C, serum protein studies**<br>- **Purpose**: exclude immune-complex vasculitis, lupus, cryoglobulinemia, infection-related disease, monoclonal disorders.<br>- **Expected result**: often non-diagnostic/negative in pauci-immune AAV; complements usually normal.<br><br>6. **Inflammatory markers and CBC**<br>- **Purpose**: assess systemic inflammation, anemia, eosinophilia, thrombocytopenia.<br>- **Expected result**: inflammatory response; anemia of inflammation/renal disease possible. Eosinophilia would raise alternative considerations such as EGPA.<br><br>7. **Chest CT**<br>- **Purpose**: characterize lung involvement and evaluate for **interstitial lung disease vs alveolar hemorrhage vs nodules**.<br>- **Expected result**: findings compatible with **interstitial pneumonia**; if |

| | |
|---|---|
| | hemorrhage is present, new diffuse ground-glass opacities may appear.<br><br>8. **Nerve conduction studies** if neuropathy is clinically significant<br>- **Purpose**: confirm **vasculitic neuropathy/mononeuritis multiplex**.<br>- **Expected result**: asymmetric axonal neuropathy.<br><br>9. **Infection screening as indicated**: blood cultures, echocardiography if endocarditis suspected<br>- **Purpose**: exclude infection before immunosuppression.<br>- **Expected result**: negative if primary AAV.<br><br>Overall, the diagnostic priority is to **confirm pauci-immune crescentic GN by kidney biopsy**, while urgently excluding **anti-GBM disease** and **infection-related mimics**. |
| With Static RAG and Mult-agent framework (Model C) | ## 4. Testing plan<br><br>### Highest-priority tests<br>1. **Urgent kidney biopsy, if feasible and safe**<br>- Best test to confirm **pauci-immune crescentic GN** and distinguish AAV from **anti-GBM** or immune-complex disease.<br>- Must consider **bleeding risk, hemodynamic stability, and dialysis status**.<br><br>2. **Anti-GBM antibody**<br>- Essential to exclude **anti-GBM disease** or **double-positive disease**.<br><br>3. **Urinalysis with microscopy and urine protein quantification**<br>- To document active nephritic sediment and quantify renal involvement.<br><br>4. **CBC, CMP/BMP, CRP/ESR**<br>- Assess anemia, inflammation, electrolyte derangements, and ongoing renal severity.<br><br>5. **ANCA testing with MPO and PR3 specificity**<br>- Helps support AAV subtype, though biopsy remains more definitive.<br><br>### Tests for mimics and classification<br>6. **Complement levels (C3/C4), ANA, anti-dsDNA, RF, cryoglobulins**<br>- To evaluate for **immune-complex or connective tissue disease-related GN**.<br><br>7. **Infectious evaluation**<br>- **Blood cultures if clinically indicated**, plus **hepatitis B/C screening** and other infection screening before immunosuppression.<br>- Consider endocarditis evaluation if suggested by history/exam.<br><br>8. **Review prior chest CT / repeat chest imaging if needed**<br>- To clarify whether the lung findings represent **interstitial lung disease, inflammatory infiltrates, or alveolar hemorrhage**.<br>- Bronchoscopy is not routine unless symptoms/imaging suggest hemorrhage and results would change management.<br><br>9. **Review prior skin biopsy; repeat skin biopsy with immunofluorescence if active lesions are present**<br>- May help better classify the cutaneous process, though prior granulomatous |

| | |
|---|---|
| | inflammation is **not specific**.<br><br>10. **Nerve conduction studies if deficits are present**<br>- To support **mononeuritis multiplex**.<br><br>11. **Imaging for severe right-sided back pain**<br>- For example, **targeted CT depending on suspected cause and renal/dialysis context**.<br>- Needed to exclude urgent complications such as **retroperitoneal hemorrhage, renal infarction, stone, or fracture**.<br><br>### Management urgency<br>Because she already has **dialysis-dependent severe RPGN**, diagnostic workup should proceed **urgently in parallel with treatment planning**, especially exclusion of **anti-GBM disease and infection** and assessment of **biopsy feasibility/safety**.<br><br>---<br><br>## Changes from the initial version<br><br>1. **Problem representation was made more cautious**<br>- I changed "interstitial pneumonia" from presumed vasculitic lung involvement to **pulmonary involvement of unclear etiology**.<br>- I also clarified that the **granulomatous skin biopsy is supportive but not specific**, rather than using it to strongly imply GPA.<br><br>2. **Differential diagnosis was reprioritized**<br>- **MPA/MPO-AAV** remains first.<br>- **Anti-GBM/double-positive disease** was moved up as the key urgent alternative.<br>- **Infection-related GN/endocarditis** and **drug-induced AAV** are now emphasized before GPA/EGPA.<br><br>3. **History and exam were refined**<br>- Added focus on **medication reconciliation**, **urine output trajectory**, **hemoptysis/hypoxemia**, and **active skin lesions for possible repeat biopsy**.<br><br>4. **Testing plan now emphasizes urgency and feasibility**<br>- I explicitly stated that **diagnostics and treatment planning should occur in parallel**.<br>- I noted that **kidney biopsy depends on procedural safety**.<br>- I avoided overcommitting to bronchoscopy and framed chest evaluation according to **symptoms, oxygenation, and imaging**. |
| **Phase II – Q5: Interpreting test results** | |
| Baseline LLM (Model A) | **5. Interpretation of the test results**<br><br>- **Marked systemic inflammation**: WBC 21,860/µL, CRP 23 mg/dL, thrombocytosis, high fibrinogen/D-dimer/FDP indicate an intense inflammatory and prothrombotic state, consistent with **active systemic vasculitis** rather than isolated kidney disease.<br>- **Renal involvement is severe and mixed**: Cr 4.44 mg/dL despite hemodialysis, hematuria (50–99/HPF), heavy proteinuria (TP/Cr 18.75 g/gCr), and granular casts |

| | |
|---|---|
| | indicate **active glomerular injury**. Very high urinary β2-microglobulin, α1-microglobulin, and NAG suggest **concomitant tubular/interstitial injury**. The degree of proteinuria is greater than typical for pure pauci-immune GN, implying extensive renal damage.<br>- **MPO-ANCA positive / PR3-ANCA negative** supports **MPO-ANCA-associated vasculitis**, while normal eosinophils/IgE argue against EGPA. Negative anti-GBM, ANA, normal complements, and negative hepatitis/HIV studies make **anti-GBM disease, immune-complex vasculitis, lupus, and infection-related causes less likely**.<br>- **Chest CT** with reticular and ground-glass opacities in both lung bases is compatible with **interstitial lung involvement** seen in MPO-AAV; there is no direct proof here of alveolar hemorrhage, but pulmonary vasculitic involvement is likely.<br>- **Renal imaging** is highly informative: **bilateral renal microaneurysms** and probable **right renal/perirenal hemorrhage** indicate **medium-vessel arteritis**, which is unusual for typical isolated AAV and raises strong concern for **polyarteritis nodosa (PAN)-like vasculitis overlap**.<br>- **Splenic hypoperfusion** suggests **visceral ischemia/infarction**, again supporting systemic vasculitis involving medium-sized arteries.<br>- **Skin biopsy** showed disruption of internal elastic lamina, fibrinoid necrosis, and neutrophilic infiltration in small-to-medium arteries, plus granulomatous tissue with giant cells. This is direct histologic evidence of **necrotizing arteritis**, not just nonspecific inflammation.<br>- **Neurologic studies** confirm multi-organ vasculitic injury: MRI-proven **acute cerebral infarction** and sural nerve biopsy showing vasculitic neuropathy with axonal damage/regeneration establish **central and peripheral nervous system involvement**. This pattern is classic for **systemic necrotizing vasculitis**.<br>- **Hypoalbuminemia (1.9 g/dL)** reflects severe inflammatory burden plus urinary protein loss.<br>- **Anemia** is compatible with inflammation, renal failure, and possibly blood loss from renal hemorrhage.<br>- **Thyroid tests** show significant hypothyroidism, but this does not explain the vasculitic syndrome.<br><br>**Overall meaning:** the data no longer fit simple MPO-AAV limited to small vessels. They indicate **highly active systemic necrotizing vasculitis with both small-vessel glomerulonephritis and medium-vessel arteritis causing aneurysms, hemorrhage, infarction, stroke, skin vasculitis, and mononeuritis multiplex**. |
| With Static RAG (Model B) | **5. Interpretation of the test results**<br><br>- **Systemic inflammation is very strong**: WBC 21,860/µL, CRP 23 mg/dL, and thrombocytosis strongly support **active systemic vasculitic inflammation** rather than isolated renal disease.<br>- **Renal involvement is severe and mixed**: Cr 4.44 mg/dL despite hemodialysis history indicates **advanced AKI/RPGN**. Urine findings of **hematuria (50–99 RBC/HPF)** and heavy **proteinuria (TP/Cr 18.75 g/gCr)** indicate marked renal injury. Very high **urinary β2-microglobulin, α1-microglobulin, and NAG** suggest substantial **tubulointerstitial injury** in addition to glomerular disease.<br>- **MPO-ANCA positivity with PR3-ANCA negativity** favors **MPO-ANCA-associated vasculitis**, but the overall pattern is not typical for isolated microscopic polyangiitis because of the medium-vessel lesions/aneurysms.<br>- **No eosinophilia or IgE elevation** argues against **eosinophilic granulomatosis with polyangiitis (EGPA)**. |

| | |
| --- | --- |
|  | - **Normal complement, negative ANA, negative anti-GBM, negative M-protein, and negative viral serologies** make immune-complex vasculitis, lupus nephritis, anti-GBM disease, paraprotein-related disease, and hepatitis-associated vasculitis less likely.<br>- **Chest CT** showing **reticular and ground-glass opacities in both lung bases** is compatible with **interstitial lung involvement**, which can occur in MPO-AAV.<br>- **Renal and splenic vascular imaging is highly important**: bilateral **renal microaneurysms**, right renal **encapsulated hemorrhage**, and **splenic hypoperfusion** indicate **medium-vessel vasculitis with aneurysm formation and ischemic organ injury**, a pattern classically seen in **polyarteritis nodosa (PAN)** rather than typical small-vessel AAV.<br>- **Skin biopsy is decisive**: disruption of the **internal elastic lamina**, **fibrinoid necrosis**, and **neutrophilic infiltration** in arteries of **68 μm and 250 μm** demonstrate **necrotizing arteritis involving small-to-medium muscular arteries**. This is pathologically much closer to **PAN-type arteritis** than pauci-immune capillaritis alone. Granulomatous tissue with giant cells suggests destructive vasculitic inflammation but does not by itself establish GPA.<br>- **Neurologic evaluation confirms active vasculitic organ damage**:<br> - **MRI**: acute cerebral infarctions explain the focal central neurologic signs.<br> - **Sural nerve biopsy**: arteriolar inflammation with myelin ovoids and regenerating clusters confirms **vasculitic neuropathy/mononeuritis multiplex**.<br>- **Hypoalbuminemia (1.9 g/dL)** reflects severe inflammatory burden and urinary protein loss.<br>- **Anemia with low iron, low TIBC, and high ferritin** is most consistent with **anemia of inflammation**, possibly compounded by renal disease and recent bleeding.<br>- **Coagulation/fibrinolysis markers** (high D-dimer, fibrinogen, FDP) fit severe inflammation/tissue injury and possibly ongoing thrombosis/bleeding burden.<br>- **TSH 76.83 with low free T4** indicates marked hypothyroidism, relevant as a comorbidity but not the main unifying diagnosis.<br><br>**Key point:** the results no longer support a pure glomerular small-vessel AAV alone. They show **overlapping small-vessel disease (glomerular/ANCA-associated features) plus clear medium-vessel necrotizing arteritis with aneurysms and infarction/hemorrhage**. |
| With Static RAG and Mult-agent framework (Model C) | ## 5. Interpretation of the test results<br><br>- **This is a severe multisystem inflammatory vasculitic syndrome, not isolated kidney disease.**<br> - Marked inflammation is shown by **WBC 21,860/μL**, **CRP 23.31 mg/dL**, **platelets 604,000/μL**, and elevated **fibrinogen/D-dimer/FDP**.<br> - In the context of skin lesions, neuropathy, cerebral infarction, renal failure, and vascular imaging abnormalities, these findings support **active systemic necrotizing vasculitis**.<br><br>- **Kidney involvement is severe and highly suspicious for vasculitic glomerular disease, with superimposed tubular injury.**<br> - She had abrupt renal failure requiring **hemodialysis**, and creatinine remains **4.44 mg/dL**.<br> - **Hematuria (50–99 RBC/HPF)** plus **heavy proteinuria (TP/Cr 18.75 g/gCr)** indicate major renal parenchymal injury with a strong glomerular component.<br> - Very high urinary **β2-microglobulin, α1-microglobulin, and NAG** support additional **tubular injury**. |

<table>
<tr><td></td><td>- **Albumin 1.9 g/dL** is explained by heavy urinary protein loss plus inflammation.<br>- However, because **renal biopsy was not performed**, the renal lesion cannot be classified with certainty as proven pauci-immune crescentic GN; the most accurate description is **severe kidney involvement highly suspicious for vasculitic GN with tubular injury**.<br><br>- **Serology supports MPO-ANCA-associated vasculitis and argues against several key mimics.**<br>- **MPO-ANCA positive (26.3 IU/mL)** with **PR3-ANCA negative** supports **MPO-AAV**.<br>- **Anti-GBM negative**, **ANA negative**, **normal complements**, and **no M-protein** make anti-GBM disease, lupus nephritis, and paraprotein/immune-complex disorders less likely.<br><br>- **Pulmonary findings are compatible with vasculitic lung involvement but are not specific.**<br>- Fine crackles and CT showing **reticular shadows and ground-glass opacities in both lung bases** may reflect **ANCA-associated interstitial lung disease or inflammatory pulmonary involvement**.<br>- Based on the provided information, there is **no clear evidence of diffuse alveolar hemorrhage**, and the CT pattern is **suggestive but not diagnostic** of vasculitic lung disease.<br><br>- **Imaging is crucial because it shows a medium-vessel vasculitic pattern.**<br>- **Multiple bilateral renal microaneurysms**, **right renal/perirenal hemorrhagic lesion**, and **splenic hypoperfusion** indicate **necrotizing arteritis with aneurysm formation and ischemic organ injury**.<br>- These findings are unusual for isolated small-vessel renal-limited AAV and strongly suggest a **PAN-like medium-vessel component**.<br><br>- **Pathology confirms true vasculitis.**<br>- The repeat **skin biopsy** showed **disruption of the internal elastic lamina, fibrinoid necrosis, and neutrophilic infiltration in small-to-medium arteries**, which is classic for **necrotizing arteritis**.<br>- Granulomatous tissue with giant cells was also present, but the decisive finding is **arterial necrotizing vasculitis**.<br>- **Sural nerve biopsy** showing arteriolar inflammation with axonal injury/regeneration supports **vasculitic neuropathy (mononeuritis multiplex)**.<br><br>- **Neurologic involvement is organ-threatening and ischemic.**<br>- MRI-confirmed **acute cerebral infarction** plus biopsy-supported **mononeuritis multiplex** indicates both **central and peripheral nervous system involvement**, consistent with systemic vasculitis affecting arteries.<br><br>- **Other relevant points**<br>- **No eosinophilia or elevated IgE** makes **EGPA less likely**.<br>- **HBV/HCV testing is negative**, which makes **virus-associated PAN less likely**.<br>- **Anemia (Hb 9 g/dL)** is likely multifactorial from inflammation, kidney failure, and possible blood loss related to renal hemorrhage.</td></tr>
<tr><td colspan="2">Phase II – Q6: Reconsidering the differential diagnosis</td></tr>
<tr><td>Baseline</td><td>**6. Updated diagnostic thinking / Working diagnosis**</td></tr>
</table>

| | |
|---|---|
| LLM (Model A) | The differential should now be reprioritized as follows:<br><br>1. **Systemic necrotizing vasculitis with overlap of MPO-ANCA-associated vasculitis and polyarteritis nodosa (PAN-like medium-vessel vasculitis)** — **most likely**<br>- Supports:<br>- MPO-ANCA positivity with RPGN/hematuria/proteinuria → **AAV component**<br>- Renal microaneurysms, renal hemorrhage, splenic ischemia, necrotizing arteritis on skin biopsy, mononeuritis multiplex, cerebral infarction → **PAN-like medium-vessel component**<br>- This mixed phenotype best explains the full picture.<br><br>2. **Microscopic polyangiitis (MPA) alone**<br>- Explains MPO-ANCA positivity, GN, neuropathy, skin lesions, and interstitial lung disease.<br>- However, **multiple renal microaneurysms and hemorrhage are atypical**, making isolated MPA less satisfactory.<br><br>3. **Polyarteritis nodosa alone**<br>- Explains aneurysms, renal hemorrhage, neuropathy, skin/visceral ischemia, and stroke.<br>- But **glomerular hematuria/proteinuria with presumed crescentic GN and MPO-ANCA positivity** argue strongly that PAN alone is incomplete.<br><br>**Working diagnosis:**<br>**MPO-ANCA-associated systemic necrotizing vasculitis with small-vessel (pauci-immune RPGN) and medium-vessel (PAN-like) involvement — an AAV/PAN overlap phenotype.** |
| With Static RAG (Model B) | **6. Updated diagnostic thinking / Working diagnosis**<br><br>The differential has shifted from "MPO-ANCA-associated vasculitis with RPGN, likely MPA" to a **polyarteritis nodosa (PAN)-like medium-vessel necrotizing vasculitis with MPO-ANCA-positive small-vessel involvement**.<br><br>**Most likely working diagnosis:**<br>**Systemic necrotizing vasculitis with features of both PAN and MPO-ANCA-associated vasculitis (polyangiitis overlap syndrome / PAN-like AAV overlap), causing RPGN, renal microaneurysms with renal hemorrhage, mononeuritis multiplex, cutaneous arteritis, cerebral infarction, and pulmonary interstitial involvement.**<br><br>**Why this is now most likely**<br>- **Supports AAV**: MPO-ANCA positivity, rapidly progressive renal injury with hematuria/proteinuria, interstitial lung abnormalities.<br>- **Supports PAN-like medium-vessel vasculitis**: bilateral renal microaneurysms, renal hemorrhage, splenic ischemia, skin/nerve biopsy showing **necrotizing arteritis of muscular arteries**, mononeuritis multiplex, and cerebral infarction.<br>- **Classic PAN alone is less satisfactory** because glomerulonephritis and ANCA positivity are usually absent.<br>- **Classic MPA alone is less satisfactory** because **microaneurysms and medium-vessel necrotizing arteritis with renal hemorrhage** are unusual. |

| | |
|---|---|
| | So the best synthesis is an **overlap phenotype of MPO-AAV and PAN-type arteritis**. |
| With Static RAG and Mult-agent framework (Model C) | ## 6. Updated diagnostic thinking / revised differential diagnosis<br><br>### Most likely diagnosis<br>**MPO-ANCA-positive systemic necrotizing vasculitis with both small-vessel features and PAN-like medium-vessel involvement.**<br><br>### Why the diagnostic thinking changed<br>Initially, the leading diagnosis was **MPO-ANCA-associated vasculitis, likely MPA**, because of:<br>- MPO-ANCA positivity,<br>- rapidly progressive kidney injury,<br>- urinary abnormalities,<br>- skin disease,<br>- neuropathic symptoms,<br>- and lung abnormalities.<br><br>However, the newer data substantially broaden the pattern:<br>- **renal microaneurysms**,<br>- **renal hemorrhage**,<br>- **splenic hypoperfusion/ischemia**,<br>- **skin biopsy-proven necrotizing arteritis of muscular arteries**,<br>- **mononeuritis multiplex**,<br>- and **acute cerebral infarction**<br><br>all strongly support **medium-vessel necrotizing arteritis**, which is **PAN-like**.<br><br>At the same time, several features make **classic PAN alone less likely**:<br>- **MPO-ANCA positivity**,<br>- **hematuria and heavy proteinuria with rapidly progressive renal failure**, suggesting glomerular disease,<br>- and **lung abnormalities**, which are more typical of **AAV** than classic PAN.<br><br>Therefore, the best interpretation is not a formal "overlap syndrome" diagnosis with certainty, but rather a **descriptive working diagnosis**:<br>> **MPO-ANCA-positive systemic necrotizing vasculitis with both small-vessel features and PAN-like medium-vessel involvement.**<br><br>### Revised prioritization of the differential<br>1. **MPO-ANCA-associated vasculitis (most likely MPA phenotype) with atypical PAN-like medium-vessel involvement**<br>2. **Systemic necrotizing arteritis/PAN-like vasculitis with coincidental MPO-ANCA positivity**<br> - Less likely than above because the renal and pulmonary findings favor AAV.<br>3. **Other mimics are now less likely**<br> - **Anti-GBM disease**: anti-GBM negative and vascular aneurysms/skin arteritis argue against it.<br> - **Immune-complex vasculitis/lupus/paraprotein disease**: complements and serologies do not support these.<br> - **EGPA**: no eosinophilia, no IgE elevation, no asthma history provided.<br> - **HBV-associated PAN**: HBV studies are negative. |

| **Phase II – Q7: Treatment planning** | |
|---|---|
| Baseline LLM (Model A) | **7. Treatment plan**<br><br>### Immediate treatment<br>- **High-dose glucocorticoids**<br>- Start **methylprednisolone pulse therapy** (e.g., 500–1000 mg/day for 3 days), then **prednisolone ~1 mg/kg/day** with taper.<br>- Rationale: life- and organ-threatening vasculitis with kidney failure, CNS ischemia, neuropathy, skin/visceral arterial disease.<br><br>- **Add a remission-induction immunosuppressant**<br>- **Cyclophosphamide** is a strong choice here.<br>- Rationale: severe multisystem necrotizing vasculitis with medium-vessel features, renal failure on dialysis, CNS involvement, and visceral arterial lesions. Cyclophosphamide has the most established role in such severe disease.<br>- **Rituximab** is an alternative for severe AAV, but evidence is strongest for small-vessel AAV; with this PAN-like overlap and very high disease severity, cyclophosphamide is particularly reasonable.<br><br>- **Continue hemodialysis as needed**<br>- For ongoing renal failure and metabolic/volume management.<br>- Renal recovery remains possible if active inflammation is controlled, though prognosis is guarded.<br><br>### Additional management considerations<br>- **Do not perform renal biopsy** given documented renal microaneurysms/hemorrhage risk.<br>- **Careful blood pressure control**<br>- Important to reduce risk of further aneurysmal rupture and stroke.<br>- **Management of renal hemorrhage / aneurysms**<br>- Close imaging and hemodynamic monitoring; if bleeding progresses, consider **interventional radiology embolization**.<br>- **Antithrombotic therapy**<br>- Very cautious individualized decision because she has **acute cerebral infarction** but also **active renal hemorrhage/microaneurysms**; bleeding risk may outweigh routine antithrombotic use initially.<br>- **Infection prophylaxis during immunosuppression**<br>- Especially **Pneumocystis jirovecii prophylaxis** unless contraindicated.<br>- **Supportive care**<br>- Gastric and bone protection, anemia management, dialysis-related care, rehabilitation for stroke/neuropathy, and thyroid hormone optimization.<br><br>### Other possible options<br>- **Rituximab instead of cyclophosphamide**<br>- Reasonable if cyclophosphamide toxicity is a major concern; especially attractive in AAV, but less validated for PAN-like medium-vessel disease.<br>- **Plasma exchange**<br>- Current evidence does **not support routine use** for AAV without diffuse alveolar hemorrhage or selected rescue situations. In this case, anti-GBM is negative and pulmonary hemorrhage is not established, so it is **not a standard first choice**.<br>- **PAN-directed strategy**<br>- For classic PAN, steroids ± cyclophosphamide are standard in severe disease; this supports the same practical induction approach here. |

| | |
|---|---|
| | **In summary:**<br>This patient has **organ-threatening systemic necrotizing vasculitis**. The most appropriate plan is **urgent induction with steroid pulses plus cyclophosphamide**, continued **dialysis/supportive care**, and close monitoring for **bleeding, ischemic complications, and infection**. |
| With Static RAG (Model B) | **7. Treatment plan**<br><br>### Initial treatment<br>1. **Immediate remission-induction immunosuppression**<br>- **High-dose glucocorticoids**<br>- e.g., methylprednisolone pulse followed by high-dose oral prednisolone.<br>- **Cyclophosphamide or rituximab** for induction.<br>- In this case, **cyclophosphamide** is particularly reasonable because of **life-threatening multiorgan disease** with medium-vessel necrotizing vasculitis, renal failure, neuropathy, cerebral infarction, and visceral vascular lesions.<br><br>2. **Continue hemodialysis as needed**<br>- for severe renal dysfunction, electrolyte/volume management, and uremic control.<br><br>3. **Careful management of renal hemorrhage and aneurysms**<br>- Close monitoring of hemoglobin, hemodynamics, and flank pain.<br>- **Interventional radiology embolization** should be considered if active bleeding progresses.<br><br>4. **Supportive care**<br>- Blood pressure control.<br>- Infection prophylaxis during immunosuppression (e.g., Pneumocystis prophylaxis unless contraindicated).<br>- Gastric and bone protection with steroids.<br>- Rehabilitation for mononeuritis multiplex and stroke deficits.<br>- Transfusion/iron strategy as clinically needed.<br>- Thyroid hormone replacement adjustment for severe hypothyroidism.<br><br>### Rationale<br>- This is **organ-threatening and potentially life-threatening systemic vasculitis**, so treatment should follow induction strategies used for **severe AAV/systemic necrotizing vasculitis**.<br>- **Steroids alone would likely be insufficient** because of the extent of renal, neurologic, cutaneous, and visceral arterial involvement.<br>- **Cyclophosphamide** has strong evidence and long clinical experience in severe necrotizing vasculitis, especially when rapid control is required.<br><br>### Other options / considerations<br>- **Rituximab** is an evidence-based alternative for severe AAV, especially if cyclophosphamide toxicity is a concern. However, evidence is strongest in classic AAV; in a PAN-like overlap phenotype, many clinicians would still favor cyclophosphamide.<br>- **Plasma exchange** is **not routinely indicated** based on the information given. It may be considered only in specific settings such as severe pulmonary hemorrhage or anti-GBM overlap, neither of which is demonstrated here.<br>- **PAN-directed therapy** (steroids + cyclophosphamide for severe disease) also aligns with this case, which supports the same practical induction approach.<br>- After remission, **maintenance therapy** could include glucocorticoid taper plus an |

| | |
|---|---|
| | agent such as **azathioprine, rituximab, or methotrexate** depending on renal function and final disease classification.<br><br>**In summary:** the patient should be treated as having **severe systemic necrotizing vasculitis with PAN/AAV overlap**, using **urgent high-dose glucocorticoids plus cyclophosphamide (or rituximab as an alternative)**, with concurrent dialysis and close monitoring for hemorrhagic and ischemic vascular complications. |
| With Static RAG and Mult-agent framework (Model C) | ## 7. Treatment plan<br><br>This is **organ-threatening systemic vasculitis** with:<br>- dialysis-requiring kidney failure,<br>- renal arterial microaneurysms and hemorrhage,<br>- CNS infarction,<br>- vasculitic neuropathy,<br>- skin necrotizing arteritis,<br>- and probable pulmonary involvement.<br><br>### Recommended treatment<br>1. **Immediate high-dose glucocorticoids**<br>- Use **IV methylprednisolone pulse therapy**, followed by high-dose oral **prednisolone/prednisone**.<br>- Rationale: rapid suppression of active necrotizing vasculitis is standard in severe AAV and PAN-like arteritis.<br><br>2. **Add remission-induction immunosuppression**<br>- **Cyclophosphamide** is the most practical first choice here because it has established use in **severe AAV** and **necrotizing medium-vessel vasculitis/PAN-like disease**.<br>- Because she is on **hemodialysis**, dosing should be **dialysis-adjusted**, with close monitoring for **cytopenias** and other toxicity.<br>- **Rituximab** is a reasonable alternative if cyclophosphamide toxicity risk is high or contraindications exist, but evidence is stronger for classic AAV than for PAN-like arteritis.<br><br>3. **Continue hemodialysis as clinically needed**<br>- For volume, electrolyte, and uremia control.<br>- Renal recovery is uncertain, but treatment is still indicated because of major extrarenal disease and possible partial renal recovery.<br><br>### Immediate practical management points<br>4. **Exclude active infection before full immunosuppression**<br>- The inflammatory markers are high, but the overall pattern supports vasculitis.<br>- Still, infection should be reassessed clinically and with cultures/imaging as appropriate before or alongside induction therapy, because immunosuppression will be intense.<br><br>5. **Manage hemorrhage risk urgently**<br>- Avoid **renal biopsy** and unnecessary anticoagulation because of **renal microaneurysms and hemorrhage**.<br>- Coordinate with **interventional radiology** in case bleeding worsens or embolization becomes necessary.<br>- Monitor hemoglobin, flank pain, and repeat imaging as needed. |

6. **Coordinate multispecialty care**
  - **Nephrology**: dialysis, renal monitoring, cyclophosphamide dosing.
  - **Neurology**: stroke and mononeuritis management.
  - **Interventional radiology/urology as needed**: bleeding and aneurysm-related complications.
  - **Rheumatology**: induction and disease monitoring.

7. **Stroke management must balance ischemia and bleeding**
  - Acute cerebral infarction usually prompts antithrombotic consideration, but this must be weighed carefully against the known **renal hemorrhage/microaneurysm bleeding risk**.

8. **Supportive and preventive care**
  - **Pneumocystis jirovecii prophylaxis** during high-dose steroids/cyclophosphamide.
  - Gastric and bone protection, glucose monitoring, vaccination review if feasible.
  - Serial CBC, liver tests, and infection surveillance.
  - Rehabilitation for stroke deficits and mononeuritis multiplex.

#### Alternative options
- **Rituximab + glucocorticoids**
  - Reasonable if fertility is irrelevant but toxicity, frailty, infection risk, or marrow concerns favor avoiding cyclophosphamide.
- **Plasma exchange**
  - **Not routinely indicated** from the information provided because **anti-GBM is negative** and there is **no clear diffuse alveolar hemorrhage**.
  - It would become more relevant only if new **alveolar hemorrhage** or **anti-GBM positivity** emerged.

---

### Changes from the initial results

1. **Renal interpretation is now more cautious**
  - I changed from implying proven **RPGN/pauci-immune GN** to:
    > **severe kidney involvement highly suspicious for vasculitic GN with superimposed tubular injury**
  - This reflects the fact that **no renal biopsy was performed**.

2. **Pulmonary interpretation is more conservative**
  - I now state that the CT findings are **suggestive but not specific** for vasculitic lung involvement, rather than treating them as definitive.

3. **Diagnostic labeling is less rigid**
  - I replaced the more confident label of **“AAV-PAN overlap syndrome”** with the more appropriate descriptive diagnosis:
    > **MPO-ANCA-positive systemic necrotizing vasculitis with both small-vessel features and PAN-like medium-vessel involvement**
  - This better reflects current classification uncertainty.

4. **Treatment plan is more individualized**
  - I added:
    - **infection exclusion before full immunosuppression**,

| | |
|---|---|
| | - **dialysis-adjusted cyclophosphamide dosing**,<br>- **close cytopenia monitoring**,<br>- and **urgent hemorrhage-risk coordination** with nephrology/neurology/interventional radiology.<br><br>Overall, the revised interpretation remains that this is a **severe, organ-threatening systemic necrotizing vasculitis**, but it now states the **diagnostic uncertainty and management priorities more precisely**. |
| **Phase III – Q8: Evaluation of treatment** | |
| Baseline LLM (Model A) | ### 8. Monitoring effectiveness, safety, risks, and countermeasures<br><br>**Effectiveness monitoring**<br>- **Clinical vasculitis activity**: regularly assess purpura, skin nodules, neuropathy/mononeuritis symptoms, new focal neurologic deficits, headache, flank pain, constitutional symptoms, and overall **BVAS**.<br>- **Renal response**: serial **serum creatinine/eGFR**, urine output, need for dialysis, **urinalysis** (hematuria, proteinuria, casts), and urine protein quantification.<br>- **Inflammation/serology**: follow **CRP** and **MPO-ANCA** trends. These support response but should be interpreted with clinical findings.<br>- **Organ-specific follow-up**:<br>- **Neurologic**: repeated neurological examination; urgent brain imaging if new deficits occur.<br>- **Pulmonary**: symptoms, oxygenation, chest imaging as needed.<br>- **Medium-vessel disease**: monitor for signs of new infarction/hemorrhage (abdominal pain, flank pain, stroke symptoms). Repeat vascular imaging can be useful if renal function allows, but risk of contrast nephrotoxicity must be weighed.<br><br>**Safety monitoring and major risks**<br>1. **Serious infection** from steroids, cyclophosphamide, and overall immunosuppression<br>- Monitor: temperature, symptoms, CBC with differential, CRP, cultures when indicated, fungal markers as appropriate.<br>- Countermeasures: prompt microbiologic workup, early empiric antimicrobials when infection is suspected, antifungal therapy for candidemia, **G-CSF** for febrile neutropenia, and reduce/hold cytotoxic therapy when necessary.<br><br>2. **Myelosuppression / febrile neutropenia from cyclophosphamide**<br>- Monitor: frequent **CBC**.<br>- Countermeasures: dose adjustment, treatment delay if cytopenic, G-CSF when needed, infection precautions.<br><br>3. **Relapse or incomplete control of vasculitis**<br>- Monitor: rising CRP, worsening renal function, recurrent purpura, new neuropathy, new ischemic events, urinary abnormalities, BVAS.<br>- Countermeasures: reassess disease phenotype (small- vs medium-vessel activity), intensify therapy when relapse is likely; in this case, **avacopan** helped avoid steroid escalation, and additional **IVCY** was used after recurrent cerebral infarction.<br><br>4. **Glucocorticoid toxicity**<br>- Risks: infection, hyperglycemia, muscle weakness, psychiatric effects, osteoporosis, GI toxicity. |

| | |
|---|---|
| | - Monitor: glucose, mental status, muscle function, GI symptoms, bone protection needs.<br>- Countermeasures: taper steroids as soon as disease control permits; use steroid-sparing therapy such as avacopan when appropriate.<br><br>5. **Hemorrhage and ischemic complications from medium-vessel vasculitis**<br>- Risks: renal hemorrhage, splenic infarction, cerebral infarction.<br>- Monitor: flank/abdominal pain, hemoglobin, hemodynamics, neurologic exams; targeted imaging if symptoms change.<br>- Countermeasures: urgent reassessment, supportive care, and escalation of vasculitis treatment; carefully balance any antithrombotic decisions against bleeding risk.<br><br>6. **Renal function decline**<br>- Monitor: creatinine, electrolytes, fluid status, dialysis need.<br>- Countermeasures: avoid nephrotoxins, adjust drug doses for kidney function, continue/withdraw dialysis based on recovery.<br><br>7. **Avacopan adverse effects**<br>- Monitor: liver enzymes, infection events, general tolerability.<br>- Countermeasures: laboratory follow-up and discontinuation if significant toxicity occurs. |
| With Static RAG (Model B) | ## 8. Monitoring of treatment effectiveness, safety, risks, and countermeasures<br><br>### A. Monitoring effectiveness<br>- **Clinical symptoms/signs**<br>- Follow **purpura, skin nodules, neuropathic symptoms, focal neurologic deficits, headache, flank pain, and constitutional symptoms**.<br>- Use **BVAS serially** to assess overall vasculitis activity.<br>- **Inflammatory activity**<br>- Monitor **CRP and WBC trend** regularly.<br>- **MPO-ANCA** can be followed as a supportive marker, but treatment decisions should not rely on ANCA alone.<br>- **Renal response**<br>- Track **serum creatinine/eGFR**, urine volume, and dialysis requirement.<br>- Repeated **urinalysis** with **proteinuria and hematuria** assessment is essential, especially for small-vessel activity.<br>- **Medium-vessel disease surveillance**<br>- Watch for new **stroke symptoms, abdominal/flank pain, or organ ischemia/bleeding**.<br>- Imaging should be reconsidered if relapse is suspected, balancing benefit against renal risk.<br>- **Neurologic follow-up**<br>- Reassess for progression or recurrence of **mononeuritis multiplex** or cerebral infarction.<br><br>### B. Main safety risks and concrete countermeasures<br>1. **Serious infection**<br>- Major risk because she already developed **candidemia and febrile neutropenia**.<br>- Monitor: **fever, cultures as indicated, CBC, CRP, procalcitonin**, fungal markers when needed.<br>- Countermeasures:<br>- Minimize steroid exposure when possible.<br>- Careful timing/dose of **IVCY**. |

| | |
|---|---|
| | - Prompt antimicrobial/antifungal treatment if infection is suspected.<br>- Consider standard infection prophylaxis during immunosuppression according to institutional practice.<br>- Use **G-CSF** for severe neutropenia as previously required.<br><br>2. **Myelosuppression from cyclophosphamide**<br>- Monitor: **frequent CBC with differential**.<br>- Countermeasures:<br>- Adjust or delay IVCY based on blood counts and clinical status.<br>- Treat neutropenia promptly; avoid further cytotoxic therapy during uncontrolled infection.<br><br>3. **Relapse of vasculitis during PSL taper**<br>- Already occurred with recurrence of purpura and later inflammatory/renal worsening.<br>- Monitor: **symptoms, CRP, renal function, urinalysis, neurologic findings**.<br>- Countermeasures:<br>- Do not judge relapse only by ANCA.<br>- If relapse is likely and infection is excluded, intensify therapy appropriately.<br>- In this case, **avacopan** was a steroid-sparing option to reduce infection risk.<br><br>4. **Progression of medium-vessel vasculitis**<br>- Risks: **cerebral infarction, splenic infarction, renal ischemia/hemorrhage**.<br>- Monitor: new focal deficits, abdominal pain, flank pain, falling hemoglobin.<br>- Countermeasures:<br>- Urgent reassessment and escalation of vasculitis therapy if active disease is suspected.<br>- Repeat vascular imaging if clinically necessary and feasible.<br><br>5. **Renal deterioration**<br>- Monitor: **Cr/eGFR, electrolytes, urine findings, fluid balance**.<br>- Countermeasures:<br>- Avoid nephrotoxins and unnecessary contrast exposure.<br>- Resume dialysis if needed.<br>- Adjust drug doses to kidney function.<br><br>6. **Glucocorticoid toxicity**<br>- Risks: **infection, hyperglycemia, muscle weakness, psychiatric effects, osteoporosis, GI complications**.<br>- Monitor clinically and with routine labs.<br>- Countermeasures:<br>- Taper steroids as disease control allows.<br>- Use bone and GI protection as appropriate.<br>- Prefer steroid-sparing strategy when possible, as done with avacopan. |
| With Static RAG and Mult-agent framework (Model C) | ## 8. Monitoring of treatment effectiveness, safety, risks, and countermeasures<br><br>### A. Monitoring treatment effectiveness<br>Treatment response in this case should be judged **primarily by organ-based clinical findings together with inflammatory and renal trends**, not by MPO-ANCA alone.<br><br>1. **Clinical disease activity**<br>- Follow **BVAS**, recurrence of **purpura**, **skin nodules**, constitutional symptoms, and changes in neuropathy or mood/functional status. |

- Because this patient had both **small-vessel manifestations** and **medium-vessel manifestations**, daily symptom review is essential during the acute phase.

2. **Neurologic monitoring should be prioritized**
  - Perform **frequent neurologic examinations** for new focal deficits, headache, gait change, dysarthria, facial weakness, or worsening mononeuritis multiplex.
  - This is especially important because she had **multiple cerebral infarctions initially** and a **recurrent mild cerebral infarction on day 88**.
  - **Immediate brain imaging** is required for any new neurologic change.

3. **Inflammatory markers and MPO-ANCA**
  - Monitor **CRP serially** and follow **MPO-ANCA trends** as supportive data.
  - In this case, both improved after induction therapy and again after avacopan-based control.
  - However, **ANCA changes should not be used alone** to define remission or relapse.

4. **Renal response**
  - Serial **serum creatinine/eGFR**, urine output, dialysis requirement, and **urinalysis including hematuria and proteinuria**.
  - Monitor **proteinuria/hematuria** for recurrence of small-vessel renal activity.
  - In this patient, successful **dialysis withdrawal on day 66** and discharge with creatinine around **2.5 mg/dL** indicate meaningful renal recovery.

5. **Monitoring for medium-vessel disease activity**
  - Follow for **new ischemic or hemorrhagic symptoms**, especially:
    - neurologic events,
    - abdominal pain,
    - flank pain,
    - skin nodules,
    - signs suggesting splenic or renal vascular events.
  - The later rise in **CRP** with worsening renal function but **no hematuria, no purpura, and only mild proteinuria** was interpreted as possible **medium-vessel-predominant relapse**.

6. **Practical monitoring schedule**
  - During acute hospitalization/intensive treatment:
    - **CBC with differential, creatinine, electrolytes, CRP**: at least several times weekly, and more often if unstable.
    - **Urinalysis/proteinuria**: regularly, such as weekly or more often if renal status changes.
    - **Focused neurologic reassessment**: daily in the acute phase.
    - **Liver function tests**: regularly after starting avacopan.
  - The exact frequency should be adjusted to clinical stability.

#### B. Monitoring safety, major risks, and countermeasures
1. **Serious infection**
  - This became a major proven complication: **candidemia** and **febrile neutropenia**.
  - Monitoring:
    - temperature and focused symptom review,
    - physical examination,
    - **CBC with differential**,
    - CRP,

| | |
|---|---|
| | - cultures and fungal tests when indicated.<br>- Countermeasures:<br>- rapid evaluation of any fever,<br>- prompt **antibacterial and antifungal therapy** when infection is suspected,<br>- **G-CSF** for neutropenia as needed,<br>- minimize glucocorticoid exposure when possible,<br>- use steroid-sparing therapy such as **avacopan** when appropriate.<br><br>2. **Cyclophosphamide toxicity**<br>- Main concerns: **neutropenia/cytopenia**, infection, and treatment interruption.<br>- Monitoring:<br>- regular **CBC with differential**,<br>- renal function for dose adjustment,<br>- overall treatment tolerance.<br>- Countermeasures:<br>- reduce or delay cyclophosphamide if cytopenia develops,<br>- support with **G-CSF** when needed,<br>- resume cautiously only after recovery.<br><br>3. **Relapse during steroid taper**<br>- This occurred once with **recurrent purpura**, and later relapse was suspected based on **CRP rise and renal deterioration** after infection improved.<br>- Monitoring:<br>- symptoms,<br>- CRP,<br>- urinalysis/proteinuria,<br>- renal function,<br>- neurologic findings.<br>- Countermeasures:<br>- temporary steroid reinforcement if clearly needed,<br>- do not rely only on steroid escalation,<br>- add or intensify remission therapy when relapse is likely, as was done with **avacopan** and later additional **IVCY**.<br><br>4. **Renal hemorrhage / aneurysm-related complications**<br>- Ongoing risk because of **renal microaneurysms** and prior **right renal hemorrhage**.<br>- Monitoring:<br>- **flank pain**,<br>- **hemoglobin**,<br>- **hemodynamics**,<br>- renal function.<br>- Countermeasures:<br>- avoid unnecessary invasive procedures,<br>- use contrast studies only when the expected benefit justifies renal and bleeding risk,<br>- obtain urgent imaging/intervention if bleeding is suspected.<br><br>5. **New ischemic events from medium-vessel vasculitis**<br>- There is continued risk of **recurrent cerebral infarction** and other ischemic events.<br>- Monitoring:<br>- close neurologic surveillance,<br>- review for abdominal or flank ischemic symptoms. |

<table>
<tr><td></td><td>- Countermeasures:<br>- urgent reassessment of disease activity,<br>- exclusion of infection and bleeding,<br>- escalation of immunosuppression if relapse is considered likely.<br><br>6. **Steroid toxicity**<br>- Risks include infection, hyperglycemia, psychiatric effects, myopathy, osteoporosis, and GI complications.<br>- Monitoring:<br>- glucose,<br>- mental status,<br>- muscle strength,<br>- GI symptoms,<br>- overall adverse effects.<br>- Countermeasures:<br>- taper steroids as disease control allows,<br>- use steroid-sparing therapy,<br>- standard preventive measures as appropriate.<br><br>7. **Avacopan safety**<br>- Monitoring:<br>- **liver function tests**,<br>- infection events,<br>- overall tolerance and clinical response.<br>- Countermeasure:<br>- reassess or discontinue if significant adverse effects or unexplained deterioration occur.</td></tr>
<tr><td colspan="2">Phase III – Q9: Formulating plans for worsening conditions</td></tr>
<tr><td>Baseline LLM (Model A)</td><td>### 9. Reassessment and action plan if the course worsens or changes unexpectedly<br><br>**Step 1: Rapidly determine whether this is**<br>- **active vasculitis/relapse**,<br>- **infection**,<br>- **drug toxicity**, or<br>- **a new vascular complication** (hemorrhage/infarction).<br><br>**How to think**<br>- A rise in **CRP** or worsening renal function alone is nonspecific. In an immunosuppressed patient, infection must be excluded first, especially after prior **candidemia** and **febrile neutropenia**.<br>- The pattern of recurrence helps:<br>- **Hematuria, proteinuria, purpura, active urinary sediment, neuropathy** → suggests **small-vessel relapse**.<br>- **New stroke, flank/abdominal pain, infarction/hemorrhage, rising inflammation without active urinary sediment or purpura** → suggests **medium-vessel activity**.<br>- Also consider treatment toxicity:<br>- Cytopenia → cyclophosphamide toxicity<br>- Fever with neutropenia → infection until proven otherwise<br>- Worsening kidney function → relapse, sepsis, drug effect, or hemodynamic causes<br><br>**Action plan**<br>1. **Immediate bedside reassessment**</td></tr>
</table>

| | |
|---|---|
| | - Vitals, hemodynamic status, full neurologic exam, skin exam, volume status, and symptom review.<br><br>2. **Urgent laboratory review**<br>- CBC with differential, creatinine/electrolytes, CRP, urinalysis, urine protein, cultures if febrile, and selected infection markers as clinically indicated.<br><br>3. **Targeted imaging based on symptoms**<br>- Brain imaging for new neurologic signs.<br>- Chest imaging for respiratory change.<br>- Abdominal/renal imaging for flank pain, anemia, or suspected hemorrhage/infarction.<br><br>4. **Temporarily modify treatment according to the leading concern**<br>- If **infection is suspected**: start empiric antimicrobials promptly, hold/reduce cytotoxic immunosuppression as needed, and support with G-CSF if neutropenic.<br>- If **vasculitis relapse is more likely** after excluding infection: intensify remission therapy while minimizing excess steroid toxicity; options include steroid pulse, adding/repeating cyclophosphamide, or using a steroid-sparing adjunct such as **avacopan**.<br>- If **vascular ischemic/hemorrhagic complication** occurs: urgent specialist evaluation and supportive management, while treating underlying vasculitis.<br><br>5. **Revise the working diagnosis dynamically**<br>- This case shows that AAV can behave as a **mixed small- and medium-vessel disease**. If usual small-vessel markers improve but inflammation persists, reassess whether **medium-vessel vasculitis** is driving the course.<br><br>6. **Rebalance risk continuously**<br>- The key clinical task is to maintain remission **without provoking fatal infection or treatment toxicity**. Therefore, each deterioration should trigger a structured re-evaluation rather than automatic steroid escalation. |
| With Static RAG (Model B) | ## 9. If the condition does not improve as expected or suddenly changes<br><br>### A. Reassess the cause quickly and broadly<br>If inflammation rises, renal function worsens, or new symptoms appear, first reconsider:<br>1. **Relapse/progression of vasculitis**<br>2. **Infection related to immunosuppression**<br>3. **Drug toxicity**, especially cytopenia from cyclophosphamide<br>4. **New vascular event** such as cerebral infarction or renal hemorrhage<br><br>### B. Practical action plan<br>- **Immediate focused reassessment**<br>- Repeat **history and physical examination**, looking for infection signs and new organ ischemia.<br>- Check **CBC, CRP, renal panel, urinalysis**, and targeted microbiology.<br>- Use **procalcitonin, cultures, fungal markers, CMV testing**, etc., when infection is a concern.<br>- **Differentiate relapse from infection**<br>- This is critical because treatment directions are opposite.<br>- In this case, absence of infectious focus and negative microbiologic/fungal testing supported relapse rather than infection. |

| | |
|---|---|
|  | - **Identify which vessel compartment is active**<br> - **Small-vessel relapse** is suggested by recurrent hematuria, proteinuria, purpura, active urinary sediment.<br> - **Medium-vessel relapse** is suggested by rising CRP with ischemic/neurologic symptoms, organ infarction, hemorrhage, or pain, even without active urine sediment or purpura.<br>- **Revise treatment flexibly**<br> - If **infection**: reduce/hold immunosuppression as feasible and treat infection aggressively.<br> - If **vasculitis relapse**: intensify remission-induction therapy while minimizing infection risk.<br> - In this patient, because further steroid escalation was risky after severe infection, adding **avacopan** was a rational steroid-sparing strategy.<br> - If a new serious medium-vessel event occurs, as with the later mild cerebral infarction, consider **additional cyclophosphamide** or other re-induction therapy.<br><br>### C. Ongoing principle<br>Management should be **dynamic**: repeatedly reassess whether the dominant problem is **active vasculitis, infection, treatment toxicity, or vascular complication**, and then modify immunosuppression and supportive care accordingly. |
| With Static RAG and Mult-agent framework (Model C) | ## 9. Reassessment and flexible revision of diagnosis/treatment when the condition worsens<br><br>### A. Immediate framework<br>If the patient does not improve or worsens suddenly, rapidly distinguish among:<br>1. **Active vasculitis / relapse**<br>2. **Infection related to immunosuppression**<br>3. **Drug toxicity / treatment complication**<br>4. **New vascular event or bleeding**<br><br>This distinction is critical because management differs in opposite directions.<br><br>### B. Immediate reassessment approach<br>1. **Reassess the pattern of organ involvement**<br> - **Purpura, hematuria, increased proteinuria, worsening neuropathy** suggest recurrence of **small-vessel activity**.<br> - **New focal neurologic symptoms, abdominal pain, flank pain, infarction, hemorrhage, or rising CRP without urinary relapse markers** suggest **medium-vessel activity**.<br> - Because this patient has mixed-vessel disease, deterioration should not be judged from urinalysis alone.<br><br>2. **Prioritize neurologic evaluation**<br> - Any new neurologic deficit requires **immediate neurologic examination and urgent brain imaging** to assess for new infarction or progression.<br> - This was especially important here because cerebral infarction recurred despite treatment.<br><br>3. **Check urgent objective data**<br> - Obtain:<br> - **CBC with differential**,<br> - CRP, |

- renal function,
  - urinalysis/proteinuria,
  - hemoglobin,
  - cultures and infection tests as indicated.
- If flank pain or anemia worsens, reassess for **renal bleeding**.
- If focal symptoms occur, obtain urgent imaging targeted to the involved organ.

4. **Exclude infection carefully before escalating immunosuppression**
- This is essential because she already developed **candidemia and febrile neutropenia**.
- In the later course, relapse was favored only after there was:
  - no clear infectious focus,
  - negative **procalcitonin**,
  - negative **bacterial cultures**,
  - negative **β-D-glucan**,
  - and negative **CMV antigenemia**.
- The same cautious approach should be repeated whenever relapse is suspected.

5. **Consider whether the active process is mainly small-vessel or medium-vessel**
- Recurrence of **purpura/hematuria/proteinuria** supports **small-vessel relapse**.
- Rising **CRP**, renal dysfunction, or new ischemic events **without hematuria or purpura** may support **medium-vessel-predominant relapse**.
- However, this inference is partly clinical, because repeat vascular imaging was not performed.

6. **Use vascular imaging selectively when feasible**
- If medium-vessel relapse, bleeding, or new infarction is suspected, consider **noninvasive or carefully selected imaging** to support the diagnosis.
- In this case, repeat contrast CT was deferred to protect renal function, so treatment decisions had to rely mainly on **clinical pattern, labs, and infection exclusion**.

#### C. How to revise treatment
1. **If relapse is likely**
- Intensify anti-vasculitis therapy according to severity:
  - temporary steroid intensification for clear inflammatory recurrence,
  - add or repeat **cyclophosphamide** for organ-threatening relapse,
  - use **avacopan** as a steroid-sparing adjunct when infection risk from steroid escalation is high.
- In this patient, this logic supported use of **avacopan on day 72** and additional **IVCY** after recurrent cerebral infarction.

2. **If infection is likely**
- Hold or reduce immunosuppression as feasible,
- obtain cultures and start prompt targeted antimicrobial therapy,
- support neutropenia when present.

3. **If bleeding or infarction is suspected**
- Obtain urgent imaging and specialist input,
- reassess both vasculitis control and supportive/interventional needs.

#### D. Key principle
Because this patient has **ANCA-associated vasculitis with medium-sized vessel involvement**, response and relapse should be assessed mainly by:

| | |
|---|---|
| | - **clinical organ-based findings**,<br>- **neurologic status**,<br>- **renal and inflammatory trends**,<br>- **careful exclusion of infection**,<br>- and, when feasible, **targeted imaging**.<br><br>MPO-ANCA trends are supportive, but they should **not** be used alone to define treatment success or failure.<br><br>---<br><br>## Changes from the initial results<br>- I **more explicitly prioritized neurologic surveillance**, including **daily neurologic reassessment in the acute phase** and **urgent brain imaging for any new deficits**, because of the recurrent cerebral infarctions.<br>- I added **practical monitoring items and timing**, including regular **CBC, creatinine, urinalysis/proteinuria, CRP, and liver function tests** after starting avacopan.<br>- I strengthened monitoring for **renal hemorrhage**, explicitly adding **hemoglobin, hemodynamics, and flank pain** because of known renal microaneurysms and prior hemorrhage.<br>- I clarified that treatment response should be judged **mainly by organ-based clinical findings plus inflammatory and renal trends**, **not by MPO-ANCA alone**.<br>- I made the later "medium-vessel relapse" interpretation **more cautious**, noting that it was clinically inferred and that **repeat vascular imaging was not performed**, so reassessment should include renewed exclusion of infection, bleeding, and stroke progression.<br>- I added that **selected noninvasive or carefully chosen vascular imaging** may be considered when feasible to support suspected medium-vessel relapse while balancing renal risk. |

| **Case 8: Hypernatremic chloride-depletion metabolic alkalosis successfully treated with high cation-gap amino acids: a case report** [16] | |
|---|---|
| **Phase I – Q1: Summarizing medical problems** | |
| Baseline LLM (Model A) | 1. **Problem representation**<br>42-year-old Japanese man with a history of atopic dermatitis presenting with **1 week of persistent abdominal pain and abdominal bloating/distension**. The key clinical issue is **subacute abdominal pain with bloating**, which raises concern for **gastrointestinal, inflammatory, obstructive, or allergic/eosinophilic causes**. His atopic background is notable because it increases the possibility of **eosinophilic gastrointestinal disease**, although more common causes must still be considered first. |
| With Static RAG (Model B) | 1. **Problem representation**<br>A **42-year-old Japanese man** with a history of **atopic dermatitis** presents with **1 week of persistent abdominal pain and abdominal bloating/distension**.<br>Key points are: **subacute course**, **abdominal pain plus bloating**, and a background of **atopic disease**, which raises the possibility of not only common gastrointestinal disorders but also **allergic/eosinophilic gastrointestinal disease**. |
| With Static RAG and | ## 1. Problem representation<br>A **42-year-old man** with **1 week of persistent abdominal pain and abdominal |

| | |
|---|---|
| Mult-agent framework (Model C) | distension/bloating** and a history of **atopic dermatitis** presents with a **subacute, nonspecific abdominal syndrome**. The atopic background is a **clue** that raises consideration of **eosinophilic gastrointestinal disease**, but it should **not outweigh more common or urgent causes** such as **bowel obstruction, ileus, constipation, inflammatory disease, pancreatitis, or ascites-related disorders**. |
| **Phase I – Q2: Differential diagnoses and rationale** | |
| Baseline LLM (Model A) | 2. **Differential diagnosis**<br>1. **Eosinophilic gastroenteritis / eosinophilic GI disease** — **most likely**<br>- Reasoning: Abdominal pain and bloating over days to a week fit this disorder, and the history of atopic dermatitis is an important clue because eosinophilic GI disease is associated with allergic/atopic conditions. It can cause bowel wall edema, ascites, nausea, diarrhea, or obstructive symptoms.<br>2. **Bowel obstruction or ileus**<br>- Reasoning: Bloating plus abdominal pain strongly suggests impaired bowel transit. This must be considered even without more details because it can become urgent, especially if there is vomiting, constipation/obstipation, or progressive distension.<br><br>3. **Inflammatory bowel or other enterocolitis (including infectious enteritis)**<br>- Reasoning: Subacute abdominal pain can reflect intestinal inflammation. If accompanied by diarrhea, fever, or weight loss, this would become more likely. Some inflammatory conditions can also present with abdominal distension.<br><br>**Must not be missed:** **Bowel obstruction/ischemia or other surgical abdomen**<br>- Because abdominal pain with distension may indicate a condition requiring urgent intervention. Red flags would include severe or worsening pain, vomiting, peritoneal signs, fever, tachycardia, absent stool/flatus, or metabolic/lactate abnormalities. |
| With Static RAG (Model B) | 2. **Differential diagnosis**<br>**Top 3 differential diagnoses:**<br>- **Bowel obstruction / subileus**<br>- **Eosinophilic gastroenteritis**<br>- **Ascites-related disorder or intra-abdominal inflammatory disease** (e.g., peritonitis, pancreatitis, malignancy-related process)<br><br>**Most likely:** **Eosinophilic gastroenteritis**<br>**Reasoning:** The combination of **abdominal pain, bloating, and atopic history** is suggestive. Eosinophilic GI disease can present with abdominal pain, distension, nausea, diarrhea, bowel wall edema, or eosinophilic ascites.<br><br>**Must not miss:** **Bowel obstruction / surgical abdomen**<br>**Reasoning:** Persistent abdominal pain and bloating for 1 week may indicate mechanical obstruction, ileus, ischemia, perforation, or other urgent causes requiring prompt diagnosis.<br><br>**Why these are considered:**<br>- **Bowel obstruction / subileus:** common and important cause of pain with distension; may present with vomiting, constipation, and tympanic abdomen.<br>- **Eosinophilic gastroenteritis:** supported by atopic background; can mimic obstruction or ascites.<br>- **Inflammatory/ascitic intra-abdominal disease:** abdominal distension may reflect fluid accumulation, inflammation, or organ disease. |

| | |
|---|---|
| With Static RAG and Mult-agent framework (Model C) | ## 2. Prioritized differential diagnosis<br>### Most important initial differential<br>1. **Bowel obstruction or ileus**<br>- Abdominal pain plus bloating/distension for a week is a classic warning pattern.<br>- This must be prioritized because it may require urgent intervention.<br>- Mechanical obstruction, severe constipation, or functional ileus are all possible.<br><br>2. **Constipation / fecal impaction**<br>- A common cause of abdominal pain and bloating over days to a week.<br>- Can mimic partial obstruction.<br><br>3. **Eosinophilic gastroenteritis / eosinophilic GI disease**<br>- Atopic dermatitis increases suspicion for allergic/eosinophilic disease.<br>- Can cause abdominal pain, distension, bowel wall edema, diarrhea, nausea, or ascites.<br>- Important consideration, but not yet the leading diagnosis based on the limited data.<br><br>### Other reasonable differentials<br>4. **Infectious or inflammatory enterocolitis / inflammatory bowel disease**<br>- More likely if diarrhea, fever, or blood in stool is present.<br><br>5. **Pancreatitis**<br>- Can cause abdominal pain, abdominal distension, ileus, and nausea/vomiting.<br><br>6. **Peptic ulcer disease / gastritis / dyspepsia**<br>- May cause abdominal pain and fullness, though marked bloating is less specific.<br><br>7. **Ascites-related disorder or intra-abdominal malignancy**<br>- Less likely from the limited information, but persistent distension warrants consideration, especially if there is weight loss or systemic illness.<br><br>### Must not miss<br>- **Strangulated bowel obstruction / bowel ischemia**<br>- **Perforation / peritonitis**<br>- **Severe pancreatitis** |
| **Phase I – Q3: Necessary physical examinations and reasons** | |
| Baseline LLM (Model A) | 3. **Additional history and physical examination**<br>- **History**<br>- Character of pain: onset, location, severity, colicky vs constant, progression<br>- Helps distinguish obstruction, inflammatory disease, or peritonitis.<br>- GI symptoms: nausea, vomiting, diarrhea, constipation, obstipation, inability to pass flatus, early satiety<br>- Clarifies obstruction vs enteritis vs functional causes.<br>- Stool features: blood, melena, mucus<br>- Suggests inflammatory, ischemic, or infectious disease.<br>- Fever, weight loss, anorexia<br>- Supports inflammatory/infectious/malignant causes.<br>- Food triggers, history of allergies/asthma, prior similar episodes<br>- Supports eosinophilic GI disease.<br>- Medication history, including NSAIDs and antibiotics<br>- NSAIDs may cause GI injury; antibiotics may suggest C. difficile or altered bowel symptoms. |

| | |
|---|---|
| | - Surgical history or hernia history<br>- Important for mechanical obstruction risk.<br>- Travel, raw food, parasite exposure<br>- Relevant to infectious/parasitic eosinophilia.<br>- **Physical examination**<br>- Vital signs<br>- Instability would suggest urgent pathology.<br>- Abdominal inspection for distension; auscultation for high-pitched or absent bowel sounds<br>- Helps evaluate obstruction/ileus.<br>- Palpation for localized tenderness, guarding, rebound<br>- Assesses for peritonitis or focal inflammation.<br>- Percussion for tympany and ascites<br>- Distinguishes gas distension vs fluid accumulation.<br>- Check for hernias<br>- Potential cause of obstruction.<br>- Skin exam for active atopic disease<br>- Supports allergic background but is not diagnostic. |
| With Static RAG (Model B) | 3. **Additional history and physical examination**<br>**History to ask:**<br>- **Pain characteristics**: location, severity, colicky vs constant, relation to meals<br>- To distinguish obstruction, inflammatory disease, pancreatitis, peptic disease.<br>- **Associated GI symptoms**: nausea, vomiting, diarrhea, constipation, obstipation, blood in stool<br>- To assess for obstruction, gastroenteritis, inflammatory bowel disease, bleeding.<br>- **Fever and systemic symptoms**: fever, weight loss, anorexia<br>- To evaluate infection, malignancy, inflammatory disease.<br>- **Allergic history**: asthma, allergic rhinitis, food allergy, recent dietary trigger<br>- To support eosinophilic GI disease.<br>- **Medication/exposure history**: NSAIDs, antibiotics, herbal drugs, new medications, travel, raw food, parasitic exposure<br>- To evaluate drug-induced disease, infection, parasites.<br>- **Past surgical history**<br>- Adhesive bowel obstruction risk.<br>- **Alcohol history / gallstone history**<br>- To assess pancreatitis.<br>- **Bowel habit change**<br>- To clarify obstruction, malignancy, inflammatory conditions.<br><br>**Physical examination to perform:**<br>- **Vital signs**<br>- To assess instability, fever, sepsis.<br>- **Abdominal inspection, auscultation, percussion, palpation**<br>- Distension, bowel sounds, focal tenderness, guarding, rebound, mass, shifting dullness.<br>- **Signs of peritonitis**<br>- To detect surgical abdomen.<br>- **Assessment for ascites**<br>- Shifting dullness/fluid wave.<br>- **Skin examination**<br>- Confirm atopic dermatitis; look for other allergic manifestations.<br>- **Rectal examination if indicated**<br>- To assess stool burden, occult blood, distal obstruction. |

| | |
|---|---|
| With Static RAG and Mult-agent framework (Model C) | ## 3. Additional history and physical examination<br>### Focused history<br>- **Pain details**: onset, location, severity, constant vs colicky, radiation<br> - Helps distinguish obstruction, pancreatitis, peptic disease, and peritonitis.<br>- **Bowel function**: last bowel movement, last passage of flatus, constipation, obstipation, diarrhea, stool caliber change<br> - Critical for obstruction, ileus, and constipation.<br>- **Nausea and vomiting**: presence, frequency, bilious or feculent<br> - Supports obstruction and helps gauge severity.<br>- **Previous similar episodes**<br> - Suggests recurrent obstruction, chronic constipation, or eosinophilic GI disease.<br>- **Prior abdominal surgery**<br> - Major risk factor for adhesive small-bowel obstruction.<br>- **Hernia history**<br> - Important mechanical cause of obstruction.<br>- **Fever, anorexia, weight loss, hematochezia, melena**<br> - Suggest infection, inflammation, ischemia, or malignancy.<br>- **Allergic history**: asthma, allergic rhinitis, food-related symptoms<br> - Supports eosinophilic GI disease.<br>- **Medication history**<br> - Opioids and anticholinergics can cause ileus/constipation; NSAIDs may suggest ulcer disease.<br><br>### Focused physical examination<br>- **Vital signs**<br> - Fever, tachycardia, or hypotension may indicate serious intra-abdominal disease.<br>- **Abdominal inspection**<br> - Degree of distension, scars, visible peristalsis.<br>- **Auscultation**<br> - High-pitched bowel sounds may suggest obstruction; reduced sounds may suggest ileus.<br>- **Palpation**<br> - Focal tenderness, masses, guarding, rebound for peritoneal irritation.<br>- **Percussion**<br> - Tympany suggests gaseous distension; shifting dullness suggests ascites.<br>- **Hernia examination**<br> - Inguinal, femoral, and umbilical hernias.<br>- **Skin/systemic exam**<br> - Confirm atopic dermatitis and look for signs of systemic illness. |
| **Phase I – Q4: Plan for investigations/tests** | |
| Baseline LLM (Model A) | 4. **Testing plan**<br> - **CBC with differential**<br> - Purpose: look for leukocytosis, anemia, and especially **eosinophilia**.<br> - Expected findings: eosinophilia would support eosinophilic GI disease, though absence does not exclude it.<br> - **CRP and basic metabolic panel**<br> - Purpose: assess inflammation, dehydration, electrolyte disturbance, renal function.<br> - Expected findings: inflammatory markers may be elevated in enteritis/inflammation; electrolyte abnormalities may occur with vomiting or obstruction.<br> - **Liver enzymes, bilirubin, amylase/lipase** |

| | |
|---|---|
| | - Purpose: exclude hepatobiliary or pancreatic causes of abdominal pain/bloating.<br>- **Abdominal CT with contrast**<br>- Purpose: most useful test to evaluate obstruction, bowel wall thickening, ascites, ischemic signs, and other intra-abdominal pathology.<br>- Expected findings:<br>- Obstruction: dilated bowel loops/transition point.<br>- Eosinophilic gastroenteritis: bowel wall thickening, edema, possible ascites.<br>- Other inflammatory disease: segmental thickening or inflammatory changes.<br>- **Stool studies if diarrhea is present** (culture, ova/parasites)<br>- Purpose: exclude infectious/parasitic causes, especially if eosinophilia is present.<br>- **Endoscopy with biopsy** if imaging/labs suggest eosinophilic or inflammatory GI disease<br>- Purpose: establish definitive diagnosis of eosinophilic gastroenteritis or other mucosal disease.<br>- Expected findings: eosinophilic infiltration on biopsy, though endoscopic appearance may be nonspecific.<br>- **Paracentesis** if ascites is present<br>- Purpose: analyze fluid; eosinophil-rich ascites can support subserosal eosinophilic gastroenteritis.<br><br>Overall, the leading consideration is **eosinophilic gastroenteritis** given the **atopic history plus subacute abdominal pain and bloating**, but **bowel obstruction/surgical abdomen must be excluded first**. |
| With Static RAG (Model B) | 4. **Tests to order**<br>**Initial laboratory tests:**<br>- **CBC with differential**<br>- Purpose: look for **eosinophilia**, leukocytosis, anemia.<br>- Expected: eosinophilia if eosinophilic gastroenteritis; leukocytosis if infection/inflammation.<br>- **CRP/ESR**<br>- Purpose: assess inflammatory activity.<br>- Expected: may be elevated but nonspecific.<br>- **Electrolytes, BUN/creatinine, liver enzymes, albumin**<br>- Purpose: assess dehydration, organ involvement, protein loss, alternative causes of ascites.<br>- **Amylase/lipase**<br>- Purpose: evaluate pancreatitis.<br>- **IgE level**<br>- Purpose: supportive evidence for allergic/eosinophilic disease.<br>- **Stool studies / parasite evaluation if exposure history suggests**<br>- Purpose: exclude parasitic infection causing eosinophilia.<br><br>**Imaging:**<br>- **Abdominal CT with contrast**<br>- Purpose: best test to assess **bowel obstruction, bowel wall thickening, ascites, inflammatory changes, ischemia, mass lesions**.<br>- Expected: in eosinophilic gastroenteritis, may show **segmental bowel wall thickening, edema, ascites**; in obstruction, dilated loops/transition point.<br>- **Abdominal ultrasound**<br>- Purpose: detect **ascites** and evaluate hepatobiliary causes; can guide paracentesis if fluid is present.<br><br>**If ascites is present:** |

| | |
|---|---|
|  | - **Diagnostic paracentesis with cell count/differential, albumin, protein, culture, cytology**<br> - Purpose: distinguish eosinophilic ascites from portal hypertensive, infectious, or malignant causes.<br> - Expected: **eosinophil-rich ascitic fluid** would support eosinophilic gastroenteritis.<br><br>**If imaging/labs suggest GI mucosal disease:**<br>- **Upper and/or lower endoscopy with biopsy**<br> - Purpose: confirm **eosinophilic infiltration** and exclude other pathology.<br> - Expected: tissue eosinophilic infiltration, though mucosa may sometimes appear nonspecific.<br><br>Overall, the diagnostic approach should first exclude **urgent obstructive or surgical causes**, while specifically evaluating for **eosinophilic gastroenteritis** given the atopic background. |
| With Static RAG and Mult-agent framework (Model C) | ## 4. Testing plan<br>### Initial tests<br>1. **CBC with differential**<br> - Looks for leukocytosis, anemia, and **eosinophilia**.<br> - Eosinophilia would support eosinophilic GI disease but is not diagnostic.<br><br>2. **Basic metabolic panel, renal function, liver tests, CRP**<br> - Assesses dehydration, electrolyte disturbance, inflammation, and alternative hepatobiliary causes.<br><br>3. **Serum lipase**<br> - Screens for pancreatitis.<br><br>4. **Contrast-enhanced abdominal CT**<br> - Best next imaging test to evaluate:<br> - bowel obstruction or transition point<br> - ileus<br> - ischemia<br> - bowel wall thickening<br> - ascites<br> - inflammatory or malignant causes<br> - This should be prioritized over plain abdominal radiography if CT is readily available.<br><br>5. **Urinalysis**<br> - Helps exclude urinary causes and assess hydration.<br><br>### Selective/next-step tests<br>- **Abdominal X-ray**<br> - Use selectively if CT is delayed or unavailable; otherwise may add limited value.<br>- **Stool studies**, including **ova and parasites**, if diarrhea or eosinophilia is present<br> - Helps exclude infectious/parasitic causes.<br>- **Total IgE**<br> - Supportive only, not confirmatory.<br>- **Upper endoscopy and/or colonoscopy with biopsies**<br> - If imaging and labs do not show a surgical cause and eosinophilic GI disease |

| | |
|---|---|
|  | remains suspected.<br> - **Biopsy is the key confirmatory test** for eosinophilic gastrointestinal disease.<br><br>## Changes from the initial version<br>- The **problem representation** was made more neutral: atopy is now treated as a **clue, not a leading diagnosis by itself**.<br>- The **differential** was **re-prioritized by urgency and commonness**, with obstruction/ileus first and eosinophilic GI disease kept as an important but not dominant possibility.<br>- **Constipation, pancreatitis, ascites-related disorders, and malignancy** were added more explicitly.<br>- The **history and exam** section now includes **previous similar episodes, last bowel movement/flatus, prior abdominal surgery, stool caliber change, weight loss, and focused assessment for ascites**.<br>- The **testing plan** now **prioritizes contrast-enhanced CT and basic labs early**, uses abdominal X-ray more selectively, and states more clearly that **endoscopic biopsy is required to confirm eosinophilic GI disease**. |
| **Phase II – Q5: Interpreting test results** | |
| Baseline LLM (Model A) | **5. Interpretation of the test results**<br><br>- **Marked colonic dilatation proximal to the sigmoid colon on CT** is the most important finding. This indicates **large-bowel obstruction at or near the sigmoid colon**, rather than a primary inflammatory or functional bloating disorder.<br>- **Distended abdomen** on examination supports a mechanically significant obstruction.<br>- **Tachycardia (HR 112/min)** suggests physiologic stress and likely **volume depletion**.<br>- **Venous blood gas: pH 7.446, HCO3 30.7 mmol/L** indicates **metabolic alkalosis**. Together with **hypokalemia (K 3.1)** and **hypochloremia (Cl 88)**, this is consistent with **chloride- and potassium-depletion from gastrointestinal fluid sequestration/losses in bowel obstruction**.<br>- **BUN 98 mg/dL and creatinine 2.69 mg/dL** indicate **acute kidney injury**, most likely **prerenal azotemia from dehydration/third spacing** in the setting of obstructive illness. The very high BUN relative to creatinine supports volume depletion.<br>- **Normal temperature** and lack of other obvious exam abnormalities do not exclude serious obstruction, but there is no clear evidence here of systemic infection or peritonitis.<br><br>**Clinical meaning:** the data support **hemodynamically relevant large-bowel obstruction with dehydration, electrolyte depletion, metabolic alkalosis, and prerenal AKI**. |
| With Static RAG (Model B) | **5. Interpretation of the test results**<br><br>- **Marked colonic dilatation proximal to the sigmoid colon on CT** is the most important finding. This strongly indicates **large-bowel obstruction at or near the sigmoid colon** rather than a functional bloating disorder. In the setting of 1 week of abdominal pain and distension, this is a structurally significant process and explains the abdominal distension on examination.<br>- **Tachycardia (112/min)** suggests significant physiologic stress, most likely from **volume depletion**, pain, or evolving obstructive illness.<br>- **VBG: pH 7.446, HCO3 30.7 mmol/L, pCO2 45.4 mmHg** indicates a |

| | |
|---|---|
| | **metabolic alkalosis with respiratory compensation**. Together with **hypokalemia (K 3.1 mEq/L)** and **hypochloremia (Cl 88 mEq/L)**, this pattern is most consistent with **chloride-responsive contraction alkalosis**, usually due to gastrointestinal fluid losses and dehydration in obstructive states.<br>- **BUN 98 mg/dL and creatinine 2.69 mg/dL** indicate **acute kidney injury**, most likely **prerenal** from reduced effective circulating volume/dehydration. The very high BUN relative to creatinine supports this interpretation.<br>- **Normal sodium (136 mEq/L)** does not exclude major volume depletion; isotonic fluid loss can still produce severe prerenal azotemia.<br>- The absence of fever and the lack of clear peritoneal findings are somewhat reassuring against perforation or advanced peritonitis at this moment, but do **not** rule out impending ischemia or need for urgent decompression.<br><br>**Overall meaning:** the data fit a patient with **mechanical large-bowel obstruction causing dehydration, hypokalemic hypochloremic metabolic alkalosis, and prerenal AKI**. |
| With Static RAG and Mult-agent framework (Model C) | 5. **Interpretation of the test results**<br><br>- **CT: marked dilatation of the colon proximal to the sigmoid colon**<br>- This strongly supports **mechanical large-bowel obstruction** with a **likely transition point at or distal to the sigmoid colon**.<br>- It is more consistent with a focal obstructive process than with nonspecific diffuse ileus.<br>- However, the **exact cause is still not defined** from the information provided; important possibilities include **sigmoid volvulus**, **obstructing colorectal cancer**, or **stricture**.<br><br>- **Abdominal distension without other obvious abnormalities on examination**<br>- This fits with **colonic obstruction**.<br>- The absence of obvious peritoneal signs is somewhat reassuring, but it **does not exclude ischemia or perforation**, especially in a patient with prolonged symptoms and marked dilation.<br><br>- **Tachycardia (112/min) with normal temperature and preserved blood pressure**<br>- This suggests **physiologic stress**, and **volume depletion is likely** in the setting of bowel obstruction and poor intake/third spacing.<br>- He is not frankly hypotensive, but the tachycardia makes this more clinically significant.<br><br>- **VBG: pH 7.446, HCO3 30.7 mmol/L, pCO2 45.4 mmHg**<br>- This is **compatible with a mild metabolic alkalemic tendency**, with roughly appropriate respiratory compensation.<br>- The alkalemic pattern plus **hypochloremia (Cl 88)** and **hypokalemia (K 3.1)** suggests **chloride- and potassium-depletion**, likely related to poor intake and gastrointestinal fluid sequestration from obstruction.<br>- This interpretation should be made **cautiously**, because isolated distal colonic obstruction is a less classic cause of metabolic alkalosis than vomiting or proximal GI losses.<br><br>- **BUN 98 mg/dL, creatinine 2.69 mg/dL, eGFR 22.5**<br>- This indicates **acute kidney injury or severe renal dysfunction**.<br>- Given the tachycardia and markedly elevated **BUN/creatinine ratio**, **prerenal azotemia from volume depletion is most likely**, but **intrinsic renal** and |

| | |
|---|---|
| | **postrenal** causes are not fully excluded by the provided data alone.<br><br>- **Overall significance**<br>- These results support **clinically significant large-bowel obstruction** with important systemic consequences: **volume depletion, hypokalemia, hypochloremia, likely contraction alkalosis, and renal dysfunction**.<br>- Even without shock or clear peritonitis, these abnormalities increase urgency because they suggest the obstruction is already causing **organ-level complications**. |
| **Phase II – Q6: Reconsidering the differential diagnosis** | |
| Baseline LLM (Model A) | **6. Updated diagnostic thinking / Working diagnosis**<br><br>The differential diagnosis changes substantially. The leading problem is now clearly **mechanical large-bowel obstruction involving the sigmoid colon**.<br><br>**Reprioritized differential**<br>1. **Sigmoid volvulus** — most likely<br>- CT shows obstruction localized to the **sigmoid region**, which is classic for sigmoid volvulus and fits abdominal distension with subacute progression.<br>2. **Obstructing sigmoid colon lesion** (e.g., colorectal cancer/stricture)<br>- Also compatible with proximal colonic dilatation, though the patient is relatively young and no further supporting data are given.<br>3. **Pseudo-obstruction (Ogilvie syndrome)**<br>- Less likely because CT specifically suggests an obstructive point near the sigmoid colon.<br><br>The previous concern for **eosinophilic gastroenteritis** becomes much less likely because the imaging now demonstrates a **focal large-bowel obstructive process**, which better explains the abdominal pain, distension, electrolyte abnormalities, and AKI.<br><br>**Working diagnosis:**<br>**Large-bowel obstruction at the sigmoid colon, most likely sigmoid volvulus, complicated by dehydration, hypokalemic hypochloremic metabolic alkalosis, and prerenal acute kidney injury.** |
| With Static RAG (Model B) | **6. Updated diagnostic thinking / Working diagnosis**<br><br>The new results substantially shift the differential toward **mechanical distal colonic obstruction**.<br><br>**Revised prioritization**<br>1. **Large-bowel obstruction at the sigmoid colon** — most likely<br>2. **Sigmoid volvulus** or **obstructing sigmoid lesion/stricture** as the immediate cause<br>3. Less likely: **pseudo-obstruction (Ogilvie syndrome)**, because CT reportedly shows dilatation **proximal to the sigmoid colon**, suggesting a distal transition/obstructive point rather than diffuse functional dilatation<br>4. **Eosinophilic gastroenteritis** becomes much less likely as the primary diagnosis because CT now supports a focal obstructive colonic process rather than diffuse allergic/infiltrative bowel disease<br><br>**Working diagnosis:** |

| | |
|---|---|
| | **Acute large-bowel obstruction centered at the sigmoid colon, complicated by volume depletion, hypokalemic hypochloremic metabolic alkalosis, and prerenal acute kidney injury.** |
| With Static RAG and Mult-agent framework (Model C) | 6. **Updated diagnostic thinking / Working diagnosis**<br><br>The new data substantially narrow the differential toward **mechanical large-bowel obstruction**.<br><br>- The CT finding of **marked colonic dilatation proximal to the sigmoid colon** indicates a **distal colonic obstructive process**, with a **likely transition point at or distal to the sigmoid colon**.<br>- This makes nonspecific functional bloating, uncomplicated constipation, peptic disease, and pancreatitis much less likely as the primary explanation.<br>- The exact etiology remains uncertain from the provided information and depends on the detailed CT morphology and/or endoscopic or operative findings.<br><br>**Most likely working diagnosis:**<br>- **Mechanical large-bowel obstruction with a likely transition point at or distal to the sigmoid colon, complicated by volume depletion, hypokalemia/hypochloremia, mild metabolic alkalemic tendency, and likely prerenal AKI.**<br><br>**Priority etiologies within this diagnosis:**<br>1. **Sigmoid volvulus**<br>- Strong consideration because the obstruction appears distal colonic and may be centered around the sigmoid region.<br>2. **Obstructing sigmoid or distal colonic malignancy / stricture**<br>- Also important, especially because CT here establishes obstruction but not the precise cause.<br>3. **Less likely alternatives**<br>- Severe functional ileus or pseudo-obstruction are less favored given the focal distal colonic pattern, though definitive exclusion depends on imaging details.<br><br>**Why this is now more probable**<br>- The combination of **subacute abdominal distension**, **focal colonic dilation on CT**, and **systemic metabolic/renal consequences** is most consistent with a **true obstructive colonic process**, not a benign functional complaint.<br>- The previous consideration of **eosinophilic GI disease** becomes much less likely as the leading diagnosis because the imaging now points to **mechanical distal colonic obstruction**. |
| **Phase II – Q7: Treatment planning** | |
| Baseline LLM (Model A) | **7. Treatment plan**<br><br>**Immediate management**<br>- **Urgent hospitalization and bowel obstruction management**<br>- **Nil per os (NPO)**<br>- **Aggressive IV isotonic fluid resuscitation**<br>- Rationale: correct **volume depletion**, improve **prerenal AKI**, and treat tachycardia.<br>- **Electrolyte correction, especially potassium and chloride**<br>- Rationale: addresses **hypokalemia/hypochloremic metabolic alkalosis** and may reduce arrhythmic risk.<br>- **Close monitoring** |

| | |
|---|---|
| | - Vitals, urine output, serial abdominal exams, renal function, electrolytes.<br><br>**Definitive/targeted treatment**<br>- If **sigmoid volvulus** is suspected and there are **no signs of perforation, peritonitis, or bowel ischemia**, the preferred initial treatment is **urgent endoscopic detorsion/decompression** (e.g., flexible sigmoidoscopy or colonoscopy).<br>- Rationale: this is guideline-consistent first-line therapy for uncomplicated sigmoid volvulus and can rapidly relieve obstruction.<br>- If endoscopic decompression fails, or if there are signs of **ischemia, necrosis, perforation, or peritonitis**, then **urgent surgery** is required.<br>- Options include operative detorsion/resection depending on bowel viability.<br>- After successful endoscopic decompression, **definitive elective surgery** is often considered because recurrence is common.<br><br>**Alternative approach if another cause is identified**<br>- If imaging/endoscopy instead shows an **obstructing sigmoid tumor or stricture**, management shifts to **surgical and/or oncologic evaluation**, with decompression or resection as appropriate.<br><br>**Why this plan fits this patient**<br>- He already has **organ impact** from obstruction: **AKI, metabolic alkalosis, hypokalemia**, and significant distension. Therefore treatment should be **urgent**, not observational. |
| With Static RAG (Model B) | **7. Treatment plan**<br><br>**Immediate management**<br>- **Urgent admission and close monitoring**<br>- Rationale: large-bowel obstruction can rapidly progress to ischemia, perforation, sepsis, and worsening AKI.<br>- **Bowel rest (NPO)**<br>- Rationale: reduces ongoing bowel distension and aspiration risk.<br>- **Aggressive isotonic IV fluid resuscitation**<br>- Preferably **normal saline**, with careful reassessment<br>- Rationale: treats **volume depletion**, supports renal perfusion, and helps correct **chloride-responsive metabolic alkalosis**.<br>- **Potassium replacement**<br>- Rationale: correct **hypokalemia**, which can worsen ileus and arrhythmia risk.<br>- **Serial abdominal examinations, urine output monitoring, repeat electrolytes/renal function**<br>- Rationale: detect deterioration, ischemia, and response to resuscitation.<br><br>**Definitive evaluation/treatment of the obstruction**<br>- **Urgent surgical consultation**<br>- Rationale: a mechanical large-bowel obstruction is potentially a surgical emergency.<br>- **Urgent lower GI decompression/endoscopic evaluation if appropriate**<br>- Especially if **sigmoid volvulus** is suspected and there are no signs of perforation or peritonitis<br>- Rationale: endoscopic detorsion/decompression can be first-line in uncomplicated sigmoid volvulus.<br>- **Operative management if there is ischemia, perforation, failed endoscopic decompression, or concern for obstructing tumor/stricture requiring resection**<br>- Rationale: definitive treatment depends on the cause and bowel viability. |

| | |
|---|---|
| | **Other supportive care**<br>- **Avoid nephrotoxins**<br>- Rationale: AKI is present.<br>- **Analgesia and antiemetics as needed**<br>- Rationale: symptom control, while avoiding medications that may mask peritoneal signs excessively.<br><br>**Alternative options depending on cause**<br>- **If sigmoid volvulus:** endoscopic detorsion first if stable and no perforation; surgery if recurrence or failure.<br>- **If malignant/structural obstruction:** surgery and, in selected cases, **colonic stenting** as bridge to surgery or palliation.<br>- **If pseudo-obstruction were later favored:** conservative management ± neostigmine, but current CT description makes this less likely.<br><br>**Bottom line:**<br>This patient needs **urgent treatment for mechanical sigmoid-level large-bowel obstruction plus immediate correction of dehydration, electrolyte abnormalities, and prerenal AKI**. |
| With Static RAG and Mult-agent framework (Model C) | 7. **Treatment plan**<br><br>### Immediate management<br>- **Urgent hospital admission**<br>- **Immediate colorectal/general surgery consultation**<br>- This is necessary because the patient has **large-bowel obstruction with renal dysfunction and electrolyte abnormalities**.<br>- **Prompt gastroenterology/endoscopy consultation**<br>- Especially important if **sigmoid volvulus** is suspected and there are **no signs of peritonitis, perforation, or ischemia**, because urgent endoscopic decompression may be therapeutic.<br><br>- **NPO**<br>- To avoid worsening distension and to prepare for possible endoscopic or surgical intervention.<br><br>- **IV isotonic chloride-rich fluids**<br>- For example, normal saline-based resuscitation is reasonable to address likely **volume depletion, hypochloremia, and renal hypoperfusion**.<br>- Because renal function is impaired, fluid replacement should be **carefully monitored** with serial vitals and urine output.<br><br>- **Potassium repletion**<br>- Correct **hypokalemia**, ideally with ECG monitoring if clinically indicated, because low potassium increases arrhythmia risk and may worsen bowel dysmotility.<br><br>- **Close monitoring**<br>- **Serial abdominal examinations**<br>- **Urine output**<br>- **Repeat electrolytes and renal function**<br>- **Repeat lactate and other labs if clinically indicated**<br>- Ongoing surveillance is important because **ischemia or perforation can develop despite initially limited exam findings**. |

<table>
<tr><td></td><td>### Management of the obstruction<br>- If imaging/clinical assessment suggests **sigmoid volvulus without peritonitis or perforation**:<br>  - **Urgent endoscopic detorsion/decompression** is an evidence-based first step.<br>- If there are signs of **peritonitis, bowel ischemia, perforation, hemodynamic deterioration**, or **failed endoscopic decompression**:<br>  - **Urgent operative management** is required.<br>- If an **obstructing tumor or stricture** is the cause:<br>  - Definitive management is usually **surgical**, with exact timing and approach depending on stability, decompression options, and specialist assessment.<br><br>### Adjuncts / alternatives<br>- **Nasogastric tube**<br>  - May be considered if there is significant vomiting or upper GI distension, though it is not definitive treatment for distal colonic obstruction.<br>- **Further etiologic evaluation after stabilization**<br>  - Depending on the course, **endoscopy** and/or more detailed radiologic interpretation will be needed to distinguish **volvulus, malignancy, or stricture**.<br><br>### Rationale<br>- The patient already shows **systemic effects of obstruction**: tachycardia, electrolyte depletion, and renal dysfunction.<br>- The immediate priorities are:<br>  1. **Resuscitate**<br>  2. **Correct electrolytes**<br>  3. **Identify and relieve the obstruction promptly**<br>  4. **Watch closely for ischemia or perforation**<br><br>---<br><br>**Changes from the initial results**<br>- The differential is now **much more narrowly focused on mechanical large-bowel obstruction**, rather than a broad abdominal pain/bloating workup.<br>- **Eosinophilic gastrointestinal disease**, previously considered because of atopy, is now **substantially deprioritized** because CT demonstrates a focal obstructive colonic pattern.<br>- The wording was revised to be more precise: **CT suggests a transition point at or distal to the sigmoid colon**, rather than claiming the lesion is definitively “in the sigmoid colon.”<br>- The acid-base and renal interpretations were made **more cautious**:<br>  - **Metabolic alkalosis** is described as a **mild alkalemic tendency compatible with chloride/potassium depletion**, rather than stated too definitively.<br>  - **Prerenal AKI** is described as **most likely**, but not proven.<br>- The treatment plan now more explicitly emphasizes:<br>  - **Immediate surgery consultation**<br>  - **Early endoscopy consultation if volvulus is suspected**<br>  - **Chloride-rich fluids with potassium repletion**<br>  - **Careful monitoring in the setting of AKI**<br>  - **Serial assessment for ischemia or perforation**</td></tr>
<tr><td colspan="2">Phase III – Q8: Evaluation of treatment</td></tr>
<tr><td>Baseline LLM (Model A)</td><td>8. **Monitoring of effectiveness, safety, risks, and countermeasures**<br><br>- **Effectiveness monitoring**</td></tr>
</table>

| | |
|---|---|
| | - **Serum sodium, chloride, potassium, bicarbonate, BUN, creatinine**: check frequently to confirm improvement of **hypernatremia, chloride-responsive metabolic alkalosis, and AKI**.<br>- **Blood gas analysis**: follow pH and bicarbonate to assess correction of alkalosis.<br>- **Fluid balance**: strict input/output charting, including **oral intake, IV fluids, urine volume, and intestinal drainage** from the ileus tube.<br>- **Clinical status**: abdominal distension, bowel function, ability to eat/drink, mental status, vital signs.<br>- **Urine studies when needed**: urine electrolytes/osmolality can help reassess ongoing renal sodium/chloride handling and free water balance.<br><br>- **Main safety risks and what to watch for**<br>1. **Overly rapid correction of hypernatremia**<br>- Risk: cerebral edema and neurologic deterioration.<br>- Monitoring: serial serum sodium and neurologic checks.<br>- Countermeasure: adjust free-water administration gradually and recheck sodium frequently.<br>2. **Persistent or worsening volume depletion from ongoing intestinal losses**<br>- Risk: recurrent prerenal AKI, worsening alkalosis, hypotension.<br>- Monitoring: urine output, heart rate, blood pressure, BUN/Cr, drainage volume.<br>- Countermeasure: replace ongoing losses appropriately and revise IV fluid composition based on electrolytes.<br>3. **Hypokalemia and hypochloremia**<br>- Risk: maintenance of metabolic alkalosis and arrhythmia.<br>- Monitoring: serial electrolytes and ECG if clinically indicated.<br>- Countermeasure: potassium and chloride repletion as needed.<br>4. **Fluid overload after renal recovery or excessive infusion**<br>- Risk: edema, pulmonary congestion.<br>- Monitoring: body weight, oxygenation, physical examination, urine output.<br>- Countermeasure: reduce infusion rate or change fluid strategy.<br>5. **Complications of HCG-AA**<br>- Risk: increased nitrogen load with rise in **BUN**, possible worsening azotemia if renal excretion is insufficient.<br>- Monitoring: BUN, creatinine, urine output, overall renal course.<br>- Countermeasure: stop or reduce HCG-AA if azotemia worsens or renal function declines.<br>6. **Recurrence or worsening of ileus/volvulus**<br>- Risk: renewed obstruction, ischemia, perforation.<br>- Monitoring: abdominal pain/distension, vomiting, bowel movement, inflammatory signs.<br>- Countermeasure: urgent imaging and surgical/endoscopic reassessment if suspected. |
| With Static RAG (Model B) | **8. Monitoring effectiveness, safety, risks, and countermeasures**<br><br>**Effectiveness monitoring**<br>- **Serum sodium, chloride, potassium, bicarbonate, BUN, creatinine**: check frequently (at least daily, and more often while sodium is changing) to confirm improvement of **hypernatremia, chloride depletion metabolic alkalosis, and AKI**.<br>- **Blood gas analysis**: follow **pH/HCO3−** to confirm resolution of metabolic alkalosis.<br>- **Fluid balance**: strict **input/output**, including ileus tube or stool losses, urine volume, and oral intake.<br>- **Clinical assessment**: thirst, mental status, abdominal distension, bowel function, |

| | |
|---|---|
| | hemodynamics.<br>- **Urine studies if needed**: repeat **urine electrolytes/osmolality** if the response is not as expected, to reassess renal handling of sodium/chloride/water.<br><br>**Main safety risks to watch for**<br>1. **Overly rapid correction of hypernatremia**<br>- Risk: cerebral edema.<br>- Countermeasure: avoid excessive fall in serum sodium; monitor sodium closely and adjust infusion/oral water gradually.<br><br>2. **Persistent or worsening volume depletion from ongoing intestinal losses**<br>- Risk: recurrent prerenal AKI, sustained alkalosis, worsening hypernatremia.<br>- Countermeasure: quantify GI losses carefully and replace ongoing losses appropriately; reassess fluid prescription daily.<br><br>3. **Hypokalemia**<br>- Risk: worsened metabolic alkalosis, arrhythmia, impaired gut motility.<br>- Countermeasure: serial potassium monitoring and active potassium replacement.<br><br>4. **Excess nitrogen/amino acid load from HCG-AA**<br>- Risk: rise in **BUN**, possible worsening azotemia if renal handling is inadequate.<br>- Countermeasure: monitor **BUN/creatinine**, urine output, and discontinue or reduce HCG-AA if azotemia worsens.<br><br>5. **Fluid overload after AKI recovery or with ongoing IV therapy**<br>- Risk: edema, heart failure, worsening abdominal status.<br>- Countermeasure: daily weight, physical examination, urine output, and adjustment of infusion rate.<br><br>6. **Recurrent ileus/obstruction or bowel ischemia**<br>- Risk: renewed GI losses, sepsis, perforation.<br>- Countermeasure: monitor abdominal symptoms, distension, bowel sounds, and promptly repeat imaging/surgical review if symptoms worsen.<br><br>7. **Hyperglycemia with 5% dextrose**<br>- Risk: osmotic diuresis and difficulty controlling sodium/water balance.<br>- Countermeasure: monitor blood glucose and adjust therapy if needed. |
| With Static RAG and Mult-agent framework (Model C) | ### 8. Monitoring of treatment effectiveness and safety<br><br>**Effectiveness monitoring**<br>- **Serum sodium, chloride, potassium, bicarbonate, BUN, and creatinine**<br>- Follow serially to assess response of **hypernatremia, chloride-depletion metabolic alkalosis, and prerenal AKI**.<br>- During active correction of hypernatremia, **serum sodium should be checked frequently**.<br>- **Potassium should also be monitored closely**, because potassium depletion can perpetuate metabolic alkalosis and increase arrhythmia risk.<br><br>- **Acid-base status**<br>- Repeat **blood gas and/or serum bicarbonate** to confirm that metabolic alkalosis is resolving after changes in fluid therapy and after HCG-AA is started. |

- **Fluid balance**
  - Maintain strict **input/output records** and **daily body weight**.
  - In this case, it is especially important to **quantify and document gastrointestinal drainage from the trans-anal ileus tube each day**, because ongoing intestinal chloride-rich losses likely contributed to persistent alkalosis and hypernatremia.

- **Urine studies**
  - Repeat **urine electrolytes** if the course is unclear or improvement is delayed.
  - A persistently **low urine chloride** would continue to support **chloride-depletion alkalosis**, while repeat urine indices may also show changing renal handling as AKI recovers.

- **Clinical bowel recovery**
  - Follow **abdominal distension, pain, bowel function, tolerance of oral intake, and imaging/procedural findings** as needed.
  - Improvement in ileus, removal of the tube, and increased oral intake all change fluid and electrolyte needs.

**Safety monitoring / complications to watch for**
- **Overly rapid correction of hypernatremia**
  - Risk: neurologic injury from overly rapid fall in serum sodium.
  - Plan: check serum sodium frequently during active treatment and adjust free-water/infusion strategy to avoid excessive daily correction.

- **Persistent or worsening hypernatremia**
  - Risk: ongoing water deficit and neurologic complications.
  - Plan: reassess **oral water allowance, ongoing intestinal losses, renal concentrating/excreting ability, and adequacy of low-sodium replacement**.

- **Persistent metabolic alkalosis**
  - Risk: worsened hypokalemia, arrhythmia, impaired physiologic recovery.
  - Plan: continue a **chloride repletion strategy**, reassess ongoing GI chloride loss, and correct potassium deficiency.

- **Hypokalemia and other electrolyte shifts**
  - Risk: arrhythmia and maintenance of alkalosis.
  - Plan: serial electrolytes with potassium supplementation as needed.

- **Fluid overload during renal recovery**
  - Risk: edema or heart failure while renal function is still changing.
  - Plan: monitor **urine output, weight, blood pressure, and clinical volume status**, and adjust infusion rates promptly.

- **Azotemia from HCG-AA**
  - Risk: increased nitrogen load, especially if renal excretion is inadequate.
  - Plan: monitor **BUN/creatinine**, and if needed urinary urea nitrogen; **reduce or stop HCG-AA** if clinically meaningful azotemia develops.

- **Need to adjust or stop HCG-AA**
  - Reconsider HCG-AA if:
    - **BUN/creatinine rises significantly**
    - **fluid overload develops**
    - **hypernatremia, alkalosis, or other electrolyte abnormalities worsen rather than improve**

| | |
|---|---|
|  | - **Failure of bowel recovery or recurrent obstruction**<br> - Risk: recurrent losses, perforation, or need for further intervention.<br> - Plan: monitor abdominal findings and oral intake tolerance; promptly reassess if distension, pain, or obstructive symptoms recur.<br><br>**Changes from the initial version**<br>- Added **more explicit priority for frequent sodium monitoring** during active correction.<br>- Added **continued close potassium monitoring** because hypokalemia can maintain alkalosis.<br>- Strengthened the need to **measure and document actual GI output daily** from the ileus tube.<br>- Added clearer conditions for **adjusting or discontinuing HCG-AA** if azotemia, overload, or electrolyte problems worsen. |
| **Phase III – Q9: Formulating plans for worsening conditions** | |
| Baseline LLM (Model A) | 9. **If the course is not as expected or changes suddenly: reassessment and action plan**<br><br>- **Reassess the problem in parallel:**<br> 1. **Is there ongoing loss of water, chloride, or potassium?**<br> - Review intestinal drainage, stool output, oral intake, and urine output.<br> 2. **Is the current fluid prescription appropriate?**<br> - Compare administered sodium/chloride/free water with ongoing losses and current labs.<br> 3. **Has kidney function changed?**<br> - Recheck BUN/Cr, urine volume, and urine electrolytes to determine whether impaired dilution/concentration or osmotic diuresis is contributing.<br> 4. **Has the bowel problem recurred or become complicated?**<br> - Consider recurrent obstruction, ischemia, or perforation if abdominal findings worsen.<br><br>- **If hypernatremia does not improve**<br> - Recalculate ongoing free-water deficit and ongoing losses.<br> - Reassess whether intestinal losses plus saline administration are still creating a net sodium gain/free-water deficit.<br> - Increase free-water replacement carefully and continue close sodium monitoring.<br> - If chloride-responsive alkalosis persists despite dextrose water alone, use a strategy that also provides **chloride with low sodium burden**, as was done with HCG-AA.<br><br>- **If metabolic alkalosis does not improve**<br> - Confirm it remains **chloride-responsive** rather than another cause.<br> - Reassess urine chloride, potassium depletion, and ongoing gastrointestinal chloride loss.<br> - Intensify chloride and potassium replacement while treating the source of losses.<br><br>- **If renal function worsens again**<br> - Distinguish recurrent prerenal AKI from intrinsic kidney injury using clinical course, urine output, and repeat urine studies.<br> - Review hemodynamics, volume status, and nephrotoxic exposures.<br> - Adjust fluids promptly and reconsider the safety of nitrogen-containing infusion.<br><br>- **If sudden clinical deterioration occurs** |

| | |
|---|---|
| | - Immediate bedside reassessment of **vital signs, mental status, abdominal examination, urine output**.<br>- Repeat urgent labs: electrolytes, blood gas, BUN/Cr.<br>- Obtain urgent abdominal imaging if obstruction/perforation/ischemia is suspected.<br>- Escalate to emergency endoscopic or surgical management if bowel complications are identified.<br><br>In summary, management should remain **dynamic**: repeatedly match treatment to **current electrolyte abnormalities, renal function, and ongoing gastrointestinal losses**, while staying alert for recurrent bowel obstruction and for complications from correction itself. |
| With Static RAG (Model B) | **9. If the condition does not improve or suddenly changes: reassessment and action plan**<br><br>**How to think**<br>- Reassess whether the initial explanation remains sufficient. In this case, persistent **hypernatremia + metabolic alkalosis** despite improving renal function should prompt review of:<br>- **Ongoing gastrointestinal chloride-rich or hypotonic fluid losses**<br>- **Insufficient free water replacement**<br>- **Renal water handling** during AKI recovery (including increased free water clearance/urea diuresis)<br>- Less likely but important alternative causes of hypernatremia or alkalosis if the course becomes atypical<br><br>**How to act**<br>1. **Repeat focused clinical assessment**<br>- Volume status, mental status, abdominal findings, urine output, net fluid balance.<br><br>2. **Repeat key labs promptly**<br>- Sodium, potassium, chloride, bicarbonate, BUN, creatinine, blood gas, glucose.<br>- Add/repeat **urine electrolytes and osmolality** if the mechanism is unclear.<br><br>3. **Recalculate ongoing losses and revise replacement**<br>- Include intestinal/ileus tube losses, urine output, and oral intake.<br>- If sodium remains high and alkalosis persists, use therapies that provide **free water and chloride without excessive sodium**, as was done with **HCG-AA** in this case.<br><br>4. **Reevaluate the abdomen and bowel course**<br>- If abdominal distension, pain, or drainage worsens, consider recurrent obstruction, persistent ileus, or bowel complications, and obtain repeat imaging/endoscopic or surgical reassessment.<br><br>5. **Escalate care if acute deterioration occurs**<br>- Especially for neurologic symptoms, hemodynamic instability, severe electrolyte shifts, or suspected bowel ischemia/perforation.<br>- Intensify monitoring and make urgent changes in fluid/electrolyte therapy.<br><br>**Practical principle**<br>- If the expected improvement does not occur, do not simply continue the same plan; instead, **redefine the dominant mechanism** of the electrolyte/acid-base disorder and **match treatment to that mechanism**. In this patient, that meant recognizing |

| | |
|---|---|
| | persistent **chloride depletion plus free-water deficit with ongoing intestinal losses**, and shifting from NS/D5W alone to **low-sodium, high-chloride therapy** while oral intake recovered. |
| With Static RAG and Mult-agent framework (Model C) | ### 9. Reassessment framework and flexible revision of diagnosis/treatment<br><br>**Reassessment framework**<br>If the course is not improving, reassess the problem in 3 parts:<br>1. **Is hypernatremia being driven by ongoing water loss, inadequate water replacement, or both?**<br>2. **Is metabolic alkalosis still chloride-responsive, or is another maintenance factor present?**<br>3. **Has the bowel condition changed again, including persistent drainage, recurrent obstruction, ischemia, or another complication?**<br><br>**If hypernatremia does not improve**<br>- Re-evaluate likely contributors identified in this case:<br> - **ongoing intestinal fluid loss**<br> - **restricted oral water intake**<br> - **impaired renal handling during AKI**<br> - **increased free water clearance during renal recovery/urea diuresis**<br> - the prior effect of **normal saline administration in the setting of hypotonic GI losses**<br>- Recheck:<br> - **serum sodium trend**<br> - **intake/output**<br> - **measured ileus-tube drainage**<br> - **renal function**<br> - **urine electrolytes and/or osmolality if needed**<br>- Then revise treatment:<br> - increase **free water** if clinically safe,<br> - but also address persistent **chloride depletion** rather than assuming water deficit alone is the whole problem.<br><br>**If metabolic alkalosis persists**<br>- Reassess whether **ongoing gastrointestinal chloride loss** is continuing to maintain alkalosis.<br>- Repeat **urine chloride** if necessary; a low value would still support **chloride-depletion alkalosis**.<br>- Reassess and correct **potassium depletion**, since hypokalemia can sustain alkalosis.<br>- If free-water replacement alone is insufficient and further sodium loading is undesirable, a **low-sodium, chloride-rich approach such as HCG-AA may be considered as an individualized rescue strategy** .<br><br>**How to interpret improvement after HCG-AA**<br>- Improvement after HCG-AA is **compatible with benefit from chloride repletion without excess sodium**.<br>- However, causality should be stated **cautiously**, because improvement may also have been influenced by:<br> - **removal of the trans-anal ileus tube**<br> - **resumption and increase of oral intake**<br> - **ongoing recovery of renal function**<br>- Therefore, HCG-AA should be described as a **plausible, case-specific treatment |

| | |
|---|---|
|  | choice**, not established standard therapy based on this information alone.<br><br>**If the patient suddenly worsens**<br>- Immediately reassess for:<br> - **recurrent or unresolved bowel obstruction**<br> - **bowel ischemia or perforation**<br> - **worsening AKI**<br> - **severe electrolyte complications**<br>- Immediate actions:<br> - urgent repeat clinical evaluation<br> - repeat labs including **electrolytes, bicarbonate, BUN/Cr, and potassium**<br> - repeat abdominal assessment/imaging as indicated<br> - prompt re-consultation with **gastroenterology and surgery**<br><br>**Principle**<br>- Management should be revised repeatedly according to the changing balance among:<br> - **intestinal losses**<br> - **sodium and chloride administration**<br> - **free water deficit**<br> - **potassium depletion**<br> - **renal recovery**<br>- In this case, the lack of response to **5% dextrose alone** appropriately prompted reconsideration of the mechanism. A **chloride-rich, low-sodium strategy** was therefore a reasonable individualized option, but its effect should be interpreted alongside concurrent improvement in bowel function and oral intake.<br><br>**Changes from the initial version**<br>- The role of **HCG-AA** is now described **more cautiously** as a plausible individualized rescue approach rather than a standard or definitively proven therapy.<br>- Added explicit reassessment of **potassium depletion** and **ongoing GI chloride loss** before concluding treatment failure.<br>- More clearly acknowledged that improvement may have been due to **multiple simultaneous factors**, not HCG-AA alone. |

| **Case 9: Tubulointerstitial nephritis with IgM-positive plasma cells complicated by liver failure** [17] | |
|---|---|
| **Phase I – Q1: Summarizing medical problems** | |
| Baseline LLM (Model A) | 1. **Problem representation**<br><br>A **70-year-old woman** with a **subacute presentation of general malaise and anorexia** was found to have **profound hypokalemia (K 1.2 mEq/L)**, **metabolic acidosis (HCO3− 14.7 mmol/L)**, **mild renal dysfunction (Cr 1.30 mg/dL)**, and **bilateral pitting leg edema**, with **normal blood pressure** and a history of **long-term antihypertensive medication use**.<br>The key clinical problem is **severe hypokalemia with metabolic acidosis**, which strongly suggests **potassium loss with bicarbonate wasting or impaired distal acid handling**, rather than the more common hypokalemia with metabolic alkalosis seen with diuretics or hyperaldosteronism. |
| With Static RAG (Model B) | 1. **Problem representation**<br><br>A **70-year-old woman** with a **subacute presentation of general malaise and anorexia** is found to have **profound hypokalemia (K 1.2 mEq/L)**, **metabolic |

| | |
|---|---|
| | acidosis ($HCO_3^-$ 14.7 mmol/L)**, **mild renal dysfunction (Cr 1.30 mg/dL)**, and **bilateral pitting leg edema**, with **normal blood pressure** at presentation and a history of long-term antihypertensive use.<br>The key medical problem is **severe potassium depletion with metabolic acidosis**, suggesting either **renal or extrarenal potassium loss associated with bicarbonate loss or impaired acid excretion**, and the edema raises concern for a disorder of **volume handling, kidney disease, medication effect, or endocrine disease**. |
| With Static RAG and Mult-agent framework (Model C) | ## 1. Problem representation<br><br>A **70-year-old woman** with a **subacute course of general malaise and anorexia** is found to have **profound hypokalemia (K 1.2 mEq/L)**, **metabolic acidosis ($HCO_3^-$ 14.7 mmol/L)**, **mild renal dysfunction (Cr 1.30 mg/dL)**, and **bilateral pitting leg edema**, with **normal blood pressure and sinus rhythm** on presentation.<br><br>This is first and foremost a **life-threatening electrolyte emergency** because **K 1.2 mEq/L** carries major risk of **arrhythmia, muscle weakness, and respiratory compromise** and requires **urgent potassium replacement and cardiac monitoring**.<br><br>Syndromically, the major diagnostic issue is **severe hypokalemia with metabolic acidosis**, which raises concern for:<br>- **renal potassium wasting with impaired urinary acidification** such as **renal tubular acidosis (RTA)**,<br>- **gastrointestinal bicarbonate/potassium loss**,<br>- or **medication-related causes**.<br><br>The **edema** is **not well explained by isolated RTA alone** and should be evaluated in parallel for **proteinuric renal disease, hypoalbuminemia, heart failure, liver disease, or medication-related fluid retention**. |
| **Phase I – Q2: Differential diagnoses and rationale** | |
| Baseline LLM (Model A) | 2. **Differential diagnoses**<br><br>**Top 3 differential diagnoses**<br>1. **Renal tubular acidosis (RTA), especially distal or proximal RTA**<br>- Severe hypokalemia plus metabolic acidosis is highly compatible.<br>- Renal dysfunction may coexist.<br>- Distal RTA is particularly important in older adults and may be associated with autoimmune disease or drugs.<br><br>2. **Gastrointestinal potassium and bicarbonate loss (e.g., chronic diarrhea/laxative abuse)**<br>- Can cause hypokalemia and non-anion gap metabolic acidosis.<br>- Would be common and should be excluded by history.<br><br>3. **Drug-induced electrolyte disorder / medication-related renal potassium wasting**<br>- Her long history of antihypertensive treatment raises concern for medication effects.<br>- Some drugs can contribute indirectly to renal tubular dysfunction or potassium loss.<br>- Classic thiazide/loop diuretics more often cause metabolic alkalosis, so they do not fully explain the current pattern unless there is another process. |

| | |
|---|---|
| | **Most likely diagnosis:**<br>**Renal tubular acidosis (especially distal RTA)**, because the combination of **severe hypokalemia + metabolic acidosis** points more strongly to a **renal acidification defect** than to routine antihypertensive adverse effects.<br><br>**Must not be missed:**<br>**Life-threatening complications of severe hypokalemia**, especially **arrhythmia and respiratory/muscle weakness**, regardless of the underlying cause. Also, if edema and renal dysfunction reflect **systemic disease (e.g., nephrotic syndrome, paraproteinemia, autoimmune disease)**, that must be identified promptly. |
| With Static RAG (Model B) | 2. **Differential diagnosis**<br><br>### Top 3 differential diagnoses<br>1. **Renal tubular acidosis (especially distal or proximal RTA) with renal potassium wasting**<br>- Hypokalemia plus **non-anion gap metabolic acidosis** strongly suggests RTA.<br>- Renal dysfunction may be related or contributory.<br>- Distal RTA is especially important in older adults and may present with profound hypokalemia.<br><br>2. **Diuretic-related electrolyte disorder / medication-induced renal potassium loss**<br>- She has a history of antihypertensive treatment for 12 years; concealed or chronic use of **diuretics** could cause severe hypokalemia.<br>- However, diuretics more typically cause **metabolic alkalosis**, so this does **not fully explain the acidosis** unless there is mixed pathology.<br><br>3. **Gastrointestinal potassium and bicarbonate loss (e.g., chronic diarrhea, laxative abuse)**<br>- This can cause **hypokalemia with metabolic acidosis**.<br>- Must be considered even if not yet reported, especially in older patients with poor intake and weakness.<br><br>### Most likely<br>**Renal tubular acidosis with renal potassium wasting** is the most likely based on the combination of **severe hypokalemia + metabolic acidosis**, which is more physiologically consistent than diuretic use alone.<br><br>### Must not be missed<br>**Life-threatening arrhythmia and neuromuscular complications from severe hypokalemia** must not be missed immediately.<br>From a diagnostic standpoint, **RTA due to an underlying systemic disease or drug toxicity**, and **occult GI losses**, should also not be overlooked. |
| With Static RAG and Mult-agent framework (Model C) | ## 2. Differential diagnosis<br><br>### Prioritized differential diagnosis<br><br>1. **Medication-related electrolyte disorder / renal potassium wasting**<br>- This should be prioritized early because she has taken **antihypertensive drugs for 12 years**, but the exact agents are unknown.<br>- **Diuretics** are especially important to exclude, although they more often cause **metabolic alkalosis** rather than acidosis. |

<table>
<tr><td></td><td>- Other prescribed or over-the-counter agents, including **laxatives, herbal medicines, or licorice**, could also contribute.<br><br>2. **Renal tubular acidosis (especially distal RTA; proximal RTA also possible)**<br>- **Hypokalemia with metabolic acidosis** strongly suggests this category.<br>- **Distal RTA** is particularly important because it classically causes **hypokalemia plus inability to acidify urine**.<br>- In an older woman, **secondary distal RTA from autoimmune disease such as Sjögren syndrome** is an important later consideration.<br>- **Proximal RTA** is less likely unless other proximal tubular defects are present.<br><br>3. **Gastrointestinal potassium and bicarbonate loss**<br>- **Chronic diarrhea** or **laxative abuse** can cause **hypokalemia with metabolic acidosis**.<br>- This remains a common and important alternative to renal causes.<br><br>4. **Chronic kidney disease or tubulointerstitial kidney disease contributing to acidosis**<br>- Mild renal dysfunction is present.<br>- CKD alone usually does **not** explain such **profound hypokalemia**, but it could contribute to acidosis or reflect underlying renal disease.<br><br>5. **Edema-related systemic disease**<br>- The bilateral pitting edema raises parallel concern for:<br>- **proteinuric kidney disease / nephrotic syndrome**<br>- **heart failure**<br>- **liver disease / hypoalbuminemia**<br>- These do not directly explain the full electrolyte pattern but may coexist.<br><br>### Most likely diagnosis<br>At this stage, the leading syndromic diagnosis is **renal potassium wasting causing hypokalemic metabolic acidosis**, with **RTA** high on the list, but this should **not be concluded before medication review, urine studies, and anion gap assessment**.<br><br>### Must not be missed<br>- **Immediate complications of profound hypokalemia**: **ventricular arrhythmia, paralysis, respiratory failure**<br>- A **secondary cause of RTA**, especially **autoimmune disease**<br>- A **proteinuric or systemic cause of edema**</td></tr>
<tr><td colspan="2">Phase I – Q3: Necessary physical examinations and reasons</td></tr>
<tr><td>Baseline LLM (Model A)</td><td>3. **Additional history and physical examination**<br><br>**History**<br>- **Medication review:** exact antihypertensives, diuretics, laxatives, herbal medicines<br>- To identify drug-induced potassium loss or tubular injury.<br>- **GI symptoms:** diarrhea, vomiting, laxative use<br>- To distinguish renal from extrarenal potassium/bicarbonate loss.<br>- **Polyuria, polydipsia, nephrolithiasis history**<br>- Suggestive of RTA, especially distal RTA.<br>- **Dry eyes/dry mouth, arthralgia, rash**<br>- To screen for autoimmune disease such as Sjögren syndrome, a cause of distal RTA.</td></tr>
</table>

| | |
|---|---|
|  | - **Weakness, paralysis, palpitations**<br> - To assess urgency and severity of hypokalemia complications.<br>- **Urinary symptoms/foamy urine**<br> - To evaluate proteinuria or kidney disease causing edema.<br><br>**Physical examination**<br>- **Focused neuromuscular exam**<br> - To assess hypokalemic weakness.<br>- **Cardiovascular exam and urgent ECG correlation**<br> - To detect hypokalemia-related conduction abnormalities.<br>- **Volume assessment**<br> - To help interpret edema and renal handling of electrolytes.<br>- **Signs of autoimmune disease** (dry mucosa, parotid enlargement, rash, arthritis)<br> - To identify secondary causes of RTA.<br>- **Abdominal exam**<br> - To look for GI causes or systemic illness. |
| With Static RAG (Model B) | 3. **Additional history and physical examination**<br><br>### Additional questions<br>- **Medication review**<br> - Ask specifically about **diuretics, laxatives, herbal medicines, OTC drugs**, and recent medication changes.<br> - Purpose: identify common causes of potassium loss and possible drug-induced tubular injury.<br><br>- **GI symptoms**<br> - Ask about **diarrhea, vomiting, stool frequency, laxative use**.<br> - Purpose: distinguish **extrarenal bicarbonate/potassium loss** from renal causes.<br><br>- **Urinary symptoms**<br> - Ask about **polyuria, nocturia, nephrolithiasis history, urinary frequency**.<br> - Purpose: RTA may be associated with **polyuria**; distal RTA may be linked to **stones/nephrocalcinosis**.<br><br>- **Symptoms of hypokalemia**<br> - Ask about **muscle weakness, cramps, paralysis, palpitations**.<br> - Purpose: assess urgency and complications.<br><br>- **Autoimmune/systemic symptoms**<br> - Ask about **dry eyes, dry mouth, arthralgia, rash**.<br> - Purpose: screen for causes such as **Sjögren syndrome**, a classic cause of distal RTA.<br><br>- **Edema-related history**<br> - Ask about **dyspnea, orthopnea, reduced urine output, frothy urine, liver disease history**.<br> - Purpose: evaluate cardiac, renal, or hepatic causes of edema.<br><br>### Physical examination<br>- **Volume assessment**<br> - JVP, lung crackles, mucous membranes, skin turgor.<br> - Purpose: determine effective circulating volume and whether edema reflects systemic fluid overload. |

| | |
|---|---|
|  | - **Neuromuscular exam**<br> - Proximal muscle strength, reflexes.<br> - Purpose: detect clinically significant hypokalemic weakness.<br><br>- **Cardiopulmonary exam**<br> - Rhythm, signs of heart failure.<br> - Purpose: identify complications of hypokalemia and causes of edema.<br><br>- **Signs of autoimmune disease**<br> - Oral dryness, parotid enlargement, rash, arthritis.<br> - Purpose: support secondary RTA. |
| With Static RAG and Mult-agent framework (Model C) | ## 3. Additional history and physical examination<br><br>### Focused history<br><br>1. **Complete medication history**<br> - Ask for the **exact antihypertensive drugs**, doses, duration, recent changes, adherence, and all **over-the-counter medications**, **herbal/traditional medicines**, **supplements**, **licorice**, and **laxatives**.<br> - Reason: medication-related disease is a key competing explanation and must be excluded early.<br><br>2. **Gastrointestinal losses**<br> - Ask about **diarrhea, chronic loose stools, vomiting, poor oral intake, laxative use**.<br> - Reason: helps distinguish **GI bicarbonate/potassium loss** from renal causes.<br><br>3. **Symptoms of hypokalemia severity**<br> - Ask about **muscle weakness, cramps, paralysis, palpitations, constipation**, and **dyspnea**.<br> - Reason: identifies urgent complications of severe hypokalemia.<br><br>4. **Renal tubular disorder clues**<br> - Ask about **polyuria, polydipsia, nephrolithiasis, fractures, bone pain**.<br> - Reason: these support possible **RTA**, especially distal RTA.<br><br>5. **Autoimmune symptoms**<br> - Ask about **dry eyes, dry mouth, dental caries, parotid swelling, arthralgia, rash**.<br> - Reason: screens for **Sjögren syndrome or other autoimmune disease** associated with distal RTA.<br><br>6. **Edema/systemic disease symptoms**<br> - Ask about **foamy urine, oliguria, dyspnea, orthopnea, abdominal distension, jaundice**.<br> - Reason: helps evaluate **proteinuria, heart failure, liver disease, hypoalbuminemia**.<br><br>### Focused physical examination<br><br>1. **Cardiac monitoring and cardiovascular exam**<br> - Assess rhythm, perfusion, signs of heart failure, and volume status.<br> - Reason: severe hypokalemia can cause **life-threatening arrhythmia**. |

| | |
|---|---|
| | 2. **Neurologic and muscle examination**<br>- Evaluate **proximal muscle strength, reflexes, and respiratory effort**.<br>- Reason: K 1.2 mEq/L can cause **severe weakness or respiratory compromise**.<br><br>3. **Respiratory assessment**<br>- Check work of breathing and inspiratory strength if needed.<br>- Reason: respiratory muscle weakness may occur in profound hypokalemia.<br><br>4. **Volume and edema assessment**<br>- Examine **JVP, lung fields, sacral/leg edema**, and signs of dehydration.<br>- Reason: helps reconcile edema and assess effective circulating volume.<br><br>5. **Autoimmune examination**<br>- Look for **dry mucosa, parotid enlargement, rash, arthritis**.<br>- Reason: supports autoimmune distal RTA if present. |
| **Phase I – Q4: Plan for investigations/tests** | |
| Baseline LLM (Model A) | 4. **Testing plan**<br><br>1. **Repeat serum electrolytes, blood gas, Mg, phosphate, renal function**<br>- **Purpose:** confirm severity, assess associated abnormalities, guide urgent treatment.<br>- **Expected:** persistent severe hypokalemia and metabolic acidosis; magnesium may also be low.<br><br>2. **ECG**<br>- **Purpose:** evaluate immediate cardiac risk from K 1.2 mEq/L.<br>- **Expected:** possible U waves, ST depression, QT/QU prolongation, arrhythmias.<br><br>3. **Urine electrolytes (especially urine K), urine pH, and urine anion gap**<br>- **Purpose:** determine whether potassium loss is renal and whether acid excretion is impaired.<br>- **Expected in RTA:** inappropriate renal potassium wasting; **urine pH often >5.5 in distal RTA**; positive urine anion gap suggesting low ammonium excretion.<br><br>4. **Urinalysis with protein quantification**<br>- **Purpose:** evaluate kidney disease contributing to edema and renal dysfunction.<br>- **Expected:** may reveal proteinuria if edema is due to renal disease; otherwise may be relatively bland in isolated RTA.<br><br>5. **Serum anion gap, glucose, albumin**<br>- **Purpose:** classify metabolic acidosis and interpret edema.<br>- **Expected:** likely **non-anion gap metabolic acidosis** if RTA or diarrhea.<br><br>6. **Autoimmune screening if RTA is supported**: ANA, anti-SSA/SSB, complements<br>- **Purpose:** identify secondary causes such as Sjögren syndrome.<br>- **Expected:** may be positive if autoimmune distal RTA is present.<br><br>7. **Renal ultrasound**<br>- **Purpose:** assess kidney structure, obstruction, nephrocalcinosis/nephrolithiasis. |

| | |
|---|---|
| | - **Expected:** nephrocalcinosis would support distal RTA; also helps assess chronic kidney disease.<br><br>Overall, the diagnostic approach should first address the **immediate danger of profound hypokalemia**, then distinguish **renal vs extrarenal potassium loss**, with **RTA as the leading unifying diagnosis**. |
| With Static RAG (Model B) | 4. **Tests to narrow the diagnosis**<br><br>### Immediate tests<br>- **ECG**<br>- Purpose: assess for hypokalemia-related arrhythmia.<br>- Expected findings: **U waves, ST depression, flattened T waves, QT/QU prolongation**.<br><br>- **Repeat basic metabolic panel, magnesium, phosphate**<br>- Purpose: confirm severity, detect associated abnormalities that worsen potassium repletion.<br>- Expected: persistent severe hypokalemia; possible **hypomagnesemia**.<br><br>### To determine cause of metabolic acidosis and potassium loss<br>- **Arterial or venous blood gas**<br>- Purpose: confirm metabolic acidosis and assess compensation.<br>- Expected: metabolic acidosis, likely **non-anion gap** if RTA or diarrhea.<br><br>- **Serum anion gap**<br>- Purpose: classify acidosis.<br>- Expected: likely **normal anion gap metabolic acidosis**.<br><br>- **Urinalysis including urine pH**<br>- Purpose: evaluate tubular function, proteinuria, hematuria, glycosuria.<br>- Expected:<br>- In **distal RTA**, urine pH may remain **>5.5** despite systemic acidosis.<br>- Proteinuria may suggest intrinsic kidney disease contributing to edema.<br><br>- **Urine electrolytes (urine K, Na, Cl) and calculation of urine anion gap or urinary ammonium surrogate**<br>- Purpose: distinguish **renal** from **extrarenal** acid loss and assess renal K wasting.<br>- Expected:<br>- **Positive urine anion gap** in RTA.<br>- Inappropriate urinary potassium excretion if renal potassium wasting is present.<br><br>- **Urine potassium-to-creatinine ratio or 24-hour urinary potassium**<br>- Purpose: confirm renal potassium wasting.<br>- Expected: inappropriately elevated if RTA or diuretic effect.<br><br>### To evaluate edema and kidney involvement<br>- **Urine protein quantification and serum albumin**<br>- Purpose: assess nephrotic-range protein loss or hypoalbuminemia as cause of edema.<br>- Expected: may help determine whether edema is renal/protein-related.<br><br>- **Renal ultrasound** |

| | |
|---|---|
| | - Purpose: evaluate kidney size, obstruction, nephrocalcinosis, stones.<br>- Expected:<br>- **Nephrocalcinosis/stones** would support distal RTA.<br><br>### Etiologic evaluation if RTA is suspected<br>- **Autoimmune serologies: ANA, anti-SSA/SSB**<br>- Purpose: assess for **Sjögren syndrome** or other autoimmune causes.<br>- Expected: may be positive if secondary distal RTA.<br><br>- **Plasma renin/aldosterone only if clinically indicated**<br>- Purpose: evaluate mineralocorticoid disorders if urinary data suggest renal K wasting not explained by RTA.<br>- Expected: less likely primary explanation here because **metabolic acidosis** argues against classic hyperaldosteronism.<br><br>Overall, the most efficient diagnostic approach is to first confirm the **type of acidosis**, determine whether potassium loss is **renal vs extrarenal**, and simultaneously assess for **urgent hypokalemia complications**. |
| With Static RAG and Mult-agent framework (Model C) | ## 4. Testing plan<br><br>### Immediate / highest priority tests<br><br>1. **ECG and continuous telemetry**<br>- Purpose: detect hypokalemia-induced arrhythmia.<br>- Why first: **K 1.2 mEq/L is an emergency**.<br><br>2. **Repeat serum chemistry**<br>- **Na, K, Cl, HCO3−, BUN, creatinine, glucose, Mg, Ca, phosphate**<br>- Purpose: confirm severity, identify associated deficits, and allow **serum anion gap calculation**.<br>- Change from initial version: **explicitly prioritizes anion gap calculation before labeling this as normal-anion-gap acidosis**.<br><br>3. **Blood gas (venous or arterial)**<br>- Purpose: confirm metabolic acidosis and physiologic compensation.<br><br>### Next tests to localize potassium loss and acidosis source<br><br>4. **Spot urine potassium, creatinine, and potassium-to-creatinine ratio**<br>- Purpose: determine whether potassium loss is **renal**.<br>- Change from initial version: **de-emphasizes TTKG**, which is less reliable in severe hypokalemia and variable urine concentration states.<br><br>5. **Urine pH and urine electrolytes (including urine sodium, potassium, chloride)**<br>- Purpose: assess renal acidification and help distinguish **RTA** from GI loss.<br><br>6. **Urine anion gap**<br>- Purpose: supports distinction between **RTA** and **extrarenal bicarbonate loss**, though interpreted together with the full clinical picture.<br><br>### Early edema/systemic disease workup<br><br>7. **Urinalysis with urine protein quantification** |

- Purpose: look for **proteinuria, hematuria**, and evidence of intrinsic renal disease.
- Reason: edema is not explained adequately by isolated RTA.

8. **Serum albumin and liver function tests**
- Purpose: evaluate **hypoalbuminemia** or liver disease as causes of edema.

9. **Chest X-ray and/or BNP if symptoms suggest cardiac disease**
- Purpose: assess for **heart failure** if clinically indicated.

#### Second-line tests depending on initial results

10. **Renal ultrasound**
- Purpose: assess kidney structure, obstruction, and possible **nephrocalcinosis/nephrolithiasis** suggestive of distal RTA.

11. **Autoimmune serologies**
- **ANA, anti-SSA/Ro, anti-SSB/La, rheumatoid factor**
- Purpose: evaluate secondary causes of distal RTA such as **Sjögren syndrome**.
- Change from initial version: these are moved **after** basic electrolyte, urine, and edema evaluation rather than being pursued too early.

---

### Changes from the initial version

1. **Stronger emphasis on emergency management**
- I now explicitly state that **K 1.2 mEq/L is a life-threatening emergency** requiring **urgent potassium replacement and cardiac monitoring**.

2. **Avoided presuming normal-anion-gap acidosis too early**
- The revised version does **not assume** the acidosis is normal anion gap before obtaining **Na and Cl** and calculating the **serum anion gap**.

3. **Broadened and reprioritized the differential**
- **Medication-related causes** are now placed higher because of the long antihypertensive history.
- **CKD/tubulointerstitial disease** is added as a possible contributor.
- The **edema differential** is addressed more explicitly in parallel.

4. **Separated RTA more carefully**
- The revision distinguishes **distal RTA** from **proximal RTA** rather than grouping them too loosely.

5. **Improved history and exam focus**
- The medication history is now more specific: **exact drugs, doses, OTC agents, herbals, licorice, laxatives**.
- The physical exam now gives greater priority to **neuromuscular and respiratory assessment** because of the extreme hypokalemia.

6. **Refined the testing plan**
- **ECG/telemetry, repeat chemistry, magnesium assessment, and anion gap calculation** are prioritized first.
- **Spot urine potassium-to-creatinine ratio** is favored over **TTKG**.

| | |
|---|---|
| | - **Edema workup** with **urinalysis/protein quantification, albumin, and hepatic/cardiac assessment** is moved earlier.<br>- **Autoimmune serologies** are deferred until after the basic diagnostic framework is established.<br><br>Overall, the revised version is **more cautious diagnostically, more explicit about urgency, broader in differential diagnosis, and more logically prioritized in testing**. |
| **Phase II – Q5: Interpreting test results** | |
| Baseline LLM (Model A) | **5. Interpretation of the test results**<br><br>- **Normal anion gap hyperchloremic metabolic acidosis with hypokalemia** indicates **renal tubular acidosis (RTA)** rather than vomiting or mineralocorticoid excess.<br>- **Urine pH 8.0** shows **inappropriately alkaline urine despite systemic acidosis**, supporting a **distal urinary acidification defect**.<br>- At the same time, **renal glycosuria (despite normal blood glucose), pan-aminoaciduria, hypophosphatemia with low tubular phosphate reabsorption, hypouricemia, markedly elevated urinary β2-microglobulin/NAG, and tubular proteinuria** indicate **generalized proximal tubular dysfunction (Fanconi syndrome)**.<br>- Therefore, the patient has **combined proximal and distal tubular dysfunction**, not isolated distal RTA alone.<br>- **Osteomalacia** is well explained by **phosphate wasting and chronic acidosis**, and the very high **ALP** supports active metabolic bone disease.<br>- **Renal biopsy** shows **tubulointerstitial nephritis (TIN)** without glomerulonephritis, which fits the urinary pattern of tubular injury and the reduced eGFR.<br>- The particularly important finding is that the infiltrate is mainly **IgM-positive CD38+ plasma cells**, together with a markedly elevated **serum IgM (1721 mg/dL)**. This suggests an **IgM-producing plasma cell/lymphoplasmacytic process involving the kidney**, rather than typical autoimmune TIN.<br>- Autoimmune serologies (ANA, SSA/SSB, ANCA, AMA, anti-M2) are negative, making common autoimmune causes such as Sjögren syndrome or PBC less likely based on the provided data. |
| With Static RAG (Model B) | **5. Interpretation of the test results**<br><br>- **Normal anion gap hyperchloremic metabolic acidosis + hypokalemia + alkaline urine (pH 8.0)**<br>This strongly indicates **renal tubular acidosis (RTA)** rather than gastrointestinal bicarbonate loss. The persistently alkaline urine despite systemic acidosis is especially characteristic of **distal acidification failure**.<br><br>- **Pan-aminoaciduria, renal glycosuria, hypophosphatemia, hypouricemia, and low tubular reabsorption of phosphate (43.4%)**<br>These show a **generalized proximal tubular reabsorptive defect (Fanconi syndrome)**. Renal glycosuria with normal blood glucose is particularly important because it confirms a tubular, not diabetic, cause. Hypophosphatemia from phosphate wasting explains the **osteomalacia**, and the markedly elevated **ALP** supports active bone disease.<br><br>- **Marked urinary β2-microglobulin and NAG elevation**<br>These are strong markers of **tubular injury**, supporting a primary |

| | |
|---|---|
| | **tubulointerstitial process** rather than glomerular disease.<br><br>- **Proteinuria (2.32 g/gCr) with little hematuria and biopsy showing no glomerulonephritis**<br>The proteinuria is most likely **tubular proteinuria**, consistent with proximal tubular damage, not primary glomerular disease.<br><br>- **Renal biopsy: tubulointerstitial nephritis with lymphocyte/plasma cell infiltration, fibrosis, and tubular atrophy**<br>This confirms **chronic tubulointerstitial nephritis (TIN)** as the structural basis of the tubular dysfunction and CKD.<br><br>- **IgM-positive CD38+ plasma cell predominance in the interstitium + very high serum IgM (1721 mg/dL)**<br>This is a particularly important clue. It suggests an **IgM-positive plasma cell–rich TIN**, a rare but recognized entity often associated with **distal RTA/Fanconi-like tubular dysfunction** and sometimes linked to **primary biliary cholangitis (PBC)** or other immune-mediated disease.<br><br>- **Negative ANA, SSA/SSB, ANCA, AMA, anti-M2**<br>These make Sjögren syndrome, ANCA-associated disease, and serologically evident PBC less likely, but do **not completely exclude** an immune-mediated TIN.<br><br>- **Renal dysfunction (Cr 1.41, eGFR 29.3), hypoalbuminemia, anemia**<br>These indicate clinically significant chronic kidney involvement and systemic illness/nutritional impact.<br><br>**Overall meaning:**<br>The data do **not** fit isolated distal RTA alone. They indicate **combined tubular dysfunction**: distal acidification failure plus generalized proximal tubular dysfunction (**Fanconi syndrome**) caused by **chronic immune-mediated tubulointerstitial nephritis**. |
| With Static RAG and Mult-agent framework (Model C) | **5. Interpretation of the test results**<br><br>- The updated chemistry confirms a **normal anion gap hyperchloremic metabolic acidosis** (**Na 139, Cl 111, HCO3− 18.2; anion gap ≈ 10**), which strongly supports **renal tubular acidosis (RTA) or gastrointestinal bicarbonate loss** rather than a high-anion-gap acidosis.<br>- However, because these results were obtained **while she was receiving potassium and sodium bicarbonate supplementation**, the **urine pH of 8.0** should be interpreted cautiously. It is **compatible with impaired urinary acidification**, but **does not by itself prove distal RTA** under alkali therapy.<br>- In contrast, the evidence for **proximal tubular dysfunction (Fanconi syndrome)** is strong:<br>- **Pan-aminoaciduria**<br>- **Renal glycosuria despite normal blood glucose/HbA1c**<br>- **Hypophosphatemia (IP 1.8 mg/dL)**<br>- **Hypouricemia (UA 2.2 mg/dL)**<br>- **Low tubular reabsorption of phosphate (43.4%)**<br>- Markedly elevated **urinary β2-microglobulin** and **NAG**, indicating tubular injury<br>- Thus, the test pattern firmly supports **Fanconi syndrome due to tubular injury**, with **RTA present**, while the exact degree of **distal acidification defect remains |

| | |
|---|---|
| | suspected rather than definitively proven** from the urine pH alone.<br>- **Proteinuria (2.32 g/gCr)** with **minimal hematuria** and very high tubular injury markers suggests that a substantial component is **tubular proteinuria**. Still, the **amount of proteinuria and hypoalbuminemia (albumin 2.7 g/dL)** mean a **glomerular contribution cannot be completely excluded**, especially since the biopsy sampled only **5 glomeruli**.<br>- **Bone scintigraphy showing osteomalacia**, together with **hypophosphatemia** and markedly elevated **ALP (1378 IU/L)**, is clinically explained by **chronic phosphate wasting and acidosis** from proximal tubular dysfunction.<br>- The renal biopsy is highly informative: it shows **tubulointerstitial nephritis (TIN)** with **lymphocyte/plasma cell infiltration, interstitial fibrosis, and tubular atrophy**, and **no glomerulonephritis** in the sampled tissue. This provides the structural explanation for the tubular abnormalities and indicates that the process is already **chronic**, not purely functional.<br>- The infiltrate being mainly **IgM-positive CD38+ plasma cells**, together with markedly elevated **serum IgM (1721 mg/dL)**, makes **IgM-positive plasma cell–rich TIN** a compelling unifying diagnosis, though this pattern is **suggestive rather than absolutely specific** on its own.<br>- Negative **ANA, SSA/SSB, ANCA, AMA, anti-M2** make common autoimmune causes less supported by the provided data, but they **do not fully exclude seronegative autoimmune TIN**, including **seronegative Sjögren-related disease**.<br>- **Anemia (Hb 9.8 g/dL)**, **reduced eGFR (29.3 mL/min/1.73 $m^2$)**, and **interstitial fibrosis/tubular atrophy** indicate **established chronic kidney injury**. |
| **Phase II – Q6: Reconsidering the differential diagnosis** | |
| Baseline LLM (Model A) | **6. Updated diagnostic thinking / Working diagnosis**<br><br>The differential now shifts strongly away from simple drug effect or isolated classic RTA and toward **Fanconi syndrome with distal RTA due to tubulointerstitial nephritis**.<br><br>**Most likely working diagnosis:**<br>**Acquired Fanconi syndrome with distal renal tubular acidosis caused by IgM-positive plasma cell–rich tubulointerstitial nephritis**, i.e. a **monoclonal gammopathy/lymphoplasmacytic disorder–related renal tubulointerstitial lesion**.<br><br>Why this is now most likely:<br>- Proximal tubular dysfunction: glycosuria, aminoaciduria, phosphaturia, hypouricemia.<br>- Distal tubular dysfunction: alkaline urine despite metabolic acidosis.<br>- Pathology localizes disease to the tubulointerstitium.<br>- IgM-predominant plasma cell infiltration plus high serum IgM points to an **IgM-related lymphoplasmacytic/plasma cell disorder** as the underlying cause. |
| With Static RAG (Model B) | **6. Updated diagnostic thinking / Working diagnosis**<br><br>### Revised prioritization<br>1. **Tubulointerstitial nephritis causing Fanconi syndrome with renal tubular acidosis**<br>2. **Immune-mediated plasma cell–rich TIN, especially IgM-positive plasma cell TIN**<br>3. Less likely: Sjögren syndrome, drug-induced TIN, monoclonal gammopathy-related tubulopathy, or other systemic autoimmune disease |

| | |
|---|---|
| | ### Most likely diagnosis (Working Diagnosis)<br>**IgM-positive plasma cell tubulointerstitial nephritis causing Fanconi syndrome and renal tubular acidosis (likely mixed proximal and distal tubular dysfunction), with hypophosphatemic osteomalacia and CKD.**<br><br>### Why this is now most likely<br>- Proximal tubular defects: **glycosuria, aminoaciduria, phosphate wasting, hypouricemia**<br>- Distal tubular defect: **alkaline urine despite hyperchloremic metabolic acidosis**<br>- Histology confirms **TIN**<br>- Immunostaining specifically shows **IgM-positive CD38+ plasma cell infiltration**<br>- High serum **IgM** supports this immune-plasma-cell process<br><br>So the diagnosis has shifted from “possible RTA” to a much more specific **immune-mediated TIN with Fanconi syndrome**, most consistent with **IgM-positive plasma cell TIN**. |
| With Static RAG and Mult-agent framework (Model C) | **6. Updated diagnostic thinking / Working diagnosis**<br><br>### **Most likely diagnosis**<br>**Tubulointerstitial nephritis with Fanconi syndrome and renal tubular acidosis, likely IgM-positive plasma cell tubulointerstitial nephritis (IgMPC-TIN), complicated by hypokalemia, chronic kidney dysfunction, and osteomalacia.**<br><br>### Why this is now the leading diagnosis<br>- The biochemical and urinary findings are classic for **Fanconi syndrome**:<br> - phosphaturia / low TRP<br> - glycosuria without hyperglycemia<br> - aminoaciduria<br> - hypouricemia<br> - tubular injury markers markedly elevated<br>- The acid-base pattern confirms **RTA physiology** in the setting of tubular disease.<br>- The biopsy proves **TIN** as the major renal lesion and does **not show glomerulonephritis** in the sampled tissue.<br>- **IgM-positive plasma cell predominance** in the interstitium plus **high serum IgM** makes **IgMPC-TIN** the most plausible specific subtype.<br><br>### Residual diagnostic uncertainty<br>- I would revise the earlier wording from **“mixed proximal and distal RTA is established”** to **“Fanconi syndrome is established, and a distal acidification defect is suspected but not definitively proven by urine pH alone during bicarbonate therapy.”**<br><br>- I would also soften **“most likely IgMPC-TIN”** to **“likely IgMPC-TIN”**, because:<br> - the biopsy sample is small (**5 glomeruli**),<br> - the immunostaining pattern is supportive but not perfectly specific,<br> - and **seronegative autoimmune TIN**, including **Sjögren-related TIN**, remains a possible alternative.<br><br>### Differential diagnosis after the new data<br>1. **TIN with Fanconi syndrome, likely IgMPC-TIN** — most likely<br>2. **Seronegative autoimmune TIN (including Sjögren-related TIN)** — still |

<table>
<tr><td></td><td>possible despite negative antibodies<br>3. **Drug-induced TIN/Fanconi syndrome** — still important to review carefully from the medication history<br>4. **Primary glomerular disease** — less likely, but not fully excluded because of the degree of proteinuria/hypoalbuminemia and limited biopsy sample</td></tr>
<tr><td colspan="2">Phase II – Q7: Treatment planning</td></tr>
<tr><td>Baseline LLM (Model A)</td><td>**7. Treatment plan**<br><br>**A. Immediate/supportive treatment**<br>- **Continue potassium supplementation** and **alkali therapy** (sodium bicarbonate or potassium citrate if tolerated/appropriate).<br>- Rationale: correct hypokalemia and chronic acidosis, reduce ongoing bone buffering, and improve symptoms.<br>- **Phosphate replacement** and management of **osteomalacia**; consider **active vitamin D** if clinically appropriate.<br>- Rationale: phosphate wasting is severe and has already caused osteomalacia.<br>- Monitor **K, HCO3−, phosphate, creatinine, calcium**, and urine findings closely.<br><br>**B. Treat the underlying renal inflammatory/plasma cell process**<br>- **Systemic corticosteroid therapy** would be a reasonable first-line treatment for active **tubulointerstitial nephritis**, especially with declining kidney function and biopsy-proven inflammatory infiltrates.<br>- Rationale: TIN is often steroid-responsive, and early treatment may preserve renal function and reduce tubular dysfunction.<br>- Because the infiltrate is **IgM-positive plasma cell rich**, the patient should also undergo **hematologic evaluation** for an **IgM monoclonal process** (e.g., lymphoplasmacytic disorder/Waldenström spectrum or related monoclonal gammopathy).<br>- If a clonal disorder is confirmed, **clone-directed therapy** may be needed in addition to or instead of steroids.<br><br>**C. Other options / alternatives**<br>- If steroids are ineffective or relapse occurs: consider **other immunosuppressive therapy**, depending on final etiologic classification and tolerance.<br>- If a definite **B-cell/plasma cell clone** is identified, treatment should be tailored to that disorder rather than treating this as nonspecific TIN alone.<br><br>In summary, treatment should combine:<br>1. **Aggressive correction of electrolyte/acid-base abnormalities**,<br>2. **Bone disease management**, and<br>3. **Etiology-directed therapy for IgM-positive plasma cell–rich TIN / possible monoclonal lymphoplasmacytic disease**.</td></tr>
<tr><td>With Static RAG (Model B)</td><td>**7. Treatment plan**<br><br>### Main treatment<br>1. **Correct electrolyte and acid-base abnormalities**<br>- **Potassium supplementation**<br>- **Alkali therapy** (sodium bicarbonate or potassium citrate)<br>- **Phosphate replacement**<br>- Consider **active vitamin D** if needed for osteomalacia/mineral bone disease<br><br>**Rationale:**</td></tr>
</table>

| | |
|---|---|
| | This treats the immediate consequences of tubular dysfunction: hypokalemia, acidosis, phosphate wasting, and bone disease.<br><br>2. **Treat the underlying tubulointerstitial inflammation**<br>- **Systemic corticosteroid therapy** is the most reasonable disease-directed treatment<br><br>**Rationale:**<br>Biopsy-proven immune/plasma cell-rich TIN is potentially steroid-responsive, and treatment may improve tubular function, kidney function, and reduce ongoing interstitial damage if fibrosis is not fully irreversible.<br><br>3. **Supportive CKD care**<br>- Monitor **creatinine/eGFR, bicarbonate, potassium, phosphate, urinary findings**<br>- Avoid **nephrotoxic drugs**<br>- Nutritional support and edema assessment<br><br>### Additional evaluation linked to treatment<br>- Reassess for associated immune disease, especially **PBC or other autoimmune disorders**, because this may influence long-term management, even though current serologies are negative.<br>- If a culprit drug is identified, **stop it immediately**.<br><br>### Other possible options<br>- If steroid response is poor or relapse occurs, **other immunosuppressive agents** may be considered on a case-by-case basis, though evidence is limited.<br>- If edema is clinically significant, use diuretics cautiously because they may worsen electrolyte losses.<br><br>**In summary:**<br>The treatment should combine **aggressive replacement of potassium, bicarbonate, and phosphate**, management of **osteomalacia**, and **steroid therapy for biopsy-proven immune-mediated IgM-positive plasma cell TIN**, while monitoring renal recovery and complications. |
| With Static RAG and Mult-agent framework (Model C) | **7. Treatment plan**<br><br>### A. Immediate and supportive management<br>1. **Continue replacement of ongoing tubular losses**<br>- **Potassium supplementation** for hypokalemia<br>- **Sodium bicarbonate or other alkali therapy** for metabolic acidosis<br>- **Careful phosphate replacement** for hypophosphatemia, especially if persistent or symptomatic<br>- **Rationale:** these abnormalities are direct consequences of tubular wasting and are responsible for weakness, arrhythmia risk, and bone disease.<br><br>2. **Treat osteomalacia / mineral-bone complications**<br>- Assess and monitor **phosphate, calcium, ALP**, and ideally broader bone-mineral parameters if available<br>- Consider **vitamin D support / active vitamin D** depending on full mineral metabolism status and kidney function<br>- **Rationale:** osteomalacia is already documented and likely driven by **phosphate wasting plus chronic acidosis**. |

#### B. Disease-directed therapy
3. **Review and stop any possible causative drugs**
   - Reassess all prescribed antihypertensives and any nonprescription/herbal agents
- **Rationale:** drug-induced TIN is a potentially reversible cause and should be addressed before or alongside immunosuppression.

4. **Corticosteroid therapy is a reasonable first-line disease-directed treatment**
   - Because biopsy shows **active inflammatory TIN with plasma cell infiltration**, steroids are a standard and evidence-based initial therapy for immune-mediated TIN
- **Rationale:** the goal is to reduce ongoing interstitial inflammation and preserve remaining renal function, recognizing that **existing fibrosis/tubular atrophy may limit reversibility**.

5. **Assess steroid response carefully**
   - Follow **creatinine/eGFR, bicarbonate, potassium, phosphate, proteinuria, urinary tubular markers, and symptoms**
   - Reassess whether the biochemical abnormalities and kidney function improve after treatment
- **Rationale:** response assessment is important both therapeutically and diagnostically.

#### C. Monitoring and patient-specific considerations
6. **Close monitoring**
   - **K, HCO3−, phosphate, calcium, creatinine/eGFR**
   - Urine findings: **proteinuria, glycosuria, tubular markers**
   - **Volume status** and edema
   - **ALP** and clinical bone symptoms
- **Rationale:** this patient has CKD, active tubular wasting, and bone disease, so over- or under-replacement can be harmful.

7. **Steroid risk monitoring**
   - Because she is **70 years old** and already has chronic kidney damage, monitor for steroid adverse effects such as:
     - infection
     - worsening edema/fluid retention
     - hyperglycemia
     - bone effects
- **Rationale:** the benefit of immunosuppression must be balanced against age and comorbidity.

#### D. Alternatives / next steps if response is incomplete
8. If steroid response is poor or relapse occurs:
   - reconsider **alternative etiologies** such as seronegative autoimmune disease or drug-related disease
   - consider additional immunosuppressive therapy only based on the confirmed course and risk-benefit assessment
- **Rationale:** the available data support immune-mediated TIN, but some uncertainty remains.

---

#### Changes from the initial results
- I have **strengthened the diagnosis of Fanconi syndrome**, because the new data clearly show **aminoaciduria, glycosuria without hyperglycemia, phosphaturia,

<table>
<tr><td></td><td>hypophosphatemia, and hypouricemia**.<br>- I have **made the distal RTA claim more cautious**: previously it was described as established from alkaline urine, but now I would say **distal acidification defect is suspected, not definitively proven**, because the urine was measured while she was taking **bicarbonate**.<br>- I have **softened the specificity of IgMPC-TIN**: it remains the **leading diagnosis**, but I now state **“likely IgMPC-TIN”** rather than presenting it as certain.<br>- I have **added residual uncertainty** regarding:<br>  - **seronegative autoimmune TIN**<br>  - **drug-induced TIN**<br>  - and a **possible glomerular contribution** to proteinuria/hypoalbuminemia<br>- I have **expanded the treatment plan** to emphasize:<br>  - careful **phosphate and bone management**<br>  - explicit **drug review**<br>  - and **steroid monitoring/response assessment** in an older patient with existing fibrosis and CKD.</td></tr>
<tr><td colspan="2">Phase III – Q8: Evaluation of treatment</td></tr>
<tr><td>Baseline LLM (Model A)</td><td>**8. Monitoring of treatment effectiveness and safety**<br><br>**Effectiveness**<br>- **Kidney response:** serial **serum creatinine/eGFR**, urinalysis, urinary **β2-microglobulin/NAG**, proteinuria, glycosuria.<br>  - Improvement would mean less active tubulointerstitial injury.<br>- **Tubular function:** monitor **K, HCO3−, Cl, phosphate, uric acid, urine pH**, and need for potassium/alkali supplementation.<br>  - Recovery of hypokalemia, acidosis, phosphaturia, and glycosuria indicates treatment response.<br>- **Bone/mineral disease:** **phosphate, calcium, ALP**, symptoms of bone pain/weakness.<br>- **IgM-related activity:** serial **serum IgM** levels.<br>- **Liver involvement:** because cholestatic enzymes were abnormal from the start, monitor **AST/ALT, ALP, γ-GT, bilirubin, albumin, PT-INR, ammonia**, and check for **ascites, edema, encephalopathy**.<br><br>**Safety / complications to watch for**<br>- **Steroid toxicity:** infection, hyperglycemia, hypertension/fluid retention, psychiatric symptoms, myopathy, gastrointestinal bleeding, osteoporosis.<br>  - **Countermeasures:** regular vitals, glucose monitoring, CBC, infection surveillance, consider gastric and bone protection as appropriate, taper promptly if risks outweigh benefit.<br>- **Electrolyte therapy risks:** overcorrection causing **hyperkalemia**, metabolic alkalosis, sodium/fluid overload from bicarbonate.<br>  - **Countermeasures:** frequent electrolyte and acid-base checks; adjust doses to labs and volume status.<br>- **Progressive liver failure / portal hypertensive complications:** worsening jaundice, coagulopathy, hyperammonemia, ascites, spontaneous bacterial peritonitis.<br>  - **Countermeasures:** repeated liver tests, diagnostic paracentesis when ascites appears, standard encephalopathy treatment, infection screening/cultures, early antibiotics if SBP suspected.<br>- **Missed systemic IgM-positive plasma cell disease:** persistence/progression despite renal response.<br>  - **Countermeasures:** reassess whole-organ involvement if IgM remains elevated</td></tr>
</table>

| | |
|---|---|
| | or new organ dysfunction appears. |
| With Static RAG (Model B) | 8. **Monitoring effectiveness, safety, risks, and countermeasures**<br><br>**A. Effectiveness monitoring**<br>- **Kidney response**<br> - Follow **serum creatinine/eGFR**, urinalysis, urinary **β2-microglobulin/NAG**, proteinuria, glycosuria, and urine pH.<br> - Goal: improvement in tubulointerstitial injury and stabilization/improvement of renal function.<br>- **Tubular dysfunction / Fanconi syndrome**<br> - Serial **K, HCO3−, phosphate, uric acid, calcium**, and if needed urinary phosphate loss.<br> - Goal: correction of **hypokalemia, metabolic acidosis, phosphate wasting**, and improvement of osteomalacia/mineral abnormalities.<br>- **Immunologic activity**<br> - Follow **serum IgM** trend.<br> - Goal: decrease in IgM as a marker of response, as occurred after prednisolone.<br>- **Liver involvement**<br> - Because this case later developed fatal liver disease, monitor **AST/ALT, ALP, γ-GT, bilirubin, albumin, PT-INR, ammonia (if mental status changes)**, and assess for **ascites/encephalopathy** clinically and by imaging when indicated.<br> - Goal: early detection of evolving hepatobiliary/portal venous disease even if autoantibodies are negative.<br><br>**B. Safety monitoring for corticosteroids and supportive therapy**<br>- **Infection**<br> - Risk is high with **prednisolone 40 mg/day**, especially in an elderly patient; spontaneous bacterial peritonitis eventually occurred.<br> - Monitor **fever, WBC/CRP, new abdominal pain, worsening ascites, respiratory/urinary symptoms**.<br> - Countermeasures: low threshold for cultures and imaging, prompt empiric antibiotics when infection is suspected, careful ascites evaluation/paracentesis if present.<br>- **Fluid retention / worsening edema or ascites**<br> - Steroids may worsen sodium/fluid retention.<br> - Monitor **body weight, edema, abdominal girth, blood pressure**.<br> - Countermeasures: sodium restriction, cautious diuretic use if needed, reassess volume status frequently.<br>- **Metabolic adverse effects**<br> - Monitor **glucose**, electrolytes, and acid-base status.<br> - Countermeasures: adjust supplements; treat steroid-induced hyperglycemia if it appears.<br>- **Electrolyte over- or under-correction**<br> - Potassium bicarbonate/phosphate therapy can lead to rebound abnormalities.<br> - Monitor **K, HCO3−, phosphate** frequently early in treatment.<br> - Countermeasures: titrate replacement based on serial labs.<br>- **Bone and muscle toxicity of steroids**<br> - She already had osteomalacia; steroids increase fracture risk.<br> - Monitor symptoms, calcium/phosphate, and support bone health.<br>- **GI bleeding / ulcer risk and neuropsychiatric effects**<br> - Monitor for abdominal pain, melena, confusion, sleep or mood changes.<br>- **Progressive liver failure despite renal response**<br> - This is a key disease-specific risk in this case. |

| | |
|---|---|
| | - Countermeasures: if liver tests worsen or ascites/encephalopathy appears, urgently reassess the diagnosis and involve hepatology; do not assume all abnormalities are steroid-related or benign. |
| With Static RAG and Mult-agent framework (Model C) | 8. **Monitoring effectiveness and safety; risks and countermeasures**<br><br>**A. Effectiveness monitoring**<br><br>- **Renal response**<br>- Follow **serum creatinine/eGFR**, urine protein, urinary **β2-microglobulin/NAG**, glycosuria, and serum/urine electrolytes.<br>- In this case, effectiveness was reflected by improved renal insufficiency, less tubular injury, and recovery from mineral abnormalities after prednisolone.<br><br>- **Tubular dysfunction / Fanconi syndrome**<br>- Monitor **K, HCO3−, phosphate, uric acid, Ca**, urine pH, glucosuria, and supplementation requirements.<br>- Clinical goals are correction of **hypokalemia**, **metabolic acidosis**, **hypophosphatemia**, and improvement of osteomalacia/mineral disorders.<br><br>- **Immunologic activity**<br>- Follow **serum IgM only as a supportive marker**, not as a disease-specific marker.<br>- In this case, the decrease in IgM from **1721 to 1026 mg/dL** paralleled renal improvement and reduced renal IgM-positive plasma-cell infiltration at autopsy.<br><br>- **Liver involvement**<br>- Because this disorder may involve the liver even when **antimitochondrial antibodies are negative**, monitor **AST/ALT, ALP, γ-GT, bilirubin, albumin, PT-INR**, and assess for **abdominal distension, ascites, edema, and encephalopathy**.<br>- Do not rely only on modest liver enzyme abnormalities: **portal venopathy/nodular regenerative hyperplasia may progress despite limited transaminase elevation**.<br>- Serial assessment should include **bilirubin, INR, albumin, ammonia**, and **abdominal imaging** if symptoms or abdominal findings change.<br><br>**B. Safety monitoring**<br><br>- **Corticosteroid toxicity**<br>- Watch for **infection**, hyperglycemia, fluid retention, hypertension, psychiatric symptoms, GI bleeding, myopathy, and osteoporosis.<br>- Countermeasures:<br>- frequent review of symptoms, **CBC/glucose**<br>- low threshold for cultures/imaging when infection is suspected<br>- bone protection and fall prevention<br>- reassess dose/taper according to benefit and adverse effects<br><br>- **Electrolyte and alkali replacement risks**<br>- Monitor for **hyperkalemia**, **metabolic alkalosis**, sodium/fluid overload, and worsening edema or ascites.<br>- Countermeasures:<br>- serial electrolytes and acid-base monitoring<br>- adjust potassium/bicarbonate promptly<br>- careful diuretic use if fluid overloaded |

| | |
|---|---|
|  | - **Progressive liver failure / portal venous disease**<br> - Specific risks include **hepatic encephalopathy, ascites, spontaneous bacterial peritonitis (SBP), and rapid liver atrophy**.<br> - Countermeasures:<br> - repeated liver tests and abdominal examination<br> - **urgent hepatology-level reassessment** if new ascites, encephalopathy, or liver atrophy develops<br> - **diagnostic paracentesis for all new ascites**, including **cell count/PMN count, culture, and albumin**<br> - prompt treatment of encephalopathy and close infection surveillance<br><br>- **Infection risk in ascites and under immunosuppression**<br> - Watch for fever, abdominal pain, leukocytosis, worsening mental status, and renal deterioration.<br> - Countermeasures:<br> - immediate paracentesis when ascites appears<br> - **early empiric antibiotics if SBP is suspected**<br> - reconsider steroid risk-benefit during severe infection or hepatic decompensation |
| **Phase III – Q9: Formulating plans for worsening conditions** | |
| Baseline LLM (Model A) | **9. If the condition does not improve or suddenly changes**<br><br>**Reassessment framework**<br>1. **Confirm whether the original target disease is actually responding**<br> - In this case, the kidney improved, so new deterioration should prompt suspicion of **another active organ process**, not simply "steroid failure."<br><br>2. **Reframe the problem by organ and syndrome**<br> - New **ascites + encephalopathy + liver atrophy + hyperammonemia** suggests **acute/subacute hepatic failure/portal venous disorder**, not a renal relapse.<br><br>3. **Reconsider the differential broadly**<br> - For sudden liver deterioration, reassess for:<br> - **Drug-induced liver injury**<br> - **Infection/sepsis**<br> - **Viral hepatitis**<br> - **Autoimmune/cholestatic liver disease**<br> - **Vascular disorders** such as **obliterative portal venopathy / nodular regenerative hyperplasia**<br> - **Direct hepatic infiltration by the same IgM-positive plasma cell process**<br><br>4. **Act urgently while re-evaluating**<br> - Repeat **liver panel, PT-INR, ammonia, renal function, CBC**, abdominal imaging, and **diagnostic paracentesis** for new ascites.<br> - Search for precipitating factors of encephalopathy and infection.<br> - Review all medications, including steroids and supportive drugs.<br><br>5. **Revise diagnosis and treatment according to new data**<br> - If evidence points to systemic IgM-positive plasma cell infiltration or portal venopathy, do not assume isolated renal disease; revise to a **multiorgan IgM-related disorder**.<br> - If steroids are insufficient or disease is progressing in another organ, consider need for **disease reclassification and alternative therapy**, while continuing organ-supportive care. |

| | |
|---|---|
| | **Practical principle:**<br>When the expected renal response occurs but a different organ rapidly fails, the correct approach is to **separate treatment response from overall disease control**, urgently **re-stage the disease**, and **change the working diagnosis from isolated kidney disease to systemic involvement** until proven otherwise. |
| With Static RAG (Model B) | 9. **If the condition does not improve as expected or suddenly changes: reassessment and action plan**<br><br>**A. Reassess the problem from first principles**<br>- Separate whether the change reflects:<br>1. **Persistent/relapsing IgMPC-TIN**<br>2. **Treatment complication** (especially infection or steroid toxicity)<br>3. **A new organ manifestation of the same systemic immune process**, particularly **liver involvement**<br>4. **An alternative diagnosis** missed initially<br><br>**B. If renal response is inadequate**<br>- Recheck adherence and adequacy of **potassium/alkali/phosphate replacement**.<br>- Repeat assessment of **urinary tubular markers, creatinine, proteinuria, urine sediment**.<br>- Reconsider competing causes of TIN/Fanconi syndrome: **drug-induced disease, other autoimmune disease, hematologic/plasma cell disorder**.<br>- If needed, discuss whether immunosuppression should be adjusted, but only after excluding infection.<br><br>**C. If there is sudden deterioration, especially abdominal distension, ascites, mental change, or abnormal liver enzymes**<br>- Treat as an emergency:<br>- Assess **vitals, mental status, ammonia, liver panel, bilirubin, PT-INR, renal function**.<br>- Perform **abdominal imaging** and **diagnostic paracentesis** if ascites is present to rule out **SBP** and characterize portal-hypertensive physiology.<br>- Rapidly broaden the differential:<br>- **Acute/subacute liver failure**<br>- **Portal venous/hepatic vascular disease** (consistent with later autopsy findings such as obliterative portal venopathy/nodular regenerative hyperplasia)<br>- **Infection/sepsis**<br>- **Drug-related liver injury**<br>- Less likely but important: viral hepatitis or autoimmune liver disease despite negative serology<br>- Do not rely on negative anti-mitochondrial or autoimmune antibodies alone; this case shows that **seronegative immune-mediated hepatobiliary disease can exist**.<br><br>**D. Flexible revision of diagnosis and treatment**<br>- If new liver disease emerges, revise the working diagnosis from isolated renal IgMPC-TIN to **systemic IgM-positive plasma cell–associated disease with hepatic involvement**.<br>- Escalate multidisciplinary care early: **hepatology, nephrology, pathology review**.<br>- Continue organ-supportive therapy while re-evaluating immunosuppression:<br>- If infection is suspected, prioritize **infection control** and reassess steroid intensity. |

| | |
|---|---|
| | - If hepatic encephalopathy develops, start standard therapy promptly and search for triggers such as infection and fluid shifts.<br>- Repeatedly compare organ-specific response:<br>- In this case the **kidney improved**, but the **liver rapidly worsened**; this discordance should trigger immediate reconsideration of the disease model rather than assuming overall treatment success.<br><br>**Bottom line:**<br>Monitor both **renal/tubular response and extra-renal complications**, especially **liver disease and infection**. If the course deviates from expectations, promptly reassess for **systemic disease extension, treatment complications, and alternative diagnoses**, and revise management without anchoring to the initial renal diagnosis alone. |
| With Static RAG and Mult-agent framework (Model C) | 9. **If the condition does not improve as expected or suddenly changes**<br><br>**A. Reassess by separating treatment response from new organ failure**<br><br>- If kidney and tubular findings improve, this supports steroid responsiveness of the **renal** disease, but it does **not** exclude progression in another organ.<br>- In this case, renal function, serum IgM, and mineral disorders improved, yet the later appearance of **ascites, encephalopathy, and rapid liver atrophy** indicated a **new major organ complication**, not simply failure of renal treatment.<br><br>- When a new organ problem appears, reconsider the overall diagnosis as a **systemic IgM-positive plasma-cell disorder with hepatobiliary/portal tract involvement**, rather than isolated kidney disease.<br><br>**B. Immediate action plan for sudden deterioration**<br><br>- **Stabilize first**<br>- For altered consciousness: assess airway, circulation, glucose, ammonia, infection, and metabolic causes.<br>- For new ascites: perform **immediate diagnostic paracentesis** with **PMN count and culture**.<br>- For suspected hepatic encephalopathy: begin standard supportive treatment promptly.<br><br>- **Urgently reassess the differential**<br>- Consider:<br>- progression of the same underlying immune-mediated disease into the liver/portal tracts<br>- **obliterative portal venopathy / nodular regenerative hyperplasia**<br>- drug-related liver injury<br>- infection/sepsis, especially **SBP**<br>- less likely classic PBC, given negative serology and absence of typical histology<br><br>- **Revise investigations based on the new organ failure**<br>- Repeat **CBC, renal panel, liver panel, bilirubin, albumin, PT-INR, ammonia**<br>- Ascitic fluid analysis: **cell count, PMN count, culture, albumin**<br>- Liver imaging to assess **ascites, liver atrophy, and vascular/portal abnormalities**<br>- Obtain **urgent hepatology consultation** and multidisciplinary reassessment<br>- If feasible and safe, consider liver tissue diagnosis when the cause remains unclear |

| | |
|---|---|
|  | **C. Revise treatment flexibly**<br><br>- If steroids help the kidney but the liver worsens:<br> - recognize that organ responses may diverge<br> - reassess whether continuing steroid monotherapy is beneficial, neutral, or harmful in the setting of evolving hepatic failure and infection risk<br><br>- If SBP is suspected:<br> - do **not** wait for confirmation before acting; start **early empiric antibiotics** after diagnostic paracentesis<br><br>- If severe infection develops:<br> - treat infection urgently and reconsider the intensity of immunosuppression<br><br>- If liver failure progresses despite standard measures:<br> - shift focus to complication control: **encephalopathy treatment, ascites management, SBP prevention/treatment, nutrition, and goals-of-care discussion**<br><br>**Changes from the initial version**<br>- I revised **serum IgM** to be a **supportive**, not disease-specific, marker.<br>- I strengthened the liver monitoring plan by adding **serial bilirubin, INR, albumin, ammonia, imaging**, and warning that routine liver enzymes may **underestimate portal venopathy/nodular regenerative hyperplasia**.<br>- I made the acute deterioration plan more operational by explicitly prioritizing **urgent hepatology involvement**, **immediate paracentesis with PMN count/culture**, and **early empiric antibiotics when SBP is suspected**.<br>- I also made the reassessment of **steroid risk-benefit** more explicit once severe hepatic decompensation or infection develops. |

| **Case 10: Pauci-immune crescentic glomerulonephritis caused to dilemma in a patient with suspected systemic lupus erythematosus: a case report** [18] | |
|---|---|
| **Phase I – Q1: Summarizing medical problems** | |
| Baseline LLM (Model A) | 1. **Problem summary**<br><br>A **37-year-old man** with **acute headache and syncope**, found on brain MRI to have **small cerebral infarcts and radiologic signs of cerebral vasculitis**, had **spontaneous neurologic recovery** but was also noted to have **elevated inflammatory markers and moderate proteinuria**. He was referred to nephrology for evaluation of **possible renal involvement in a systemic inflammatory/vasculitic process**.<br>Key features are: **young age, cerebrovascular ischemic events, inflammatory state, and renal abnormality (proteinuria)**, which together strongly suggest a **systemic disease rather than isolated stroke**. |
| With Static RAG (Model B) | 1. **Problem representation**<br><br>A **37-year-old man** with **acute headache and syncope** was found on brain MRI to have **small cerebral infarcts and radiologic signs of cerebral vasculitis**, with **spontaneous neurologic recovery**. He has **no known chronic disease or recent medication exposure**. Laboratory evaluation showed **elevated inflammatory markers** and **moderate proteinuria**, prompting nephrology referral for possible **systemic inflammatory/vasculitic disease with renal |

| | |
|---|---|
| | involvement**.<br><br>Key points:<br>- Young patient with **ischemic CNS events**<br>- Imaging suggests **vasculitic process**<br>- Concurrent **systemic inflammation**<br>- **Proteinuria** suggests possible **glomerular kidney involvement**<br>- Multisystem pattern raises concern for **systemic vasculitis or autoimmune disease** |
| With Static RAG and Mult-agent framework (Model C) | ## 1. Problem representation<br><br>A **37-year-old man** with **acute headache and syncope** was found on brain MRI to have **small cerebral infarcts and radiologic features suggestive of cerebral vasculitis**, with **spontaneous neurologic recovery**. He also had **elevated inflammatory markers** and **moderate proteinuria**, raising concern for **possible renal involvement as part of a systemic process**.<br><br>**Key clinical issue:** in a young patient with cerebral ischemic lesions plus inflammation and proteinuria, the priority is to distinguish **systemic vasculitis** from **thromboembolic/autoimmune disease** and **infection-related embolic disease**. |
| **Phase I – Q2: Differential diagnoses and rationale** | |
| Baseline LLM (Model A) | 2. **Differential diagnoses**<br><br>**Top 3 differential diagnoses**<br>1. **ANCA-associated vasculitis (AAV)**, especially a small-vessel systemic vasculitis with renal involvement<br>- Explains inflammatory markers, proteinuria, and possible CNS ischemic manifestations from vasculitic involvement.<br>2. **Systemic lupus erythematosus / antiphospholipid-associated disease**<br>- Can cause stroke/syncope through thrombosis, and kidney disease through glomerulonephritis; inflammatory markers may be elevated.<br>3. **Primary angiitis of the central nervous system (PACNS) with separate renal disease or secondary systemic vasculitis**<br>- CNS imaging may suggest vasculitis, but isolated PACNS does not usually explain proteinuria well; therefore systemic disease is more likely.<br><br>**Most likely:** **Systemic vasculitis, especially ANCA-associated vasculitis.**<br>Reasoning: the combination of **cerebral ischemic lesions + inflammatory markers + proteinuria** is highly compatible with a systemic small-vessel vasculitis affecting both brain and kidney.<br><br>**Must not be missed:**<br>- **Rapidly progressive glomerulonephritis due to systemic vasculitis**<br>- **Antiphospholipid syndrome / other hypercoagulable state causing recurrent stroke**<br>These are critical because delayed diagnosis risks **irreversible renal failure** or **recurrent CNS ischemia**. |
| With Static RAG | 2. **Differential diagnosis**<br><br>**Top 3 differentials** |

| | |
|---|---|
| (Model B) | 1. **ANCA-associated vasculitis (AAV)** with renal and CNS involvement<br>2. **Systemic lupus erythematosus / antiphospholipid syndrome-related disease**<br>3. **Primary angiitis of the central nervous system (PACNS)** or other systemic vasculitis such as **polyarteritis nodosa**<br><br>**Most likely: ANCA-associated vasculitis**<br>- Explains **elevated inflammatory markers**<br>- Can cause **proteinuria** from pauci-immune glomerulonephritis<br>- May involve the **CNS**, though less common than renal/ENT/pulmonary involvement<br>- Fits a **systemic small-vessel vasculitic** process better than isolated CNS disease<br><br>**Must not be missed: Lupus nephritis/antiphospholipid syndrome or infective endocarditis with embolic stroke**<br>- **APS/SLE** can cause **stroke in a young patient** plus **proteinuria**<br>- **Infective endocarditis** can cause **cerebral emboli, glomerulonephritis, and inflammatory marker elevation**; missing it would be dangerous, especially before immunosuppression<br>- PACNS is important, but kidney findings make a **systemic process** more likely |
| With Static RAG and Mult-agent framework (Model C) | ## 2. Prioritized differential diagnosis<br><br>### 1. **Systemic vasculitis with renal involvement**<br>Includes **ANCA-associated vasculitis** and less likely other systemic vasculitides.<br>- Fits the combination of **inflammation**, **possible cerebral vasculitic changes**, and **proteinuria**.<br>- Renal involvement is possible, but **proteinuria alone does not yet prove glomerulonephritis or true kidney injury**.<br><br>### 2. **Primary angiitis of the central nervous system (PACNS)**<br>- Suggested by MRI findings interpreted as **cerebral vasculitis**.<br>- Important alternative if systemic workup is unrevealing.<br>- However, isolated PACNS would not easily explain proteinuria unless this is unrelated or coincidental.<br><br>### 3. **Antiphospholipid syndrome (APS), with or without SLE**<br>- Young patient with **cerebral infarcts** strongly raises a thrombotic disorder.<br>- **Proteinuria** may occur with lupus nephritis or APS-associated renal vascular disease.<br>- This may fit better than ANCA vasculitis if urine sediment is not nephritic.<br><br>### 4. **Infective endocarditis or other embolic source**<br>- Can cause **multiple small cerebral infarcts**, inflammatory marker elevation, and renal abnormalities.<br>- Must not be missed because treatment differs greatly from vasculitis.<br><br>### 5. **Other systemic vasculitides / vascular disorders**<br>Examples: **polyarteritis nodosa**, cryoglobulinemic disease, hepatitis-associated vasculitis, less likely other inflammatory vascular conditions.<br>- These can involve brain and kidney, though the exact pattern here is not yet specific.<br><br>### 6. **Severe hypertension or other vascular/thrombotic stroke etiologies in the young**<br>- Important because hypertension can cause **headache, syncope, renal protein loss, |

| | |
|---|---|
| | and cerebrovascular events**.<br>- Also consider other embolic or hypercoagulable causes if vasculitis is not confirmed.<br><br>### Most likely at this stage<br>The data support **a possible systemic inflammatory or vascular disorder**, but it is **too early to rank ANCA-associated vasculitis as definitively most likely**. The leading categories are:<br>- **Systemic vasculitis**<br>- **Primary CNS vasculitis**<br>- **APS/SLE or other thrombotic disease**<br>- **Infectious/embolic disease**<br><br>### Must not miss<br>- **Infective endocarditis / embolic source**<br>- **APS**<br>- **Rapidly progressive glomerulonephritis if renal involvement is confirmed** |
| **Phase I – Q3: Necessary physical examinations and reasons** | |
| Baseline LLM (Model A) | 3. **Additional history and physical examination**<br><br>**History**<br>- **Urinary symptoms**: dark urine, foamy urine, oliguria, edema<br> - To assess glomerular involvement.<br>- **Systemic vasculitis symptoms**: fever, weight loss, fatigue, arthralgia, myalgia, rash, purpura, sinus symptoms, epistaxis, cough, hemoptysis, dyspnea, neuropathy<br> - To identify multisystem vasculitis pattern.<br>- **Autoimmune/thrombotic clues**: oral ulcers, photosensitivity, Raynaud phenomenon, prior thrombosis, miscarriages/family history of thrombosis<br> - To evaluate SLE/APS.<br>- **Infectious risk factors**: recent infection, endocarditis symptoms, hepatitis risk<br> - Important because infection can mimic vasculitis and some secondary vasculitides are infection-related.<br>- **Drug/substance exposure**: cocaine, amphetamines, supplements, recent new drugs<br> - Can trigger vasculitis or stroke syndromes.<br><br>**Physical examination**<br>- **Blood pressure and volume status**<br> - Hypertension or edema would support renal involvement.<br>- **Skin exam** for purpura, livedo, ulcers<br> - Suggests systemic vasculitis or APS.<br>- **ENT and lung exam**<br> - Upper/lower respiratory involvement supports AAV.<br>- **Cardiac exam**<br> - Murmur may suggest cardioembolic source or infective endocarditis.<br>- **Neurologic exam**<br> - To document any persistent focal deficit.<br>- **Funduscopic exam**<br> - Can show vasculitic or hypertensive changes. |
| With Static RAG (Model B) | 3. **Additional history and physical examination**<br><br>**History**<br>- **Constitutional symptoms**: fever, weight loss, night sweats |

| | |
|---|---|
| | - To assess systemic inflammatory or infectious disease<br>- **ENT/pulmonary symptoms**: sinusitis, epistaxis, cough, hemoptysis, dyspnea<br>- Support AAV, especially GPA<br>- **Skin/joint symptoms**: purpura, ulcers, arthralgia<br>- Look for systemic vasculitis or lupus<br>- **Urinary symptoms**: foamy urine, hematuria, edema, oliguria<br>- Clarify glomerular renal involvement<br>- **Thrombotic/autoimmune history**: oral ulcers, photosensitivity, rash, miscarriages/family thrombosis<br>- Suggest SLE/APS<br>- **Infectious clues**: dental work, IV drug use, recent infection, fevers, heart murmur history<br>- Evaluate endocarditis<br>- **Substance exposure**: cocaine/amphetamines, supplements, toxins<br>- Can mimic vasculitis or cause stroke<br><br>**Physical examination**<br>- **Blood pressure, volume status, edema**<br>- Assess kidney disease severity<br>- **Skin exam** for purpura, livedo, ulcers, splinter hemorrhages<br>- Distinguish vasculitis, APS, endocarditis<br>- **Cardiac exam** for murmur<br>- Screen for endocarditis/cardioembolism<br>- **ENT exam**<br>- Evidence of granulomatous disease/AAV<br>- **Chest exam**<br>- Pulmonary hemorrhage or other systemic involvement<br>- **Focused neurologic exam**<br>- Confirm residual deficits or multifocal disease pattern |
| With Static RAG and Mult-agent framework (Model C) | ## 3. Focused history and physical examination<br><br>## Targeted history<br><br>### Renal questions<br>- **Foamy urine, dark/red urine, reduced urine output, edema**<br>- To look for glomerular disease or nephritic/nephrotic features.<br>- **Timing of proteinuria and any prior urinalyses**<br>- To determine whether this is new and clinically significant.<br><br>### Blood pressure / hypertensive symptoms<br>- **History of hypertension, severe headache, visual symptoms**<br>- Hypertension could connect neurologic and renal findings.<br><br>### Vasculitis/systemic inflammatory symptoms<br>- **Fever, weight loss, fatigue, arthralgia, myalgia**<br>- Support systemic inflammatory disease.<br>- **Purpura, rash, ulcers, livedo**<br>- Helpful for vasculitis or thrombotic disease.<br>- **Sinus symptoms, epistaxis, cough, dyspnea, hemoptysis**<br>- Suggest ANCA-associated vasculitis.<br><br>### Thrombotic/autoimmune history<br>- **Prior DVT/PE, family history of thrombosis** |

| | |
|---|---|
| | - Supports APS or inherited thrombophilia.<br>- **Oral ulcers, photosensitivity, Raynaud symptoms**<br>- Suggest SLE/connective tissue disease.<br><br>### Infectious/embolic history<br>- **Fever, chills, recent dental procedure, IV drug use, known murmur**<br>- Important for endocarditis or systemic infection.<br><br>### Exposure history<br>- **Cocaine/amphetamine use, vasculitis-inducing drugs**<br>- May explain stroke or vasculopathy.<br><br>---<br><br>## Focused physical examination<br><br>### Vital signs, especially **blood pressure**<br>- Crucial to assess for severe hypertension.<br><br>### General and volume assessment<br>- **Edema**<br>- Supports renal involvement.<br><br>### Skin examination<br>- **Purpura, livedo reticularis, splinter hemorrhages, ulcers**<br>- Helps distinguish vasculitis, APS, or endocarditis.<br><br>### Cardiovascular examination<br>- **Murmur, peripheral embolic signs**<br>- Screens for cardiac embolic source or infective endocarditis.<br><br>### Neurologic examination<br>- Confirm absence of persistent focal deficits.<br><br>### Funduscopic examination<br>- Look for **hypertensive retinopathy or vascular changes**<br>- May support severe hypertension/systemic vascular disease.<br><br>### ENT and chest examination<br>- Looks for systemic vasculitis clues, especially AAV. |
| **Phase I – Q4: Plan for investigations/tests** | |
| Baseline LLM (Model A) | 4. **Tests to narrow the diagnosis**<br><br>**Urine and kidney assessment**<br>- **Urinalysis with microscopy**<br>- Purpose: detect dysmorphic RBCs, RBC casts, degree of proteinuria; expected if glomerulonephritis: **hematuria with dysmorphic RBCs/casts and proteinuria**.<br>- **Urine protein quantification** (spot protein/creatinine or albumin/creatinine ratio)<br>- To define severity of renal involvement.<br>- **Serum creatinine, BUN, electrolytes, albumin**<br>- To assess kidney function and systemic impact.<br><br>**Inflammatory/autoimmune workup** |

| | |
|---|---|
|  | - **CBC, CRP, ESR**<br> - To confirm systemic inflammation; may show anemia of inflammation.<br>- **ANCA (PR3-ANCA, MPO-ANCA)**<br> - To evaluate AAV; expected possibly positive in systemic small-vessel vasculitis.<br>- **ANA, anti-dsDNA, complements (C3/C4)**<br> - To assess SLE/immune-complex disease; lupus would often show **positive ANA/dsDNA with low complement**.<br>- **Antiphospholipid antibodies**: lupus anticoagulant, anticardiolipin, anti-β2 glycoprotein I<br> - To evaluate APS as a cause of cerebral infarcts.<br>- **IgA level, cryoglobulins, hepatitis B/C serologies**<br> - To assess other secondary/systemic vasculitides.<br><br>**Exclude mimics**<br>- **Blood cultures if infection suspected**<br> - Endocarditis can cause stroke, glomerulonephritis, and elevated inflammatory markers.<br>- **Echocardiography**<br> - To look for cardiac embolic source or endocarditis.<br>- **Coagulation profile**<br> - To evaluate thrombophilia context.<br><br>**Kidney-specific definitive test**<br>- **Kidney biopsy**<br> - Most important to define the renal lesion and guide therapy.<br> - Expected result if AAV: **pauci-immune necrotizing/crescentic glomerulonephritis**.<br> - If lupus/immune-complex disease: **immune deposits on immunofluorescence**.<br><br>Overall, the workup should focus first on confirming **systemic vasculitis with glomerulonephritis**, while urgently excluding **APS, infection, and other stroke mimics**. |
| With Static RAG (Model B) | 4. **Tests to narrow the diagnosis**<br><br>**Urine and kidney assessment**<br>- **Urinalysis with microscopy**<br> - Purpose: detect glomerular disease<br> - Expected: **hematuria, dysmorphic RBCs, RBC casts, proteinuria** if nephritic process<br>- **Urine protein quantification** (spot protein/creatinine or albumin/creatinine ratio)<br> - Purpose: quantify renal involvement<br>- **Serum creatinine, BUN, electrolytes, albumin**<br> - Purpose: assess kidney function and severity<br><br>**Autoimmune/vasculitis workup**<br>- **ANCA (PR3-ANCA, MPO-ANCA)**<br> - Purpose: evaluate AAV<br> - Expected: positive in AAV, though not definitive alone<br>- **ANA, anti-dsDNA, C3/C4**<br> - Purpose: evaluate SLE/immune-complex disease<br> - Expected: lupus would often show **positive ANA/dsDNA and low complement**<br>- **Antiphospholipid antibodies**: lupus anticoagulant, anticardiolipin, anti-β2 glycoprotein I |

| | |
|---|---|
| | - Purpose: assess APS as cause of stroke in a young patient<br>- **ESR/CRP**<br>- Purpose: confirm inflammatory burden and follow trend<br><br>**Infectious evaluation**<br>- **Blood cultures**<br>- Purpose: exclude infective endocarditis before immunosuppression<br>- **Echocardiography**<br>- Purpose: look for vegetations/cardioembolic source<br>- Consider **hepatitis B/C and HIV testing**<br>- Purpose: secondary vasculitis or glomerular disease causes<br><br>**Neurologic/vascular evaluation**<br>- **Brain/vascular imaging review or angiography as indicated**<br>- Purpose: characterize vasculitic vs embolic pattern<br>- **Lumbar puncture** if PACNS remains a concern and no contraindication<br>- Purpose: supportive evidence for CNS vasculitis; helps exclude infection<br><br>**Definitive renal test**<br>- **Kidney biopsy**<br>- Purpose: establish the exact renal lesion and guide therapy<br>- Expected:<br>- **AAV**: pauci-immune necrotizing/crescentic glomerulonephritis<br>- **Lupus**: immune-complex glomerulonephritis<br>- **Endocarditis-related GN**: immune-complex pattern<br><br>**Overall diagnostic strategy**<br>Because he has **CNS ischemic lesions + inflammation + proteinuria**, the priority is to distinguish **systemic vasculitis/autoimmune glomerulonephritis** from **infection-associated disease**. The most informative next steps are **urine sediment, kidney function, serologic vasculitis/autoimmune tests, blood cultures/echo, and kidney biopsy**. |
| With Static RAG and Mult-agent framework (Model C) | ## 4. Logical testing plan<br><br>## First: confirm whether there is true renal involvement<br><br>### 1. **Serum creatinine, BUN, electrolytes, eGFR, albumin**<br>- Establish whether kidney function is actually impaired.<br><br>### 2. **Repeat urinalysis with microscopy**<br>- Look for **hematuria, dysmorphic RBCs, RBC casts**<br>- This is critical to determine whether proteinuria is glomerular/nephritic.<br><br>### 3. **Quantify proteinuria**<br>- Spot **urine protein-creatinine ratio** or **albumin-creatinine ratio**<br>- Determines severity and significance.<br><br>### 4. **Blood pressure measurement**<br>- Essential bedside discriminator before advancing to invasive testing.<br><br>---<br><br>## Second: evaluate major etiologic categories |

#### If inflammatory/systemic disease remains likely:
- **CBC, ESR, CRP**
- **ANCA (MPO/PR3)**
- **ANA, anti-dsDNA, C3, C4**
- **Antiphospholipid antibodies**: lupus anticoagulant, anticardiolipin, anti-β2 glycoprotein I
- **Hepatitis B/C serologies**, possibly **cryoglobulins**

These tests help separate:
- pauci-immune vasculitis,
- immune-complex disease,
- lupus/APS,
- infection-associated or hepatitis-associated vasculitis.

#### If embolic/infectious source is a concern:
- **Blood cultures**
- **Echocardiography**

These are important before considering immunosuppression.

---

### Third: targeted adjunctive studies

#### 5. **Renal ultrasound**
- Low-cost, noninvasive assessment for kidney size and structural disease.

#### 6. **Additional imaging based on symptoms**
- **Chest X-ray/CT** if pulmonary symptoms are present
  - Pulmonary-renal syndrome would support AAV.

---

### Fourth: definitive testing if renal findings justify it

#### 7. **Kidney biopsy**
Consider if there is:
- impaired kidney function,
- active urine sediment,
- significant persistent proteinuria,
- or strong suspicion of intrinsic renal disease.

Purpose:
- Distinguish **glomerular**, **vascular**, or **thrombotic** renal pathology.
- Guide therapy and avoid inappropriate empiric treatment.

---

### Changes from the initial version

1. **Problem representation revised**
  - I changed **“kidney injury”** to **“possible renal involvement”**, because moderate proteinuria alone does not confirm intrinsic renal dysfunction.

| | |
|---|---|
| | 2. **Differential diagnosis re-prioritized**<br>- I removed the premature conclusion that **ANCA-associated vasculitis is the most likely diagnosis**.<br>- I now explicitly separate **systemic vasculitis** from **primary CNS vasculitis**.<br>- I gave more weight to **APS/embolism/infective endocarditis** as competing explanations for cerebral infarcts in a young patient.<br><br>3. **History and exam refined**<br>- I emphasized **blood pressure**, **hematuria/urine color**, **funduscopic exam**, **skin findings**, and **focused cardiovascular evaluation**.<br><br>4. **Testing sequence clarified**<br>- I now prioritize first confirming renal involvement with:<br>- **creatinine/eGFR**<br>- **repeat urinalysis with microscopy**<br>- **protein quantification**<br>- **blood pressure**<br>- Serologies, echocardiography, and biopsy are then used selectively based on whether the pattern looks **glomerular, vascular, thrombotic, or infectious**. |
| **Phase II – Q5: Interpreting test results** | |
| Baseline LLM (Model A) | **5. Interpretation of the test results**<br><br>- **Renal function is still preserved**: creatinine 0.98 mg/dL and eGFR 98 mL/min/1.73 m² indicate that, despite active urinary abnormalities, there is not yet a major fall in filtration. This is important because it suggests **early but active glomerular disease**, a potentially reversible stage if treated promptly.<br>- **Urine findings strongly indicate glomerulonephritis**: microscopic hematuria, **dysmorphic RBCs**, and **RBC casts** are classic for **glomerular bleeding** rather than urologic bleeding. The **1.8 g/day proteinuria** is subnephrotic and fits an **inflammatory glomerular process** rather than primary nephrotic syndrome.<br>- **CRP 84 mg/L with mild leukocytosis and thrombocytosis** supports a significant inflammatory process. In context, this is compatible with **systemic vasculitic/inflammatory disease**.<br>- **Normal albumin and absence of edema** are consistent with the proteinuria being subnephrotic and not causing nephrotic syndrome.<br>- **Normal complement (C3/C4)** argues **against classic immune-complex lupus nephritis**, cryoglobulinemic GN, or infection-related GN, although it does not absolutely exclude autoimmune disease.<br>- **ANA high-titer, anti-dsDNA positive, anti-ribosomal P positive** suggest **lupus autoimmunity/serologic lupus phenotype**. However, these serologies must be interpreted together with the biopsy and clinical picture.<br>- **Negative antiphospholipid antibodies** make APS less likely as the explanation for the cerebral infarcts.<br>- **Negative ANCA (IFA, PR3, MPO)** does not exclude pauci-immune GN; a subset of patients have **ANCA-negative pauci-immune crescentic GN**.<br>- **Negative anti-GBM** helps exclude anti-GBM disease.<br>- **Kidney biopsy is the key result**: **pauci-immune crescentic glomerulonephritis** with **cellular crescents in 8/19 glomeruli** indicates **active necrotizing small-vessel glomerular injury**. The absence of Ig/complement staining argues strongly **against immune-complex lupus nephritis**, despite positive ANA/dsDNA. This biopsy pattern is most consistent with **pauci-immune GN**, not |

| | |
|---|---|
| | membranous disease, not amyloidosis, and not thrombotic microangiopathy.<br><br>**Bottom line:** the patient has **active crescentic glomerulonephritis with preserved kidney function**, and the biopsy pattern outweighs the lupus-like serologies in defining the renal lesion. |
| With Static RAG (Model B) | **5. Interpretation of the test results**<br><br>- **Renal function is currently preserved**: creatinine 0.98 mg/dL and eGFR 98 mL/min/1.73 $m^2$ indicate that despite active glomerular injury, overall filtration is still normal. This is important because it suggests **early or limited renal involvement**, not advanced chronic kidney damage.<br>- **Urine findings strongly indicate active glomerulonephritis**: microscopic hematuria, **dysmorphic RBCs**, and **RBC casts** are classic for a **glomerular inflammatory process** rather than lower urinary tract bleeding. Proteinuria of **1.8 g/day** is subnephrotic and fits a **nephritic pattern**.<br>- **CRP 84 mg/L** and mild leukocytosis support a significant **active inflammatory process**.<br>- **Normal kidney ultrasound** supports a **non-chronic, potentially reversible** renal disease.<br>- **Serology is mixed but important**:<br>- **ANA high titer**, **anti-dsDNA positive**, and **anti-ribosomal P positive** raise concern for **SLE**.<br>- However, **normal C3/C4**, absence of clear extrarenal SLE manifestations, and most importantly the **kidney biopsy pattern** argue **against classic lupus nephritis**.<br>- **ANCA negative** does **not exclude pauci-immune crescentic GN**, because **ANCA-negative pauci-immune GN** is a recognized entity.<br>- **Anti-GBM negative** makes anti-GBM disease unlikely.<br>- Negative antiphospholipid antibodies make **APS less likely** as the explanation for the cerebral infarcts.<br>- **Kidney biopsy is the key result**:<br>- **Cellular crescents in 8/19 glomeruli** indicate **active severe crescentic GN**.<br>- **Pauci-immune immunofluorescence** (no Ig/complement deposition) strongly favors **pauci-immune crescentic glomerulonephritis**, not immune-complex disease such as lupus nephritis or IgA nephropathy.<br>- Lack of thrombi argues against thrombotic microangiopathy/APS-related renal injury.<br>- Mild tubular atrophy/interstitial inflammation suggests some associated tubulointerstitial injury, but not advanced chronic scarring.<br><br>**Overall meaning:** the data support an **active pauci-immune necrotizing/crescentic glomerular disease with preserved kidney function**, occurring in a patient who also had CNS ischemic/vasculitic findings. The positive lupus serologies are notable, but the renal pathology is **not typical for lupus nephritis**. |
| With Static RAG and Mult-agent framework (Model C) | **5. Interpretation of the test results**<br><br>- **There is definite active glomerulonephritis despite preserved serum kidney function.**<br>Creatinine is **0.98 mg/dL** and eGFR **98 mL/min/1.73 $m^2$**, so filtration is still preserved. However, this does **not** mean the kidney disease is mild, especially in a **very large man (192 cm, 105 kg)**, because substantial active glomerular injury can exist before creatinine rises. The key point is that the patient has **histologically |

<table>
<tr><td></td><td>active nephritis with normal current filtration**.<br><br>- **Urine findings clearly indicate a nephritic glomerular process.**<br>- **Microscopic hematuria (50 RBC/HPF)** with **dysmorphic erythrocytes** and **erythrocyte casts** strongly localizes bleeding to the **glomeruli**.<br>- **Proteinuria 1.8 g/day** is **subnephrotic**, fitting **nephritic glomerular inflammation** rather than a pure nephrotic syndrome.<br>- **Pyuria** may reflect renal inflammation in this setting rather than infection, although infection always needs clinical exclusion.<br><br>- **The inflammatory burden is high.**<br>**CRP 84 mg/L** with mild leukocytosis supports an active inflammatory process, consistent with ongoing renal inflammation and possibly a broader systemic process.<br><br>- **The absence of edema and the normal albumin are clinically coherent.**<br>Albumin is **4.5 g/dL** and there are no signs of hypervolemia or edema, which supports that this is **not nephrotic syndrome** and matches the **subnephrotic proteinuria**.<br><br>- **Serology is autoimmune-positive but not typical for immune-complex lupus nephritis.**<br>- **ANA 1:1000**, **anti-dsDNA positive**, and **anti-ribosomal P positive** are notable and keep **systemic autoimmunity** in the discussion.<br>- However, **normal C3/C4**, absence of clear extrarenal SLE features, and most importantly the **renal biopsy** argue strongly **against classic lupus nephritis**.<br>- **Negative antiphospholipid antibodies** make **APS** less likely as a unifying explanation for the cerebral infarcts.<br>- **Negative ANCA (IFA, PR3, MPO)** does **not** exclude pauci-immune GN, because **ANCA-negative pauci-immune crescentic GN** is well recognized.<br>- **Negative anti-GBM** helps exclude anti-GBM disease.<br><br>- **The kidney biopsy is the decisive result.**<br>- The biopsy shows **pauci-immune crescentic glomerulonephritis** with **cellular crescents in 8 of 19 glomeruli**, indicating **active aggressive glomerular injury**.<br>- The lack of staining for **IgA, IgG, IgM, C3, C4, and C1q** argues strongly against **immune-complex GN**, including typical **lupus nephritis**.<br>- The absence of **glomerular thrombi** argues against **APS-associated nephropathy / thrombotic microangiopathy**.<br>- The absence of arterial wall vasculitis on the biopsy does **not** exclude a pauci-immune small-vessel process centered in the glomeruli.<br><br>**Established conclusion:**<br>The patient has **biopsy-proven ANCA-negative pauci-immune crescentic glomerulonephritis with preserved renal function**.<br><br>**Inferred but not proven:**<br>Given the prior cerebral infarcts and MRI suggestion of vasculitis, a **systemic vasculitic process with CNS involvement is suspected**, but the renal biopsy alone does **not prove** that the brain lesions arose from the same disease.</td></tr>
<tr><td colspan="2">Phase II – Q6: Reconsidering the differential diagnosis</td></tr>
<tr><td>Baseline LLM (Model A)</td><td>**6. Updated diagnostic thinking / Working diagnosis**<br><br>### Revised differential diagnosis</td></tr>
</table>

| | |
|---|---|
| | 1. **ANCA-negative pauci-immune crescentic glomerulonephritis** with probable systemic small-vessel vasculitic involvement<br>2. **Serology-positive lupus with pauci-immune crescentic GN ("lupus-like" autoimmune background, but not immune-complex lupus nephritis)**<br>3. Much less likely: other secondary pauci-immune GN or atypical vasculitis<br><br>### Why the diagnosis changed<br>Initially, SLE/APS and ANCA-associated vasculitis were both plausible because of stroke-like CNS disease, inflammation, and proteinuria. After biopsy, the differential narrows substantially:<br><br>- **Biopsy shows pauci-immune crescentic GN**, which is the defining lesion of **pauci-immune vasculitic GN**.<br>- **No immune deposits** on IF argues strongly against **lupus nephritis**, which typically shows immune-complex deposition.<br>- **ANCA negativity** does not rule out this diagnosis.<br>- **APS is unlikely** because antiphospholipid testing is negative and the kidney biopsy does not show thrombotic lesions.<br>- Positive ANA/dsDNA suggest an autoimmune background, but the **renal pathology is not typical for lupus nephritis**.<br><br>### Working diagnosis<br>**ANCA-negative pauci-immune crescentic glomerulonephritis, likely as part of a systemic small-vessel vasculitic syndrome with CNS involvement.** |
| With Static RAG (Model B) | **6. Updated diagnostic thinking / Working diagnosis**<br><br>My diagnostic thinking changes substantially after the **kidney biopsy**, because pathology outweighs serologic ambiguity.<br><br>- Before biopsy, major considerations included:<br> - **SLE/lupus nephritis**<br> - **ANCA-associated vasculitis**<br> - Other systemic vasculitides<br>- After biopsy, the leading diagnosis becomes:<br> - **ANCA-negative pauci-immune crescentic glomerulonephritis**, likely within the spectrum of **pauci-immune small-vessel vasculitis**, with **possible CNS vasculitic involvement**.<br><br>**Why this is now most likely**<br>- The urine sediment proves **true glomerulonephritis**.<br>- The biopsy shows **pauci-immune crescentic GN**, which is the defining renal lesion of **pauci-immune vasculitis**.<br>- **ANCA negativity does not rule this out**.<br>- Although ANA and anti-dsDNA are positive, the biopsy lacks the **immune-complex deposition** expected in lupus nephritis ("full house" pattern is absent), complements are normal, and there are no clear clinical SLE features.<br>- APS is less likely because antiphospholipid testing is negative and renal biopsy shows **crescentic GN rather than thrombotic lesions**.<br><br>**Working diagnosis:**<br>**ANCA-negative pauci-immune crescentic glomerulonephritis, likely part of a systemic pauci-immune small-vessel vasculitis with CNS involvement.**<br>A secondary note is that he has **lupus-like autoantibody positivity**, but current |

| | |
|---|---|
| | evidence does **not support classic lupus nephritis**. |
| With Static RAG and Mult-agent framework (Model C) | **6. Updated diagnostic thinking / Working diagnosis**<br><br>The biopsy substantially refines the differential diagnosis.<br><br>- **Most likely diagnosis:**<br>**ANCA-negative pauci-immune crescentic glomerulonephritis**<br><br>- **How the differential changes:**<br>- **Lupus nephritis becomes much less likely** because classic lupus nephritis is usually an **immune-complex GN** with immunofluorescence positivity; that pattern is absent here.<br>- However, lupus should not be dismissed absolutely, because the patient does have unusual autoimmune serologies (**ANA/dsDNA/ribosomal P positivity**). These may represent **concurrent autoimmune seropositivity** rather than the cause of the renal lesion.<br>- **APS is less likely** because antiphospholipid antibodies are negative and there are **no thrombotic renal lesions** on biopsy.<br>- **Infective endocarditis or other embolic disease** may still be relevant to the prior cerebral infarcts, but they do **not** explain the renal biopsy pattern well.<br>- The pathology best fits the spectrum of **pauci-immune small-vessel vasculitis**, which may be **renal-limited** or **systemic**.<br><br>**Working diagnosis:**<br>**ANCA-negative pauci-immune crescentic glomerulonephritis, likely renal-limited or part of a systemic small-vessel vasculitic process. CNS association is suspected but not definitively established from the provided data.**<br><br>A cautious unifying interpretation is:<br>- **Established:** pauci-immune crescentic GN<br>- **Suspected:** systemic vasculitis linking kidney and CNS findings<br>- **Still worth excluding clinically before final unification:** infection and other causes of cerebral infarction in a young patient<br><br>**Changes from the initial impression:**<br>- I would now place **biopsy-proven ANCA-negative pauci-immune crescentic GN** clearly at the top.<br>- I would be **less definitive** than before in stating that the patient already has proven **systemic vasculitis with CNS involvement**; that remains **probable but not confirmed**.<br>- I would also keep a brief note that **autoimmune serology is discordant** and does not override the biopsy. |
| **Phase II – Q7: Treatment planning** | |
| Baseline LLM (Model A) | **7. Treatment plan**<br><br>### Main treatment<br>Because this is **active crescentic GN** with probable extrarenal vasculitic involvement (CNS), treatment should follow an **induction regimen for severe pauci-immune vasculitis**, even though kidney function is currently preserved.<br><br>1. **High-dose glucocorticoids**<br>- Example approach: **IV methylprednisolone pulses** initially, followed by **oral |

| | |
|---|---|
|  | prednisone/prednisolone** with taper.<br> - **Rationale:** rapidly suppresses glomerular and systemic vascular inflammation and may prevent irreversible renal/cerebral injury.<br><br>2. **Add an induction immunosuppressive agent**<br> - **Rituximab** or **cyclophosphamide**<br> - **Rationale:** crescentic pauci-immune GN generally requires more than steroids alone.<br> - In this patient, **rituximab** is an attractive option because he is young and cyclophosphamide has cumulative toxicity; however, **cyclophosphamide** is also an accepted standard, especially in severe organ-threatening disease.<br><br>### Supportive management<br>- **Close monitoring** of creatinine, urinalysis, proteinuria, CRP, CBC, liver tests<br>- **Blood pressure and renoprotective management**; if proteinuria persists and blood pressure allows, consider **RAS blockade**<br>- **Glucocorticoid toxicity prevention**: gastric protection as appropriate, calcium/vitamin D, consider bone protection depending on dose/duration<br>- **Pneumocystis prophylaxis** if high-dose steroids are combined with rituximab/cyclophosphamide<br>- Reassess the need for **ongoing anticoagulation**, since APS is not supported and the main process appears inflammatory/vasculitic rather than thrombotic<br><br>### Other options / comments<br>- **Plasma exchange** is generally reserved for selected situations such as severe pulmonary hemorrhage or severe anti-GBM disease; based on the provided data, it is **not routinely indicated here**.<br>- If future findings support definite SLE rather than isolated pauci-immune vasculitis, treatment could be modified, but **current biopsy-based evidence favors vasculitis-directed induction therapy**.<br>- After remission, transition to **maintenance therapy** (commonly rituximab or azathioprine, depending on induction choice and response).<br><br>**Overall treatment goal:** induce rapid remission of **organ-threatening pauci-immune vasculitic GN**, preserve renal function, and reduce risk of further CNS events. |
| With Static RAG (Model B) | **7. Treatment plan**<br><br>**Primary treatment**<br>- Treat as **active pauci-immune crescentic GN / systemic vasculitis** with **induction immunosuppression**:<br> 1. **High-dose glucocorticoids**<br> - Typically pulse **IV methylprednisolone** in severe organ-threatening disease, followed by **oral prednisone**<br> - Rationale: rapid control of glomerular and possible CNS vasculitic inflammation<br> 2. **Rituximab or cyclophosphamide** for remission induction<br> - Either is evidence-based for severe pauci-immune GN<br> - Given **crescentic biopsy** and **extrarenal CNS involvement**, this is **organ-threatening disease**, so steroid alone is insufficient<br><br>**Which induction agent?**<br>- **Rituximab**: effective alternative to cyclophosphamide, often preferred when fertility preservation or cumulative toxicity is a concern |

| | |
|---|---|
|  | - **Cyclophosphamide**: also appropriate, especially in severe rapidly progressive disease or when broader historical experience is preferred<br><br>**Supportive management**<br>- Close monitoring of:<br> - creatinine/eGFR<br> - urine protein and sediment<br> - blood pressure<br> - CRP and blood counts<br>- Add standard steroid/immunosuppression safety measures as appropriate:<br> - infection prophylaxis (commonly **PJP prophylaxis** when using high-dose steroids plus another immunosuppressant)<br> - gastric and bone protection depending on regimen/duration<br>- Continue to reassess the need for anticoagulation based on neurologic and vascular evaluation, but the current core disease process appears **inflammatory vasculitic**, not APS-mediated thrombosis.<br><br>**Why not treat as lupus nephritis first?**<br>- Because the **biopsy pattern is pauci-immune**, not immune-complex lupus nephritis. Treatment overlap exists, but diagnostic classification and long-term management are better aligned with **pauci-immune vasculitis**.<br><br>**Other options / considerations**<br>- **Plasma exchange** is generally reserved for selected situations such as severe pulmonary hemorrhage or advanced rapidly progressive kidney failure; based on the provided data, there is **no clear indication** here.<br>- If later features of definite SLE emerge, treatment may need reassessment, but with current information, the biopsy-driven diagnosis should guide therapy.<br><br>**Practical plan:**<br>Initiate **high-dose corticosteroids plus rituximab or cyclophosphamide** for induction of remission, with supportive prophylaxis and close renal/neurologic follow-up. |
| With Static RAG and Mult-agent framework (Model C) | **7. Treatment plan**<br><br>**Recommended approach: start treatment promptly once active infection has been reasonably excluded.**<br><br>### A. Induction therapy<br>Because this is **active crescentic GN**, treatment should not be delayed simply because creatinine is still normal.<br><br>1. **High-dose glucocorticoids**<br> - In organ-threatening disease, many clinicians would use **IV methylprednisolone initially**, followed by **oral prednisone/prednisolone** with taper.<br> - Rationale: rapid suppression of active glomerular inflammation.<br><br>2. **Add a remission-induction agent: cyclophosphamide or rituximab**<br> - **Cyclophosphamide** is a strong choice here, especially if the prior CNS lesions are felt to represent **true vasculitic organ-threatening disease**.<br> - **Rituximab** is also evidence-based for pauci-immune GN and may be preferred in some patients depending on toxicity profile, fertility concerns, and clinician experience. |

#### B. Which induction agent may be favored here?
- If the case is treated as **renal-limited pauci-immune GN**, either **rituximab** or **cyclophosphamide** is reasonable.
- If **CNS vasculitis is strongly suspected**, many clinicians would favor **cyclophosphamide** because of the severity and multiorgan concern.
- The decision should consider:
  - age,
  - fertility wishes,
  - cumulative toxicity concerns,
  - local expertise.

#### C. Safety and supportive measures
These should be stated explicitly:

- **Exclude active infection as far as reasonably possible before full immunosuppression**, especially because cerebral infarcts can also occur with infectious embolic disease.
- **Pneumocystis jirovecii prophylaxis** during intensive immunosuppression, according to local practice.
- **Gastric and bone protection** as appropriate with prolonged steroids.
- **Fertility counseling** before cyclophosphamide if relevant.
- Close monitoring of:
  - **serum creatinine / eGFR**
  - **urine sediment**
  - **proteinuria**
  - **CBC**
  - **liver tests**
  - **CRP**
  - **blood pressure**
  - **drug toxicities**
  - **neurologic status**

- **RAS blockade** can be considered later for persistent proteinuria if blood pressure permits, but this is supportive, not primary therapy.

#### D. Plasma exchange
- **Not routinely indicated** from the provided information.
- There is **no severe renal failure**, **no anti-GBM disease**, and **no diffuse alveolar hemorrhage** described.

#### E. Maintenance therapy after remission
After induction, maintenance usually involves:
- **rituximab** or
- **azathioprine**,
with a continued **glucocorticoid taper**, depending on response and final disease classification.

#### Rationale
- The biopsy shows **active pauci-immune crescentic GN**, which can progress rapidly even when kidney function initially appears normal.
- The patient's preserved creatinine should not provide false reassurance.
- Positive ANA/dsDNA should **not redirect therapy toward lupus nephritis**, because the biopsy does not support immune-complex disease.

<table>
<tr><td></td><td>### Alternative considerations<br>- If future evaluation shows the disease is truly **renal-limited**, the treatment framework is still largely similar.<br>- If a competing cause for the CNS lesions is proven, renal treatment would still be directed by the **biopsy-confirmed pauci-immune crescentic GN**.<br><br>**Changes from the initial plan:**<br>- I would now state more explicitly: **begin induction promptly once infection is reasonably excluded**.<br>- I would add concrete safety steps: **Pneumocystis prophylaxis, fertility counseling if cyclophosphamide is used, and structured monitoring for toxicity and response**.<br>- I would also phrase the CNS component more cautiously: **cyclophosphamide is especially attractive if CNS vasculitis is strongly suspected, but that link is not definitively proven by the kidney biopsy alone**.</td></tr>
<tr><td colspan="2">Phase III – Q8: Evaluation of treatment</td></tr>
<tr><td>Baseline LLM (Model A)</td><td>8. **Monitoring effectiveness, safety, risks, and countermeasures**<br><br>**Effectiveness monitoring**<br>- **Kidney response:** follow **serum creatinine/eGFR**, **urinalysis with sediment**, and **quantitative proteinuria** regularly.<br>- Main targets: stable kidney function, disappearance of active sediment (especially hematuria/RBC casts), and progressive fall in proteinuria.<br>- **Systemic lupus activity:** monitor **anti-dsDNA titers**, **complements (C3/C4)**, **CRP/ESR**, and assess for any new neurologic, mucocutaneous, serosal, or joint manifestations.<br>- **Blood pressure:** monitor regularly, because BP control supports renal protection and reduction of proteinuria.<br>- **Clinical status:** confirm absence of relapse symptoms such as headache, focal neurologic deficits, edema, gross hematuria, or constitutional symptoms.<br><br>**Safety monitoring**<br>- **Glucocorticoids**<br>- Risks: infection, hyperglycemia, hypertension, weight gain, mood/psychiatric effects, myopathy, gastric injury, osteoporosis.<br>- Monitoring/countermeasures: check **blood pressure**, **glucose**, weight, and infection symptoms; taper steroids as disease allows; provide **bone protection** (calcium/vitamin D ± osteoporosis prophylaxis depending on duration/risk); consider gastric protection when appropriate.<br>- **Cyclophosphamide**<br>- Risks: **myelosuppression**, serious infection, hemorrhagic cystitis, infertility/gonadal toxicity, hepatotoxicity, and long-term malignancy risk.<br>- Monitoring/countermeasures: perform **CBC with differential** before each cycle and after treatment as needed; monitor **urinalysis** for hematuria and ensure **good hydration**; adjust or delay dosing if cytopenia or infection occurs; monitor liver tests; counsel regarding fertility risk before/during treatment if relevant.<br>- **Hydroxychloroquine**<br>- Risks: retinal toxicity, less commonly cytopenia or myopathy.<br>- Monitoring/countermeasures: arrange **baseline and periodic ophthalmologic examination** and review for visual symptoms.<br>- **Losartan**<br>- Risks: hyperkalemia and creatinine rise.<br>- Monitoring/countermeasures: check **serum creatinine and potassium** after</td></tr>
</table>

| | |
|---|---|
|  | initiation/dose adjustment and periodically thereafter.<br><br>**Specific risks to watch closely in this case**<br>- **Relapsing nephritis** despite preserved kidney function<br> - Countermeasure: frequent urine/proteinuria and renal function follow-up; escalate/reassess treatment if active sediment or proteinuria returns.<br>- **New CNS lupus/vascular events**<br> - Countermeasure: urgent reassessment of any headache, syncope, focal deficit, seizure, or mental status change; involve neurology promptly and repeat neuroimaging if indicated.<br>- **Treatment-related infection**<br> - Countermeasure: low threshold for evaluation of fever or localizing symptoms; hold or adjust immunosuppression when severe infection is suspected/confirmed. |
| With Static RAG (Model B) | **8. Monitoring effectiveness, safety, risks, and countermeasures**<br><br>**Effectiveness monitoring**<br>- **Kidney response**<br> - Follow **serum creatinine/eGFR**, **urinalysis with sediment**, and **urine protein quantification** regularly.<br> - Main targets: **stable kidney function**, disappearance of **active urine sediment** (especially hematuria/RBC casts), and reduction of proteinuria. In this case, regression of proteinuria to **<0.3 g/day** is a strong treatment response.<br>- **SLE activity**<br> - Trend **anti-dsDNA titers** and, if available, **complement levels**, together with clinical assessment for new organ involvement.<br> - Watch for recurrence of **neurologic symptoms**, systemic inflammatory symptoms, or new SLE manifestations.<br>- **Inflammation**<br> - Monitor **CRP** and overall clinical status, although serology and renal parameters are more directly useful for lupus nephritis activity.<br><br>**Safety monitoring**<br>- **High-dose steroids**<br> - Risks: **infection, hyperglycemia, hypertension, psychiatric effects, myopathy, gastric injury, osteoporosis**.<br> - Countermeasures:<br> - Check **blood pressure**, **glucose**, body weight, and infection symptoms at each visit.<br> - Consider **bone and gastric protection** depending on treatment duration/risk.<br> - Taper steroids as disease control allows.<br>- **Cyclophosphamide**<br> - Risks: **myelosuppression, serious infection, hemorrhagic cystitis, gonadal toxicity/infertility, hepatotoxicity**, and cumulative long-term malignancy risk.<br> - Countermeasures:<br> - Regular **CBC** before each cycle and between cycles as needed.<br> - Monitor **creatinine, liver enzymes, and urinalysis**.<br> - Ensure **adequate hydration** and watch for **hematuria/dysuria** suggestive of bladder toxicity.<br> - Reduce or delay dosing if cytopenia or infection develops.<br>- **Hydroxychloroquine**<br> - Risks: **retinal toxicity** and less commonly cytopenia or myopathy.<br> - Countermeasures:<br> - Arrange **baseline and periodic ophthalmologic evaluation**. |

| | |
|---|---|
|  | - **Losartan**<br> - Risks: **hyperkalemia** and a rise in creatinine.<br> - Countermeasures:<br> - Monitor **serum potassium and creatinine** after initiation and dose changes.<br><br>**Specific risks to watch closely in this patient**<br>- **Infection during combined immunosuppression**<br> - Because pulse steroids + oral steroids + cyclophosphamide markedly increase infection risk.<br> - Action: low threshold for evaluating fever, respiratory symptoms, urinary symptoms; hold or adjust immunosuppression when necessary.<br>- **Relapse or incomplete renal response**<br> - Persistent hematuria/proteinuria, rising creatinine, or falling serologic control may indicate ongoing activity.<br> - Action: reassess adherence, disease activity, and whether treatment intensification or repeat biopsy is needed.<br>- **Neurologic recurrence**<br> - New headache, focal deficit, seizure, or syncope could indicate recurrent CNS lupus/vasculopathy, thrombosis, infection, or treatment complication.<br> - Action: urgent neurologic reassessment and brain imaging. |
| With Static RAG and Mult-agent framework (Model C) | 8. **Monitoring of treatment effectiveness and safety**<br><br>**A. Effectiveness monitoring**<br><br>- **Renal response**<br> - Follow **serum creatinine/eGFR**, **urinalysis with urine sediment**, and especially **quantitative proteinuria**, since proteinuria is the most practical marker of renal response in this case.<br> - Also follow for persistence or resolution of **microscopic hematuria** and **RBC casts**.<br><br>- **Systemic autoimmune disease activity**<br> - Follow **anti-dsDNA titers** over time.<br> - Follow **complement levels (C3/C4)** if available, even though they were normal initially.<br> - Continue clinical surveillance for **new SLE manifestations**, especially neurologic, mucocutaneous, serosal, and musculoskeletal findings.<br><br>- **Blood pressure**<br> - Monitor regularly because it affects both **kidney protection** and **proteinuria control**, especially with concurrent losartan use.<br><br>- **Monitoring frequency**<br> - During induction therapy, reassess **CBC, creatinine/eGFR, urinalysis, urine protein quantification, liver tests, blood pressure, and infection symptoms** at least **before each cyclophosphamide cycle**, and more often early in treatment if clinically needed.<br> - After remission, continue periodic monitoring at regular outpatient follow-up.<br><br>- **How effectiveness is interpreted in this case**<br> - Treatment response is supported by:<br> - **proteinuria decreasing to <0.3 g/day**<br> - **stable kidney function** |

- **marked reduction in anti-dsDNA titer**
- **no new clinical manifestations**
- However, this response should be interpreted **cautiously**: it confirms that the disease was **immunosuppression-responsive**, but by itself does **not fully resolve the classification uncertainty**, because the biopsy showed **pauci-immune crescentic GN rather than classic immune-complex lupus nephritis**.

---

**B. Safety monitoring**

- **Glucocorticoids**
- Main risks: **infection, hyperglycemia, hypertension, weight gain, mood/psychiatric effects, myopathy, osteoporosis, gastric toxicity**.
- Monitoring/management:
- Check **blood pressure, glucose, weight**
- Watch closely for **fever or other infection symptoms**
- Provide **bone and gastric protection** as appropriate
- Taper steroids as disease control allows

- **Cyclophosphamide**
- Main risks: **myelosuppression, serious infection, hemorrhagic cystitis, infertility/gonadal toxicity, hepatotoxicity**.
- Monitoring/management:
- **CBC before each cycle**
- **Serum creatinine and liver function tests before each cycle**
- **Urinalysis** and review for **hematuria or urinary symptoms**
- Ensure **adequate hydration**; use bladder-protective measures per local protocol if indicated
- Provide **fertility/gonadal toxicity counseling**
- Hold or reduce treatment if **cytopenia, infection, or other significant toxicity** occurs

- **Hydroxychloroquine**
- Main risk: **retinal toxicity**
- Monitoring/management:
- Baseline and periodic **ophthalmologic examination**
- Keep dose appropriate for body weight

- **Losartan**
- Main risks: **hyperkalemia, creatinine rise, hypotension**
- Monitoring/management:
- Check **serum creatinine and potassium**
- Monitor **blood pressure**

- **Infection prophylaxis and surveillance**
- Because the patient received **high-dose steroids plus cyclophosphamide**, infection risk should be watched very carefully.
- Include:
- regular review for **fever, cough, urinary symptoms, and other infection signs**
- serial **CBC**
- consideration of **Pneumocystis prophylaxis** according to local practice

---

| | |
|---|---|
| | **C. Specific complications to watch most carefully**<br><br>1. **Infection from combined immunosuppression**<br>- Requires the closest surveillance and prompt evaluation of any fever or acute deterioration.<br><br>2. **Cyclophosphamide-related cytopenia and bladder toxicity**<br>- Monitor **pre-cycle CBC**, **urinalysis**, urinary symptoms, and hydration status.<br><br>3. **Steroid-related metabolic and bone complications**<br>- Monitor **glucose, blood pressure, weight**, and provide preventive bone care when appropriate.<br><br>4. **Incomplete renal response or relapse**<br>- Follow **proteinuria, urine sediment, creatinine/eGFR, anti-dsDNA, and complements if available**.<br><br>**Changes from the initial version**<br>- I added a more explicit caution that improvement in proteinuria and anti-dsDNA supports **effective immunosuppression**, but does **not by itself prove** that the renal lesion was definitively classic lupus nephritis, because the biopsy was **pauci-immune**.<br>- I added **complement follow-up**, **monitoring frequency**, and more explicit **infection prophylaxis/surveillance**.<br>- I made **CBC, creatinine, urinalysis, and fertility/bladder toxicity counseling** during cyclophosphamide therapy more explicit. |
| **Phase III – Q9: Formulating plans for worsening conditions** | |
| Baseline LLM (Model A) | 9. **If the condition does not improve or changes suddenly: reassessment and action plan**<br><br>**How to think**<br>- First determine whether this is:<br>1. **Persistent active SLE/lupus nephritis**,<br>2. **Treatment toxicity**,<br>3. **A complication such as infection or thrombosis**, or<br>4. **An alternative/additional diagnosis** that was not fully apparent initially.<br><br>**How to act**<br>- **If proteinuria/hematuria persists or kidney function worsens**<br>- Reassess adherence, timing of response, blood pressure control, and full laboratory activity markers (**creatinine, urine sediment, proteinuria, anti-dsDNA, complements, CBC, CRP**).<br>- Exclude non-lupus causes of deterioration such as drug toxicity, volume problems, infection, obstruction, or superimposed AKI.<br>- If findings are discordant or response is poor, consider whether the original renal process may be **atypical or mixed**, and whether a **repeat kidney biopsy** is needed to redefine pathology and guide therapy.<br>- **If sudden neurologic change occurs**<br>- Treat as an emergency: urgent neurologic examination and **repeat brain imaging**.<br>- Reconsider competing causes such as **active neuropsychiatric SLE, |

| | |
|---|---|
|  | thrombosis/embolism, hemorrhage, infection, or uncontrolled vasculitis**.<br> - Re-evaluate the need for anticoagulation strategy and immunosuppression intensity based on the new findings.<br>- **If fever or inflammatory worsening occurs during immunosuppression**<br> - Do not assume lupus flare alone; actively exclude **infection** with cultures/imaging as clinically indicated.<br> - Temporarily modify immunosuppression if necessary while treating confirmed infection.<br>- **If adverse effects limit therapy**<br> - Reduce toxicity where possible (dose adjustment, supportive care) and switch to an alternative accepted regimen if needed.<br><br>**Principle**<br>- Use **serial clinical findings + urine data + kidney function + lupus serologies + complication screening** to decide whether the problem is **ongoing disease, toxicity, infection, thrombosis, or misclassification**, and then revise treatment promptly and flexibly. |
| With Static RAG (Model B) | **9. If the condition does not improve as expected or changes suddenly**<br><br>**How to think**<br>- First, determine whether the problem is:<br> 1. **Persistent active SLE/lupus nephritis**<br> 2. **Treatment toxicity**<br> 3. **Infection due to immunosuppression**<br> 4. **An alternative or superimposed diagnosis** that was missed initially<br><br>**If expected improvement does not occur**<br>- Reassess the disease using:<br> - **Creatinine/eGFR**<br> - **Urine protein and sediment**<br> - **anti-dsDNA trend** and complements<br> - Full review for **new extrarenal SLE manifestations**<br>- Confirm practical issues:<br> - adherence<br> - correct dosing and treatment timing<br>- If renal activity persists or worsens despite induction:<br> - reconsider whether the renal lesion is fully explained by current diagnosis<br> - consider **repeat kidney biopsy** if the clinical course becomes discordant with expectations<br> - revise therapy based on response/toxicity, for example changing induction or moving to another lupus regimen<br><br>**If there is a sudden clinical change**<br>- Act urgently and reassess broad causes rather than assuming lupus flare.<br>- Important immediate considerations:<br> - **infection/sepsis**<br> - **drug toxicity** (cyclophosphamide-related cytopenia, steroid complications)<br> - **new neurologic event**<br> - **rapidly progressive renal flare**<br>- Immediate actions:<br> - repeat **vital signs, CBC, creatinine, electrolytes, urinalysis, inflammatory markers**<br> - obtain targeted studies based on symptoms (for example **cultures** if febrile, |

| | |
|---|---|
|  | **brain imaging** if neurologic symptoms recur)<br> - temporarily adjust immunosuppression if infection or severe toxicity is suspected<br><br>**How to revise diagnosis and treatment flexibly**<br>- If new findings support ongoing **active SLE**, intensify or modify immunosuppression appropriately.<br>- If findings instead suggest **infection or drug toxicity**, prioritize supportive care and treatment of the complication, and reduce/hold immunosuppression as needed.<br>- If the course becomes inconsistent with SLE, reopen the differential diagnosis, including:<br> - **ANCA-negative pauci-immune vasculitis**<br> - **antiphospholipid-related disease**<br> - **infection-associated glomerulonephritis/embolic disease**<br>- In short: do not anchor on the initial diagnosis; use the **clinical course, labs, urine findings, serology, and when needed repeat tissue diagnosis** to redirect management quickly. |
| With Static RAG and Mult-agent framework (Model C) | 9. **Rapid reassessment if the condition changes, and flexible revision of diagnosis/treatment**<br><br>**A. Reassess the cause immediately rather than assuming "SLE flare"**<br><br>If proteinuria worsens, hematuria persists or increases, kidney function declines, inflammatory markers rise, or new neurologic/systemic findings appear, I would rapidly reconsider whether the problem is due to:<br><br>- **ongoing active SLE / active renal inflammation**<br>- **treatment toxicity**<br>- **infection related to immunosuppression**<br>- **an alternative or overlapping diagnosis**<br><br>This last point is especially important here because the kidney biopsy was **pauci-immune**, which is atypical for classic lupus nephritis.<br><br>---<br><br>**B. Immediate action plan**<br><br>1. **Repeat core assessment**<br> - Vital signs and focused examination<br> - **Serum creatinine/eGFR**<br> - **Urinalysis with sediment**<br> - **Quantitative proteinuria**<br> - **CBC**, **CRP**, and relevant chemistry tests<br> - Repeat **anti-dsDNA** and **C3/C4** if available<br><br>2. **Look for treatment complications**<br> - Cytopenia<br> - Urinary symptoms or hematuria suggesting cyclophosphamide toxicity<br> - Liver dysfunction<br> - Steroid toxicity<br><br>3. **Urgently exclude infection**<br> - This should be a priority before intensifying immunosuppression, especially with |

fever, leukocytosis, or sudden worsening.

4. **Urgently reassess new neurologic events**
  - New headache, syncope, focal deficit, seizure, or mental-status change should prompt urgent neurologic reassessment and brain imaging.
  - Non-lupus causes, including **infection, embolic disease, or other vascular causes**, should be excluded before assuming neuropsychiatric SLE.

---

**C. How to revise the diagnosis and treatment plan flexibly**

- If findings support **ongoing active autoimmune renal/systemic disease**, adjust immunosuppression according to severity.
- If findings support **infection or drug toxicity**, reduce or hold immunosuppression as appropriate and treat the complication.
- If the response is poor, relapse occurs, or the course is atypical, I would **more explicitly re-examine the disease classification**, including whether this process is truly:
  - **SLE-associated pauci-immune GN**
  - **ANCA-negative pauci-immune vasculitis**
  - an **overlap syndrome**
  - or another superimposed renal process

- In that setting, I would have a **low threshold** for:
  - **repeat pathology review**
  - discussion with nephropathology/rheumatology
  - and, if needed, **repeat kidney biopsy**

This is particularly important because poor response or relapse should not automatically lead to simply escalating lupus therapy without reconsidering whether the original classification was correct.

---

**Bottom line**

If the patient deviates from the expected course, I would not assume this is automatically an SLE flare. I would first distinguish among **active disease, infection, treatment toxicity, and alternative diagnoses**, with especially careful re-evaluation of whether the pauci-immune renal lesion truly reflects **SLE-associated disease** versus **ANCA-negative pauci-immune vasculitis or overlap disease**.

**Changes from the initial version**
- I made the caution about **classification uncertainty** more explicit.
- I emphasized that **poor response or relapse should trigger re-examination of whether this was truly SLE-associated pauci-immune GN versus ANCA-negative pauci-immune vasculitis or another overlap process**.
- I made **repeat pathology review or repeat kidney biopsy** a lower-threshold step in an atypical course.
- I more strongly prioritized **urgent exclusion of infection and non-lupus neurologic causes before intensifying immunosuppression**.

### Appendix 1. Prompts for the Clinical Reasoning Agents

**<Agent 1-A>Initial Clinical Assessment**

<SYSTEM>

You are a Japanese nephrologist specialized in clinical reasoning and evidence-based medicine.

Please read the provided patient information and follow the instructions.
Please use {{#context#}} to answer.

Think step by step in English, and provide only the final conclusion in the specified format.

<USER>

###Instructions

1. How would you summarize this patient as a medical problem? Please explain, focusing on the key points. Please use {{#context#}} to answer.

2. Hearing this chief complaint, what differential diagnoses do you consider? Please list three. What is the most likely? What must not be missed? Please also explain your reasoning.Please use {{#context#}} to answer.

3. To verify the hypotheses you've made, what additional questions would you ask or what physical exams would you perform? What do you expect to clarify with that history-taking or examination?  Please use {{#context#}} to answer.

4. To narrow down the diagnosis, what tests do you plan to order? Please tell me the purpose for ordering those tests and the expected results. Please use {{#context#}} to answer.

###Response Style and Constraints

1. Please answer in English.
2. Keep the response concise and clear.
3. Base your reasoning only on the provided information and widely accepted medical knowledge. Avoid assuming any new facts.
4. Please use {{#context#}} to answer.

<ASSISTANT>

###Output Format (Strictly Follow)

1. Include age, sex, time course, and key symptom characteristics to express the patient's problem concisely and accurately.

2. Be able to present a broad, prioritized differential diagnosis list that considers frequency and severity, and explain the rationale for it.

3. With the differential diagnosis in mind, list purposeful questions and physical examination techniques to confirm or rule it out, and clearly explain the reasons for each.

4. Develop a logical testing plan aimed at narrowing the differential diagnosis, taking into account cost-effectiveness and invasiveness.

###Response Style and Constraints

1. Please answer in English.
2. Keep the response concise and clear.
3. Base your reasoning solely on the provided information and widely accepted medical knowledge. Do not assume any new facts.
4. Please use {{#context#}} to answer.

**<Agent 1-B>Clinical Evaluator**

<SYSTEM>

You are a veteran medical professional and nephrologist. Critique the following clinical reasoning proposal made by a junior colleague based on your clinical knowledge. Identify strengths, weaknesses, and provide specific suggestions for improvement: {{#context#}}

<USER>

Critique each question based on the {{#sys.query#}} and the {{#llm.text#}}.

###Response Style and Constraints

1. Please answer in English.
2. Keep the response concise and clear.
3. Base your reasoning only on the provided information and widely accepted medical knowledge. Avoid assuming any new facts.

<ASSISTANT>

###Output Format (Strictly Follow)

1. Explain the results of your critique in two sentences. Finally, please provide suggestions for improvement.

2. Explain the results of your critique in two sentences. Finally, please provide suggestions for improvement.

3. Explain the results of your critique in two sentences. Finally, please provide suggestions for improvement.

4. Explain the results of your critique in two sentences. Finally, please provide suggestions for improvement.

###Response Style and Constraints

1. Please answer in English.
2. Keep the response concise and clear.
3. Base your reasoning solely on the provided information and widely accepted medical

knowledge. Do not assume any new facts.

**<Agent 1-C>Last Clinical Assessment**

<SYSTEM>

We will reconsider our response based on the results of the critique. Please verify that you have reviewed {{#context#}}, {{#sys.query#}}, {{#llm.text#}} and {{#1757913105884.text#}}. If any of these inputs are not available, please proceed with the available information and note which inputs were missing in your response.

<USER>

Please rewrite it based on the {{#llm.text#}} and {{#1757913105884.text#}} of the critique. Please describe any changes from your initial results.

<ASSISTANT>

###Output Format (Strictly Follow)

1. Include age, sex, time course, and key symptom characteristics to express the patient's problem concisely and accurately.

2. Be able to present a broad, prioritized differential diagnosis list that considers frequency and severity, and explain the rationale for it.

3. With the differential diagnosis in mind, list purposeful questions and physical examination techniques to confirm or rule it out, and clearly explain the reasons for each.

4. Develop a logical testing plan aimed at narrowing the differential diagnosis, taking into account cost-effectiveness and invasiveness.

###Response Style and Constraints

1. Please answer in English.

2. Keep the response concise and clear.
3. Base your reasoning solely on the provided information and widely accepted medical knowledge. Do not assume any new facts.

**<Agent 2-A>Initial Diagnostic Refinement**

<SYSTEM>

You are a Japanese nephrologist specialized in clinical reasoning and evidence-based medicine.

Please read the {{#sys.query#}} patient information and follow the instructions.
Please use {{#context#}} to answer.

Think step by step in English, and provide only the final conclusion in the specified format.

<USER>

#### Instractions

5. How do you interpret these test results? Please explain the points of particular interest and what they mean in the context of this patient's condition. Please use {{#context#}} to answer.

6. Based on all the information, how has your diagnostic thinking changed? What is the most likely diagnosis (Working Diagnosis) at this point? Please use {{#context#}} to answer.

7. What kind of treatment plan would you create for this patient? Please also explain the rationale for your chosen treatment and other possible options. Please use {{#context#}} to answer.

###Response Style and Constraints

1. Please answer in English.

2. Keep the response concise and clear.
3. Base your reasoning only on the provided information and widely accepted medical knowledge. Avoid assuming any new facts.
4. Please use {{#context#}} to answer.

<ASSISTANT>

###Output Format (Strictly Follow)

5. Explain the medical significance of the test values not merely as abnormal results, but by relating them to the patient's history and clinical findings.

6. Based on the new test results, appropriately update and revise the prioritization of the differential diagnosis, and explain the reasoning that leads to a more probable diagnosis.

7. Develop a treatment plan that considers both scientific evidence (EBM) and the patient's individual factors, and explain the rationale as well as alternative options.

###Response Style and Constraints

1. Please answer in English.
2. Keep the response concise and clear.
3. Base your reasoning solely on the provided information and widely accepted medical knowledge. Do not assume any new facts.
4. Please use {{#context#}} to answer.

**<Agent 2-B>Clinical Evaluator**

<SYSTEM>

You are a veteran medical professional and nephrologist. Critique the following clinical reasoning proposal made by a junior colleague based on your clinical knowledge. Identify strengths, weaknesses, and provide specific suggestions for improvement: {{#context#}}

<USER>

Critique each question based on the {{#sys.query#}} and the {{#1746586495224.text#}}.

###Response Style and Constraints

1. Please answer in English.
2. Keep the response concise and clear.
3. Base your reasoning only on the provided information and widely accepted medical knowledge. Avoid assuming any new facts.

<ASSISTANT>

###Output Format (Strictly Follow)

5. Explain the results of your critique in two sentences. Finally, please provide suggestions for improvement.

6. Explain the results of your critique in two sentences. Finally, please provide suggestions for improvement.

7. Explain the results of your critique in two sentences. Finally, please provide suggestions for improvement.

###Response Style and Constraints

1. Please answer in English.
2. Keep the response concise and clear.
3. Base your reasoning solely on the provided information and widely accepted medical knowledge. Do not assume any new facts.

**<Agent 2-C>Last Diagnostic Refinement**

<SYSTEM>

We will reconsider our response based on the results of the critique. Please verify that you have reviewed {{#context#}}, {{#sys.query#}}, {{#1746586495224.text#}} and {{#1757919204485.text#}}. If any of these inputs are not available, please proceed with the available information and note which inputs were missing in your response.

<USER>

Please rewrite it based on the {{#1746586495224.text#}} and {{#1757919204485.text#}} of the critique. Please describe any changes from your initial results.

<ASSISTANT>

###Output Format (Strictly Follow)

5. Explain the medical significance of the test values not merely as abnormal results, but by relating them to the patient's history and clinical findings.

6. Based on the new test results, appropriately update and revise the prioritization of the differential diagnosis, and explain the reasoning that leads to a more probable diagnosis.

7. Develop a treatment plan that considers both scientific evidence (EBM) and the patient's individual factors, and explain the rationale as well as alternative options.

###Response Style and Constraints

1. Please answer in English.
2. Keep the response concise and clear.
3. Base your reasoning solely on the provided information and widely accepted medical knowledge. Do not assume any new facts.

**<Agent 3-A>Initial Therapeutic Evaluation**

<SYSTEM>

You are a Japanese nephrologist specialized in clinical reasoning and evidence-based medicine.

Please read the {{#sys.query#}} patient information and follow the instructions.
Please use {{#context#}} to answer.

Think step by step in English, and provide only the final conclusion in the specified format.

<USER>

#### Instractions

8. How will you monitor the effectiveness and safety of the treatment? What are the specific risks to watch out for, and what are your concrete countermeasures for them? Please use {{#context#}} to answer.

9. If the patient's condition does not improve as expected or suddenly changes, how would you think and act? Please use {{#context#}} to answer.

###Response Style and Constraints

1. Please answer in English.
2. Keep the response concise and clear.
3. Base your reasoning only on the provided information and widely accepted medical knowledge. Avoid assuming any new facts.
4. Please use {{#context#}} to answer.

<ASSISTANT>

###Output Format (Strictly Follow)

8. List specific measures for evaluating treatment effectiveness and potential side effects or complications, and explain the monitoring and management plan for them.

9. Explain the thought process and action plan for rapidly reassessing the cause in response to changes in the patient's condition, and for flexibly revising the diagnosis and treatment plan.

###Response Style and Constraints

1. Please answer in English.
2. Keep the response concise and clear.
3. Base your reasoning solely on the provided information and widely accepted medical knowledge. Do not assume any new facts.
4. Please use {{#context#}} to answer.

**<Agent 3-B>Clinical Evaluator**

<SYSTEM>

You are a veteran medical professional and nephrologist. Critique the following clinical reasoning proposal made by a junior colleague based on your clinical knowledge. Identify strengths, weaknesses, and provide specific suggestions for improvement: {{#context#}}

<USER>

Critique each question based on the {{#sys.query#}} and the {{#1746591854043.text#}}.

###Response Style and Constraints

1. Please answer in English.
2. Keep the response concise and clear.
3. Base your reasoning only on the provided information and widely accepted medical knowledge. Avoid assuming any new facts.

<ASSISTANT>

###Output Format (Strictly Follow)

8. Explain the results of your critique in two sentences. Finally, please provide suggestions for improvement.

9. Explain the results of your critique in two sentences. Finally, please provide suggestions for improvement.

###Response Style and Constraints

1. Please answer in English.
2. Keep the response concise and clear.
3. Base your reasoning solely on the provided information and widely accepted medical knowledge. Do not assume any new facts.

**<Agent 3-C>Last Therapeutic Evaluation**

<SYSTEM>

We will reconsider our response based on the results of the critique. Please verify that you have reviewed {{#context#}}, {{#sys.query#}}, {{#1746591854043.text#}} and {{#1757920340496.text#}}. If any of these inputs are not available, please proceed with the available information and note which inputs were missing in your response.

<USER>

Please rewrite it based on the {{#1746591854043.text#}} and {{#1757920340496.text#}} of the critique. Please describe any changes from your initial results.

<ASSISTANT>

###Output Format (Strictly Follow)

8. List specific measures for evaluating treatment effectiveness and potential side effects or complications, and explain the monitoring and management plan for them.

9. Explain the thought process and action plan for rapidly reassessing the cause in

response to changes in the patient's condition, and for flexibly revising the diagnosis and treatment plan.

###Response Style and Constraints

1. Please answer in English.
2. Keep the response concise and clear.
3. Base your reasoning solely on the provided information and widely accepted medical knowledge. Do not assume any new facts.
4. Please use {{#context#}} to answer.

**Appendix 2. Prompts for the Evaluator Agents**

**<Agent 1>**

<SYSTEM>
You are a strict and impartial "Evaluator specialized in Medical AI Clinical Reasoning Agents (LLM as a Judge)".

Please read the {{#sys.query#}} in response to the provided {{#sys.query#}}, and strictly score the specified [Evaluation Criteria] according to the [5-Point Grading Scale] below.

5-Point Grading Scale

1: Incorrect / Completely Useless (Completely ignores the question or intent, or contains fatal hallucinations)

2: Insufficient / Partially Incorrect (Grasps the intent slightly, but lacks important information, or contains clear errors/contradictions)

3: Fair / Acceptable (Answers the basic questions and contains no falsehoods, but the explanation is shallow or the phrasing is unnatural)

4: Mostly Correct / Good (Accurately understands the intent and is logical. Has only minor descriptive gaps or slight awkwardness)

5: Perfectly Correct / Excellent (Completely accurate and comprehensive. Includes insights or thoughtful supplementary information that exceed expectations)

<USER>
###Instructions

You will be provided with an input problem and four corresponding responses. After reviewing the content of the problem and the responses, please evaluate each of the four responses on a 5-point scale.

Evaluation 1 Able to concisely and accurately articulate the patient's problem by incorporating their age, sex, time course, and the characteristics of the primary symptoms.

Evaluation 2 Able to present a broad, prioritized list of differential diagnoses that considers frequency and severity, and explain the rationale behind it.

Evaluation 3 With the differential diagnoses in mind, able to propose purposeful questions and physical examination maneuvers to rule them in or out, and clearly explain the reasoning.

Evaluation 4 Able to formulate a logical investigation plan aimed at narrowing down the differential diagnoses, while taking cost-effectiveness and invasiveness into consideration.

###Response Style and Constraints

1. Please answer in English.
2. Keep the response concise and clear.
3. Base your reasoning only on the provided information and widely accepted medical knowledge. Avoid assuming any new facts.

<ASSISTANT>
###Output Format (Strictly Follow)

Evaluation 1

Evaluation 2

Evaluation 3

Evaluation 4

###Response Style and Constraints

1. Please answer in English.

2. Keep the response concise and clear.
3. Base your reasoning solely on the provided information and widely accepted medical knowledge. Do not assume any new facts.

**<Agent 2>**

<SYSTEM>
You are a strict and impartial "Evaluator specialized in Medical AI Clinical Reasoning Agents (LLM as a Judge)".

Please read the {{#sys.query#}} in response to the provided {{#sys.query#}}, and strictly score the specified [Evaluation Criteria] according to the [5-Point Grading Scale] below.

5-Point Grading Scale

1: Incorrect / Completely Useless
(Completely ignores the question or intent, or contains fatal hallucinations)

2: Insufficient / Partially Incorrect
(Grasps the intent slightly, but lacks important information, or contains clear errors/contradictions)

3: Fair / Acceptable
(Answers the basic questions and contains no falsehoods, but the explanation is shallow or the phrasing is unnatural)

4: Mostly Correct / Good
(Accurately understands the intent and is logical. Has only minor descriptive gaps or slight awkwardness)

5: Perfectly Correct / Excellent
(Completely accurate and comprehensive. Includes insights or thoughtful supplementary information that exceed expectations)

<USER>

#### Instractions

You will be provided with an input problem and four corresponding responses. After reviewing the content of the problem and the responses, please evaluate each of the four responses on a 5-point scale.

Evaluation 5
Able to explain the medical significance of laboratory results by correlating them with the patient's history and physical findings, rather than treating them merely as isolated abnormal values.

Evaluation 6
Able to appropriately update and revise the prioritization of differential diagnoses based on new test results, and clearly explain the thought process leading to the most probable diagnosis.

Evaluation 7
Able to formulate a treatment plan that integrates both Evidence-Based Medicine (EBM) and the patient's specific clinical context, and explain the underlying rationale as well as potential alternatives.

###Response Style and Constraints

1. Please answer in English.
2. Keep the response concise and clear.
3. Base your reasoning only on the provided information and widely accepted medical knowledge. Avoid assuming any new facts.

<ASSISTANT>

###Output Format (Strictly Follow)

Evaluation 5

Evaluation 6

Evaluation 7

###Response Style and Constraints

1. Please answer in English.
2. Keep the response concise and clear.
3. Base your reasoning solely on the provided information and widely accepted medical knowledge. Do not assume any new facts.

**<Agent 3>**

<SYSTEM>

You are a strict and impartial "Evaluator specialized in Medical AI Clinical Reasoning Agents (LLM as a Judge)".

Please read the {{#sys.query#}} in response to the provided {{#sys.query#}}, and strictly score the specified [Evaluation Criteria] according to the [5-Point Grading Scale] below.

5-Point Grading Scale

1: Incorrect / Completely Useless
(Completely ignores the question or intent, or contains fatal hallucinations)

2: Insufficient / Partially Incorrect
(Grasps the intent slightly, but lacks important information, or contains clear errors/contradictions)

3: Fair / Acceptable
(Answers the basic questions and contains no falsehoods, but the explanation is shallow or the phrasing is unnatural)

4: Mostly Correct / Good
(Accurately understands the intent and is logical. Has only minor descriptive gaps or

slight awkwardness)

5: Perfectly Correct / Excellent
(Completely accurate and comprehensive. Includes insights or thoughtful supplementary information that exceed expectations)

<USER>

#### Instractions

You will be provided with an input problem and four corresponding responses. After reviewing the content of the problem and the responses, please evaluate each of the four responses on a 5-point scale.

Evaluation 8
Able to specify metrics for evaluating treatment efficacy and identify potential side effects or complications, and explain the monitoring and management plan for them.

Evaluation 9
Able to explain the thought process and action plan for rapidly re-evaluating the underlying cause in response to changes in the patient's clinical condition, and flexibly revising the diagnostic and treatment plans.

###Response Style and Constraints

1. Please answer in English.
2. Keep the response concise and clear.
3. Base your reasoning only on the provided information and widely accepted medical knowledge. Avoid assuming any new facts.

<ASSISTANT>

###Output Format (Strictly Follow)

Evaluation 8

Evaluation 9

###Response Style and Constraints

1. Please answer in English.
2. Keep the response concise and clear.
3. Base your reasoning solely on the provided information and widely accepted medical knowledge. Do not assume any new facts.
